# The effect of exercise on aerobic capacity in individuals with spinal cord injury: A systematic review with meta-analysis and meta-regression

**DOI:** 10.1101/2022.08.05.22278397

**Authors:** Daniel D. Hodgkiss, Gurjeet S. Bhangu, Carole Lunny, Catherine R. Jutzeler, Shin-Yi Chiou, Matthias Walter, Samuel J. E. Lucas, Andrei V. Krassioukov, Tom E. Nightingale

## Abstract

**Background:** A low level of cardiorespiratory fitness [CRF; defined as peak oxygen uptake (V̇O_2peak_) or peak power output (PPO)] is a widely reported consequence of spinal cord injury (SCI) and a major risk factor associated with chronic disease. However, CRF can be modified by exercise. This systematic review with meta-analysis and meta-regression aimed to assess whether certain SCI characteristics and/or specific exercise considerations are moderators of changes in CRF.

**Methods and Findings:** Databases (MEDLINE, EMBASE, CENTRAL and Web of Science) were searched from inception to March 2023. A primary meta-analysis was conducted including randomised controlled trials (RCTs; exercise interventions lasting >2 weeks relative to control groups). A secondary meta-analysis pooled independent exercise interventions >2 weeks from longitudinal pre-post and RCT studies to explore whether subgroup differences in injury characteristics and/or exercise intervention parameters explained CRF changes. Further analyses included cohort, cross-sectional and observational study designs. Outcome measures of interest were absolute (AV̇O_2peak_) or relative V̇O_2peak_ (RV̇O_2peak_), and/or PPO. Bias/quality was assessed via The Cochrane Risk of Bias 2 and the National Institute of Health Quality Assessment Tools. Certainty of the evidence was assessed using the Grading of Recommendations Assessment, Development and Evaluation (GRADE) approach. Random effects models were used in all meta-analyses and meta-regressions.

Of 21,020 identified records, 120 studies comprising 29 RCTs, 67 pre-post studies, 11 cohort, 7 cross-sectional and 6 observational studies were included. The primary meta-analysis revealed significant improvements in AV̇O_2peak_ [0.16 (0.07, 0.25) L/min], RV̇O_2peak_ [2.9 (1.8, 3.9) mL/kg/min] and PPO [9 (5, 14) W] with exercise, relative to controls (*p*<0.001). Ninety-six studies (117 independent exercise interventions comprising 1,331 participants) were included in the secondary, pooled meta-analysis which demonstrated that exercise interventions significantly improve AV̇O_2peak_ [0.22 (0.17, 0.26) L/min], RV̇O_2peak_ [2.8 (2.2, 3.3) mL/kg/min], and PPO [11 (9, 13) W] (*p*<0.001). There were subgroup differences for RV̇O_2peak_ based on exercise modality (*p*=0.002) and intervention length (*p*=0.01), but there were no differences for AV̇O_2peak_. There were subgroup differences (*p*≤0.008) for PPO based on time since injury, neurological level of injury, exercise modality, relative exercise intensity, exercise intensity prescription method, and frequency. The meta-regression found that studies with a higher mean age of participants were associated with smaller changes in AV̇O_2peak_ and RV̇O_2peak_ (*p*<0.10). GRADE indicated a moderate level of certainty in the estimated effect for RV̇O_2peak_, but low levels for AV̇O_2peak_ and PPO. This review may be limited by the small number of RCTs, which prevented a subgroup analysis within this specific study design.

**Conclusions:** Performing exercise >2 weeks results in significant improvements to AV̇O_2peak_, RV̇O_2peak_ and PPO in individuals with SCI. Subgroup comparisons identified that exercise interventions lasting up to 12 weeks yield the greatest change in RV̇O_2peak_. Upper-body aerobic exercise and resistance training also appear the most effective at improving RV̇O_2peak_ and PPO. Furthermore, acutely-injured, individuals with paraplegia, exercising at a moderate-to-vigorous intensity, prescribed via a percentage of oxygen consumption or heart rate, for ≥3 sessions/week will likely experience the greatest change in PPO. Ageing seemingly diminishes the adaptive CRF responses to exercise training in individuals with SCI.

**Registration:** PROSPERO: CRD42018104342

**AUTHOR SUMMARY:** *Why was this research done?:* - Individuals with spinal cord injury typically exhibit low levels of cardiorespiratory fitness. As such, these individuals are at a higher risk for the development of chronic diseases in comparison to the non-injured population. - The current spinal cord injury-specific exercise guidelines encourage moderate-to-vigorous intensity aerobic exercise 40 minutes per week for fitness benefits or 90 minutes per week for cardiometabolic health benefits. Yet, others have suggested individuals with spinal cord injury should be achieving 150 minutes per week in line with non-injured population guidelines. - This systematic review with meta-analysis and meta-regression aimed to identify whether specific injury characteristics (e.g., time, level or severity of injury) or exercise intervention parameters (e.g., modality, intensity, volume etc.) result in the greatest changes in cardiorespiratory fitness in individuals with spinal cord injury.

*What did the researchers do and find?:* - We searched for studies that investigated the effects of exercise interventions lasting longer than 2 weeks on changes in absolute and relative peak oxygen consumption and/or peak power output in individuals with spinal cord injury. In total, we included 120 studies of various study designs: 29 randomised controlled trials, 67 pre-post studies, 11 cohort comparisons, 7 cross-sectional studies and 6 observational studies. - The greatest changes in peak power output may be achieved by individuals with acute spinal cord injury or paraplegia. Upper-body aerobic and resistance exercise were identified as the most optimal exercise modalities. Furthermore, prescribing moderate-to-vigorous intensity aerobic exercise using either a percentage of the individual’s peak heart rate or oxygen consumption, for three or more sessions per week, resulted in the greatest improvements in peak power output. - Our findings support the minimum 40 minutes of weekly moderate-to-vigorous intensity exercise recommended by the spinal cord injury-specific exercise guidelines to significantly improve fitness. However, while not statistically significant, a two-fold greater improvement in peak power output was shown for interventions with exercise performed ≥90 min/week in comparison to ≥40 min/week. Cross-sectional comparisons also revealed that individuals with spinal cord injury performing higher levels of physical activity were associated with higher cardiorespiratory fitness.

*What do these findings mean?:* - Exercise interventions >2 weeks can significantly improve cardiorespiratory fitness in individuals with a spinal cord injury by a clinically meaningful change greater than one spinal cord injury adjusted metabolic equivalent (i.e., ≥2.7 mL/kg/min). A one metabolic equivalent improvement has been associated with a reduction in cardiovascular related mortality risk in non-injured individuals. - Our findings indicate that certain participant/injury characteristics and exercise intervention parameters are moderators of the changes observed in cardiorespiratory fitness across studies. These factors should be considered in the design of future exercise interventions. Future research should consider: following spinal cord injury-specific reporting guidelines (ensuring transparency of reporting), investigating the dose-response relationship between exercise and cardiorespiratory fitness or influence of exercise intensity in this population, and consider how different injury characteristics impact the benefits of exercise on cardiorespiratory fitness. - The main limitation of the study was the lack of randomised controlled trials (RCT) comparing changes in CRF following an exercise intervention relative to a control group. This prevented subgroup comparisons in this study design specifically and therefore we pooled pre-post and RCT exercise interventions to explore these effects.

## 1. INTRODUCTION

Spinal cord injury (SCI) is a complex neurological condition, caused by trauma, disease or degeneration, which results in sensory-motor deficits (i.e., paralysis or paresis) below the level of lesion and autonomic dysfunctions. Progressive physical deconditioning following injury results in increased health care utilisation, reliance on personal assistance services and a greater predisposition towards developing chronic diseases [1,2]. Individuals with SCI are at an increased risk of stroke, cardiovascular disease (CVD), and type-2 diabetes mellitus compared to non-injured counterparts [3–5]. The elevated incidence of these conditions in people with SCI emphasises the need for targeted interventions to address modifiable risk factors for these chronic diseases, such as cardiorespiratory fitness (CRF). In clinical populations CRF is typically defined as an individual’s peak oxygen uptake (V̇O_2peak_) or peak power output (PPO). V̇O_2peak_ and PPO are determined during graded cardiopulmonary exercise testing (CPET) to the point of volitional exhaustion, and represents the integrated functioning of different bodily systems (pulmonary, cardiovascular and skeletal) to uptake, transport and utilise oxygen for metabolic processes [6]. A number of prospective studies have indicated that CRF is at least as important, if not more so, than other traditional CVD risk factors (e.g., obesity, hypertension and smoking) and is strongly associated with mortality [7–12].

Low levels of CRF have been widely reported in the SCI-population [13], with the between-person variability partially explained by the neurological level and severity of injury (i.e., lower CRF reported in individuals with tetraplegia) [14]. A large proportion of the variance in CRF is also explained by physical activity [15], which is reduced in the SCI-population [16,17]. Performing regular physical activity and/or structured exercise has long been promoted for improving CRF in individuals with SCI [18,19]. In 2011, the first evidence-based exercise guidelines, specifically for individuals with SCI were developed [20], which stated that *“for important fitness benefits, adults with SCI should engage in at least 20 minutes of moderate-to-vigorous-intensity aerobic activity and strength-training exercises 2 times per week”*. This guideline has since been updated, yet remains the same with regards to CRF benefits [21]. Although this implies adults with SCI can accrue fitness benefits from volumes of activity well below that promoted in the general population, others have advocated that adults with a physical disability [22,23] and individuals with SCI [24] should aim to perform at least 150 minutes of aerobic exercise per week. For additional health benefits it has been suggested that adults should perform closer to 300 minutes per week of moderate-intensity physical activity [25,26]. While the current SCI-specific guidelines likely represent the *“minimum”* threshold required to achieve CRF benefits, it has been suggested that this creates an impression that individuals with SCI do not need to be as physically active as the general population [27]. The dose-response relationship between exercise volume and CRF improvements in individuals with SCI remains to be elucidated.

It is noteworthy that the aforementioned SCI-specific exercise guidelines utilise the terminology of *“moderate-to-vigorous”* to describe the desired exercise intensity. This is in contrast to accepted guidelines in the general population whereby moderate and vigorous-intensity exercise are distinguished from one another with specific thresholds (e.g., ≥150 minutes of moderate-intensity or ≥75 minutes of vigorous-intensity activity per week) [23]. Exercise intervention intensity has been shown to influence the magnitude of change in CRF in patients undergoing cardiac rehabilitation [28,29]. The feasibility/effectiveness of higher intensity exercise is also currently a topical area of research in the SCI-population [30–32]. There is the potential for vigorous-intensity exercise to be more time efficient or lead to superior health benefits, although the impact of different high-intensity interval training protocols on CRF in individuals with SCI compared to moderate-intensity exercise is yet to be determined. A recent systematic review identified that exercise interventions of a specific modality yield distinct changes in certain cardiometabolic health outcomes, but not other outcomes, in individuals with SCI [33]. This provides rationale for wanting to investigate the efficacy of different exercise modalities on CRF in this population. Consequently, a number of research questions requiring further attention include:

1. Do injury-specific characteristics (e.g., tetraplegia vs. paraplegia, acute vs. chronic injuries, motor-complete vs. incomplete) mediate CRF responses to exercise?
2. What is the best intensity, frequency, and volume of weekly exercise?
3. Is there an optimal conditioning modality [e.g., upper-body aerobic exercise, resistance training, functional electrical stimulation (FES), hybrid or multimodal exercise interventions etc.]?

To address these questions, we performed a systematic review with meta-analysis and meta-regression to investigate the impact of different exercise interventions on changes in CRF in individuals with SCI. Moreover, we gathered evidence to determine whether key moderators (e.g., participant/injury characteristics, intervention/study characteristics and risk of bias) influence these intervention effects.

## 2. METHODS

This review is reported as per the Preferred Reporting Items for Systematic Reviews and Meta-Analyses (PRISMA) guidelines [34] (S1 Checklist) and was prospectively registered (PROSPERO ID CRD42018104342). The primary meta-analysis of this review included randomised controlled trials (RCTs; exercise intervention versus a comparison control group). A secondary, pooled meta-analysis was also conducted that combined the intervention arms of RCTs with non-randomised study designs (RCTs and pre-post exercise interventions without a comparison control group). Further secondary meta-analyses were conducted that included cohort comparisons (e.g., physically inactive vs. habitual exercisers) and observational studies (e.g., standard of care rehabilitation), along with RCTs that specifically compared the impact of different exercise intensities (e.g., low or moderate-intensity vs. vigorous or supramaximal) on CRF outcomes. Lastly, we qualitatively reviewed cross-sectional studies that reported associations between physical activity and CRF outcomes.

### 2.1. Eligibility criteria

Studies were required to meet the following inclusion criteria: 1) Adult (≥18 years) participants; 2) any acquired (traumatic, infection, cancer) SCI (*note, studies were included if >80% of the sample met these two aforementioned inclusion criteria*); 3) an exercise or physical activity intervention lasting >2 weeks (RCTs, pre-post and observational trials) ; 4) report a measurable exposure variable (i.e., cohort studies: athletes vs. non-athletes or sedentary vs. active participants; and cross-sectional studies: self-reported or objectively measured habitual physical activity level) and; 5) report CRF-specific outcomes [i.e., absolute (AV̇O_2peak_) or expressed as relative to body mass V̇O_2peak_ (RV̇O_2peak_), evaluated via analysis of expired air during a peak (or symptom-limited) CPET or submaximal prediction, or PPO].

Studies were excluded if they met the following criteria: 1) non-human; 2) non-original work (i.e., reviews, guideline documents, editorials, viewpoints, letter-to-editor, protocol paper); 3) case-reports and case series with a number (n) of participants <5 (to increase the robustness of our findings given the inclusion of smaller sample sizes in previous reviews [18,35,36]); 4) non-peer reviewed (i.e., conference proceeding/abstracts/posters); 5) children or adolescents (<18 years); 6) non-SCI (non-injured participants or other neurological conditions); 7) does not report a CRF-specific outcome; 8) single exercise sessions or an intervention <2 weeks; 9) no suitable comparison (i.e., control group or baseline data pre-intervention) or exposure variable measured; and 10) no full text. Studies with concurrent interventions (i.e., diet, lifestyle or respiratory training) were included only if the effects of exercise could be isolated.

### 2.2. Search strategy

A search of the following electronic databases: MEDLINE (via Pubmed), Excerpta Medica Database (EMBASE; via Ovid), Web of Science and the Cochrane Central Register of Controlled Trials (CENTRAL) was conducted from their respective inception through to March 25, 2023. Search terms were developed by the corresponding author (TN) and agreed upon by co-authors (AK, MW). The search strategy combined key words describing the following: 1) condition (e.g., SCI); 2) ‘intervention or exposure variable’ (e.g., rehabilitation, exercise and physical activity); and 3) ‘outcome’ (e.g., V̇O_2peak_ or PPO). Details of the complete search strategy can be found in online supplementary material (S2). Search results were collated using Endnote software (Thomson Reuters, NY) and duplicates removed.

### 2.3. Study selection and data extraction

The citations retrieved from the search strategy were screened by title, abstract, and full text by two independent reviewers (DH, GB). At each stage of the evaluation, studies were excluded if the inclusion criteria were not satisfied. A conservative approach was taken, whereby if insufficient information was available to warrant study exclusion during the title and abstract stages of the screening, studies were retained in the sample for full text screening. TN resolved any disagreement with regards to study inclusion. There was no restriction on the language of studies. Where necessary, reviewers screened using Google translate [37] or sought assistance from bilingual co-authors.

Two authors (DH, GB) independently extracted data in duplicate using Microsoft Excel. Any disagreements were resolved via mutual consensus. Where more than one publication was apparent for the same participants, data were extracted from the study with the largest sample size to avoid duplication. Author, year, study design, sample size, participant demographics/injury characteristics, exercise parameters (including the type, frequency, duration, intensity and weekly volume), or physical activity exposure details (training history, objective wearable device or validated self-report questionnaire) and adverse events were extracted. For RCTs, pre-post interventions and observational studies, mean ± standard deviation (SD) for V̇O_2peak_ and PPO outcomes at baseline and post-intervention/control or observation period were extracted to assess change in CRF. For cross-sectional studies, mean ± SD outcomes were extracted for the unique cohorts, along with the significance and magnitude of associations between CRF and habitual physical activity. Where possible, V̇O_2peak_ values were extracted in relative (mL/kg/min) and absolute (L/min) terms or calculated using pre- and post-intervention body mass values when provided. As it is widely accepted that V̇O_2peak_ should be expressed in relative terms, to account for changes in body mass, results are presented for both AV̇O_2peak_ and RV̇O_2peak_, but the discussion focuses primarily on RV̇O_2peak_. PPO values were extracted in watts (W) only. If there was insufficient information, the authors were contacted via email (N=13) and given a two-week window to provide additional data or clarity (responses received, N=8 [38–45]). Detailed notes were recorded outlining the reasons for study inclusion/exclusion and the number of studies included and excluded at each stage.

### 2.4. Risk of bias

Study quality was appraised by at least two independent reviewers in duplicate (DH, GB, SYC), with any conflicts resolved by a third reviewer (TN). The Cochrane Risk of Bias 2 (RoB 2) was used to assess the risk of bias of the RCTs [46]. Reviewers determined the level of bias for each domain using the RoB 2 algorithms and is presented visually using Robvis [47]. Non-randomised designs were assessed using assessment tools generated by the National Institutes of Health (NIH) and National Heart, Lung and Blood Institute (NHLBI, Bethesda, MD). Pre-post studies were rated using the Quality Assessment Tool for Before-After (Pre-Post) Studies with No Control Group (12 items) and observational and cross-sectional studies were rated using the Quality Assessment Tool for Observational Cohort and Cross-Sectional Studies (14 items). Studies were subsequently classified as good, fair or poor quality using the guidance provided within each tool and is presented visually in online supplementary material.

### 2.5. Data synthesis and analysis

A variety of methods [i.e., indices of heart rate (HR), V̇O_2_ or ratings of perceived exertion (RPE)] have been utilised in the literature to establish, prescribe and regulate exercise intensity in the SCI-population, which creates complexity when classifying the intensity of exercise. Each intervention was classified as having prescribed either light, moderate, vigorous or supramaximal-intensity aerobic exercise, based on thresholds proposed by the American College of Sports Medicine (ACSM) [48] (S3). If a study reported a progression in intensity that spanned the moderate and vigorous-intensity categories (e.g., 60-65% V̇O_2peak_), it was classified as ‘moderate-to-vigorous’. If insufficient data were provided, studies were classified as ‘mixed-intensity/cannot determine’. Furthermore, where a study reported frequency of sessions or length of interventions as a range (e.g., 6-8 weeks), the midpoint was extracted and if a study reported duration as a range (e.g., 40-45 min), the greater value was extracted. Descriptions of adverse events in the included studies were also collated. These were categorised into the following subgroups: 1) bone, joint or muscular pain, 2) autonomic or cardiovascular function, 3) skin irritation or pressure sores, and 4) other.

Means ± SD were estimated from median and interquartile range (IQR) [49] or median and range [50], where required. Where CRF data was only presented in figures, data were extrapolated using Photoshop (Adobe Inc). To combine within-study subgroups and to estimate SD of the delta (Δ) change in CRF using correlation factors, we followed guidance from the Cochrane handbook [49]. Correlation factors were calculated for AV̇O_2peak_, RV̇O_2peak_ and PPO using studies that reported pre-post SD and SD of the Δ change using the following equation:

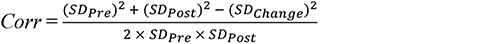

The specific correlation factors that were calculated for each study were averaged across each study design (S4) and applied in the following equation to calculate SD of the change for studies where these values were not reported:

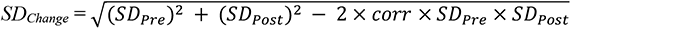

where *corr* represents the correlation coefficient.

Since AV̇O_2peak_, RV̇O_2peak_, and PPO are continuous variables, expressed using the same units across studies, we utilised weighted mean differences (WMDs) and 95% confidence intervals (CI) as summary statistics. A primary meta-analysis was carried out that included RCTs comparing Δ in CRF outcomes following an exercise intervention relative to control groups. A secondary, pooled meta-analysis describing Δ in CRF outcomes in response to prospective, well-characterised exercise interventions lasting >2 weeks (e.g., combining exercise intervention-arms from RCTs and pre-post studies) was also conducted to facilitate subgroup comparisons. Nine separate subgroup analyses were performed for this pooled, secondary meta-analysis to describe Δ in each CRF outcome with studies categorised into subgroups based on the following: 1) time since injury [(TSI), e.g., acute (<1-year), chronic (≥1-year)]; 2) neurological level of injury (e.g., tetraplegia, paraplegia); 3) injury severity [e.g., grading in accordance with the American Spinal Injury Association Impairment Scale (AIS): motor-complete (AIS A-B), motor-incomplete (AIS C-D)]; 4) exercise modality [e.g., aerobic volitional upper-body, resistance training, FES, gait training, behaviour change]; 5) relative exercise intensity (e.g., light, moderate, moderate-to-vigorous, vigorous, supramaximal); 6) method used to prescribe exercise intensity (e.g., V̇O_2_, HR, RPE, workload); 7) frequency of exercise sessions per week (<3, ≥3 to <5, ≥5); 8) exercise volume [e.g., SCI-specific exercise guidelines for fitness (40 - 89 min/wk) [21], SCI-specific exercise guidelines for cardiometabolic health (90 - 149 min/wk) [21], achieving general population exercise guidelines (≥150 min/wk) [23], and 9) length of intervention (≤6 weeks, >6 to ≤12 weeks, >12 weeks). Studies were also classified as ‘mixed’ or ‘not reported/cannot determine’. Three additional secondary meta-analyses were conducted for different trial designs: 1) comparing inactive vs active participants (e.g., cross-sectional cohort studies); 2) describing Δ in CRF outcomes with standard of care inpatient rehabilitation or free-living follow up (e.g., observational studies) and 3) head-to-head comparison of different exercise intensities (RCTs with exercise interventions comparing low or moderate vs. vigorous or supramaximal-intensity exercise). Meta-analyses were conducted in R (Version 3.5.1, R Foundation for Statistical Computing, Vienna, Austria) using the package metafor [51]. Statistical heterogeneity was assessed using the *I*^2^ and accompanying *p*-value from the chi-squared test. A random effects model was chosen to account for the variability in the true effect size across studies, given the expected between-study variability of different exercise intervention components and participant characteristics (by nature of SCI being a heterogeneous condition depending on the neurological level and severity of injury). Evidence for differences in effects between the subgroups in the secondary, pooled meta-analysis was explored by comparing effects in the subgroups and the corresponding *p-*values for interaction (metagen function from the R package meta [52]). Thresholds for statistically significant subgroup differences were adjusted for the number of subgroup comparisons and individual subgroup p-values were adjusted for multiple comparisons via the Bonferroni correction method. To assess the effect of potential outlier studies, leave-one-out analyses were performed with studies removed and pooled WMD recalculated. Sensitivity analyses were also conducted by comparing the WMDs of low and high risk of bias studies, as well as studies with and without imputed data (i.e., extracted from figures or where mean ± SD were calculated from median, IQR or range), to confirm the robustness of our findings. A further subgroup analysis was performed to compare Δ in CRF outcomes following exercise interventions that matched the CPET modality to the intervention modality (i.e., using an incremental arm-crank ergometry CPET to test the effects of an arm-crank exercise intervention). Potential small study effects in the dataset were assessed using funnel plots. Egger’s tests were also conducted in R when there was a minimum of 10 studies included in a meta-analysis [53]. Study design statistical power for both the summary effect size and a range of hypothetical effect sizes was calculated and visualised in firepower plots using the metameta R package, recently described by Quintana [54]. Plots were produced for the pre-post exercise interventions alone and for the RCT exercise interventions alone relative to controls (i.e., the primary meta-analysis studies) to facilitate comparisons between study designs. Data is visualised in R (see Github for scripts: https://github.com/jutzca/Exercise-and-fitness-in-SCI). A 2.7 mL/kg/min, and thus 1 metabolic equivalent in SCI (1 SCI-MET) [55], change in RV̇O_2peak_ was considered clinically meaningful.

To explore potential sources of heterogeneity, a random-effects meta-regression was performed using preselected moderator variables in Stata (Version 13, StataCorp LLC, College Station, TX, USA), adjusted for multiple testing. As per Cochrane recommendations [53], for each included covariate in the model a minimum of 10 studies were required. To achieve this, and to also overcome the issue of collinearity between moderators, some moderators were not included in the analysis. Moderators were selected *a priori*, based on their potential to influence CRF responses. Exercise intensity prescription was later added as a moderator in the meta-regression in light of a recent study challenging strategies for prescribing exercise intensity in individuals with SCI [56]. Moderators fell into two categories: model 1) participant/injury characteristics [continuous variables: age, TSI and baseline CRF; categorical variables: sex (n=male), neurological level of injury (n=PARA), severity (n=motor-complete)]; or model 2) intervention/study characteristics [continuous variables: exercise session duration, frequency, weekly exercise volume, intervention length; categorical variables: exercise modality, exercise intensity, method of exercise intensity prescription, and risk of bias classification]. Any potential covariates of the effect of AV̇O_2peak_, RV̇O_2peak_, and PPO with *p* ≤ 0.10 identified via univariate meta-regression were subsequently included in multivariate meta-regression modelling. The level of significance for multivariate meta-regression was set at *p* ≤ 0.10. Because meta-regression can result in inflated false-positive rates when heterogeneity is present, or when there are few studies, a permutation test described by Higgins and Thompson [57] was used to verify the significance of the predictors in the final model, whereby 10,000 permutations were generated.

### 2.6. Certainty on the body of the evidence assessment using the GRADE approach

The Grading of Recommendations Assessment, Development and Evaluation (GRADE) approach [58] was used to evaluate the certainty of the evidence for AV̇O_2peak_, RV̇O_2peak_ and PPO for the pooled, pre-post and RCT exercise interventions secondary meta-analysis. It was decided that the greater number of studies included in the pooled meta-analysis, in comparison to the primary meta-analysis consisting of RCTs only, would provide a more accurate assessment of the current body of evidence. Two authors (DH, SYC) independently assessed the certainty of evidence for each outcome, with any conflicts resolved by the corresponding author (TN). The certainty of the evidence was graded from ‘High’ to ‘Moderate’, ‘Low’ or ‘Very Low’. GRADE certainty in the evidence was downgraded if one or more of the following criteria were present: 1) risk of bias, 2) inconsistency in the results for a given outcome, 3) indirectness, 4) imprecision, and 5) small study effects.

## 3. RESULTS

The initial database search identified 14,248 articles after removal of duplicates. A further 12,322 studies were removed following the screening of titles and abstracts. The remaining 1,926 articles were selected for full-text review based on inclusion and exclusion criteria (S2). Of these, a total of 120 eligible studies, across each specific study design (RCT = 29, pre-post = 67, observational = 6, cross-sectional cohort = 11, cross-sectional association = 7), were included in this review. Twenty-nine RCTs were included in the primary meta-analysis. Ninety-six studies, comprising the RCTs and pre-post studies (total = 117 independent interventions), were included in the secondary, pooled meta-analysis. Summaries of the pooled cohorts and descriptions of the individual studies included within each additional secondary meta-analysis are provided as supplementary material.

### 3.1. Primary meta-analysis: RCT exercise intervention vs control groups

Twenty-two RCTs assessed changes in CRF outcomes between exercise intervention (n=283 participants) and control (n=252 participants) groups. Participant demographics, injury characteristics, exercise intervention parameters and changes in CRF outcomes for each RCT can be found in Figure 2. A summary of the pooled cohort characteristics, summary statistics for the meta-analysis and forest plots for each CRF outcome are presented in supplementary material (S5). The meta-analysis of RCTs revealed a significantly higher AV̇O_2peak_ [0.16 (0.07, 0.25) L/min, *p*<0.001], RV̇O_2peak_ [2.9 (1.8, 3.9) mL/kg/min, *p*<0.001], and PPO [9 (5, 14) W, *p*<0.001] following exercise interventions relative to controls. There was significant heterogeneity present across all CRF outcomes (*p*<0.001), with I^2^ values of 87%, 93% and 88% for AV̇O_2peak_, RV̇O_2peak_, and PPO, respectively.

**Figure 1.**
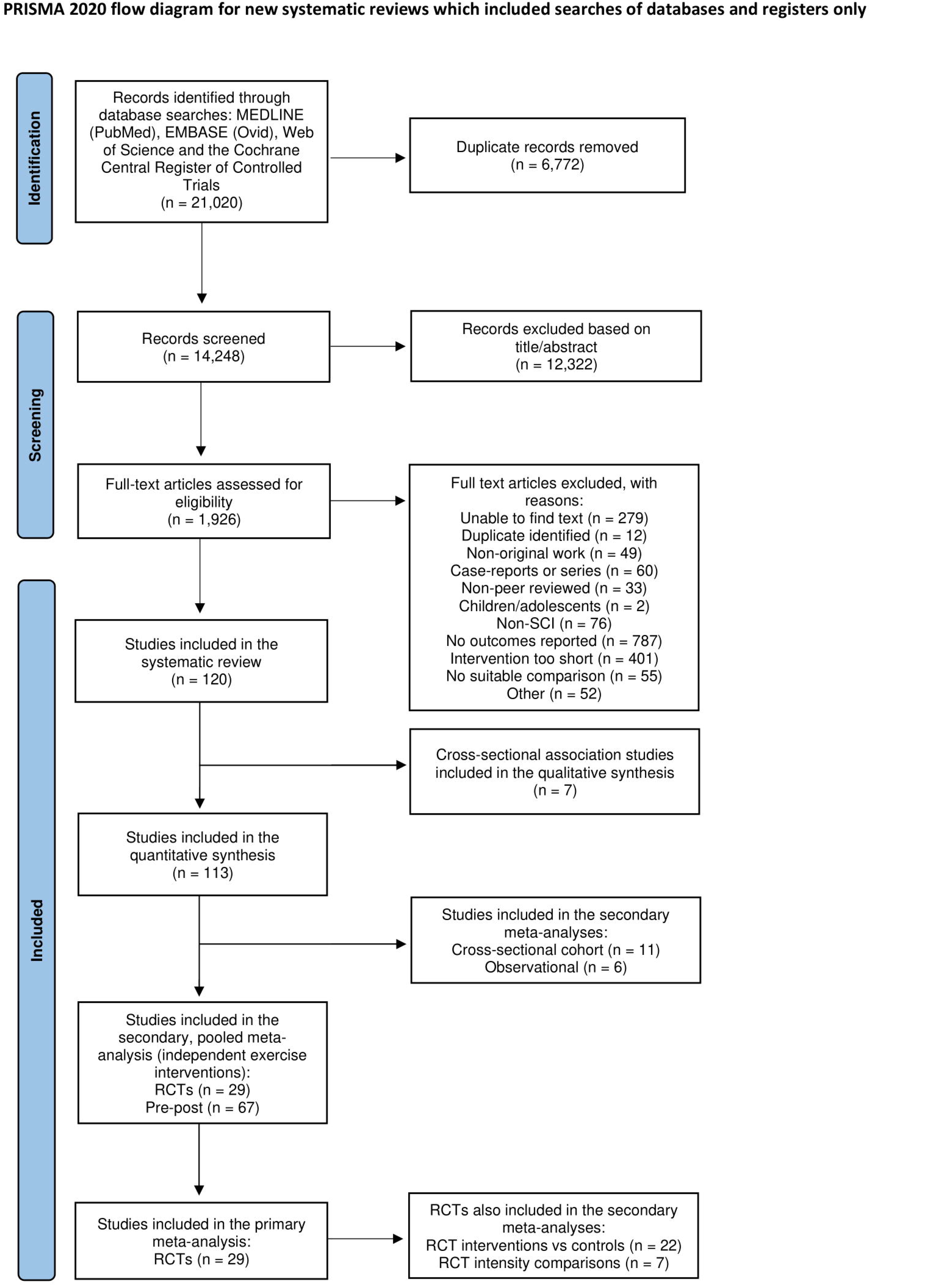
PRISMA flow diagram. Abbreviation: PRISMA, Preferred Reporting Items for Systematic Reviews and Meta-Analyses.

**Figure 2.**
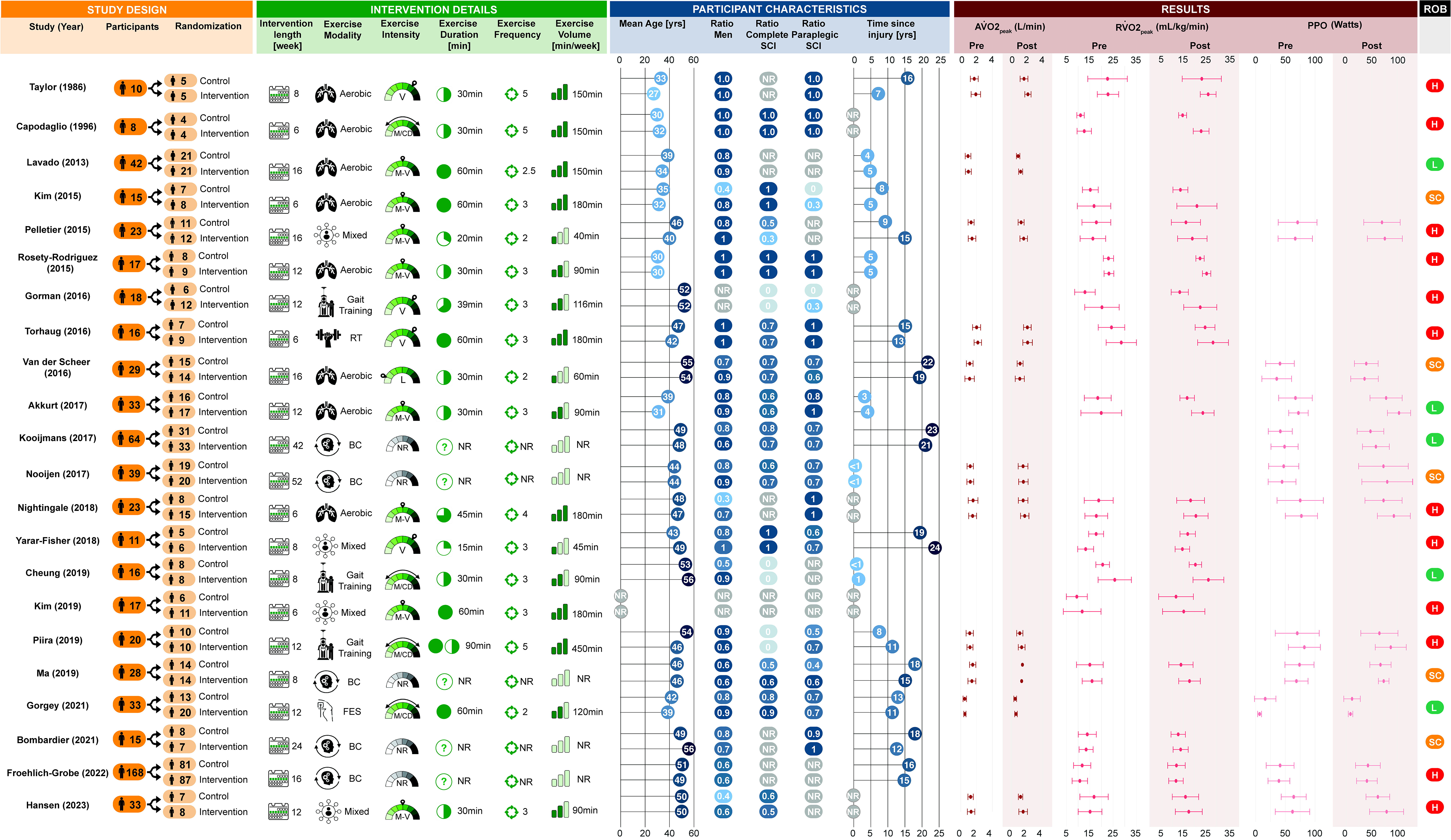
Summary of the study design, exercise intervention parameters, participant demographics/injury characteristics and changes in cardiorespiratory fitness outcomes for each randomised controlled trial study included in the primary meta-analysis. Studies were assessed for risk of bias (ROB) and rated as either low (L), some concerns (SC) or high (H) risk. AV̇O_2peak_, absolute peak oxygen uptake; BC, behaviour change; FES, functional electrical stimulation; L, light-intensity; M, moderate-intensity; M/CD, mixed/cannot determine; M-V, moderate-to-vigorous-intensity; NR, not reported; PPO, peak power output; RT, resistance training; RV̇O_2peak_, relative peak oxygen uptake; SCI, spinal cord injury; V, vigorous-intensity.

### 3.2. Secondary, pooled meta-analysis: Effects of prescribed, prospective exercise interventions from pre-post and RCT studies

CRF responses were pooled across 96 studies, comprising 117 exercise interventions in total, taken from 81 pre-post exercise interventions and 36 independent exercise intervention arms from RCTs. Some studies included multiple exercise intervention arms/phases, hence the greater total number of exercise interventions than studies. A summary of the demographic/injury characteristics and intervention parameters for the pooled cohort (i.e., all 117 interventions) included in the analyses for AV̇O_2peak_, RV̇O_2peak,_ and PPO are presented in supplementary material (S6). Participant demographics and exercise intervention parameters across the total 117 exercise interventions are summarised below. Summaries of the interventions for each CRF outcome, along with details of the specific studies is presented in online supplementary material (S6).

#### 3.2.1. Participants

Across the 117 exercise interventions, there were a total of 1,331 participants. Most interventions included both males and females (63% of studies), where females made up between 6-80% of the mixed cohorts. There were no female-only cohorts. Mean age ranged between 23 to 58 years and the majority of participants had chronic injuries (64% >1-year), with mean TSI ranging between 56 days to 24 years. Sixty-three interventions included a mixed cohort of paraplegia and tetraplegia, of which individuals with paraplegia made up between 10-88% of the mixed cohorts. Four interventions recruited individuals with tetraplegia-only, 39 paraplegia-only, and 11 did not specify. Participants across all AIS groups were included, of which 41 interventions were motor-complete-only, 19 were motor-incomplete-only, and 20 did not report. Thirty-seven interventions recruited both motor-complete and incomplete individuals, of which 33% were motor-incomplete. Weighted mean AV̇O_2peak_ and RV̇O_2peak_ at baseline was 1.28 (0.51-3.50) L/min and 17.8 (7.3-36.9) mL/kg/min, respectively, and PPO was 49 (0-168) W.

#### 3.2.2. Exercise intervention characteristics

Length of interventions ranged from 4 to 52 weeks, and whilst most studies reported a specific, predetermined intervention length, some reported a range [59–61], a total or targeted number of sessions [60,62–66], or provided an average [65,67,68]. Exercise sessions were completed between two to seven times per week. Eleven studies reported a range (e.g., “two to three sessions”) or maximum frequency (e.g., “up to three sessions/week”) [60,63,66,69–76], and frequency was either not reported or could not be determined in seven studies [39,77–82]. The remainder reported an exact frequency (e.g., three sessions per week). The duration of exercise sessions ranged from 5 to 90 minutes, with five studies reporting a range (e.g., 20-30 min) [60,76,83–85] and six studies reporting a progression to a target duration [63,86–90]. Duration was not reported or could not be determined in 15 studies. Based on current exercise guidelines, 23 interventions prescribed exercise within the SCI-specific exercise guidelines for fitness (40-89 min/week), 45 interventions targeted the SCI-specific exercise guidelines for cardiometabolic health (90-149 min/week), and 30 were greater than general population exercise guidelines (≥150 min/week).

Forty-two interventions utilised aerobic upper-body exercise, 7 upper-body resistance training, 22 FES, 15 gait training, 5 behaviour change, and 26 mixed/multimodal interventions. Following the ACSM thresholds, one intervention prescribed light-intensity (1%), 17 prescribed moderate-intensity (14%), 35 prescribed moderate-to-vigorous-intensity (30%), 26 prescribed vigorous-intensity (22%), and 2 prescribed supramaximal-intensity exercise (2%). Intensity could not be determined from 36 interventions (31%). With regards to exercise intensity prescription methods, 35 interventions used HR, regulated either via HR_peak_ (%HR_peak_, i.e., determined via a CPET; N=10), HR_max_ (%HR_max_, i.e., age-predicted; N=10), or HR reserve (%HRR; N=15). Fourteen interventions established intensity using V̇O_2peak_ (%V̇O_2peak_; N=13) or V̇O_2_ reserve (%V̇O_2reserve_; N=1) calculated from the pre-intervention CPET. Fourteen interventions utilised RPE, using either the Borg CR10 scale (N=7) or the Borg 6-20 scale (N=7). Workload was used to prescribe intensity in 11 interventions, via a percentage of PPO (%PPO; N=6), one repetition maximum (%1RM; N=4), or maximal tolerated power (%MTP; N=1). Forty-three interventions either used a mixture of prescription methods or intensity could not be classified.

#### 3.2.3. Adverse events

Adverse events were described in 18 interventions, comprising at least 49/1,331 (3.7%) participants (S7). These events were related to: 1) bone, joint or muscular pain (n=10 participants), 2) autonomic or cardiovascular function (n=8 participants), 3) skin irritation or pressure sores (n=18 participants), and 4) other events including anxiety, nausea, dizziness and issues with testing equipment (n=3 participants). The following adverse events were reported in four other pre-post studies but could not be categorised as above. Beillot et al. [77] stated that participants experienced *“spontaneous fractures of lower limbs, occurrence of a syringomyelia and pressure sores at the foot and ankle”* (n=10), but did not define the number of participants who sustained each event. Likewise, Janssen and Pringle [70] reported *“lightheadedness in some subjects”* and Gibbons et al. [91] stated that *“a number of participants showed some level of autonomic dysreflexia during the FES response test”*, but both studies did not quantify further. Vestergaard et al. [92] reported adverse events relating to “slight non-persisting pain in neck (n=1), arms and shoulders (n=4) during and between training sessions, dizziness that disappeared after 5-min (n=1), feeling tired in the head/dizziness that disappeared after training with no other signs of autonomic hyperreflexia (n=2), increased spasms (n=2), and vomiting just after training (n=2)”. However, as only seven participants completed the intervention, it cannot be determined whether these events were reported for one or multiple participants.

#### 3.2.4. Change in CRF outcomes

Seventy-four exercise interventions assessed the change in AV̇O_2peak_, revealing a significant increase of 0.22 [0.17, 0.26] L/min (*p*<0.001). There were no significant subgroup differences for any of the nine subgroup analyses. Seventy-nine exercise interventions assessed the change in RV̇O_2peak_, revealing a significant increase of 2.8 [2.3, 3.3] mL/kg/min (*p*<0.001). There were significant subgroup differences for exercise modality (*p*=0.002) and length of intervention (*p*=0.01), but there were no other differences (Figure 3). Sixty-five exercise interventions assessed the change in PPO, revealing a significant increase of 11 [9, 13] W (*p*<0.001). There were significant subgroup differences for TSI (*p*<0.001), neurological level of injury (*p*<0.001), exercise modality (*p*=0.002), relative exercise intensity (*p*=0.001), method of exercise intensity prescription (*p*<0.001), and frequency (*p*<0.001) (Figure 3). There was significant heterogeneity present across all CRF outcomes (*p*<0.001), with I^2^ values of 72%, 53% and 78% for AV̇O_2peak_, RV̇O_2peak_, and PPO, respectively. Forest plots for each subgroup analysis are presented in supplementary material (S6).

**Figure 3.**
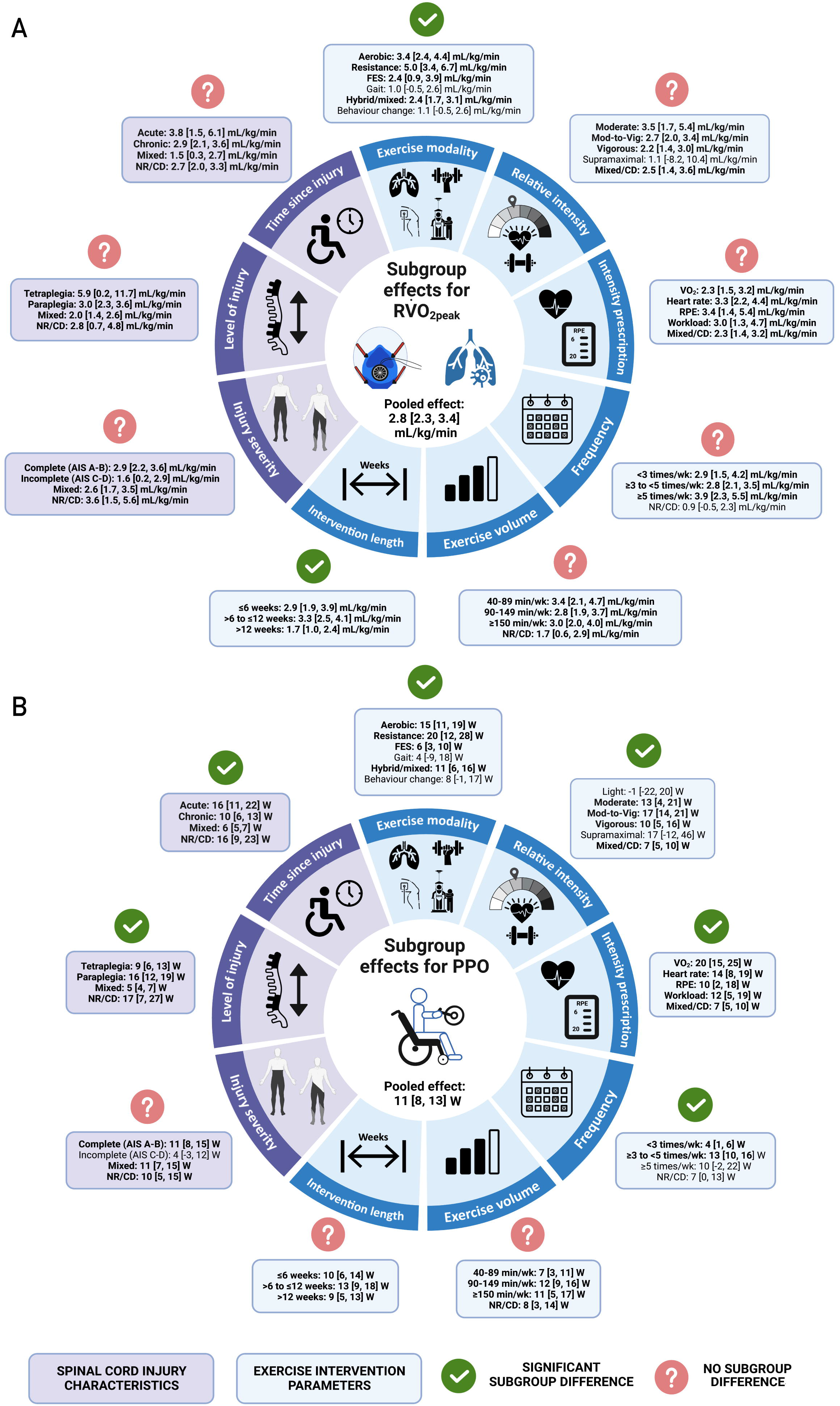
Overview of the subgroup effects for relative peak oxygen uptake (A; RV̇O_2peak_) and peak power output (B; PPO) for pooled pre-post and randomised-controlled trial exercise interventions based on injury-specific characteristics (purple) and exercise intervention parameters (blue). Individual subgroup effects are highlighted in bold. Differences within each subgroup category are represented by green ticks whereas subgroup categories without significant differences are represented by red question marks. AIS, American Spinal Injury Association Impairment Scale; FES, functional electrical stimulation; Mod-to-Vig, moderate-to-vigorous; NR/CD, not reported/cannot determine; RPE, rating of perceived exertion; V̇O_2_, volume of oxygen consumption W, watts.

An additional subgroup analysis grouped interventions into those that matched the CPET modality to the exercise intervention and those that did not. Following the adjustment for subgroup comparisons, there were no significant differences in AV̇O_2peak_, RV̇O_2peak_ or PPO (S9). However, there were trends for a greater AV̇O_2peak_ (*p*=0.05) and RV̇O_2peak_ (*p*=0.06) in studies with matched CPET and intervention modalities in comparison to interventions without matched modalities. A sub-analysis for gait training interventions alone also revealed no subgroup differences between studies that used upper-body or treadmill CPETs to determine CRF outcomes (S10).

#### 3.2.5. Sensitivity analyses

Leave-one-out analyses identified outliers for AV̇O_2peak_ [42][61,93]. A sensitivity analysis for risk of bias revealed no differences in the pooled effects for low and high risk of bias studies (S8). A sensitivity analysis comparing studies with imputed data via conversion of medians (IQR), extrapolated data from figures, and studies without imputed data revealed no differences in the pooled effects for any outcome. (S8).

#### 3.2.6. Study-design statistical power considerations

Median statistical power and observed effect sizes for each CRF outcome and study design (i.e., pre-post studies compared to RCT interventions relative to controls) are reported in Figure 4. Across each CRF outcome, median statistical power was higher for the RCTs included in the primary meta-analysis in comparison to the pre-post studies. In general, this indicates that the RCT studies were designed to reliably detect a wider range of effect sizes in comparison to the pre-post studies.

**Figure 4.**
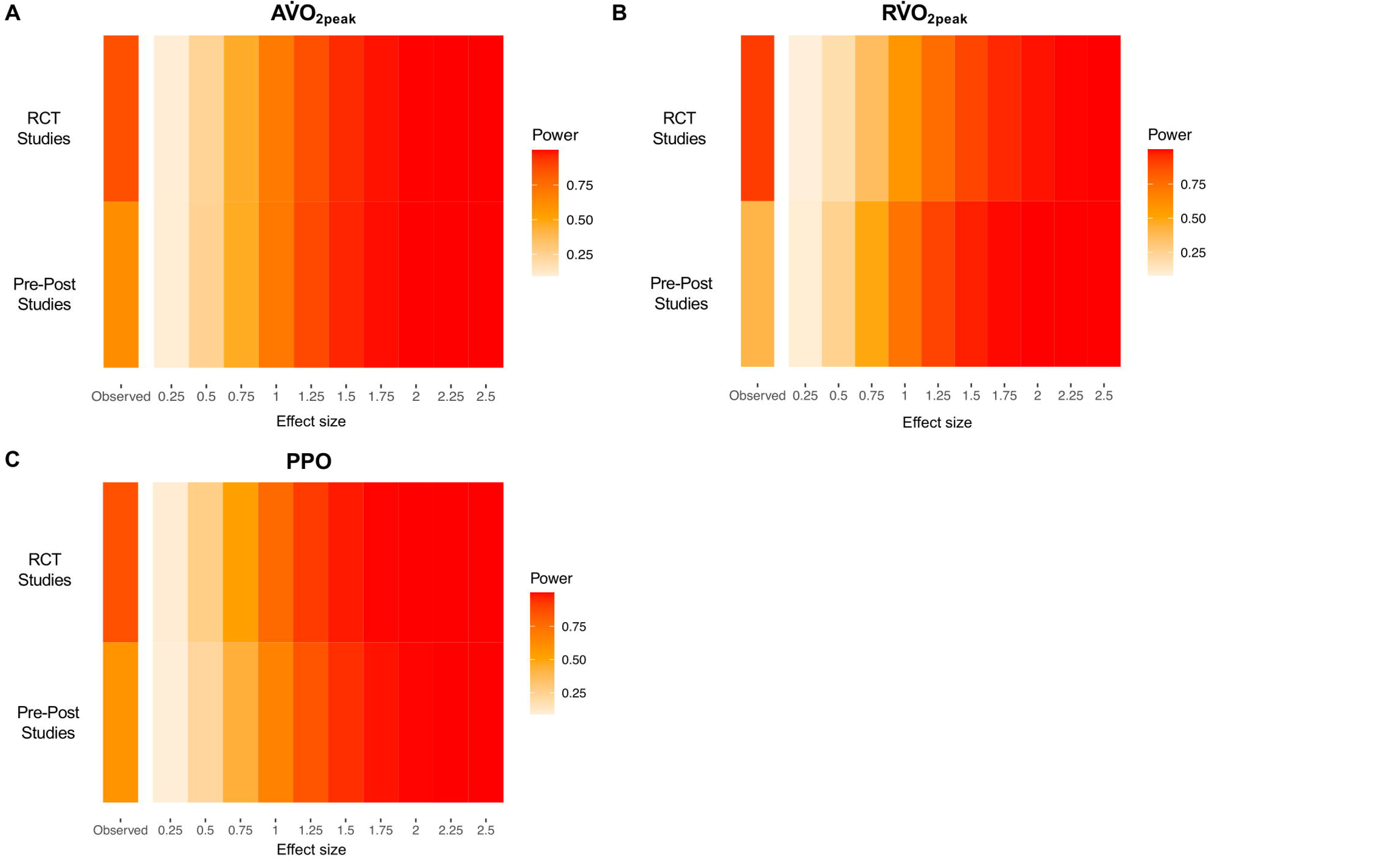
Firepower plots visualising study-level statistical power for a range of effect sizes between meta-analyses for pre-post studies alone in comparison to RCT studies included in the primary meta-analysis (i.e., exercise interventions relative to control groups) for each cardiorespiratory fitness outcome. Hedges’ g effect sizes were calculated for each meta-analysis and used as the observed effect size, as described by Quintana [54]. (A) Meta-analysis of pre-post studies describing changes in absolute peak oxygen uptake (AV̇O_2peak_) had a median power of 62% for detecting an effect size of 0.90, whereas the RCTs had a median power of 86% for detecting an effect size of 1.25. (B) Meta-analysis of pre-post studies describing changes in relative peak oxygen uptake (RV̇O_2peak_) had a median power of 41% for detecting an effect size of 0.67, whereas the RCTs had a median power of 91% for detecting an effect size of 1.57. (C) Meta-analysis of pre-post studies describing changes in peak power output (PPO) had a median power of 58% for detecting an effect size of 0.91, whereas the RCTs had a median power of 86% for detecting an effect size of 1.13.

#### 3.2.7. Meta-regression

##### Model 1 - Participant and injury characteristics

Exercise interventions with a greater mean age of participants were associated with smaller changes in the effect estimates for AV̇O_2peak_ (*p*=0.08) and RV̇O_2peak_ (*p*=0.01). The coefficients indicate that for every one-year increase in mean age of participants in an exercise intervention, the effect on AV̇O_2peak_ and RV̇O_2peak_ decreases on average by 0.003 L/min and 0.041 mL/kg/min, respectively, holding all other covariates constant (Table 1). There were no associations between the other moderator variables included in this model and AV̇O_2peak_ or RV̇O_2peak_. Whilst there were no significant associations between PPO and the other moderator variables (Table 1), there was a trend for an association between PPO and TSI (p=0.18). The coefficient for TSI indicates that for every additional year since injury, the effect on PPO following an exercise intervention decreases by 1.5W on average (Table 1).

**Table 1.**
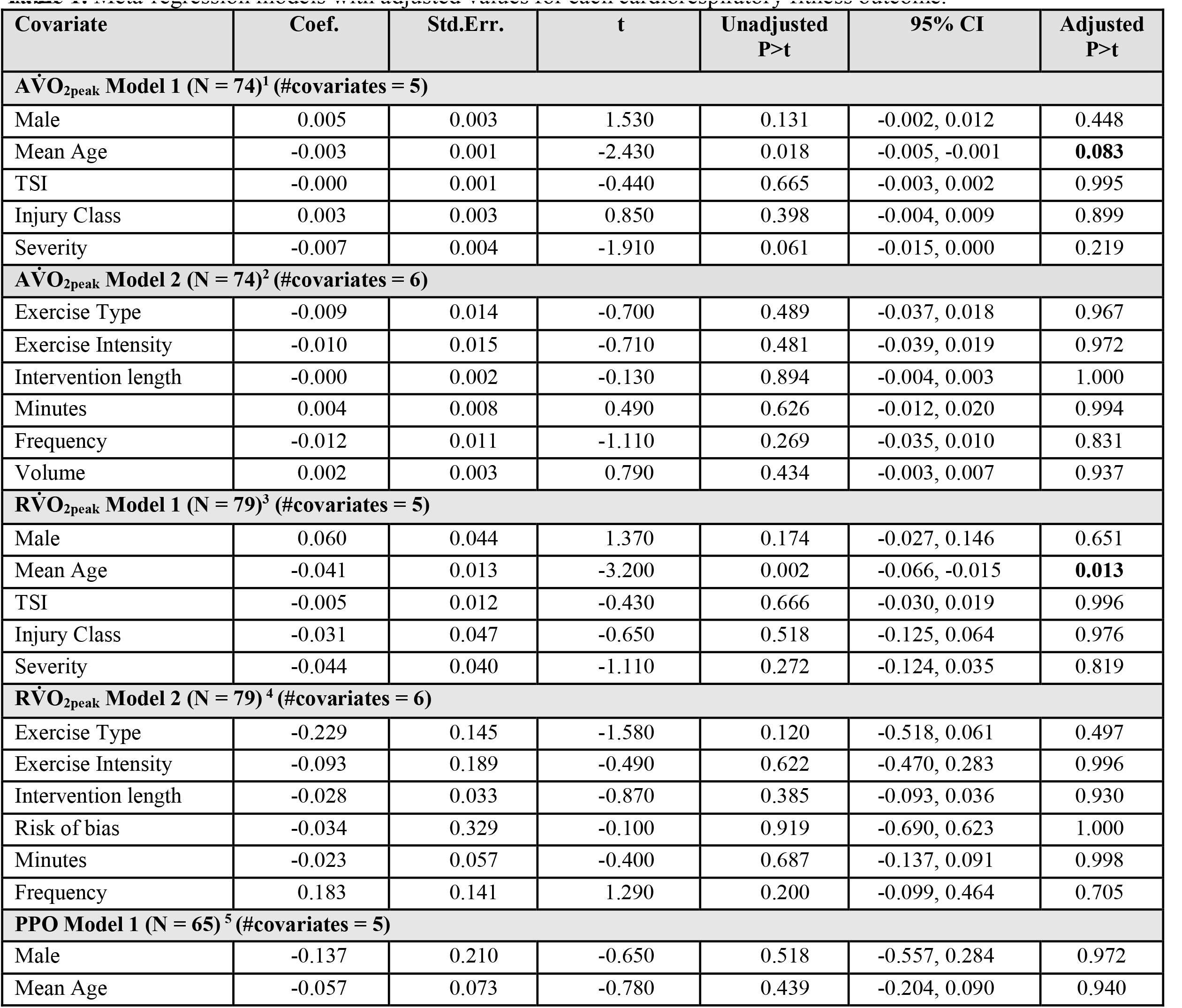
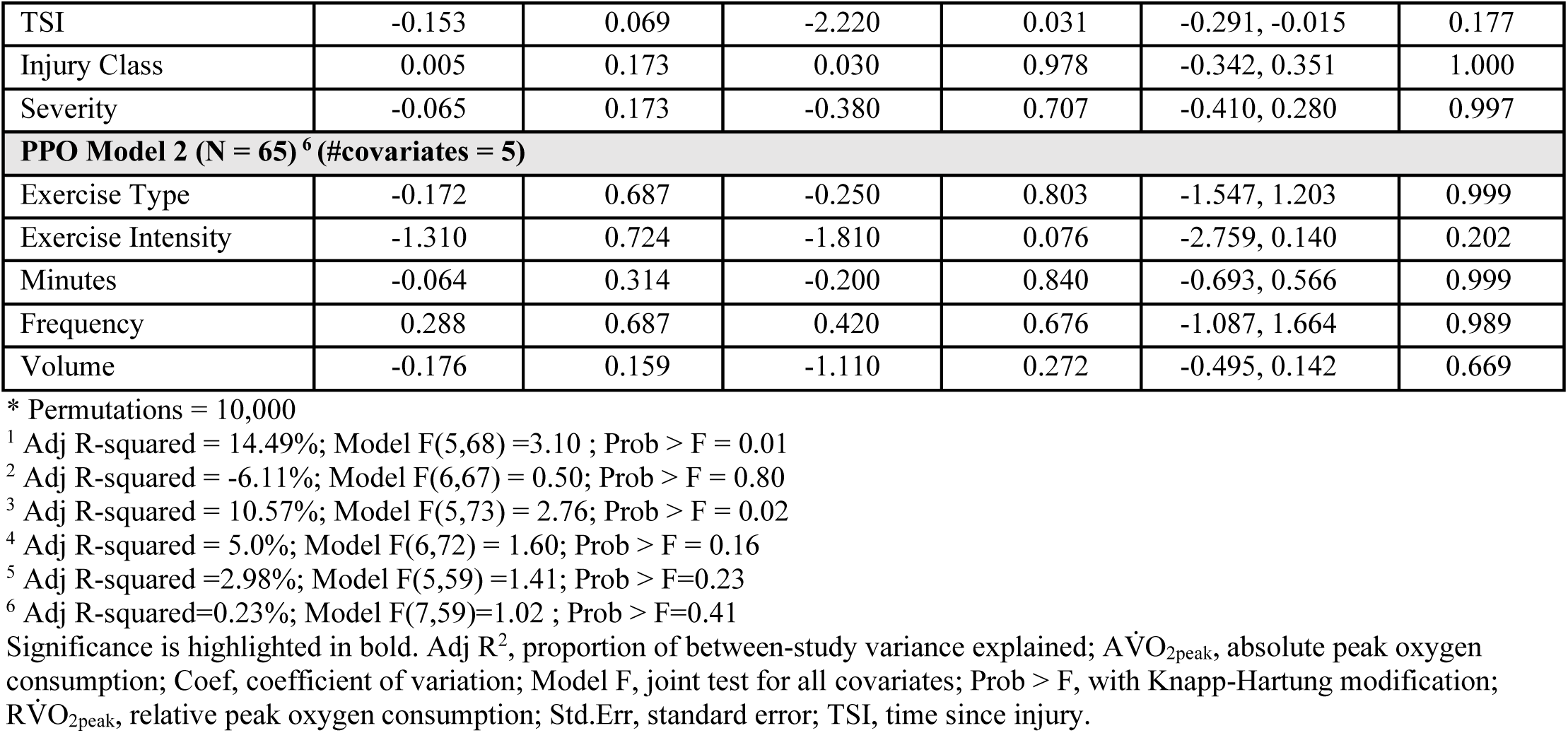
Meta-regression models with adjusted values for each cardiorespiratory fitness outcome.

##### Model 2 - Exercise intervention and study characteristics

There were no significant associations between the exercise intervention and study characteristics included in model 2 for AV̇O_2peak,_ RV̇O_2peak_, or PPO (Table 1).

#### 3.2.8. Small study effects

Egger’s tests for funnel plot asymmetry were not statistically significant for AV̇O_2peak_ (Z = -1.20, *p* = 0.23), RV̇O_2peak_ (Z = -0.44, *p* = 0.66), or PPO (Z = 0.76, *p* = 0.45). Funnel plots are provided in supplementary material (S6).

### 3.3. Secondary Meta-Analyses

#### 3.3.1. Cross-sectional studies

Eleven studies included cross-sectional data comparing CRF outcomes in active (n=182 participants) vs. inactive (n=134 participants) individuals with SCI. Inactive participants were mainly classified as sedentary, whereas active participants varied from recreationally active wheelchair sport players to paralympic athletes A meta-analysis of cross-sectional cohort studies revealed significantly (*p*<0.001) higher AV̇O_2peak_ [0.55 (0.43, 0.67) L/min], RV̇O_2peak_, [9.1 (7.0, 11.2) mL/kg/min] and PPO [38 (32, 45) W] in active compared to inactive individuals with SCI (S11). Given the significant heterogeneity in RV̇O_2peak_, a sensitivity analysis was conducted to compare inactive individuals with either ‘active’ or ‘elite athletes’. There was a significantly higher RV̇O_2peak_ [6.4 (4.7, 8.1) mL/kg/min, *p*<0.001] in ‘active’ compared to inactive individuals, but an even higher RV̇O_2peak_ [11.2 (9.6, 12.9) mL/kg/min, *p*<0.001] in ‘elite athletes’ compared to inactive.

Seven studies (n=581 participants) included cross-sectional data and assessed associations between habitual physical activity level (as a continuous variable) and CRF outcomes (S12). Six studies assessed physical activity exposure using self-report methods [38,40,94–97], whereas one study used a validated research-grade wearable device [98]. The measurement period used to capture physical activity dimensions ranged from 3 to 7 days. There was considerable variability across studies with regards to the physical activity dimensions captured: hours per week of exercise/sport, minutes per day or week of mild, moderate, heavy-intensity for the subcategories of leisure time physical activity (LTPA), lifestyle or household activity or cumulative activity. Collectively, data indicates significant positive correlations of a larger magnitude between CRF/PPO outcomes and the volume of sport, exercise or LTPA rather than household activity. The only study to use a validated wearable device indicated that participants performing ≥150 min/week of moderate-to-vigorous physical activity (MVPA) had a significantly higher CRF relative to a low activity group (performing <40 min/wk). Whereas, there was no significant difference in CRF between the low activity group and participants achieving the SCI-specific exercise guidelines (40 - 149 min/week) [98]. These data have been replicated in a recent study, which compared CRF outcomes for participants achieving either the SCI-specific exercise guidelines for fitness (40 - 89 min/week) and cardiometabolic health (90 - 149 min/week) [21] with the Exercise and Sport Science Australia (ESSA) position statement recommending that individuals with SCI achieve ≥150 min/week [24]. There were no differences in CRF outcomes between individuals classified as inactive and those meeting the current SCI-specific exercise guidelines, yet there were significant differences across all CRF outcomes between inactive individuals and those achieving the ESSA guidelines. Furthermore, AV̇O_2peak_ and PPO were greater for individuals meeting the ESSA guidelines, relative to the current SCI-specific exercise guidelines. Across studies, significant, positive correlations were reported for the amount of moderate-to-vigorous LTPA or cumulative activity with CRF/PPO outcomes, which was not the case for mild or light-intensity activity.

#### 3.3.2. Observational inpatient rehabilitation or community free-living studies

Six studies (n=354 participants) included observational longitudinal data and assessed changes in CRF outcomes following either standard of care inpatient rehabilitation [99–101] or a period of community free-living [99,102,103]. Whilst one study described a training programme for elite-level wheelchair rugby players, it could not be determined whether players adhered to the pre-specified training programme throughout the season and therefore the study was categorised as a community free-living observational study in this review [104]. The duration between assessments for standard of care varied, ranging from 5 to 28 weeks, whereas the follow-up period for community observations ranged from 30 weeks to 2.9 years. Reporting on the therapies used within standard of care was poor and only one study included a measurement of physical activity during the community-based free-living follow-up (self-reported mean sport activity) [102]. There were significant improvements following standard of care, but not following community-based free-living, in AV̇O_2peak_ [0.12 (0.07, 0.17) L/min, *p*<0.001 vs. 0.09 (0.00, 0.19) L/min, *p*=0.06] and RV̇O_2peak_ [2.1 (1.0, 3.2) mL/kg/min, *p*<0.001 vs -0.5 (-2.5, 1.5) mL/kg/min, *p*=0.64] (S13). Significant improvements in PPO were identified following both standard of care [6 (3, 9) W, *p*<0.001] and community-based free-living [7 (2, 12) W, *p*=0.006] (S13).

#### 3.3.3. RCT exercise intensity comparisons

Seven RCTs compared changes in CRF outcomes between low or moderate (n=52 participants) and vigorous or supramaximal (n=51 participants) exercise intensity groups. These studies utilised upper-body aerobic exercise and gait training. A meta-analysis revealed no significant differences between low or moderate and vigorous or supramaximal intensity in AV̇O_2peak_ (*p*=0.67), RV̇O_2peak_ (*p*=0.88) or PPO (*p*=0.62) (S14). There were also no significant subgroup differences between studies that matched

exercise volume between intensity groups and those that did not.

### 3.4. Risk of Bias

Full risk of bias assessments for pre-post and RCT interventions can be found in supplementary material (S5, S6, S14). Twenty-six pre-post studies were rated as having good, 27 as having fair, and 14 as having poor methodological quality. Six RCTs were rated as having a low risk of bias, 8 as having some concerns, and 15 as having a high risk of bias. The most common domains in the RCTs with either some concerns or high risk were ‘bias in the measurement of the outcome’ and ‘bias in selection of the reported result’. Reporting was inadequate in many of the included studies, which made the assessment of risk of bias challenging. Notably, reporting of blinding, eligibility or selection criteria, as well as the enrolment of participants (i.e., a lack of CONSORT flow diagrams) was poor. Individual risk of bias assessments for each study design are provided in supplementary material (S5, S6, S11-14).

### 3.5. Evidence appraisal using GRADE

Overall, the GRADE assessment revealed a Moderate certainty in the body of evidence for improvements in RV̇O_2peak_, but a ‘Low’ certainty in the body of evidence for improvements in AV̇O_2peak_ and PPO (Table 2). The certainty rating for AV̇O_2peak_ was downgraded due to imprecision and a lack of high-quality study designs, whereas RV̇O_2peak_ was downgraded as a result of imprecision. The certainty rating for PPO was downgraded due to imprecision and inconsistency, resulting from considerable heterogeneity in the included exercise interventions.

**Table 2.**
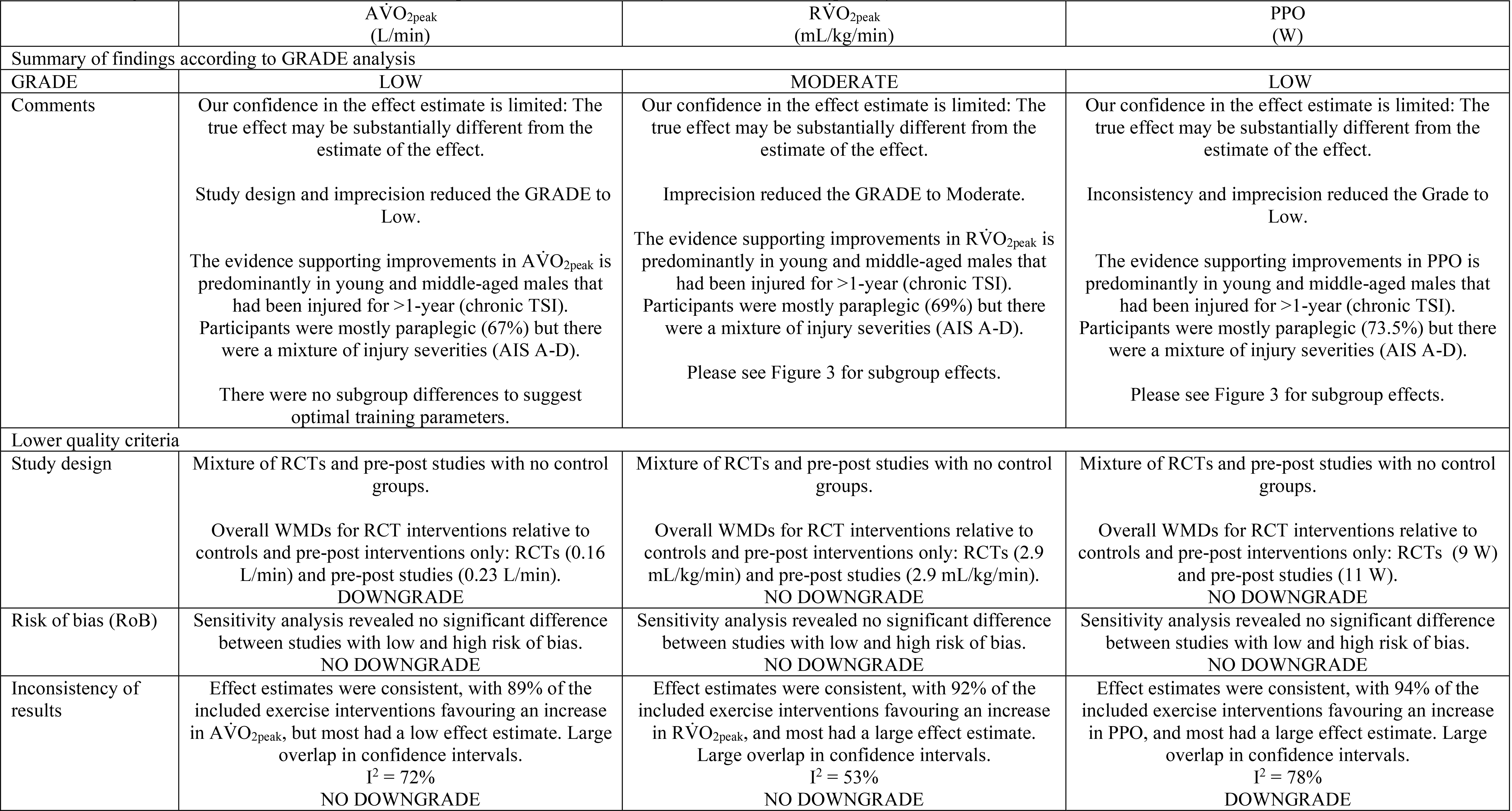
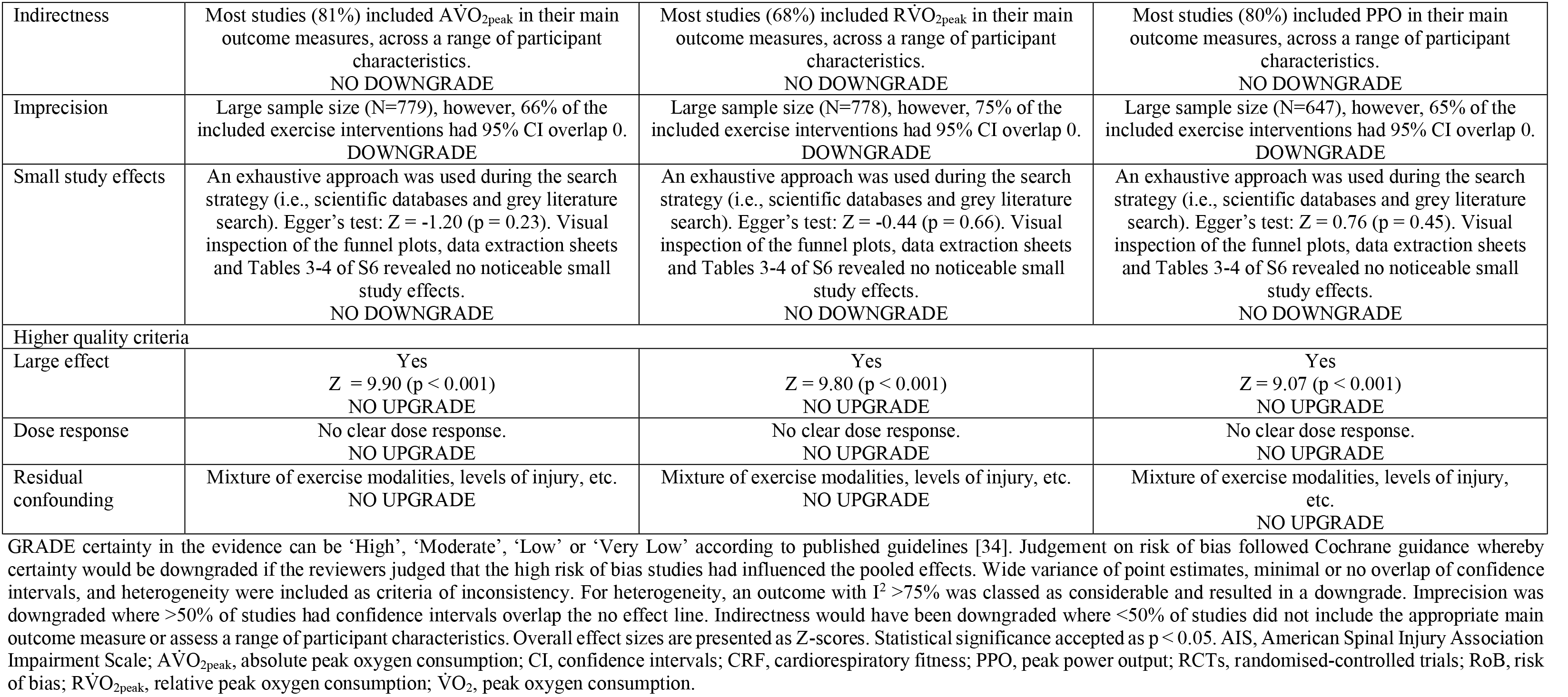
Grading of recommendations assessment, development and evaluation analysis for each cardiorespiratory fitness outcome.

## 4. DISCUSSION

This review provides a large evidence-based summary and appraisal on the effects of prescribed and prospective exercise interventions >2 weeks on CRF in individuals with SCI. The results from the primary meta-analysis of RCTs support the role of exercise in improving CRF in this population by 0.16 L/min and 9W in AV̇O_2peak_ and PPO, respectively. The primary meta-analysis also indicates a clinically meaningful change in RV̇O_2peak_ of 2.9 mL/kg/min. Subgroup analyses in the secondary, pooled meta-analysis revealed no effects of injury characteristics or exercise intervention parameters on AV̇O_2peak_, but an effect of exercise modality and length of intervention on RV̇O_2peak_. There were also significant subgroup differences for PPO based on TSI, neurological level of injury, exercise modality, exercise intensity, method of exercise intensity prescription, and frequency of sessions per week. The GRADE assessment including RCTs and pre-post intervention studies revealed ‘Moderate’ certainty in the evidence for improvements in RV̇O_2peak_, yet ‘Low’ certainty in the evidence for significant improvements in AV̇O_2peak_ and PPO.

Following exercise interventions, V̇O_2peak_ improves in individuals with both acute and chronic SCI. However, this review highlights the need for more exercise interventions in the acute phase post-SCI. Indeed, a recent review by Van der Scheer et al. [35] rated the confidence in the evidence base for exercise in acute SCI as ‘Very Low’, and called for more RCTs to control for the deteriorations in fitness and health occurring almost immediately following SCI. With regards to PPO in the current review, the meta-regression revealed a trend for an association between TSI and changes in PPO, with individuals with long-term injuries exhibiting smaller changes in PPO in comparison to those with relatively newer injuries. The subgroup analysis based on TSI also revealed that individuals with acute SCI exhibit a greater change than individuals with chronic SCI. This may be due to exercise being delivered in combination with standard of care inpatient rehabilitation for individuals with acute injuries, representing an additive effect. Indeed, the secondary meta-analysis with longitudinal observational studies indicates a 6W improvement in PPO with standard of care inpatient rehabilitation alone during the subacute period. This could be influenced by spontaneous motor recovery in the first few months following SCI [105]. Or alternatively, this subacute period may provide a familiarisation effect to novel modalities of exercise that patients were previously unaccustomed to and/or provides a notable stimulus to naive upper-body musculature following a period of bed rest and deconditioning, possibly representing a regression to the mean artefact. Ultimately, more rigorous RCTs are required in the subacute phase post-SCI that compare standard of care versus standard of care plus a specific exercise intervention to truly quantify the independent effects of a prescribed exercise intervention in the inpatient setting.

Exercise results in improved V̇O_2peak_ regardless of the neurological level of injury. In particular, this review reveals a pooled improvement of 5.9 mL/kg/min in studies that included only individuals with tetraplegia (N=3 interventions). For comparison, there is a considerably larger evidence-base for studies including only individuals with paraplegia (N=31 interventions). A recent systematic review suggested that aerobic exercise results in minimal returns on investment in individuals with tetraplegia, with V̇O_2peak_ improving on average only 9% following 10-37 weeks of training [106]. However, their review excluded studies with a sample size <10. Consequently, the Dicarlo study [93], which reported a 94% increase in RV̇O_2peak_ was excluded from their analysis. Whilst the inclusion of this study in the current analysis may have augmented the overall effect, as identified by our leave-one-out analysis, there was nothing untoward in this study to suggest a reason for this exaggerated response. Therefore, our findings indicate that exercise improves CRF in individuals with tetraplegia and that the magnitude of change is not significantly different to individuals with paraplegia.

However, the pooled meta-analysis highlights that individuals with paraplegia (16W) are likely to accrue greater absolute changes in PPO than those with tetraplegia (9W). Typically, higher neurological levels of injury result in a loss of trunk control, motor impairments in the upper-limbs and reduced mechanical efficiency, compared to lower levels of injury [107,108]. Therefore, individuals with tetraplegia may not have the physical or motor capacity to adapt as effectively as individuals with paraplegia, and thus could experience a ceiling effect with training. Indeed, a recent study identified lesion level as a significant predictor of PPO in a group of handcyclists with SCI [109]. To account for baseline motor function differences between individuals with tetraplegia and paraplegia, we determined relative percentage change for studies that included upper-body aerobic exercise interventions only. The relative percentage change was similar between neurological level of injury classifications: 46% tetraplegia (N=1 interventions) vs. 53% paraplegia (N=9 interventions). While only one tetraplegia-only intervention was included in this subgroup analysis [110], normalising for baseline values seems to indicate similar relative magnitudes of change in PPO.

Williams et al. [111] recently demonstrated that individuals with a lower level of injury (<T6) significantly improved PPO compared to individuals with a higher level of injury (≥T6), suggesting a potential role of disrupted cardiovascular control in mediating changes in PPO [112,113]. Whilst methods for ameliorating the reduction in sympathetic cardiovascular control typically associated with injuries ≥T6 have been investigated (e.g., abdominal binding [114], lower-body positive pressure [115], and midodrine [116]), the evidence for an improved CRF is still mixed. A recent case-report has indicated that epidural spinal cord stimulation (SCS) can safely and effectively restore cardiovascular control and improve CRF [117]. With an explosion in SCS studies over the last few years [118], particularly including transcutaneous SCS, the pairing of exercise with novel and non-invasive neuromodulatory approaches will likely continue to receive considerable research attention. Future, adequately powered, research may want to consider separating participants into paraplegic and tetraplegic groups or dichotomize by injuries above and below T6 to account for differences in sympathetic cardiovascular control. Currently, there is a paucity of studies analysing data in this fashion, which limits our understanding of how neurological level of injury and the degree of impaired sympathetic cardiovascular control influences the magnitude of change in CRF following an exercise intervention. Researchers may want to consider conducting a battery of autonomic nervous system stress tests at baseline (e.g., Valsalva manoeuvre, head-up tilt, sympathetic skin responses etc. [119]), to determine the degree of supraspinal sympathetic disruption rather than relying on a neurological level of injury derived from a motor-sensory examination. This is important as recent research has indicated that cardiovascular instability cannot be predicted by motor-sensory level and completeness of SCI [120].

There were no significant subgroup differences in CRF based on injury severity. However, the subgroup analysis suggests that individuals with a motor-incomplete SCI may not yield PPO improvements of the same magnitude as individuals with a motor-complete SCI. This is most likely due to the majority of motor-incomplete studies implementing gait training as its exercise modality, which we reveal is the least effective modality for improving CRF. The gait training interventions that measured PPO (N=2 interventions) used arm-crank ergometry (ACE) as the CPET modality, demonstrating no transfer effect from lower-body to upper-body exercise. During data extraction, reviewers noted poor reporting of injury severity in a number of studies. Whilst this may be due to older studies having used alternative severity scales (e.g., International Stoke Mandeville Games Federation or Frankel), researchers should endeavour to perform an International Standards for Neurological Classification of SCI (ISNCSCI) exam during screening, and subsequently report an AIS grade, to enable better comparisons to be made between injury severities in the future.

Van der Scheer et al. [35] concluded that there was high certainty in the evidence that exercise interventions ≥2 weeks can improve CRF in young and middle-aged adults. However, they revealed that there was a lack of studies exploring the effects of exercise in older adults with SCI (>65 years). The oldest mean age included in our review is 57.9 years [121], and thus supports their call for more research to be conducted in the older SCI population. Interestingly, our meta-regression identified that exercise interventions with a higher mean age were associated with smaller changes in V̇O_2peak_, suggesting that older adults do not achieve the same CRF benefits as younger or middle-aged adults. This is not surprising given the progressive physical deconditioning that occurs naturally with age, as previously shown in the non-injured population [122]. Research indicates that SCI represents a model of advanced ageing [123], with the ageing process being exacerbated in individuals with SCI possibly due to diminished mobility independence resulting in physical deconditioning. Older adults with SCI find it harder to change body position, transfer, and move around independently in comparison to younger adults with SCI [124]. Moreover, it has been suggested that older adults with SCI do not perform volumes or intensities of leisure time physical activity required to achieve fitness benefits [125]. These changes will likely result in reduced incidental physical activity outside of a prescribed exercise intervention. Ageing skeletal muscle is also susceptible to mitochondrial dysfunction, which may be related to chronic inflammation (e.g., “inflammaging”), possibly explaining the diminished responses in CRF for older adults with SCI. Future research may want to investigate optimal strategies for improving CRF in older adults. Moreover, there is a need for more longitudinal studies that explore the age-related decline in CRF in the SCI population, and whether this is accelerated relative to the non-injured population.

Despite a number of recent reviews summarising the effects of specific exercise modalities on the change in CRF following SCI, including aerobic ACE [126], FES-cycling [36], and aerobic plus muscle strength training (mixed multimodal) interventions [127], to the best of our knowledge our secondary, pooled meta-analysis is the first to directly compare the effects of a wide range of exercise modalities on the change in CRF in individuals with SCI. This review revealed there was a significant subgroup difference in RV̇O_2peak_, with the greatest changes gained via upper-body aerobic exercise or resistance training. The change in RV̇O_2peak_ for upper-body aerobic exercise in the current review (21%) is equivalent to the average 21% improvement reported in a recent systematic review on the effects of ACE in chronic SCI [126]. Whilst the current review did not exclusively investigate ACE, it is evident that aerobic, volitional upper-body exercise training can improve CRF in individuals with SCI. Activating larger amounts of skeletal muscle mass via FES exercise interventions also appears to improve RV̇O_2peak_, yet it is noteworthy that more accessible and less expensive training modalities such as aerobic and resistance training may yield similar or even greater increases in RV̇O_2peak_, despite utilising less muscle mass. Additionally, RV̇O_2peak_ improves following multimodal/hybrid exercise interventions, which challenges a 2015 review reporting inconclusive findings on the effects of combined upper-body aerobic and muscle strength training on CRF [127]. Yet, as the current review included a wide range of interventions not restricted to the upper-body (e.g., aquatic treadmill [63], hybrid cycling [64,69,128], multimodal exercises [129,130], etc.), it is recommended that more research is conducted to delineate whether the improvements in RV̇O_2peak_ with multimodal/hybrid exercise interventions are due to the combination of upper- and lower-body exercise modalities, or due to concurrent training modalities that predominantly use the upper-body (e.g., aerobic plus muscle strength training). Finally, both gait training and behaviour change interventions appear less effective at improving RV̇O_2peak_ and PPO.

Aerobic, upper-body exercise and resistance training modalities demonstrate the greatest improvements in PPO, by 15W and 20W, respectively. It is perhaps unsurprising that resistance training resulted in the largest change in PPO given that these interventions included upper-body exercises prescribed to increase muscular strength, as shown by Jacobs et al. [131]. Volitional exercise, as opposed to activity-based therapy modalities (i.e., interventions that provide activation of the neuromuscular system below the level of lesion with the goal of retraining the nervous system, such as FES and gait training), may therefore be more beneficial at improving PPO. Ultimately, these improvements have important ramifications for individuals with SCI that are dependent on performing functional upper-body movements such as transfers or wheelchair propulsion [99,103], and may lead to increased quality of life with more functional independence [132].

Several studies directly compared the effects of specific exercise modalities on the change in CRF [42,63,86]. Notably, Gorman et al. [63] demonstrated that there were no transfer effects from a robotic treadmill exercise intervention to ACE performance in a CPET. This review also demonstrates a trend for greater changes in RV̇O_2peak_ are likely achieved when the CPET modality is matched to the intervention (S9). Therefore, researchers should endeavour to match the CPET modality to their exercise intervention, or at the very least be careful when interpreting changes in CRF when using different modalities.

The current SCI-specific exercise guidelines recommend that exercise should be performed at a moderate-to-vigorous intensity [21]. A recent overview of systematic reviews also advocated the use of moderate-to-vigorous intensity for improving aerobic fitness [133]. The current secondary, pooled meta-analysis demonstrates robust improvements across all CRF outcomes for interventions prescribing exercise at this particular intensity. Furthermore, the secondary meta-analysis including cross-sectional studies reveals significant associations of a greater magnitude between MVPA and CRF, as compared to lower-intensity activity. Despite this, our classification of moderate-to-vigorous exercise intensity spans two of the ACSM exercise intensity thresholds (S3). There may be considerable variation in the actual intensity performed by participants given the noticeable range across the two thresholds (e.g., 46-90% V̇O_2peak_, 64-95% HR_peak_, 12-17 RPE etc.). Therefore, individuals with SCI and exercise practitioners should be cautious when prescribing such a broad exercise intensity.

The secondary meta-analysis comparing RCT exercise intensities reveals similar changes in CRF outcomes between moderate- and vigorous-intensity interventions. This is in agreement with a previous review [30] and supports the viewpoint from a special communication on high-intensity interval training (HIIT) [31], which suggested that vigorous-intensity exercise is more time efficient and may result in similar if not superior CRF and skeletal muscle oxidative capacity improvements in comparison to moderate-intensity exercise. Interestingly, in a response to a Letter-to-the-Editor [27], the SCI-specific exercise guideline developers acknowledge the need for shorter, effective protocols to be documented in the literature [134]. Furthermore, recent evidence has suggested that HIIT may be more enjoyable than moderate-intensity exercise for individuals with SCI [135], and so this form of training may offer a more time efficient and readily available alternative to moderate-intensity protocols. In the current review, a number of HIIT-based arm-crank ergometry, wheelchair propulsion, and FES hybrid cycling and skiing interventions demonstrated improvements in CRF [69,92,128,136–140]. The potential of “real-world” strategies such as virtual HIIT have also been discussed in a recent review by McMillan et al. [141]. Future exercise interventions should look to compare the effects of different HIIT modalities/protocols (i.e., virtual home-based HIIT versus supervised arm-crank HIIT, the most appropriate number and/or length of intervals and recovery periods, vigorous or supramaximal exercise intensities) before concrete recommendations can be made on the most optimal HIIT prescription for improving CRF. Whilst efforts are ongoing to corroborate the safety and feasibility of HIIT [32,92,142] both researchers and individuals with SCI should be vigilant in identifying risks associated with HIIT such as shoulder discomfort or pain, skin irritation or pressure sores caused by abrasive movements, or increased spasticity, along with monitoring for post-exercise hypotension [143]. As higher-intensity exercise generates a greater metabolic heat load than lower-intensity exercise, which increases core body temperature, considerations should be made for individuals with higher neurological levels of injury exercising at a high-intensity and/or in warm to hot ambient temperatures given their greater likelihood of experiencing thermoregulatory issues [144]. General contraindications for performing HIIT have been discussed in the non-injured population [145] yet they also apply for individuals with SCI.

This review reveals that RV̇O_2peak_ improves regardless of the method used to prescribe exercise intensity. With regards to PPO, subgroup difference indicates that the magnitude of change is greater when prescribing intensity via indices of HR (i.e., %HR_peak_, %HR_max_, %HRR) or V̇O_2_ (i.e., %V̇O_2peak_, %V̇O_2reserve_), compared to RPE and workload. A recent systematic review concluded that exercise interventions using RPE to prescribe relative exercise intensities improved PPO in individuals with SCI [146]. Previous research has revealed that RPE results in inter-individual responses to exercise, with the potential for two individuals to perform the same bout of exercise either above or below lactate threshold despite being prescribed the same perceptually regulated intensity, which prevents the development of SCI-specific RPE recommendations [147]. These differences identified in our meta-analysis may also be due to individuals with SCI being unaccustomed to subjective measures of exertion. Accordingly, there have been calls for better reporting of the standardisation and familiarisation procedures used for RPE [146] and have only tentatively recommended its use before the evidence base is expanded [148]. Therefore, it seems plausible to suggest that the blunted improvements in PPO with intensity prescribed via RPE, as compared to other prescription methods, may have resulted from insufficient familiarisation before an exercise intervention.

Although HR and V̇O_2_ have long been used to prescribe exercise intensity, these approaches can result in large training ranges and ignore individual metabolic responses. Particularly, issues may arise with using HR for individuals with a neurological level of injury ≥T6, given that these individuals typically exhibit a lower HR_peak_ [149]. The use of fixed percentages (i.e, %HR_peak_, %V̇O_2peak_) in the non-injured population has been questioned [150] and has recently been investigated in individuals with SCI, whereby Hutchinson et al. [56] showed that fixed %HR_peak_ and %V̇O_2peak_ could not guarantee a homogenous domain-specific exercise intensity prescription. Notably, individuals were spread across moderate, heavy and severe domains at the “moderate” and “vigorous” intensity classifications; thereby questioning whether the “moderate-to-vigorous” terminology used in the SCI-specific exercise guidelines is suitable for adults with SCI.

Given that prescribing exercise intensity via HR and V̇O_2_ can typically be resource and cost-intensive, there is some scope for using RPE as a cheaper and more practical method for community-based exercise prescription. However, this may not be as effective as other objective methods. Future research should aim to identify the optimal methods of exercise intensity prescription, as well as consider revisiting the current “moderate-to-vigorous intensity” recommendations. Moreover, further research may want to consider using traditional intensity anchors (e.g., the gas exchange threshold, critical power or lactate threshold) rather than prescribing exercise relative to physiological thresholds to see whether this results in more homogeneous CRF responses to exercise, given research in non-injured individuals suggests this may increase the precision of exercise intensity prescription [151,152]. However, it is worth acknowledging that it is not always possible to identify such traditional intensity anchors in individuals with higher levels of SCI [153,154].

Subgroup analyses based on frequency of sessions and exercise volume reveal no differences in RV̇O_2peak_, thereby supporting the minimal volume of exercise required to attain CRF benefits in individuals with SCI. Furthermore, although there are no statistically significant subgroup differences for PPO based on exercise volume, there is a greater magnitude of change observed for individuals exercising 90-149 min/wk in comparison to 40-89 min/wk (12W vs 7W change, respectively). A greater weekly exercise volume may therefore accrue greater changes in PPO and, as already described, may be important in improving the capacity to perform daily tasks such as bed or wheelchair transfers [99,103]. Although changes in CRF are similar between each exercise volume subgroup, and thus corresponding exercise guidelines, the secondary meta-analysis on cross-sectional cohorts indicates a significant cumulative impact of prolonged participation in physical activity and exercise. To support this point, a sensitivity analysis revealed a larger difference in RV̇O_2peak_ between inactive individuals and elite athletes, compared to between inactive and active individuals, suggesting that those who exercise more exhibit a greater CRF. Indeed, two cross-sectional association studies [40,98], reported significantly higher CRF in individuals with SCI that were habitually performing greater volumes of physical activity. Looking forward, longitudinal RCTs with multiple intervention arms would be the best way to explore dose-response changes with regards to differing volumes of exercise, as has been done in the non-injured population [155–158].

Subgroup analysis based on length of intervention indicated that exercise interventions of 12 weeks or less yield greater changes in RV̇O_2peak_ than those lasting >12 weeks. This may be explained by compliance and adherence issues during prolonged interventions (i.e., >12 weeks). Indeed, the two behaviour change studies included in this subgroup analysis observed minimal changes in RV̇O_2peak_ following 16 [82] and 24 weeks [39]. This perhaps emphasises the benefit of short and intensive exercise interventions ≤12 weeks as well as the need for supervised exercise sessions in prolonged interventions to ensure compliance and meaningful changes in RV̇O_2peak_.

Adverse events were reported for at least 3.7% of the total included participants, with the majority of events related to skin irritation, pressure sores or ulcers. Qualitatively, there was no particular exercise modality that suggested an increased risk for an adverse event, but higher-intensity exercise appeared to reveal more adverse events, albeit being swayed by one study in particular [130]. Reporting was poor in a number of studies with reviewers at times unable to determine the exact number of events per participant. Furthermore, there is generally a lack of follow-up assessments following exercise interventions, so it is currently unknown whether there are any detrimental long-term effects of exercise in the SCI population. Going forward, researchers are encouraged to follow a standardised adverse event reporting procedure (including serious and non-serious adverse events) and ensure that they are transparent with reporting of both the nature and the total numbers of each event, either related or unrelated to the exercise intervention.

### 4.1. Strengths and limitations of the review and future directions

#### 4.1.1. Limitations of the included studies

Poor reporting of injury characteristics and exercise parameters prevented a perfect comparison of exercise interventions. Overall, studies could have provided more precise descriptions of training parameters to aid with any future refinements to the SCI-specific exercise guidelines. Reporting of adherence to interventions was also poor and should be encouraged to provide an indication of the feasibility or applicability of specific exercise interventions for individuals with SCI. Moreover, adverse events should be transparently reported, even if none occur so that practitioners are able to identify forms of exercise that are most likely to be safe for this population. Additionally, studies typically failed to utilise the training principle of progression, which during prolonged exercise interventions is essential for preventing a plateau in training adaptations and perhaps particularly important in this population for supporting the transition from an inactive lifestyle to higher levels of activity, and ultimately achieving greater CRF benefits [24]. On the whole, the reporting of V̇O_2peak_ attainment criteria was poor, with only 24% of the included exercise interventions using at least two criterion methods for identifying when an individual had reached peak capacity, as recently recommended by Alrashidi et al. [159]. At least two methods [e.g., RPE ≥17, respiratory exchange ratio (RER) ≥1.1, plateau in oxygen uptake] should be adopted for confirming the attainment of a true V̇O_2peak_ to prevent magnitudes of change in CRF from being inflated or underestimated. Furthermore, to the best of our knowledge, only 30% of interventions had a prospectively registered clinical trial entry and only 7.7% had a protocol manuscript published. To sustain the integrity and transparency of reporting in this field, researchers are encouraged to prospectively register any planned clinical trials using publicly available repositories.

The risk of bias assessments on pre-post studies revealed that no study conducted multiple baseline or follow-up assessments. Whilst often time-consuming and impractical with larger sample sizes, multiple assessments ensure reproducibility by accounting for any technical or biological variation, as shown previously in non-injured individuals at risk for type-2 diabetes [160]. In the SCI population, individuals are typically deconditioned and often exhibit variable responses to a CPET. This variance may be explained by profound blood pressure instability [161], including unintentional ‘boosting’ via episodes of autonomic dysreflexia [162]. Researchers should therefore consider performing multiple CPETs at baseline and follow-up to attain reliable assessments of CRF.

There are also several limitations with regards to the studies included in the secondary meta-analyses for this review. First, there is only one cross-sectional study using a research-grade wearable device to investigate the association between physical activity and CRF [98]. Whilst self-report questionnaires are valid tools for estimating levels of physical activity [97,163–165], there are important drawbacks including the difficulty of accurately capturing intensity, lack of questionnaires measuring activities of daily living, and recall bias. Secondly, there is a lack of RCTs comparing near-maximal, maximal or supramaximal exercise intensities to moderate-intensity exercise. The only supramaximal intervention included in this review demonstrated a 17W improvement in PPO [139]. The inclusion of more RCTs comparing vigorous-intensity to lower intensity exercise could identify whether there are, in fact, benefits to performing shorter but more vigorous-intensity exercise bouts, in comparison to longer continuous forms of exercise.

#### 4.1.2. Strengths and limitations of the review

A major strength of the current study is that we pre-planned and prospectively registered (PROSPERO ID CRD42018104342) our systematic review. We used GRADE to assess the certainty in the body of evidence and used quality appraisal tools for the specific study designs included in this review. Our GRADE assessment demonstrates generalisability within the SCI population, through the inclusion of participants across the lifespan and with a wide range of injury characteristics. Yet, the ‘Low’ confidence in the evidence for AV̇O_2peak_ and PPO emphasises the need for more rigorous exercise interventions to address current gaps in the literature [35]. The disparity in GRADE confidence ratings across the specific CRF outcomes likely is a factor of the variability of the total number of included interventions across outcomes AV̇O_2peak_ (N=74), RV̇O_2peak_ (N=79) and PPO (N=65).

As there were not enough RCTs to perform subgroup comparisons and a meta-regression on this study design specifically, we pooled pre-post (N=81) and RCT (N=36) exercise interventions. This is in accordance with Cochrane guidance stating that the inclusion of non-randomised study designs is justified when there are only a small number of RCTs available to provide evidence of the effects of interventions [53]. To consider this limitation, we generated firepower plots to explore the statistical power of the included RCTs and pre-post studies. These plots demonstrate that the RCTs had greater median statistical power across CRF outcomes and were designed to reliably detect a wider range of effect sizes than the pre-post studies. However, the changes in RV̇O_2peak_ and PPO in the primary meta-analysis of RCT interventions relative to controls (2.9 mL/kg/min and 9W, respectively) are similar to those reported in the secondary, pooled meta-analysis (2.8 mL/kg/min and 11W, respectively), and thus confirms the robustness of our overall findings. Furthermore, our rigorous approach of adjusting for multiple comparisons minimises any erroneous interpretations of subgroup differences and therefore strengthens our conclusions on the available evidence.

Despite this, the categorisation of interventions within each subgroup could be considered a limitation of the current review. Whilst this was done to directly compare the effects of different subgroups (i.e., acute vs chronic, tetraplegia vs paraplegia, aerobic vs resistance vs FES etc.), it resulted in an unequal number of interventions within each classification and likely underpowered the subgroup comparisons. For example, the subgroup analysis based on exercise intensity reveals an effect of exercise intensity on PPO, yet this may be influenced by the small number of interventions for light- and supramaximal-intensity. Despite reporting some significant subgroup differences across dichotomised studies, these variables were not identified as significant moderator variables in the random-effects meta-regression, meaning these findings should be viewed with caution. It is perhaps more of a limitation of the evidence-base *per se*, rather than our meta-analysis, in that more RCTs should be conducted to increase the power of these subgroups. Another limitation is that despite our comprehensive search strategy we may have missed relevant studies as abstracts, theses, and other unpublished work were not included.

### 4.2. Clinical implications and future directions

Our results support the current guidelines regarding the minimal weekly volume of exercise necessary to improve CRF in the SCI population. However, our pooled analysis indicates subgroup differences for PPO based on certain exercise intervention parameters. To the best of our knowledge, there are no large-scale epidemiological studies investigating the dose-response relationship between physical activity and CRF in this population using sensitive and validated methods to quantify the exposure variable (e.g. free-living physical activity). Such studies have been performed in non-injured individuals [166,167]. To identify the optimal stimulus for beneficial CRF responses in this population, dose-ranging studies, akin to those that are used in the pharmaceutical industry, should be conducted. A recent overview of systematic reviews [168] highlighted the poor reporting in exercise interventions in health and disease and called upon the inclusion of checklists [e.g., the Consensus on Exercise Reporting Template (CERT) [169] or the Template for Intervention Description and Replication (TIDieR) [170]] to improve study quality. This would ultimately lead to a better understanding of the ‘dose’ of exercise as medicine required to optimise CRF outcomes in this population.

Both the primary meta-analysis of RCTs (Δ2.9 mL/kg/min) and the secondary, pooled meta-analysis (Δ2.8 mL/kg/min) reveal that exercise interventions >2 weeks result in an overall increase in RV̇O_2peak_ which is roughly equivalent to 1 MET-SCI [metabolic equivalent in SCI (2.7 mL/kg/min)] [55]. An increase in maximal aerobic capacity (an estimate of CRF) by 1 MET (3.5 mL/kg/min) in non-injured individuals is associated with a 13% and 15% reduction in all-cause and cardiovascular mortality, respectively [171]. The current review shows that individuals meeting the SCI-specific guidelines for cardiometabolic health [21] can improve RV̇O_2peak_ to a similar magnitude to the overall pooled effect (∼1 MET-SCI), highlighting that these guidelines may offer a reduction in CVD risk, and therefore mortality. Nonetheless, an association between an improvement in CRF and a reduction in mortality is yet to be established specifically in the SCI population and remains an important avenue of research for the future.

## 5. CONCLUSION

This systematic review with meta-analysis provides an updated, evidence-based summary of the effects of exercise interventions on CRF in individuals with SCI. Based on evidence of moderate-to-low certainty, exercise interventions >2 weeks are associated with significant improvements in CRF, and in particular, a clinically meaningful change in RV̇O_2peak_. Subgroup comparisons identified that upper-body, aerobic exercise and resistance training appear the most effective at improving RV̇O_2peak_ and PPO. Furthermore, acutely-injured, paraplegic individuals, exercising at a moderate-to-vigorous intensity, prescribed via V̇O_2_ or HR (at a minimum of either 46% V̇O_2peak_, 64% HR_peak_, or 40% HRR/V̇O_2reserve_, in accordance with ACSM classifications), for more than 3 sessions/week will likely experience the greatest change in PPO. Exercise interventions up to 12-weeks are also most likely to lead to improvements in RV̇O_2peak_. The meta-regression revealed that older adults may experience smaller changes in V̇O_2peak_ following an exercise intervention. Importantly, there is an ever-growing need for studies to establish a dose-response relationship between exercise and CRF in the SCI population to determine the most optimal form of exercise prescription to reduce the wide-ranging consequences typically associated with SCI. To improve the certainty of evidence in the field moving forward, we call for the development of an SCI-specific reporting template for exercise interventions, as well as encourage researchers to pre-register and/or publish protocol papers for prospective clinical exercise trials. Researchers should also consider whether injury characteristics or participant demographics (e.g., impact of neurological level of injury/severity on motor-sensory or autonomic cardiovascular control, age, sex) influence changes in CRF outcomes with a period of exercise training.

## Supporting information

S1 PRISMA checklist

S2 Search strategy

S3 Exercise intensity classifications

S4 Correlation factors

S5 Primary meta-analysis (RCTs)

S6 Secondary pooled meta-analysis (pre-post and RCT exercise interventions) (

S7 Adverse events

S8 Sensitivity analyses

S9 Cardiopulmonary exercise test modalities sub-analysis

S10 Gait training sub-analysis

S11 Cohort comparisons

S12 Cross-sectional associations

S13 Observational studies

S14 RCT exercise intensity comparisons

## Data Availability

All code/scripts and data used for the analysis are available at https://github.com/jutzca/Exercise-and-fitness-in-SCI

## SUPPLEMENTARY MATERIAL

**Supplementary material 1 (S1): PRISMA Checklist**

**Supplementary file 2 (S2): Systematic Review Search Strategy**

**Supplementary file 3 (S3): Description of Exercise Intensity Classifications as per the American College of Sports Medicine (ACSM) guidelines**

**Supplementary file 4 (S4): Calculated Correlation Factors**

**Supplementary file 5 (S5): Primary meta-analysis (RCT interventions relative to controls)**

1. Overview of participant demographics and injury characteristics for the pooled RCTs (exercise intervention vs. true-world control or standard of care)

2. Summary of the individual RCTs included in the review

3. Quality assessment rating for each study using the Cochrane Risk of Bias 2 tool

4. Forest plots and funnel plots for each CRF outcome

5. References

**Supplementary file 6 (S4): Change in CRF outcomes in response to prospective, well-characterised exercise interventions lasting >2 weeks from pre-post and RCT studies (Secondary, pooled meta-analysis)**

1. Overview of participant demographics and injury characteristics for the pooled exercise interventions from pre-post and RCT studies

2. Summary statistics of the subgroup analyses

3. Figure visualising the worldwide coverage of exercise interventions included in the secondary, pooled meta-analysis

4. Summary of the individual studies included in the review

5. Forest and funnel plots for change in each CRF outcome for each subgroup comparison (time since injury, neurological level of injury, injury severity, exercise modality, length of intervention, relative exercise intensity, method of exercise intensity prescription, frequency of exercise sessions, and exercise volume)

6. Quality assessment ratings for each pre-post study included in the primary meta-analysis

7. Risk of bias for each RCT intervention arm included in the primary meta-analysis

8. References

**Supplementary file 7 (S7): Adverse events**

**Supplementary file 8 (S8): Sensitivity analyses**

**Supplementary file 9 (S9): CPET vs. exercise intervention modality**

1. Forest plots for each CRF outcome

**Supplementary file 10 (S10): Gait-training sub-analysis**

**1. Forest plots for each CRF outcome**

**Supplementary file 11 (S11): Cross-sectional cohort comparisons summary (secondary meta-analysis)**

1. Overview of participant demographics and injury characteristics for the pooled cohort comparisons

2. Summary of the individual studies included in the review

3. Quality assessment rating for each study using the NIH tool for observational cohort and cross-sectional studies

4. Forest plots and funnel plots for each CRF outcome

5. Sensitivity analysis on RV̇O_2peak_

6. References

**Supplementary file 12 (S12): Cross-sectional associations between physical activity and CRF outcomes**

1. Overview of participant demographics and injury characteristics for the pooled association comparisons

2. Summary of the individual studies included in the review

3. Quality assessment rating for each study using the NIH tool for observational cohort and cross-sectional studies

4. Visualisation of correlation coefficients between physical activity dimensions and CRF outcomes across included studies

5. References

**Supplementary file 13 (S13): Observational studies (secondary meta-analysis)**

1. Overview of participant demographics and injury characteristics for the pooled observational studies

2. Summary of the individual studies included in the review

3. Quality assessment rating for each study using the NIH tool for observational cohort and cross-sectional studies

4. Forest plots and funnel plots for each CRF outcome

5. References

**Supplementary file 14 (S14): RCTs intensity comparisons (secondary meta-analysis)**

1. Overview of participant demographics and injury characteristics for the pooled RCTs comparing the effects of different exercise intensities

2. Summary of the individual RCTs comparing exercise intensity included in the review

3. Quality assessment rating for each study using the Cochrane Risk of Bias 2 too1

4. Forest plots and funnel plots for each CRF outcome

5. References

## 6. ACKNOWLEDGEMENTS

We would like to thank Mr. Sajjad Tavassoly (Faculty of Medicine, UBC, Vancouver, Canada) for his assistance with piloting the initial database search. We would also like to thank Dr. Matthew Querée (Department of Physical Therapy, UBC, Spinal Cord Injury Research Evidence Team, GF Strong Rehabilitation Centre, Vancouver, Canada) for his assistance with the risk of bias assessments. We appreciate the assistance of Mr Adrian Cheng (ICORD) and Dr. Alex Williams (ICORD), who helped pilot the inclusion/exclusion of articles and supported the use of Photoshop to extract data from figures, respectively. We thank Dr. Sam Weaver (University of Birmingham) for support with the meta-analysis using R. Figure 3 was created using Biorender.com.

## 7. LIST OF ABBREVIATIONS

1RM: One repetition maximum Arm-crank ergometry
ACSM: American College of Sports Medicine
AIS: American Spinal Injury Association Impairment Scale
AV̇O2peak: Absolute peak oxygen uptake
CENTRAL: Cochrane Central Register of Controlled Trials
CERT: Consensus on Exercise Reporting Template
CI: Confidence interval
CPET: Cardiopulmonary exercise test
CRF: Cardiorespiratory fitness
CVD: Cardiovascular disease
EMBASE: Excerpta Medica Database
FES: Functional electrical stimulation
GRADE: Grading of Recommendations, Assessment, Development and Evaluation
HIIT: High-intensity interval training
HR: Heart rate
HRmax: Maximum heart rate (age-predicted)
HRpeak: Peak heart rate
HRR: Heart rate reserve
IQR: Interquartile range
ISNCSCI: International Standards for Neurological Classification of Spinal Cord Injury
LTPA: Leisure time physical activity
MET: Metabolic equivalent
MTP: Maximal tolerated power
MVPA: Moderate-to-vigorous physical activity
PPO: Peak power output
PRISMA: Preferred Reporting Items for Systematic Reviews
RCT: Randomised-controlled trial
RoB 2: The Cochrane Risk of Bias 2 tool
RPE: Rating of perceived exertion
RV̇O2peak: Relative peak oxygen uptake
SCI: Spinal cord injury
SCS: Spinal cord stimulation
SD: Standard deviation
TIDieR: Template for Intervention Description and Replication
TSI: Time since injury
V̇O2: Oxygen uptake
V̇O2peak: Peak oxygen uptake
V̇O2reserve: Reserve oxygen uptake
W: Watts
WMD: Weighted mean difference

## REFERENCES

1. Miller LE, Herbert WG. Health and economic benefits of physical activity for patients with spinal cord injury. Clinicoecon Outcomes Res. 2016;8: 551–558.

2. Rimmer JH, Schiller W, Chen M-D. Effects of disability-associated low energy expenditure deconditioning syndrome. Exerc Sport Sci Rev. 2012;40: 22–29.

3. Cragg JJ, Noonan VK, Dvorak M, Krassioukov A, Mancini GBJ, Borisoff JF. Spinal cord injury and type 2 diabetes: results from a population health survey. Neurology. 2013;81: 1864–1868.

4. Cragg JJ, Noonan VK, Krassioukov A, Borisoff J. Cardiovascular disease and spinal cord injury: results from a national population health survey. Neurology. 2013;81: 723–728.

5. Wu J-C, Chen Y-C, Liu L, Chen T-J, Huang W-C, Cheng H, et al. Increased risk of stroke after spinal cord injury: a nationwide 4-year follow-up cohort study. Neurology. 2012;78: 1051–1057.

6. Ross R, Blair SN, Arena R, Church TS, Després J-P, Franklin BA, et al. Importance of Assessing Cardiorespiratory Fitness in Clinical Practice: A Case for Fitness as a Clinical Vital Sign: A Scientific Statement From the American Heart Association. Circulation. 2016;134: e653–e699.

7. Clausen Johan S.R., Marott Jacob L., Holtermann Andreas, Gyntelberg Finn, Jensen Magnus T. Midlife Cardiorespiratory Fitness and the Long-Term Risk of Mortality. J Am Coll Cardiol. 2018;72: 987–995.

8. Lee D-C, Sui X, Artero EG, Lee I-M, Church TS, McAuley PA, et al. Long-term effects of changes in cardiorespiratory fitness and body mass index on all-cause and cardiovascular disease mortality in men: the Aerobics Center Longitudinal Study. Circulation. 2011;124: 2483–2490.

9. Myers J, Prakash M, Froelicher V, Do D, Partington S, Atwood JE. Exercise capacity and mortality among men referred for exercise testing. N Engl J Med. 2002;346: 793–801.

10. Laukkanen JA, Kurl S, Salonen R, Rauramaa R, Salonen JT. The predictive value of cardiorespiratory fitness for cardiovascular events in men with various risk profiles: a prospective population-based cohort study. Eur Heart J. 2004;25: 1428–1437.

11. Erikssen G, Liestøl K, Bjørnholt J, Thaulow E, Sandvik L, Erikssen J. Changes in physical fitness and changes in mortality. Lancet. 1998;352: 759–762.

12. Lee DC, Sui X, Ortega FB, Kim Y-S, Church TS, Winett RA, et al. Comparisons of leisure-time physical activity and cardiorespiratory fitness as predictors of all-cause mortality in men and women. Br J Sports Med. 2011;45: 504–510.

13. Haisma JA, van der Woude LHV, Stam HJ, Bergen MP, Sluis TAR, Bussmann JBJ. Physical capacity in wheelchair-dependent persons with a spinal cord injury: a critical review of the literature. Spinal Cord. 2006;44: 642–652.

14. Simmons OL, Kressler J, Nash MS. Reference fitness values in the untrained spinal cord injury population. Arch Phys Med Rehabil. 2014;95: 2272–2278.

15. Stofan JR, DiPietro L, Davis D, Kohl HW 3rd, Blair SN. Physical activity patterns associated with cardiorespiratory fitness and reduced mortality: the Aerobics Center Longitudinal Study. Am J Public Health. 1998;88: 1807–1813.

16. Nightingale TE, Williams S, Thompson D, Bilzon JLJ. Energy balance components in persons with paraplegia: daily variation and appropriate measurement duration. Int J Behav Nutr Phys Act. 2017;14: 132.

17. van den Berg-Emons RJ, Bussmann JB, Stam HJ. Accelerometry-based activity spectrum in persons with chronic physical conditions. Arch Phys Med Rehabil. 2010;91: 1856–1861.

18. Hicks AL, Martin Ginis KA, Pelletier CA, Ditor DS, Foulon B, Wolfe DL. The effects of exercise training on physical capacity, strength, body composition and functional performance among adults with spinal cord injury: a systematic review. Spinal Cord. 2011;49: 1103–1127.

19. Valent L, Dallmeijer A, Houdijk H, Talsma E, van der Woude L. The effects of upper body exercise on the physical capacity of people with a spinal cord injury: a systematic review. Clin Rehabil. 2007;21: 315–330.

20. Ginis KAM, Hicks AL, Latimer AE, Warburton DER, Bourne C, Ditor DS, et al. The development of evidence-informed physical activity guidelines for adults with spinal cord injury. Spinal Cord. 2011;49: 1088–1096.

21. Martin Ginis KA, van der Scheer JW, Latimer-Cheung AE, Barrow A, Bourne C, Carruthers P, et al. Evidence-based scientific exercise guidelines for adults with spinal cord injury: an update and a new guideline. Spinal Cord. 2018;56: 308–321.

22. Smith B, Kirby N, Skinner B, Wightman L, Lucas R, Foster C. Infographic. Physical activity for disabled adults. Br J Sports Med. 2019;53: 335–336.

23. Bull FC, Al-Ansari SS, Biddle S, Borodulin K, Buman MP, Cardon G, et al. World Health Organization 2020 guidelines on physical activity and sedentary behaviour. Br J Sports Med. 2020;54: 1451–1462.

24. Tweedy SM, Beckman EM, Geraghty TJ, Theisen D, Perret C, Harvey LA, et al. Exercise and sports science Australia (ESSA) position statement on exercise and spinal cord injury. J Sci Med Sport. 2017;20: 108–115.

25. Piercy KL, Troiano RP, Ballard RM, Carlson SA, Fulton JE, Galuska DA, et al. The Physical Activity Guidelines for Americans. JAMA. 2018;320: 2020–2028.

26. Powell KE, Paluch AE, Blair SN. Physical activity for health: What kind? How much? How intense? On top of what? Annu Rev Public Health. 2011;32: 349–365.

27. Tweedy SM, Beckman EM, Connick MJ, Geraghty TJ, Theisen D, Perret C, et al. Correspondence re “Evidence-based scientific exercise guidelines for adults with spinal cord injury: an update and new guideline.” Spinal cord. 2018. pp. 406–408.

28. Yue T, Wang Y, Liu H, Kong Z, Qi F. Effects of High-Intensity Interval vs. Moderate-Intensity Continuous Training on Cardiac Rehabilitation in Patients With Cardiovascular Disease: A Systematic Review and Meta-Analysis. Front Cardiovasc Med. 2022;9: 845225.

29. Hannan AL, Hing W, Simas V, Climstein M, Coombes JS, Jayasinghe R, et al. High-intensity interval training versus moderate-intensity continuous training within cardiac rehabilitation: a systematic review and meta-analysis. Open Access J Sports Med. 2018;9: 1–17.

30. Peters J, Abou L, Rice LA, Dandeneau K, Alluri A, Salvador AF, et al. The effectiveness of vigorous training on cardiorespiratory fitness in persons with spinal cord injury: a systematic review and meta-analysis. Spinal Cord. 2021;59: 1035–1044.

31. Nightingale TE, Metcalfe RS, Vollaard NB, Bilzon JL. Exercise Guidelines to Promote Cardiometabolic Health in Spinal Cord Injured Humans: Time to Raise the Intensity? Arch Phys Med Rehabil. 2017;98: 1693–1704.

32. Astorino TA, Hicks AL, Bilzon JLJ. Viability of high intensity interval training in persons with spinal cord injury-a perspective review. Spinal Cord. 2021;59: 3–8.

33. Farrow M, Nightingale TE, Maher J, McKay CD, Thompson D, Bilzon JLJ. Effect of Exercise on Cardiometabolic Risk Factors in Adults With Chronic Spinal Cord Injury: A Systematic Review. Arch Phys Med Rehabil. 2020;101: 2177–2205.

34. Moher D, Liberati A, Tetzlaff J, Altman DG, PRISMA Group. Preferred reporting items for systematic reviews and meta-analyses: the PRISMA statement. BMJ. 2009;339: b2535.

35. van der Scheer JW, Martin Ginis KA, Ditor DS, Goosey-Tolfrey VL, Hicks AL, West CR, et al. Effects of exercise on fitness and health of adults with spinal cord injury: A systematic review. Neurology. 2017;89: 736–745.

36. van der Scheer JW, Goosey-Tolfrey VL, Valentino SE, Davis GM, Ho CH. Functional electrical stimulation cycling exercise after spinal cord injury: a systematic review of health and fitness-related outcomes. J Neuroeng Rehabil. 2021;18: 99.

37. Jackson JL, Kuriyama A, Anton A, Choi A, Fournier J-P, Geier A-K, et al. The Accuracy of Google Translate for Abstracting Data From Non-English-Language Trials for Systematic Reviews. Ann Intern Med. 2019;171: 677–679.

38. Lannem AM, Sørensen M, Lidal IB, Hjeltnes N. Perceptions of exercise mastery in persons with complete and incomplete spinal cord injury. Spinal Cord. 2010;48: 388–392.

39. Bombardier CH, Dyer JR, Burns P, Crane DA, Takahashi MM, Barber J, et al. A tele-health intervention to increase physical fitness in people with spinal cord injury and cardiometabolic disease or risk factors: a pilot randomized controlled trial. Spinal Cord. 2021;59: 63–73.

40. Hoevenaars D, Holla JFM, Postma K, van der Woude LHV, Janssen TWJ, de Groot S. Associations between meeting exercise guidelines, physical fitness, and health in people with spinal cord injury. Disabil Rehabil. 2022; 1–8.

41. Piira A, Lannem AM, Sørensen M, Glott T, Knutsen R, Jørgensen L, et al. Manually assisted body-weight supported locomotor training does not re-establish walking in non-walking subjects with chronic incomplete spinal cord injury: A randomized clinical trial. J Rehabil Med. 2019;51: 113–119.

42. Farkas GJ, Gorgey AS, Dolbow DR, Berg AS, Gater DR Jr. Energy Expenditure, Cardiorespiratory Fitness, and Body Composition Following Arm Cycling or Functional Electrical Stimulation Exercises in Spinal Cord Injury: A 16-Week Randomized Controlled Trial. Top Spinal Cord Inj Rehabil. 2021;27: 121–134.

43. Dallmeijer AJ, Hopman MT, van der Woude LH. Lipid, lipoprotein, and apolipoprotein profiles in active and sedentary men with tetraplegia. Arch Phys Med Rehabil. 1997;78: 1173–1176.

44. Davis GM, Shephard RJ, Leenen FH. Cardiac effects of short term arm crank training in paraplegics: echocardiographic evidence. Eur J Appl Physiol Occup Physiol. 1987;56: 90–96.

45. Hansen RK, Samani A, Laessoe U, Handberg A, Mellergaard M, Figlewski K, et al. Rowing exercise increases cardiorespiratory fitness and brachial artery diameter but not traditional cardiometabolic risk factors in spinal cord-injured humans. Eur J Appl Physiol. 2023; 1–15.

46. Higgins JPT, Sterne JAC, Savovic J, Page MJ, Hróbjartsson A, Boutron I, et al. A revised tool for assessing risk of bias in randomized trials. Cochrane Database Syst Rev. 2016;10: 29–31.

47. McGuinness LA, Higgins JPT. Risk-of-bias VISualization (robvis): An R package and Shiny web app for visualizing risk-of-bias assessments. Res Synth Methods. 2021;12: 55–61.

48. American College Medicine. ACSM’s guidelines for exercise testing and prescription. 9th ed. Lippincott Williams & Wilkins; 2013. Available: https://shop.lww.com/ACSM-s-Guidelines-for-Exercise-Testing-and-Prescription/p/9781975150181

49. Higgins JPT, Li T, Deeks JJ. Choosing effect measures and computing estimates of effect. Cochrane Handbook for Systematic Reviews of Interventions. Wiley; 2019. pp. 143–176. doi:10.1002/9781119536604.ch6

50. Hozo SP, Djulbegovic B, Hozo I. Estimating the mean and variance from the median, range, and the size of a sample. BMC Med Res Methodol. 2005;5: 13.

51. Viechtbauer W. Conducting Meta-Analyses in R with the metafor Package. J Stat Softw. 2010;36: 1–48.

52. Balduzzi S, Rücker G, Schwarzer G. How to perform a meta-analysis with R: a practical tutorial. Evid Based Ment Health. 2019;22: 153–160.

53. Higgins JPT, Thomas J, Chandler J, Cumpston M, Li T, Page MJ, et al. Cochrane Handbook for Systematic Reviews of Interventions. John Wiley & Sons; 2019.

54. Quintana DS. A Guide for Calculating Study-Level Statistical Power for Meta-Analyses. Advances in Methods and Practices in Psychological Science. 2023;6: 25152459221147260.

55. Collins EG, Gater D, Kiratli J, Butler J, Hanson K, Langbein WE. Energy cost of physical activities in persons with spinal cord injury. Med Sci Sports Exerc. 2010;42: 691–700.

56. Hutchinson MJ, Goosey-Tolfrey VL. Rethinking aerobic exercise intensity prescription in adults with spinal cord injury: time to end the use of “moderate to vigorous” intensity? Spinal Cord. 2021; 1–7.

57. Higgins JPT, Thompson SG. Controlling the risk of spurious findings from meta-regression. Stat Med. 2004;23: 1663–1682.

58. Guyatt GH, Oxman AD, Vist GE, Kunz R, Falck-Ytter Y, Alonso-Coello P, et al. GRADE: an emerging consensus on rating quality of evidence and strength of recommendations. BMJ. 2008;336: 924–926.

59. Han D-S, Hsiao M-Y, Wang T-G, Chen S-Y, Yang W-S. Association of serum myokines and aerobic exercise training in patients with spinal cord injury: an observational study. BMC Neurol. 2016;16: 142.

60. Hoekstra F, van Nunen MPM, Gerrits KHL, Stolwijk-Swüste JM, Crins MHP, Janssen TWJ. Effect of robotic gait training on cardiorespiratory system in incomplete spinal cord injury. J Rehabil Res Dev. 2013;50: 1411–1422.

61. Pollack SF, Axen K, Spielholz N, Levin N, Haas F, Ragnarsson KT. Aerobic training effects of electrically induced lower extremity exercises in spinal cord injured people. Arch Phys Med Rehabil. 1989;70: 214–219.

62. Barstow TJ, Scremin AM, Mutton DL, Kunkel CF, Cagle TG, Whipp BJ. Changes in gas exchange kinetics with training in patients with spinal cord injury. Med Sci Sports Exerc. 1996;28: 1221–1228.

63. Gorman PH, Scott W, VanHiel L, Tansey KE, Sweatman WM, Geigle PR. Comparison of peak oxygen consumption response to aquatic and robotic therapy in individuals with chronic motor incomplete spinal cord injury: a randomized controlled trial. Spinal Cord. 2019;57: 471–481.

64. Heesterbeek PJC, Berkelmans HWA, Thijssen DHJ, van Kuppevelt HJM, Hopman MTE, Duysens J. Increased physical fitness after 4-week training on a new hybrid FES-cycle in persons with spinal cord injury. Technol Disabil. 2005;17: 103–110.

65. Mutton DL, Scremin AM, Barstow TJ, Scott MD, Kunkel CF, Cagle TG. Physiologic responses during functional electrical stimulation leg cycling and hybrid exercise in spinal cord injured subjects. Arch Phys Med Rehabil. 1997;78: 712–718.

66. Ozturk ED, Lapointe MS, Kim D-I, Hamner JW, Tan CO. Effect of 6-Month Exercise Training on Neurovascular Function in Spinal Cord Injury. Med Sci Sports Exerc. 2021;53: 38–46.

67. Hooker SP, Scremin AM, Mutton DL, Kunkel CF, Cagle G. Peak and submaximal physiologic responses following electrical stimulation leg cycle ergometer training. J Rehabil Res Dev. 1995;32: 361–366.

68. Hooker SP, Figoni SF, Rodgers MM, Glaser RM, Mathews T, Suryaprasad AG, et al. Physiologic effects of electrical stimulation leg cycle exercise training in spinal cord injured persons. Arch Phys Med Rehabil. 1992;73: 470–476.

69. Hasnan N, Engkasan JP, Husain R, Davis GM. High-Intensity Virtual-reality Arm plus FES-leg Interval Training in Individuals with Spinal Cord Injury. Biomed Tech . 2013;58 Suppl 1. doi:10.1515/bmt-2013-4028

70. Janssen TWJ, Pringle DD. Effects of modified electrical stimulation-induced leg cycle ergometer training for individuals with spinal cord injury. J Rehabil Res Dev. 2008;45: 819–830.

71. Jeon JY, Hettinga D, Steadward RD, Wheeler GD, Bell G, Harber V. Reduced plasma glucose and leptin after 12 weeks of functional electrical stimulation-rowing exercise training in spinal cord injury patients. Arch Phys Med Rehabil. 2010;91: 1957–1959.

72. Midha M, Schmitt JK, Sclater M. Exercise effect with the wheelchair aerobic fitness trainer on conditioning and metabolic function in disabled persons: a pilot study. Arch Phys Med Rehabil. 1999;80: 258–261.

73. Milia R, Roberto S, Marongiu E, Olla S, Sanna I, Angius L, et al. Improvement in hemodynamic responses to metaboreflex activation after one year of training in spinal cord injured humans. Biomed Res Int. 2014;2014: 893468.

74. Lavado EL, Cardoso JR, Silva LGA, Dela Bela LF, Atallah AN. Effectiveness of aerobic physical training for treatment of chronic asymptomatic bacteriuria in subjects with spinal cord injury: a randomized controlled trial. Clin Rehabil. 2013;27: 142–149.

75. Brazg G, Fahey M, Holleran CL, Connolly M, Woodward J, Hennessy PW, et al. Effects of Training Intensity on Locomotor Performance in Individuals With Chronic Spinal Cord Injury: A Randomized Crossover Study. Neurorehabil Neural Repair. 2017;31: 944–954.

76. Schleifer G, Solinsky R, Hamner JW, Picard G, Taylor JA. Hybrid Functional Electrical Stimulation Improves Anaerobic Threshold in First Three Years after Spinal Cord Injury. J Neurotrauma. 2022;39: 1050–1056.

77. Beillot J, Carré F, Le Claire G, Thoumie P, Perruoin-Verbe B, Cormerais A, et al. Energy consumption of paraplegic locomotion using reciprocating gait orthosis. Eur J Appl Physiol Occup Physiol. 1996;73: 376–381.

78. Koch I, Schlegel M, Pirrwitz A, Jaschke B, Schlegel K. [Objectification of the training effect of sports therapy for wheelchair users]. Int J Rehabil Res. 1983;6: 439–448.

79. Kooijmans H, Post MWM, Stam HJ, van der Woude LHV, Spijkerman DCM, Snoek GJ, et al. Effectiveness of a Self-Management Intervention to Promote an Active Lifestyle in Persons With Long-Term Spinal Cord Injury: The HABITS Randomized Clinical Trial. Neurorehabil Neural Repair. 2017;31: 991–1004.

80. Ma JK, West CR, Martin Ginis KA. The Effects of a Patient and Provider Co-Developed, Behavioral Physical Activity Intervention on Physical Activity, Psychosocial Predictors, and Fitness in Individuals with Spinal Cord Injury: A Randomized Controlled Trial. Sports Med. 2019;49: 1117–1131.

81. Nooijen CF, Stam HJ, Sluis T, Valent L, Twisk J, van den Berg-Emons RJ. A behavioral intervention promoting physical activity in people with subacute spinal cord injury: secondary effects on health, social participation and quality of life. Clin Rehabil. 2017;31: 772–780.

82. Froehlich-Grobe K, Lee J, Ochoa C, Lopez A, Sarker E, Driver S, et al. Effectiveness and feasibility of the workout on wheels internet intervention (WOWii) for individuals with spinal cord injury: a randomized controlled trial. Spinal Cord. 2022;60: 862–874.

83. Bakkum AJT, de Groot S, Stolwijk-Swüste JM, van Kuppevelt DJ, ALLRISC, van der Woude LHV, et al. Effects of hybrid cycling versus handcycling on wheelchair-specific fitness and physical activity in people with long-term spinal cord injury: a 16-week randomized controlled trial. Spinal Cord. 2015;53: 395–401.

84. Capodaglio P, Grilli C, Bazzini G. Tolerable exercise intensity in the early rehabilitation of paraplegic patients. A preliminary study. Spinal Cord. 1996;34: 684–690.

85. Gorgey AS, Lai RE, Khalil RE, Rivers J, Cardozo C, Chen Q, et al. Neuromuscular electrical stimulation resistance training enhances oxygen uptake and ventilatory efficiency independent of mitochondrial complexes after spinal cord injury: a randomized clinical trial. J Appl Physiol. 2021;131: 265–276.

86. Alrashidi AA, Nightingale TE, Currie KD, Hubli M, MacDonald MJ, Hicks AL, et al. Exercise Improves Cardiorespiratory Fitness, but Not Arterial Health, after Spinal Cord Injury: The CHOICES Trial. J Neurotrauma. 2021;38: 3020–3029.

87. Berry HR, Perret C, Saunders BA, Kakebeeke TH, Donaldson NDN, Allan DB, et al. Cardiorespiratory and power adaptations to stimulated cycle training in paraplegia. Med Sci Sports Exerc. 2008;40: 1573–1580.

88. Duffell LD, Paddison S, Alahmary AF, Donaldson N, Burridge J. The effects of FES cycling combined with virtual reality racing biofeedback on voluntary function after incomplete SCI: a pilot study. J Neuroeng Rehabil. 2019;16: 149.

89. Durán FS, Lugo L, Ramírez L, Eusse E. Effects of an exercise program on the rehabilitation of patients with spinal cord injury. Arch Phys Med Rehabil. 2001;82: 1349–1354.

90. Gorman PH, Scott W, York H, Theyagaraj M, Price-Miller N, McQuaid J, et al. Robotically assisted treadmill exercise training for improving peak fitness in chronic motor incomplete spinal cord injury: A randomized controlled trial. J Spinal Cord Med. 2016;39: 32–44.

91. Gibbons RS, Stock CG, Andrews BJ, Gall A, Shave RE. The effect of FES-rowing training on cardiac structure and function: pilot studies in people with spinal cord injury. Spinal Cord. 2016;54: 822–829.

92. Vestergaard M, Jensen K, Juul-Kristensen B. Hybrid high-intensity interval training using functional electrical stimulation leg cycling and arm ski ergometer for people with spinal cord injuries: a feasibility study. Pilot Feasibility Stud. 2022;8: 43.

93. DiCarlo SE. Effect of arm ergometry training on wheelchair propulsion endurance of individuals with quadriplegia. Phys Ther. 1988;68: 40–44.

94. Janssen TWJ, Dallmeijer AJ, Veeger DJHEJ, van der Woude LHV. Normative values and determinants of physical capacity in individuals with spinal cord injury. J Rehabil Res Dev. 2002;39: 29–39.

95. Latimer AE, Ginis KAM, Craven BC, Hicks AL. The physical activity recall assessment for people with spinal cord injury: validity. Med Sci Sports Exerc. 2006;38: 208–216.

96. Manns PJ, McCubbin JA, Williams DP. Fitness, inflammation, and the metabolic syndrome in men with paraplegia. Arch Phys Med Rehabil. 2005;86: 1176–1181.

97. Martin Ginis KA, Úbeda-Colomer J, Alrashidi AA, Nightingale TE, Au JS, Currie KD, et al. Construct validation of the leisure time physical activity questionnaire for people with SCI (LTPAQ-SCI). Spinal Cord. 2021;59: 311–318.

98. Nightingale TE, Walhin J-P, Thompson D, Bilzon JL. Biomarkers of cardiometabolic health are associated with body composition characteristics but not physical activity in persons with spinal cord injury. J Spinal Cord Med. 2019;42: 328–337.

99. Haisma JA, Post MW, van der Woude LH, Stam HJ, Bergen MP, Sluis TA, et al. Functional independence and health-related functional status following spinal cord injury: a prospective study of the association with physical capacity. J Rehabil Med. 2008;40: 812–818.

100. Leving MT, de Groot S, Woldring FAB, Tepper M, Vegter RJK, van der Woude LHV. Motor learning outcomes of handrim wheelchair propulsion during active spinal cord injury rehabilitation in comparison with experienced wheelchair users. Disabil Rehabil. 2021;43: 1429–1442.

101. Stewart MW, Melton-Rogers SL, Morrison S, Figoni SF. The measurement properties of fitness measures and health status for persons with spinal cord injuries. Arch Phys Med Rehabil. 2000;81: 394–400.

102. Dallmeijer AJ, van der Woude LH, Hollander PA, Angenot EL. Physical performance in persons with spinal cord injuries after discharge from rehabilitation. Med Sci Sports Exerc. 1999;31: 1111–1117.

103. Janssen TW, van Oers CA, Rozendaal EP, Willemsen EM, Hollander AP, van der Woude LH. Changes in physical strain and physical capacity in men with spinal cord injuries. Med Sci Sports Exerc. 1996;28: 551–559.

104. Szymczak Ł, Podgórski T, Lewandowski J, Janiak A, Michalak E, Domaszewska K. Physical Fitness and Inflammatory Response to the Training Load of Wheelchair Rugby Players. Int J Environ Res Public Health. 2022;19. doi:10.3390/ijerph19042228

105. Steeves JD, Kramer JK, Fawcett JW, Cragg J, Lammertse DP, Blight AR, et al. Extent of spontaneous motor recovery after traumatic cervical sensorimotor complete spinal cord injury. Spinal Cord. 2011;49: 257–265.

106. Figoni SF, Dolbow DR, Crawford EC, White ML, Pattanaik S. Does aerobic exercise benefit persons with tetraplegia from spinal cord injury? A systematic review. J Spinal Cord Med. 2021;44: 690–703.

107. de Groot S, Dallmeijer AJ, Kilkens OJ, van Asbeck FW, Nene AV, Angenot EL, et al. Course of gross mechanical efficiency in handrim wheelchair propulsion during rehabilitation of people with spinal cord injury: a prospective cohort study. Arch Phys Med Rehabil. 2005;86: 1452–1460.

108. Sprigle S, Maurer C, Holowka M. Development of valid and reliable measures of postural stability. J Spinal Cord Med. 2007;30: 40–49.

109. Kouwijzer I, Valent L, Osterthun R, van der Woude L, de Groot S, HandbikeBattle group. Peak power output in handcycling of individuals with a chronic spinal cord injury: predictive modeling, validation and reference values. Disabil Rehabil. 2020;42: 400–409.

110. Hjeltnes N, Wallberg-Henriksson H. Improved work capacity but unchanged peak oxygen uptake during primary rehabilitation in tetraplegic patients. Spinal Cord. 1998;36: 691–698.

111. Williams AM, Ma JK, Martin Ginis KA, West CR. Effects of a Tailored Physical Activity Intervention on Cardiovascular Structure and Function in Individuals With Spinal Cord Injury. Neurorehabil Neural Repair. 2021;35: 692–703.

112. Cruz S, Blauwet CA. Implications of altered autonomic control on sports performance in athletes with spinal cord injury. Auton Neurosci. 2018;209: 100–104.

113. West CR, Gee CM, Voss C, Hubli M, Currie KD, Schmid J, et al. Cardiovascular control, autonomic function, and elite endurance performance in spinal cord injury. Scand J Med Sci Sports. 2015;25: 476–485.

114. West CR, Campbell IG, Goosey-Tolfrey VL, Mason BS, Romer LM. Effects of abdominal binding on field-based exercise responses in Paralympic athletes with cervical spinal cord injury. J Sci Med Sport. 2014;17: 351–355.

115. Pitetti KH, Barrett PJ, Campbell KD, Malzahn DE. The effect of lower body positive pressure on the exercise capacity of individuals with spinal cord injury. Med Sci Sports Exerc. 1994;26: 463–468.

116. Nieshoff EC, Birk TJ, Birk CA, Hinderer SR, Yavuzer G. Double-blinded, placebo-controlled trial of midodrine for exercise performance enhancement in tetraplegia: a pilot study. J Spinal Cord Med. 2004;27: 219–225.

117. Nightingale TE, Walter M, Williams AMM, Lam T, Krassioukov AV. Ergogenic effects of an epidural neuroprosthesis in one individual with spinal cord injury. Neurology. 2019;92: 338–340.

118. Laskin JJ, Waheed Z, Thorogood NP, Nightingale TE, Noonan VK. Spinal Cord Stimulation Research in the Restoration of Motor, Sensory, and Autonomic Function for Individuals Living With Spinal Cord Injuries: A Scoping Review. Arch Phys Med Rehabil. 2022. doi:10.1016/j.apmr.2022.01.161

119. Berger MJ, Kimpinski K, Currie KD, Nouraei H, Sadeghi M, Krassioukov AV. Multi-Domain Assessment of Autonomic Function in Spinal Cord Injury Using a Modified Autonomic Reflex Screen. J Neurotrauma. 2017;34: 2624–2633.

120. Wang S, Wecht JM, Legg Ditterline B, Ugiliweneza B, Maher MT, Lombard AT, et al. Heart rate and blood pressure response improve the prediction of orthostatic cardiovascular dysregulation in persons with chronic spinal cord injury. Physiol Rep. 2020;8: e14617.

121. DiPiro ND, Embry AE, Fritz SL, Middleton A, Krause JS, Gregory CM. Effects of aerobic exercise training on fitness and walking-related outcomes in ambulatory individuals with chronic incomplete spinal cord injury. Spinal Cord. 2016;54: 675–681.

122. Fleg JL, Morrell CH, Bos AG, Brant LJ, Talbot LA, Wright JG, et al. Accelerated longitudinal decline of aerobic capacity in healthy older adults. Circulation. 2005;112: 674–682.

123. Groah SL, Charlifue S, Tate D, Jensen MP, Molton IR, Forchheimer M, et al. Spinal cord injury and aging: challenges and recommendations for future research. Am J Phys Med Rehabil. 2012;91: 80–93.

124. Hinrichs T, Lay V, Arnet U, Eriks-Hoogland I, Koch HG, Rantanen T, et al. Age-related variation in mobility independence among wheelchair users with spinal cord injury: A cross-sectional study. J Spinal Cord Med. 2016;39: 180–189.

125. Jörgensen S, Martin Ginis KA, Lexell J. Leisure time physical activity among older adults with long-term spinal cord injury. Spinal Cord. 2017;55: 848–856.

126. Chiou SY, Clarke E, Lam C, Harvey T, Nightingale TE. Effects of Arm-Crank Exercise on Fitness and Health in Adults With Chronic Spinal Cord Injury: A Systematic Review. Front Physiol. 2022;13: 831372.

127. Bochkezanian V, Raymond J, de Oliveira CQ, Davis GM. Can combined aerobic and muscle strength training improve aerobic fitness, muscle strength, function and quality of life in people with spinal cord injury? A systematic review. Spinal Cord. 2015;53: 418–431.

128. Brurok B, Helgerud J, Karlsen T, Leivseth G, Hoff J. Effect of aerobic high-intensity hybrid training on stroke volume and peak oxygen consumption in men with spinal cord injury. Am J Phys Med Rehabil. 2011;90: 407–414.

129. Gant KL, Nagle KG, Cowan RE, Field-Fote EC, Nash MS, Kressler J, et al. Body System Effects of a Multi-Modal Training Program Targeting Chronic, Motor Complete Thoracic Spinal Cord Injury. J Neurotrauma. 2018;35: 411–423.

130. Lotter JK, Henderson CE, Plawecki A, Holthus ME, Lucas EH, Ardestani MM, et al. Task-Specific Versus Impairment-Based Training on Locomotor Performance in Individuals With Chronic Spinal Cord Injury: A Randomized Crossover Study. Neurorehabil Neural Repair. 2020;34: 627–639.

131. Jacobs PL, Nash MS, Rusinowski JW. Circuit training provides cardiorespiratory and strength benefits in persons with paraplegia. Med Sci Sports Exerc. 2001;33: 711–717.

132. Sweet SN, Martin Ginis KA, Tomasone JR. Investigating intermediary variables in the physical activity and quality of life relationship in persons with spinal cord injury. Health Psychol. 2013;32: 877–885.

133. Eitivipart AC, de Oliveira CQ, Arora M, Middleton J, Davis GM. Overview of Systematic Reviews of Aerobic Fitness and Muscle Strength Training after Spinal Cord Injury. J Neurotrauma. 2019;36: 2943–2963.

134. Martin Ginis KA, van der Scheer JW, Latimer-Cheung AE, Barrow A, Bourne C, Carruthers P, et al. Response to correspondence from the ESSA Statement authors. Spinal cord. 2018. pp. 409–411.

135. Astorino TA, Thum JS. Within-session responses to high-intensity interval training in spinal cord injury. Disabil Rehabil. 2018;40: 444–449.

136. Tordi N, Dugue B, Klupzinski D, Rasseneur L, Rouillon JD, Lonsdorfer J. Interval training program on a wheelchair ergometer for paraplegic subjects. Spinal Cord. 2001;39: 532–537.

137. de Groot PCE, Hjeltnes N, Heijboer AC, Stal W, Birkeland K. Effect of training intensity on physical capacity, lipid profile and insulin sensitivity in early rehabilitation of spinal cord injured individuals. Spinal Cord. 2003;41: 673–679.

138. Gauthier C, Brosseau R, Hicks AL, Gagnon DH. Feasibility, Safety, and Preliminary Effectiveness of a Home-Based Self-Managed High-Intensity Interval Training Program Offered to Long-Term Manual Wheelchair Users. Rehabil Res Pract. 2018;2018: 8209360.

139. Mcleod JC, Diana H, Hicks AL. Sprint interval training versus moderate-intensity continuous training during inpatient rehabilitation after spinal cord injury: a randomized trial. Spinal Cord. 2020;58: 106–115.

140. Graham K, Yarar-Fisher C, Li J, McCully KM, Rimmer JH, Powell D, et al. Effects of High-Intensity Interval Training Versus Moderate-Intensity Training on Cardiometabolic Health Markers in Individuals With Spinal Cord Injury: A Pilot Study. Top Spinal Cord Inj Rehabil. 2019;25: 248–259.

141. McMillan DW, Astorino TA, Correa MA, Nash MS, Gater DR. Virtual Strategies for the Broad Delivery of High Intensity Exercise in Persons With Spinal Cord Injury: Ongoing Studies and Considerations for Implementation. Front Sports Act Living. 2021;3: 703816.

142. Maher JL, Whitmarsh C, Smith P, Taylor H, Fard A, Bilzon J. Feasibility study of high-intensity interval training to reduce cardiometabolic disease risks in individuals with acute spinal cord injury. BMJ Open. 2023;13: e068507.

143. Claydon VE, Hol AT, Eng JJ, Krassioukov AV. Cardiovascular responses and postexercise hypotension after arm cycling exercise in subjects with spinal cord injury. Arch Phys Med Rehabil. 2006;87: 1106–1114.

144. Griggs KE, Leicht CA, Price MJ, Goosey-Tolfrey VL. Thermoregulation during intermittent exercise in athletes with a spinal-cord injury. Int J Sports Physiol Perform. 2015;10: 469–475.

145. Weston KS, Wisløff U, Coombes JS. High-intensity interval training in patients with lifestyle-induced cardiometabolic disease: a systematic review and meta-analysis. Br J Sports Med. 2014;48: 1227–1234.

146. Valentino SE, Hutchinson MJ, Goosey-Tolfrey VL, MacDonald MJ. Effects of Perceptually Regulated Exercise Training on Cardiorespiratory Fitness and Peak Power Output in Adults With Spinal Cord Injury: A Systematic Review and Meta-analysis. Arch Phys Med Rehabil. 2022. doi:10.1016/j.apmr.2022.03.008

147. Hutchinson MJ, Kouwijzer I, de Groot S, Goosey-Tolfrey VL. Comparison of two Borg exertion scales for monitoring exercise intensity in able-bodied participants, and those with paraplegia and tetraplegia. Spinal Cord. 2021;59: 1162–1169.

148. van der Scheer JW, Hutchinson MJ, Paulson T, Martin Ginis KA, Goosey-Tolfrey VL. Reliability and Validity of Subjective Measures of Aerobic Intensity in Adults With Spinal Cord Injury: A Systematic Review. PM R. 2018;10: 194–207.

149. Cowley KC. A new conceptual framework for the integrated neural control of locomotor and sympathetic function: implications for exercise after spinal cord injury. Appl Physiol Nutr Metab. 2018;43: 1140–1150.

150. Jamnick NA, Pettitt RW, Granata C, Pyne DB, Bishop DJ. An Examination and Critique of Current Methods to Determine Exercise Intensity. Sports Med. 2020;50: 1729–1756.

151. Meyler S, Bottoms L, Wellsted D, Muniz-Pumares D. Variability in exercise tolerance and physiological responses to exercise prescribed relative to physiological thresholds and to maximum oxygen uptake. Exp Physiol. 2023;108: 581–594.

152. Anselmi F, Cavigli L, Pagliaro A, Valente S, Valentini F, Cameli M, et al. The importance of ventilatory thresholds to define aerobic exercise intensity in cardiac patients and healthy subjects. Scand J Med Sci Sports. 2021;31: 1796–1808.

153. Au JS, Sithamparapillai A, Currie KD, Krassioukov AV, MacDonald MJ, Hicks AL. Assessing Ventilatory Threshold in Individuals With Motor-Complete Spinal Cord Injury. Arch Phys Med Rehabil. 2018;99: 1991–1997.

154. Kouwijzer I, Cowan RE, Maher JL, Groot FP, Riedstra F, Valent LJM, et al. Interrater and intrarater reliability of ventilatory thresholds determined in individuals with spinal cord injury. Spinal Cord. 2019;57: 669–678.

155. Tjønna AE, Leinan IM, Bartnes AT, Jenssen BM, Gibala MJ, Winett RA, et al. Low- and high-volume of intensive endurance training significantly improves maximal oxygen uptake after 10-weeks of training in healthy men. PLoS One. 2013;8: e65382.

156. Stensvold D, Viken H, Steinshamn SL, Dalen H, Støylen A, Loennechen JP, et al. Effect of exercise training for five years on all cause mortality in older adults-the Generation 100 study: randomised controlled trial. BMJ. 2020;371: m3485.

157. Church TS, Earnest CP, Skinner JS, Blair SN. Effects of different doses of physical activity on cardiorespiratory fitness among sedentary, overweight or obese postmenopausal women with elevated blood pressure: a randomized controlled trial. JAMA. 2007;297: 2081–2091.

158. Foulds HJA, Bredin SSD, Charlesworth SA, Ivey AC, Warburton DER. Exercise volume and intensity: a dose-response relationship with health benefits. Eur J Appl Physiol. 2014;114: 1563–1571.

159. Alrashidi AA, Nightingale TE, Bhangu GS, Bissonnette-Blais V, Krassioukov AV. Post-processing of peak oxygen uptake data obtained during cardiopulmonary exercise testing in individuals with spinal cord injury: A scoping review and analysis of different post-processing strategies. Arch Phys Med Rehabil. 2022. doi:10.1016/j.apmr.2022.11.015

160. Phillips BE, Kelly BM, Lilja M, Ponce-González JG, Brogan RJ, Morris DL, et al. A Practical and Time-Efficient High-Intensity Interval Training Program Modifies Cardio-Metabolic Risk Factors in Adults with Risk Factors for Type II Diabetes. Front Endocrinol. 2017;8: 229.

161. Katzelnick CG, Weir JP, Jones A, Galea M, Dyson-Hudson TA, Kirshblum SC, et al. Blood Pressure Instability in Persons With SCI: Evidence From a 30-Day Home Monitoring Observation. Am J Hypertens. 2019;32: 938–944.

162. Nightingale TE, Eginyan G, Balthazaar SJT, Williams AMM, Lam T, Krassioukov AV. Accidental boosting in an individual with tetraplegia - considerations for the interpretation of cardiopulmonary exercise testing. J Spinal Cord Med. 2021; 1–6.

163. Martin Ginis KA, Phang SH, Latimer AE, Arbour-Nicitopoulos KP. Reliability and validity tests of the leisure time physical activity questionnaire for people with spinal cord injury. Arch Phys Med Rehabil. 2012;93: 677–682.

164. Ginis KAM, Latimer AE, Hicks AL, Craven BC. Development and evaluation of an activity measure for people with spinal cord injury. Med Sci Sports Exerc. 2005;37: 1099–1111.

165. de Groot S, van der Woude LHV, Niezen A, Smit CAJ, Post MWM. Evaluation of the physical activity scale for individuals with physical disabilities in people with spinal cord injury. Spinal Cord. 2010;48: 542–547.

166. Healy GN, Wijndaele K, Dunstan DW, Shaw JE, Salmon J, Zimmet PZ, et al. Objectively measured sedentary time, physical activity, and metabolic risk: the Australian Diabetes, Obesity and Lifestyle Study (AusDiab). Diabetes Care. 2008;31: 369–371.

167. O’Donovan G, Hillsdon M, Ukoumunne OC, Stamatakis E, Hamer M. Objectively measured physical activity, cardiorespiratory fitness and cardiometabolic risk factors in the Health Survey for England. Prev Med. 2013;57: 201–205.

168. Hansford HJ, Wewege MA, Cashin AG, Hagstrom AD, Clifford BK, McAuley JH, et al. If exercise is medicine, why don’t we know the dose? An overview of systematic reviews assessing reporting quality of exercise interventions in health and disease. Br J Sports Med. 2022;56: 692– 700.

169. Slade SC, Dionne CE, Underwood M, Buchbinder R. Consensus on Exercise Reporting Template (CERT): Explanation and Elaboration Statement. Br J Sports Med. 2016;50: 1428–1437.

170. Hoffmann TC, Glasziou PP, Boutron I, Milne R, Perera R, Moher D, et al. Better reporting of interventions: template for intervention description and replication (TIDieR) checklist and guide. BMJ. 2014;348: g1687.

171. Kodama S, Saito K, Tanaka S, Maki M, Yachi Y, Asumi M, et al. Cardiorespiratory fitness as a quantitative predictor of all-cause mortality and cardiovascular events in healthy men and women: a meta-analysis. JAMA. 2009;301: 2024–2035.

