## Supplementary material for "The effect of exercise on aerobic capacity in individuals with spinal cord injury: A systematic review with meta-analysis and meta-regression": S2 Search strategy

**Electronic Supplementary Material 2**

**PLoS Medicine**

Hodgkiss, D.D^1^, Bhangu, G^2,3^, Lunny, C^4^, Jutzeler C.R^5,6^, Chiou S.Y^1,7,8,9^ Walter, M^2,10^, Lucas S.E^1,7^, Krassioukov, A.V.^2,11,12^, Nightingale, T.E.^1,2,9^

**^1^** School of Sport, Exercise and Rehabilitation Sciences, University of Birmingham, UK.

**^2^** International Collaboration on Repair Discoveries (ICORD), University of British Columbia, Vancouver, British Columbia, Canada. **^3^** MD Undergraduate Program, Faculty of Medicine, University of British Columbia, Vancouver, Canada. **^4^** Knowledge Translation Program, Li Ka Shing Knowledge Institute, St. Michael’s Hospital, Toronto, and the University of British Columbia, Vancouver, British Columbia, Canada.

**^5^** Department of Health Sciences and Technology, ETH Zurich, Zurich, Switzerland. **^6^** Schulthess Clinic, Zurich, Switzerland. **^7^** Centre for Human Brain Health, University of Birmingham, United Kingdom. **^8^** MRC Versus Arthritis Centre for Musculoskeletal Ageing Research, University of Birmingham, United Kingdom.

**^9^** Centre for Trauma Science Research, University of Birmingham, United Kingdom. **^10^** Department of Urology, University Hospital Basel, University of Basel, Basel, Switzerland. **^11^** Department of Medicine, Division of Physical Medicine and Rehabilitation, University of British Columbia, Vancouver, British Columbia, Canada. **^12^** GF Strong Rehabilitation Centre, Vancouver Coastal Health, Vancouver, British Columbia, Canada.

**Table 1**: Example of a search strategy (Ovid: Medline and Embase).

| **#** | **Searches** | **Results** |
| --- | --- | --- |
| Search keywords for spinal cord injury | | |
| 1 | tetrapleg*.mp. | 10968 |
| 2 | parapleg*.mp. | 57254 |
| 3 | quadripleg*.mp. | 30436 |
| 4 | spinal cord injur*.mp. | 123750 |
| 5 | spinal cord lesion*.mp. | 12285 |
| 6 | spinal cord transection*.mp. | 3300 |
| 7 | spinal cord impair*.mp. | 239 |
| 8 | spinal injur*.mp. | 22487 |
| 9 | spinal lesion*.mp. | 4693 |
| 10 | spinal transection*.mp. | 1631 |
| 11 | spinal impairm*.mp. | 60 |
| 12 | brown-sequard syndrome.mp. | 1606 |
| 13 | central cord.mp. | 1169 |
| 14 | myelitis.mp. | 19043 |
| 15 | spinal cord diseas*.mp. | 29885 |
| 16 | myelopath*.mp. | 35505 |
| 17 | spinal paraly*.mp. | 570 |
| 18 | hemipleg*.mp. | 41532 |
| 19 | syringomy*.mp. | 11672 |
| 20 | 1 or 2 or 3 or 4 or 5 or 6 or 7 or 8 or 9 or 10 or 11 or 12 or 13 or 14 or 15 or 16 or 17 or 18 or 19 | 319022 |
| Search keywords for exercise/fitness | | |
| 21 | exercise*.mp. | 968425 |
| 22 | aerobic exercise*.mp. | 34799 |
| 23 | exercise condition*.mp. | 3285 |
| 24 | exercise prescription*.mp. | 6029 |
| 25 | exercise therap*.mp. | 52021 |
| 26 | exercise train*.mp. | 44870 |
| 27 | physical activit*.mp. | 363264 |
| 28 | sport*.mp. | 271414 |
| 29 | strength train*.mp. | 13970 |
| 30 | resistance train*.mp. | 40147 |
| 31 | endurance exercise*.mp. | 10884 |
| 32 | endurance train*.mp. | 19681 |
| 33 | interval train*.mp. | 9012 |
| 34 | activity level.mp. | 32014 |
| 35 | neuromuscular electrical stimulation*.mp. | 4086 |
| 36 | functional electrical stimulation*.mp. | 6793 |
| 37 | power output*.mp. | 18842 |
| 38 | cardiorespiratory fitness.mp. | 16131 |
| 39 | peak oxygen uptake.mp. | 9896 |
| 40 | peak oxygen consumption*.mp. | 7686 |
| 41 | functional capacity.mp. | 36814 |
| 42 | aerobic capacity.mp. | 21507 |
| 43 | 21 or 22 or 23 or 24 or 25 or 26 or 27 or 28 or 29 or 30 or 31 or 32 or 33 or 34 or 35 or 36 or 37 or 38 or 39 or 40 or 41 or 42 | 1456041 |
| Combined search for spinal cord injury and exercise/fitness keywords | | |
| 44 | 20 and 33 | 17786 |

**Table 2**: Example of a search strategy (Ovid: Central)

| 1 | (Tetraplegia or paraplegia or Quadriplegia or spinal cord injury or spinal cord lesion or spinal cord transection or spinal cord impairment or spinal injury or spinal lesion or spinal transection or spinal impairment or brown-sequard syndrome or central cord or myelitis or spinal cord disease or myelopathy or spinal paralysis or hemiplegia or Syringomyelia).mp. [mp=title, original title, abstract, mesh headings, heading words, keyword] | 7098 |
| --- | --- | --- |
| 2 | (Exercise or aerobic exercise or exercise conditioning or exercise prescription or exercise therapy or exercise training or physical activity or sport or strength training or resistance training or endurance exercise or endurance training or interval training or neuromuscular electrical stimulation or Functional electrical stimulation or activity level).mp. [mp=title, original title, abstract, mesh headings, heading words, keyword] | 131064 |
| 3 | (power output or cardiorespiratory fitness or peak oxygen uptake or peak oxygen consumption or functional capacity or aerobic capacity).mp. [mp=title, original title, abstract, mesh headings, heading words, keyword] | 12654 |
| 4 | 1 and 2 and 3 | 131 |

**Table 3**: Example of a search strategy (Web of Science)

| 1 | TS=(Tetraplegia or paraplegia or Quadriplegia or spinal cord injury or spinal cord lesion or spinal cord transection or spinal cord impairment or spinal injury or spinal lesion or spinal transection or spinal impairment or brown-sequard syndrome or central cord or myelitis or spinal cord disease or myelopathy or spinal paralysis or hemiplegia or Syringomyelia) | 198360 |
| --- | --- | --- |
| 2 | TS=(Exercise or aerobic exercise or exercise conditioning or exercise prescription or exercise therapy or exercise training or physical activity or sport or strength training or resistance training or endurance exercise or endurance training or interval training or neuromuscular electrical stimulation or Functional electrical stimulation or activity level) | 2245923 |
| 3 | TS=(power output or cardiorespiratory fitness or peak oxygen uptake or peak oxygen consumption or functional capacity or aerobic capacity) | 389028 |
| 4 | 1 and 2 and 3 | 1093 |
