## Supplementary material for "The effect of exercise on aerobic capacity in individuals with spinal cord injury: A systematic review with meta-analysis and meta-regression": S4 Correlation factors

**Electronic Supplementary Material 4**

**PLoS Medicine**

Hodgkiss, D.D^1^, Bhangu, G^2,3^, Lunny, C^4^, Jutzeler C.R^5,6^, Chiou S.Y^1,7,8,9^ Walter, M^2,10^, Lucas S.E^1,7^, Krassioukov, A.V.^2,11,12^, Nightingale, T.E.^1,2,9^

**^1^** School of Sport, Exercise and Rehabilitation Sciences, University of Birmingham, UK.

**^2^** International Collaboration on Repair Discoveries (ICORD), University of British Columbia, Vancouver, British Columbia, Canada. **^3^** MD Undergraduate Program, Faculty of Medicine, University of British Columbia, Vancouver, Canada. **^4^** Knowledge Translation Program, Li Ka Shing Knowledge Institute, St. Michael’s Hospital, Toronto, and the University of British Columbia, Vancouver, British Columbia, Canada. **^5^** Department of Health Sciences and Technology, ETH Zurich, Zurich, Switzerland. **^6^** Schulthess Clinic, Zurich, Switzerland. **^7^** Centre for Human Brain Health, University of Birmingham, United Kingdom. **^8^** MRC Versus Arthritis Centre for Musculoskeletal Ageing Research, University of Birmingham, United Kingdom. **^9^** Centre for Trauma Science Research, University of Birmingham, United Kingdom. **^10^** Department of Urology, University Hospital Basel, University of Basel, Basel, Switzerland. **^11^** Department of Medicine, Division of Physical Medicine and Rehabilitation, University of British Columbia, Vancouver, British Columbia, Canada. **^12^** GF Strong Rehabilitation Centre, Vancouver Coastal Health, Vancouver, British Columbia, Canada.

| **Table 1.** The correlation coefficients averaged across each study design for each cardiorespiratory fitness outcome. | | | |
| --- | --- | --- | --- |
|  | **AV̇O_2peak_** | **RV̇O_2peak_** | **PPO** |
| Observational studies | 0.87 | 0.92 | 0.92 |
| RCT controls | 0.98 | 0.99 | 0.99 |
| RCT exercise interventions | 0.97 | 0.96 | 0.95 |
| Combined pre-post and RCT exercise interventions | 0.87 | 0.92 | 0.95 |

AV̇O_2peak_, absolute peak oxygen consumption; PPO, peak power output; RCT, randomised-controlled trial; RV̇O_2peak_, relative peak oxygen consumption.
