## Supplementary material for "The effect of exercise on aerobic capacity in individuals with spinal cord injury: A systematic review with meta-analysis and meta-regression": S5 Primary meta-analysis (RCTs)

**Electronic Supplementary Material 5**

**PLoS Medicine**

Hodgkiss, D.D^1^, Bhangu, G^2,3^, Lunny, C^4^, Jutzeler C.R^5,6^, Chiou S.Y^1,7,8,9^ Walter, M^2,10^, Lucas S.E^1,7^, Krassioukov, A.V.^2,11,12^, Nightingale, T.E.^1,2,9^

**^1^** School of Sport, Exercise and Rehabilitation Sciences, University of Birmingham, UK.

**^2^** International Collaboration on Repair Discoveries (ICORD), University of British Columbia, Vancouver, British Columbia, Canada. **^3^** MD Undergraduate Program, Faculty of Medicine, University of British Columbia, Vancouver, Canada. **^4^** Knowledge Translation Program, Li Ka Shing Knowledge Institute, St. Michael’s Hospital, Toronto, and the University of British Columbia, Vancouver, British Columbia, Canada.

**^5^** Department of Health Sciences and Technology, ETH Zurich, Zurich, Switzerland. **^6^** Schulthess Clinic, Zurich, Switzerland. **^7^** Centre for Human Brain Health, University of Birmingham, United Kingdom. **^8^** MRC Versus Arthritis Centre for Musculoskeletal Ageing Research, University of Birmingham, United Kingdom.

**^9^** Centre for Trauma Science Research, University of Birmingham, United Kingdom. **^10^** Department of Urology, University Hospital Basel, University of Basel, Basel, Switzerland. **^11^** Department of Medicine, Division of Physical Medicine and Rehabilitation, University of British Columbia, Vancouver, British Columbia, Canada. **^12^** GF Strong Rehabilitation Centre, Vancouver Coastal Health, Vancouver, British Columbia, Canada.

***Purpose:*** This supplementary file includes the pooled and individual participant demographics, injury characteristics and exercise intervention parameters of randomised-controlled trials (RCTs) assessing the change in cardiorespiratory fitness (CRF) following an exercise or physical activity intervention relative to a control or standard of care intervention.

***Conclusion:*** There are significantly greater changes in absolute (AV̇O_2peak_) and relative peak oxygen consumption (RV̇O_2peak_) and peak power output (PPO) in favour of exercise interventions in comparison to control interventions. The pooled effect estimates for RV̇O_2peak_ and PPO are very similar to those reported in the secondary, pooled meta-analysis and therefore confirms the robustness of our findings in this meta-analysis.

| **Table 1:** Characteristics of randomised controlled trials (exercise intervention and controls arms) reporting on each CRF outcome. | | | | | | |
| --- | --- | --- | --- | --- | --- | --- |
|  | **Cardiorespiratory fitness outcome**  **Total number of interventions [sum of participants]**  **Mean (range)** | | | | | |
|  | AV̇O_2peak_ (L/min) | | RV̇O_2peak_ (mL/kg/min) | | PPO (W) | |
|  | Intervention | Control | Intervention | Control | Intervention | Control |
|  | 12 [181] | 12 [168] | 16 [232] | 16 [201] | 11 [247] | 9 [135] |
| *Participant demographics* | | | | | | |
| Age (years) | 44 (27 – 54) | 46 (33 – 55) | 45 (27 – 56) | 46 (30 – 53) | 46 (31 – 56) | 46 (39 – 55) |
| Baseline CRF | 1.41 (0.51 – 3.50) | 1.41 (0.49 – 3.20) | 17.4 (10.7 – 28.5) | 16.8 (9.4 – 24.3) | 52 (7 – 82) | 54 (16 – 75) |
| *Sex* | | | | | | |
| Male, n (%) | 3 [26] | 2 [12] | 6 [45] | 4 [24] | 1 [12] | - |
| Female | - | - | - | - | - | - |
| Mixed (% F) | 9 [155] (25%) | 10 [149] (28%) | 8 [164] (33%) | 10 [165] (41%) | 10 [235] (30%) | 9 [135] (27%) |
| Not reported/cannot determine | - | - | 2 [23] | 2 [12] | - | - |
| *Injury characteristics* | | | | | | |
| *Time since injury (years)* | 13 (0 – 21) | 13 (0 – 23) | 12 (1 – 24) | 13 (1 – 19) | 14 (0 – 21) | 15 (0 – 23) |
| Acute (<1-year) | 1 [20] | 1 [19] | 1 [4] | 1 [4] | 1 [20] | 1 [19] |
| Chronic (>1-year) | 11 [161] | 11 [149] | 12 [116] | 12 [92] | 8 [123] | 8 [107] |
| Mixed (% acute) | - | - | - | - | - | - |
| Not reported/cannot determine | - | - | 3 [112] | 3 [105] | 2 [94] | 2 [97] |
| *Neurological level of injury (TETRA/PARA)* | | | | | | |
| TETRA | - | - | - | 2 [13] | - | - |
| PARA | 3 [29] | 3 [22] | 7 [66] | 5 [34] | 3 [39] | 1 [8] |
| Mixed (% PARA) | 6 [111] (67%) | 6 [102] (64%) | 4 [40] (48%) | 4 [43] (70%) | 5 [101] (66%) | 7 [116] (70%) |
| Not reported/cannot determine | 3 [41] | 3 [39] | 5 [126] | 5 [113] | 3 [107] | 3 [99] |
| *Severity* | | | | | | |
| Motor-complete | - | - | 4 [27] | 4 [24] | - | - |
| Motor-incomplete | 1 [10] | 1 [10] | 2 [20] | 2 [14] | - | - |
| Mixed (% incomp.) | 7 [122] (34%) | 8 [117] (32%) | 5 [60] (47%) | 5 [55] (44%) | 8 [138] (35%) | 6 [126] (33%) |
| Not reported/cannot determine | 3 [41] | 3 [41] | 5 [125] | 5 [115] | 3 [109] | 3 [97] |
| *Exercise intervention parameters* | | | | | | |
| *Type* | | | | | | |
| Upper-body aerobic exercise | 4 [55] |  | 6 [58] |  | 3 [46] |  |
| Upper-body resistance training/circuits | 1 [9] |  | 1 [9] |  | - |  |
| Functional electrical stimulation | 1 [20] |  | - |  | 1 [20] |  |
| Gait/locomotor training | 1 [10 |  | 2 [20] |  | - |  |
| Mixed/multimodal | 2 [20] |  | 4 [37] |  | 2 [20] |  |
| Behaviour change | 3 [67] |  | 3 [108] |  | 5 [161] |  |
| *Relative intensity* | | | | | | |
| Light | 1 [14] |  | - |  | 1 [14] |  |
| Moderate | - |  | - |  | - |  |
| Moderate-to-vigorous | 4 [56] |  | 7 [80] |  | 4 [52] |  |
| Vigorous | 2 [14] |  | 4 [32] |  | - |  |
| Supramaximal | - |  | - |  | - |  |
| Mixed/cannot determine | 5 [97] |  | 5 [120] |  | 6 [181] |  |
| *Relative intensity prescription* | | | | | | |
| VO_2_ (%peak, %reserve) | 1 [15] |  | 3 [38] |  | 2 [32] |  |
| Heart rate (%HRR, %HR_peak_, _-%HR_max_) | 2 [26] |  | 3 [26] |  | - |  |
| RPE | 1 [8] |  | 3 [23] |  | 1 [8] |  |
| Workload (%PPO, %MTP, __ _  -_%1RM) | 1 [9] |  | 1 [9] |  | - |  |
| Mixed/cannot determine | 7 [123] |  | 6 [136] |  | 8 [207] |  |
| *Session duration (min)* | 49 (20 – 90) |  | 38 (15 – 60) |  | 38 (20 – 60) |  |
| *Frequency (sessions/week)* | 3 (2 – 5) |  | 3 (2 – 5) |  | 3 (2 – 4) |  |
| < 3 | 4 [67] |  | 1 [12] |  | 3 [46] |  |
| ≥ 3 and < 5 | 3 [32] |  | 10 [103] |  | 3 [40] |  |
| ≥ 5 | 2 [15] |  | 2 [9] |  | - |  |
| Not reported | 3 [67] |  | 3 [108] |  | 5 [161] |  |
| *Volume (min/week)* | 151 (40 – 450) |  | 121 (40 – 180) |  | 101 (40 – 180) |  |
| SCI-specific exercise guidelines [fitness (40 – 89 min/wk)] | 2 [26]  51 (40 – 60) |  | 2 [18]  42 (40 – 45) |  | 2 [26]  51 (40 – 60) |  |
| SCI-specific exercise guidelines [cardiometabolic (90 – 149 min/wk)] | 2 [28]  111 (90 – 120) |  | 5 [54]  96 (90 – 116) |  | 3 [45]  103 (90 – 120) |  |
| Achieving general population exercise guidelines (≥150 min/wk) | 5 [60]  212 (150 – 450) |  | 6 [52]  175 (150 – 180) |  | 1 [15]  180 (N/A) |  |
| Cannot classify | 3 [67] |  | 3 [108] |  | 5 [161] |  |
| *Length (weeks)* | 22 (6 – 52) |  | 12 (6 – 24) |  | 21 (6 – 52) |  |
| ≤ 6 weeks | 2 [24] |  | 5 [47] |  | 1 [15] |  |
| > 6 and ≤ 12 weeks | 5 [57] |  | 8 [79] |  | 1 [59] |  |
| > 12 weeks | 5 [100] |  | 3 [106] |  | 6 [173] |  |
| *Adverse events reported* | | | | | | |
| Bone, joint or muscular pain | - |  | - |  | - |  |
| Autonomic or cardiovascular ...function | - |  | - |  | - |  |
| Skin irritation or pressure sores | - |  | 1 [2] |  | - |  |
| Other ^a^ | - |  | 1 [1] |  | 1 [1] |  |
| Total number of studies (N) and participants [Σ] along with descriptive characteristics for RCTs included in this systematic review that describes Δ in CRF outcomes in response to an exercise intervention or a true-world control or standard of care arm. Continuous variables are displayed as weighted means (range: lowest – highest mean values reported from studies). Categorical variables are displayed as n (%). Weighted means were calculated to account for differences in sample size between studies using the following formula: Σn*x̅ /Σn, where Σ = the sum of, n = number of participants in each study and, x̅ = mean CRF outcome of each study. F, females; HR _max_, maximal heart rate; HR _peak_, peak heart rate; HRR, heart rate reserve; 1RM, one repetition maximum; M, males; MTP, maximal tolerated power; NR, not reported; PARA, paraplegia; PPO, peak power output; TETRA, tetraplegia; V̇O_2 peak_, peak oxygen consumption; W, watts. ^a^ Other adverse events included: anxiety, nausea, dizziness and issues with testing equipment. | | | | | | |

| **Table 2:** Summaries of individual randomised controlled trials. | | | | |
| --- | --- | --- | --- | --- |
| **RANDOMISED-CONTROLLED TRIALS** | | | | |
| **Author/Year/**  **Country** | **Group** | **Population** | **Training Details** | **Cardiorespiratory Fitness Outcomes** |
| Akkurt et al. (2017)  Turkey | Intervention | *N =* 17 (16 M/ 1 F)    *Age =* 31 ± 7.8 years  *TSI =* 3.7 ± 3.5 years (no. acute/chronic NR)  *Classification =* 0 T/ 17 P  *Severity =* 10 comp./ 7 incomp.  *CPET =* Progressive ACE  *CPET same modality of intervention? =* Yes | *Type of Exercise =* ACE  *Relative Intensity and Class =* Moderate-to-vigorous (50-70% VO_2peak_)  *Session Duration (min) =* 30  *Frequency (times/week) =* 3  *Intervention Length (weeks) =* 12  *Adverse Events =* NR | *AV̇O_2peak_ (L/min)*   - *Pre:* NR - *Post:* NR   *RV̇O_2peak_ (mL/kg/min)*   - *Pre:* 19.9 ± 8.8 - *Post:* 23.9 ± 5.1   *PPO (W)*   - *Pre:* 72 ±16 - *Post:* 103 ± 23 |
|  | Control | *N =* 16 (13 M/ 3 F)    *Age =* 38.8 ± 12.4 years  *TSI =* 3.2 ± 2.8 years (no. acute/chronic NR)  *Classification =* 1 T/ 15 P  *Severity =* 10 comp./ 6 incomp.  *CPET =* Progressive ACE | General exercises (including locomotor training, 50% 1RM strength training, ROM exercises) | *AV̇O_2peak_ (L/min)*   - *Pre:* NR - *Post:* NR   *RV̇O_2peak_ (mL/kg/min)*   - *Pre:* 18.4 ± 5.8 - *Post:* 16.8 ± 3.3   *PPO (W)*   - *Pre:* 67 ± 28 - *Post:* 78 ± 31 |
| Capodaglio et al. (1996)  Italy | Intervention | *N =* 4 (4 M/ 0 F)    *Age =* 31.8 ± 11.0 years  *TSI =* CD (4 acute/ 0 chronic)  *Classification =* 0 T/ 4 P  *Severity =* 4 comp./ 0 incomp.  *CPET =* Progressive ACE  *CPET same modality of intervention? =* Yes | *Type of Exercise =* ACE in addition to conventional rehabilitation  *Relative Intensity and Class =* Mixed/cannot determine (‘moderate subjective perception of effort’)  *Session Duration (min) =* 20-30  *Frequency (times/week) =* 5  *Intervention Length (weeks) =* 6  *Adverse Events =* NR | *AV̇O_2peak_ (L/min)*   - *Pre:* NR - *Post:* NR   *RV̇O_2peak_ (mL/kg/min)*   - *Pre:* 11.0 ± 1.6 - *Post:* 14.7 ± 1.8   *PPO (W)*   - *Pre:* NR - *Post:* NR |
|  | Control | *N =* 4 (4 M/ 0 F)    *Age =* 29.8 ± 7.3 years  *TSI =* CD (4 acute/ 0 chronic)  *Classification =* 0 T/ 4 P  *Severity =* 4 comp./ 0 incomp.  *CPET =* Progressive ACE | Conventional rehabilitation program (strengthening, ROM + stretching, training in transfers and ADLs, wheelchair position and standing, respiratory training, bowel + bladder management, psychological support). | *AV̇O_2peak_ (L/min)*   - *Pre:* NR - *Post:* NR   *RV̇O_2peak_ (mL/kg/min)*   - *Pre:* 12.6 ± 3.1 - *Post:* 23.1 ± 3.6   *PPO (W)*   - *Pre:* NR - *Post:* NR |
| Cheung et al. (2019)  China | Intervention | *N =* 8 (7 M/ 1 F)    *Age =* 55.6 ± 4.98 years  *TSI =* 1.42 ± 0.58 years (CD acute/chronic)  *Classification =* CD  *Severity =* 0 comp./ 8 incomp.  *CPET =* Submaximal ACE to predict maximal outcomes  *CPET same modality of intervention? =* No | *Type of Exercise =* Physiotherapy programme plus RABWSTT  *Relative Intensity and Class =* CD  *Session Duration (min) =* 30  *Frequency (times/week) =* 3  *Intervention Length (weeks) =* 8  *Adverse Events =* None | *AV̇O_2peak_ (L/min)*   - *Pre:* NR - *Post:* NR   *RV̇O_2peak_ (mL/kg/min)*   - *Pre:* 25.7 ± 7.16 - *Post:* 26.4 ± 6.99   *PPO (W)*   - *Pre:* NR - *Post:* NR |
|  | Control | *N =* 8 (4 M/ 4 F)    *Age =* 53 ± 12.94 years  *TSI =* 0.87 ± 0.53 years (CD acute/chronic)  *Classification =* CD  *Severity =* 0 comp./ 8 incomp.  *CPET =* Submaximal ACE to predict maximal outcomes | Standard physiotherapy including limb mobilisation and strengthening, trunk stabilisation, wheelchair manoeuvre training and overground walking. | *AV̇O_2peak_ (L/min)*   - *Pre:* NR - *Post:* NR   *RV̇O_2peak_ (mL/kg/min)*   - *Pre:* 20.5 ± 2.93 - *Post:* 20.5 ± 2.84   *PPO (W)*   - *Pre:* NR - *Post:* NR |
| Gorgey et al. (2021)  USA | Intervention | *N =* 20 (18 M/ 2 F)    *Age =* 39 ± 11 years  *TSI =* 13 ± 11 years (0 acute/ 20 chronic)  *Classification =* 6 T/ 14 P  *Severity =* 17 comp./ 3 incomp.  *CPET =* FES-LCE  *CPET same modality of intervention? =* No | *Type of Exercise =* NMES-RT  *Relative Intensity and Class =* Cannot determine (4 sets of 10 reps per leg knee extension)  *Session Duration (min) =* 45-60  *Frequency (times/week) =* 2  *Intervention Length (weeks) =* 12  *Adverse Events =* NR | *AV̇O_2peak_ (L/min)*   - *Pre:* 0.513 ± 0.144 - *Post:* 0.579 ± 0.169   *RV̇O_2peak_ (mL/kg/min)*   - *Pre:* NR - *Post:* NR   *PPO (W)*   - *Pre:* 6.6 ± 3.3 - *Post:* 9 ± 4.4 |
|  | Control | *N =* 13 (10 M/ 3 F)    *Age =* 41.5 ± 13 years  *TSI =* 11 ± 11 years (0 acute/ 13 chronic)  *Classification =* 4 T/ 9 P  *Severity =* 11 comp./ 2 incomp.  *CPET =* FES-LCE | Passive movement training. | *AV̇O_2peak_ (L/min)*   - *Pre:* 0.494 ± 0.218 - *Post:* 0.472 ± 0.178   *RV̇O_2peak_ (mL/kg/min)*   - *Pre:* NR - *Post:* NR   *PPO (W)*   - *Pre:* 16 ± 18 - *Post:* 12 ± 16 |
| Gorman et al. (2016)  USA | Intervention | *N =* 12 (M/F NR)    *Age =* 51.5 ± 12.7 years  *TSI =* NR (0 acute/ 12 chronic)  *Classification =* 8 T/ 4 P  *Severity =* 0 comp./ 12 incomp.  *CPET =* Peak robotic treadmill testing  *CPET same modality of intervention? =* Yes | *Type of Exercise =* RABWSTT  *Relative Intensity and Class =* Vigorous (80-85% HRR)  *Session Duration (min) =* 38.75 (First week 20-min and increased by 5-min each week thereafter)  *Frequency (times/week) =* 3  *Intervention Length (weeks) =* 12  *Adverse Events =* Two subjects developed skin irritation or abrasion in hip, groin, penis, back, wrist, glutei, and scapula. | *AV̇O_2peak_ (L/min)*   - *Pre:* NR - *Post:* NR   *RV̇O_2peak_ (mL/kg/min): RABWSTT*   - *Pre:* 20.2 ± 7.4 - *Post:* 22.7 ± 7.5   *RV̇O_2peak_ (mL/kg/min): ACE*   - *Pre:* 20.0 ± 6.5 - *Post:* 21.7 ± 7.5   *PPO (W)*   - *Pre:* NR - *Post:* NR |
|  | Control | *N =* 6 (M/F NR)    *Age =* 52 ± 15.4 years  *TSI =* NR (0 acute/ 6 chronic)  *Classification =* 6 T/ 0 P  *Severity =* 0 comp./ 6 incomp.  *CPET =* Peak robotic treadmill testing | Home stretching programme. | *AV̇O_2peak_ (L/min)*   - *Pre:* NR - *Post:* NR   *RV̇O_2peak_ (mL/kg/min)*   - *Pre:* 12.9 ± 4.4 - *Post:* 13.4 ± 3.9   *PPO (W)*   - *Pre:* NR - *Post:* NR |
| Hansen et al. (2023)  Denmark | Intervention | *N =* 8 (5 M/ 3 F)    *Age =* 50 ± 4 years  *TSI =* NR (0 acute/ 8 chronic)  *Classification =* NR  *Severity =* 4 comp./ 4 incomp.  *CPET =* Graded arm-crank ergometry exercise test  *CPET same modality of intervention? =* No | *Type of Exercise =* Hybrid, ski ergometry  *Relative Intensity and Class =* Moderate-to-vigorous (RPE 12 – 17).  *Session Duration (min) =* 30  *Frequency (times/week) =* 3  *Intervention Length (weeks) =* 12  *Adverse Events =* NR | *AV̇O_2peak_ (L/min)*   - *Pre:* 1.30 ± 0.47 - *Post:* 1.49 ± 0.55   *RV̇O_2peak_ (mL/kg/min)*   - *Pre:* 15.1 ± 5.1 - *Post:* 17.5 ± 6.1   *PPO (W)*   - *Pre:* 62 ± 29 - *Post:* 79 ± 33 |
|  | Control | *N =* 7 (3 M/ 4 F)    *Age =* 50 ± 13 years  *TSI =* NR (0 acute/ 7 chronic)  *Classification =* NR  *Severity =* 4 comp./ 3 incomp.  *CPET =* Graded arm-crank ergometry exercise test | Asked to maintain normal lifestyle habits throughout the 12 weeks. | *AV̇O_2peak_ (L/min)*   - *Pre:* 1.24 ± 0.34 - *Post:* 1.16 ± 0.28   *RV̇O_2peak_ (mL/kg/min)*   - *Pre:* 16.8 ± 6.1 - *Post:* 16.0 ± 6.0   *PPO (W)*   - *Pre:* 64 ± 21 - *Post:* 62 ± 23 |
| Kim et al. (2015)  South Korea | Intervention | *N =* 8 (6 M/ 2 F)    *Age =* 31.5 ± 5.5 years  *TSI =* 5 ± 3.2 years (0 acute/ 8 chronic)  *Classification =* 6 T/ 2 P  *Severity =* 8 comp./ 0 incomp.  *CPET =* Maximal graded handbike exercise test  *CPET same modality of intervention? =* Yes | *Type of Exercise =* Handbike exercise  *Relative Intensity and Class =* Moderate-to-vigorous (Wk1-2: RPE 5, 70% HR_max_; Wk2-4: RPE 6, 75% HR_max_; Wk4-6: RPE 7, 80% HR_max_)  *Session Duration (min) =* 60  *Frequency (times/week) =* 3  *Intervention Length (weeks) =* 6  *Adverse Events =* NR | *AV̇O_2peak_ (L/min)*   - *Pre:* NR - *Post:* NR   *RV̇O_2peak_ (ml/kg/min)*   - *Pre:* 16.8 ± 7.2 - *Post:* 21.2 ± 9.1   *PPO (W)*   - *Pre:* NR - *Post:* NR |
|  | Control | *N =* 7 (3 M/ 4 F)    *Age =* 35 ± 5.1 years  *TSI =* 8.3 ± 4.5 years (0 acute/ 7 chronic)  *Classification =* 7 T/ 0 P  *Severity =* 7 comp./ 0 incomp.  *CPET =* Maximal graded handbike exercise test | Continue with usual activities. | *AV̇O_2peak_ (L/min)*   - *Pre:* NR - *Post:* NR   *RV̇O_2peak_ (mL/kg/min)*   - *Pre:* 15.2 ± 3.4 - *Post:* 13.7 ± 3.4   *PPO (W)*   - *Pre:* NR - *Post:* NR |
| Kim et al. (2019)  South Korea | Intervention | *N =* 11 (M/F NR)    *Age =* NR  *TSI =* NR (0 acute/ 11 chronic)  *Classification =* NR  *Severity =* NR  *CPET =* Maximal ACE  *CPET same modality of intervention? =* Yes | *Type of Exercise =* Mixed (ACE and resistance exercises)  *Relative Intensity and Class =* Moderate-to-vigorous (6-8 RPE)  *Session Duration (min) =* 60  *Frequency (times/week) =* 3  *Intervention Length (weeks) =* 6  *Adverse Events =* NR | *AV̇O_2peak_ (L/min)*   - *Pre:* NR - *Post:* NR   *RV̇O_2peak_ (mL/kg/min)*   - *Pre:* 11.7 ± 8.1 - *Post:* 15.2 ± 9.6   *PPO (W)*   - *Pre:* NR - *Post:* NR |
|  | Control | *N =* 6 (M/F NR)    *Age =* NR  *TSI =* NR (0 acute/ 6 chronic)  *Classification =* NR  *Severity =* NR  *CPET =* Maximal ACE | Standard care. | *AV̇O_2peak_ (L/min)*   - *Pre:* NR - *Post:* NR   *RV̇O_2peak_ (mL/kg/min)*   - *Pre:* 9.4 ± 4.5 - *Post:* 11.7 ± 7.9   *PPO (W)*   - *Pre:* NR - *Post:* NR |
| Lavado et al. (2013)  Brazil | Intervention | *N =* 21 (18 M/ 3 F)    *Age =* 34.1 ± 11.1 years  *TSI =* 4.8 ± 2.2 years (0 acute/ 21 chronic)  *Classification =* NR  *Severity =* NR  *CPET =* Maximal ACE  *CPET same modality of intervention? =* Yes | *Type of Exercise =* Physiotherapy sessions plus aerobic training (cycloergometer of upper limbs, performed distance with a wheelchair, and general exercises to gain muscle power)  *Relative Intensity and Class =* Moderate-to-vigorous (70-80% HR_max_ or VO_2peak_)  *Session Duration (min) =* 60  *Frequency (times/week) =* 2-3  *Intervention Length (weeks) =* 16  *Adverse Events =* None | *AV̇O_2peak_ (L/min)*   - *Pre:* 0.96 ± 0.37 - *Post:* 1.17 ± 0.26   *RV̇O_2peak_ (mL/kg/min)*   - *Pre:* NR - *Post:* NR   *PPO (W)*   - *Pre:* NR - *Post:* NR |
|  | Control | *N =* 21 (17 M/ 4 F)    *Age =* 38.5 ± 10.6 years  *TSI =* 4.05 ± 1.6 years (0 acute/ 21 chronic)  *Classification =* NR  *Severity =* NR  *CPET =* Maximal ACE | Physiotherapy sessions (passive mobilizations, stretching, and functional activity training). | *AV̇O_2peak_ (L/min)*   - *Pre:* 0.91 ± 0.36 - *Post:* 0.85 ± 0.22   *RV̇O_2peak_ (mL/kg/min)*   - *Pre:* NR - *Post:* NR   *PPO (W)*   - *Pre:* NR - *Post:* NR |
| Nightingale et al. (2018)  UK | Intervention | *N =* 15 (11 M/ 4 F)    *Age =* 46.7 ± 5.6 years  *TSI =* NR (0 acute/ 15 chronic)  *Classification =* 0 T/ 15 P  *Severity =* NR  *CPET =* Discontinuous incremental submaximal ACE  *CPET same modality of intervention? =* Yes | *Type of Exercise =* ACE  *Relative Intensity and Class =* Moderate (60-65% VO_2peak_)  *Session Duration (min) =* 45  *Frequency (times/week) =* 4  *Intervention Length (weeks) =* 6  *Adverse Events =* One participant did not complete the trial due to illness, not related to intervention. | *AV̇O_2peak_ (L/min)*   - *Pre:* 1.49 ± 0.50 - *Post:* 1.69 ± 0.54   *RV̇O_2peak_ (mL/kg/min)*   - *Pre:* 17.78 ± 4.89 - *Post:* 20.73 ± 5.53   *PPO (W)*   - *Pre:* 77 ± 27 - *Post:* 93 ± 32 |
|  | Control | *N =* 8 (2 M/ 6 F)    *Age =* 48 ± 10.3 years  *TSI =* NR (0 acute/ 8 chronic)  *Classification =* 0 T/ 8 P  *Severity =* NR  *CPET =* Discontinuous incremental submaximal ACE | Lifestyle maintenance. | *AV̇O_2peak_ (L/min)*   - *Pre:* 1.55 ± 0.61 - *Post:* 1.49 ± 0.61   *RV̇O_2peak_ (mL/kg/min)*   - *Pre:* 18.81 ± 6.21 - *Post:* 18.32 ± 6.30   *PPO (W)*   - *Pre:* 75 ± 39 - *Post:* 73 ± 36 |
| Pelletier et al. (2015)  Canada | Intervention | *N =* 12 (12 M/ 0 F)    *Age =* 40 ± 12.3 years  *TSI =* 15 ± 8.52 years (0 acute/ 12 chronic)  *Classification =* NR  *Severity =* 4 comp./ 8 incomp.  *CPET =* Symptom-limited graded ACE exercise test  *CPET same modality of intervention? =* Yes | *Type of Exercise =* Mixed: aerobic ACE and RT  *Relative Intensity and Class =* Moderate-to-vigorous (3-6 RPE; 50-70% 1RM RT)  *Session Duration (min) =* 20  *Frequency (times/week) =* 2  *Intervention Length (weeks) =* 16  *Adverse Events =* NR | *AV̇O_2peak_ (L/min)*   - *Pre:* 1.42 ± 0.48 - *Post:* 1.56 ± 0.48   *RV̇O_2peak_ (mL/kg/min)*   - *Pre:* 16.3 ± 5.5 - *Post:* 19.1 ± 6.7   *PPO (W)*   - *Pre:* 67 ± 29 - *Post:* 76 ± 34 |
|  | Control | *N =* 11 (9 M/ 2 F)    *Age =* 45.9 ± 11.5 years  *TSI =* 9.25 ± 10 years (0 acute/ 11 chronic)  *Classification =* NR  *Severity =* 5 comp./ 6 incomp.  *CPET =* Symptom-limited graded ACE exercise test | Members of weekly community exercise program for adult SCI and given no specific guidance or training. | *AV̇O_2peak_ (L/min)*   - *Pre:* 1.30 ± 0.38 - *Post:* 1.23 ± 0.38   *RV̇O_2peak_ (ml/kg/min)*   - *Pre:* 17.8 ± 6.2 - *Post:* 16.2 ± 6.6   *PPO (W)*   - *Pre:* 71 ± 33 - *Post:* 70 ± 35 |
| Piira et al. (2019)  Norway | Intervention | *N =* 10 (6 M/ 4 F)    *Age =* 46 ± 14 years  *TSI =* 11.3 ± 9 years (0 acute/ 10 chronic)  *Classification =* 3 T/ 7 P  *Severity =* 0 comp./ 10 incomp.  *CPET =* Progressive ACE  *CPET same modality of intervention? =* No | *Type of Exercise =* Gait training (body-weight supported locomotor training)  *Relative Intensity and Class =* Cannot determine (“increase towards 3-5 km/h”)  *Session Duration (min) =* 90  *Frequency (times/week) =* 5  *Intervention Length (weeks) =* 12  *Adverse Events =* NR | *AV̇O_2peak_ (L/min)*   - *Pre:* 1.40 ± 0.50 - *Change:* -0.10 ± 0.20   *RV̇O_2peak_ (mL/kg/min)*   - *Pre:* NR - *Post:* NR   *PPO (W)*   - *Pre:* NR - *Post:* NR |
|  | Control | *N =* 10 (9 M/ 1 F)    *Age =* 54 ± 13 years  *TSI =* 7.5 ± 5.8 years (0 acute/ 10 chronic)  *Classification =* 5 T/ 5 P  *Severity =* 0 comp./ 10 incomp.  *CPET =* Progressive ACE | Standard of care from a physical therapist. | *AV̇O_2peak_ (L/min)*   - *Pre:* 1.50 ± 0.40 - *Change:* -0.10 ± 0.20   *RV̇O_2peak_ (mL/kg/min)*   - *Pre:* NR - *Post:* NR   *PPO (W)*   - *Pre:* NR - *Post:* NR |
| Rosety-Rodriguez et al. (2014)  Spain | Intervention | *N =* 9 (9 M/ 0 F)    *Age =* 29.6 ± 3.6 years  *TSI =* 4.6 ± 0.3 years (0 acute/ 9 chronic)  *Classification =* 0 T/ 9 P  *Severity =* 9 comp./ 0 incomp.  *CPET =* Continuous incremental workload test on an ACE  *CPET same modality of intervention? =* Yes | *Type of Exercise =* Upper-body, aerobic ACE  *Relative Intensity and Class =* Moderate-to-vigorous (50-65% HRR)  *Session Duration (min) =* 20-30 (increase of 2.5 min every 3 weeks)  *Frequency (times/week) =* 3  *Intervention Length (weeks) =* 12  *Adverse Events =* NR | *AV̇O_2peak_ (L/min)*   - *Pre:* NR - *Post:* NR   *RV̇O_2peak_ (mL/kg/min)*   - *Pre:* 23.2 ± 2.1 - *Post:* 25.6 ± 1.9   *PPO (W)*   - *Pre:* NR - *Post:* NR |
|  | Control | *N =* 8 (8 M/ 0 F)    *Age =* 30.2 ± 3.8 years  *TSI =* 4.6 ± 0.3 years (0 acute/ 8 chronic)  *Classification =* 0 T/ 8 P  *Severity =* 8 comp./ 0 incomp.  *CPET =* Continuous incremental workload test on an ACE | No participation in the training programme. | *AV̇O_2peak_ (L/min)*   - *Pre:* NR - *Post:* NR   *RV̇O_2peak_ (mL/kg/min)*   - *Pre:* 23.0 ± 2.2 - *Post:* 22.6 ± 1.9   *PPO (W)*   - *Pre:* NR - *Post:* NR |
| Taylor et al. (1986)  Canada | Intervention | *N =* 5 (5 M/ 0 F)    *Age =* 27 ± 3 years  *TSI =* 7.2 ± 4.4 years (0 acute/ 5 chronic)  *Classification =* 0 T/ 5 P  *Severity =* NR  *CPET =* Maximal ACE exercise test  *CPET same modality of intervention? =* Yes | *Type of Exercise =* Upper-body aerobic ACE  *Relative Intensity and Class =* Vigorous (80% HR_peak_)  *Session Duration (min) =* 30  *Frequency (times/week) =* 5  *Intervention Length (weeks) =* 8  *Adverse Events =* NR | *AV̇O_2peak_ (L/min)*   - *Pre:* 1.9 ± 0.6 - *Post:* 2.1 ± 0.4   *RV̇O_2peak_ (mL/kg/min)*   - *Pre:* 22.8 ± 4.6 - *Post:* 26.3 ± 3.6   *PPO (W)*   - *Pre:* NR - *Post:* NR |
|  | Control | *N =* 5 (5 M/ 0 F)    *Age =* 33 ± 8 years  *TSI =* 15.8 ± 13.9 years (0 acute/ 5 chronic)  *Classification =* 0 T/ 5 P  *Severity =* NR  *CPET =* Maximal ACE exercise test | No formalised exercise routine. | *AV̇O_2peak_ (L/min)*   - *Pre:* 1.70 ± 0.50 - *Post:* 1.60 ± 0.50   *RV̇O_2peak_ (mL/kg/min)*   - *Pre:* 22.6 ± 8.5 - *Post:* 23.4 ± 8.9   *PPO (W)*   - *Pre:* NR - *Post:* NR |
| Torhaug et al. (2016)  Norway | Intervention | *N =* 9 (9 M/ 0 F)    *Age =* 42 ± 12.4 years  *TSI =* 13.2 ± 7.2 years (0 acute/ 9 chronic)  *Classification =* 0 T/ 9 P  *Severity =* 6 comp./ 3 incomp.  *CPET =* Maximal wheelchair ergometry test  *CPET same modality of intervention? =* No | *Type of Exercise =* RT (bench press)  *Relative Intensity and Class =* Vigorous (85-95% 1RM)  *Session Duration (min) =* 60  *Frequency (times/week) =* 3  *Intervention Length (weeks) =* 6  *Adverse Events =* NR | *AV̇O_2peak_ (L/min)*   - *Pre:* 2.15 ± 0.47 - *Post:* 2.06 ± 0.61   *RV̇O_2peak_ (mL/kg/min)*   - *Pre:* 28.5 ± 6.5 - *Post:* 28.5 ± 7.0   *PPO (W)*   - *Pre:* NR - *Post:* NR |
|  | Control | *N =* 7 (7 M/ 0 F)    *Age =* 47.1 ± 5.8 years  *TSI =* 15.1 ± 10.2 years (0 acute/ 7 chronic)  *Classification =* 0 T/ 7 P  *Severity =* 5 comp./ 2 incomp.  *CPET =* Maximal wheelchair ergometry test | No formalised exercise routine. | *AV̇O_2peak_ (L/min)*   - *Pre:* 2.00 ± 0.51 - *Post:* 2.03 ± 0.49   *RV̇O_2peak_ (mL/kg/min)*   - *Pre:* 24.3 ± 5.7 - *Post:* 24.9 ± 4.5   *PPO (W)*   - *Pre:* NR - *Post:* NR |
| Van der Scheer et al. (2016)  The Netherlands | Intervention | *N =* 14 (12 M/ 2 F)    *Age =* 53.6 ± 16.3 years  *TSI =* 19.3 ± 11.9 years (0 acute/ 14 chronic)  *Classification =* 5 T/ 9 P  *Severity =* 10 comp./ 4 incomp.  *CPET =* Peak incremental treadmill test  *CPET same modality of intervention? =* Yes | *Type of Exercise =* Wheelchair propulsion*  *Relative Intensity and Class =* Light (1-3 RPE; 30-40% HRR)  *Session Duration (min) =* 30  *Frequency (times/week) =* 2  *Intervention Length (weeks) =* 16  *Adverse Events =* NR  **Exercise was continuous in participants with the highest baseline fitness levels, while those with lower levels followed protocols with intermittent exercise (4 × 7.5 or 10 × 3 min with 1–2 min rest intervals).* | *AV̇O_2peak_ (L/min)*   - *Pre:* 1.11 ± 0.60 - *Post:* 1.06 ± 0.58   *RV̇O_2peak_ (mL/kg/min)*   - *Pre:* NR - *Post:* NR   *PPO (W)*   - *Pre:* 35 ± 25 - *Post:* 36 ± 27 |
|  | Control | *N =* 15 (10 M/ 5 F)    *Age =* 55 ± 11.9 years  *TSI =* 21.7 ± 12.6 years (0 acute/ 15 chronic)  *Classification =* 4 T/ 11 P  *Severity =* 10 comp./ 5 incomp.  *CPET =* Peak incremental treadmill test | No intervention. | *AV̇O_2peak_ (L/min)*   - *Pre:* 1.13 ± 0.42 - *Post:* 1.10 ± 0.38   *RV̇O_2peak_ (mL/kg/min)*   - *Pre:* NR - *Post:* NR   *PPO (W)*   - *Pre:* 41 ± 24 - *Post:* 40 ± 23 |
| Yarar-Fisher et al. (2018)  USA | Intervention | *N =* 6 (6 M/ 0 F)    *Age =* 48.5 ± 6.6 years  *TSI =* 23.8 ± 7.5 years (0 acute/ 6 chronic)  *Classification =* 2 T/ 4 P  *Severity =* 6 comp./ 0 incomp.  *CPET =* Progressive ACE test  *CPET same modality of intervention? =* Yes | *Type of Exercise =* Mixed (NMES, RT and ACE)  *Relative Intensity and Class =* Vigorous (80-90% VO_2peak_)  *Session Duration (min) =* 15  *Frequency (times/week) =* 3  *Intervention Length (weeks) =* 8  *Adverse Events =* NR | *AV̇O_2peak_ (L/min)*   - *Pre:* NR - *Post:* NR   *RV̇O_2peak_ (mL/kg/min)*   - *Pre:* 13.2 ± 3.4 - *Post:* 14.6 ± 3.3   *PPO (W)*   - *Pre:* NR - *Post:* NR |
|  | Control | *N =* 5 (4 M/ 1 F)    *Age =* 43 ± 5.4 years  *TSI =* 19.4 ± 4 years (0 acute/ 5 chronic)  *Classification =* 2 T/ 3 P  *Severity =* 5 comp./ 0 incomp.  *CPET =* Progressive ACE test | High protein diet only. | *AV̇O_2peak_ (L/min)*   - *Pre:* NR - *Post:* NR   *RV̇O_2peak_ (mL/kg/min)*   - *Pre:* 17.7 ± 3.2 - *Post:* 17.0 ± 3.5   *PPO (W)*   - *Pre:* NR - *Post:* NR |
| **RANDOMISED-CONTROLLED TRIALS (BEHAVIOUR CHANGE)** | | | | |
| Bombardier et al. (2021)  USA | Intervention | *N =* 7 (5 M/ 2 F)    *Age =* 56 ± 13 years  *TSI =* 12 ± 8 years (0 acute/ 7 chronic)  *Classification =* 0 T/ 7 P  *Severity =* NR  *CPET =* Graded maximal ACE exercise test  *CPET same modality of intervention? =* No | *Type of Behaviour Change Counselling =* Tele-health intervention (target of 150 min of MVPA per week) plus DVD*  *Number of Contact Sessions =* 16 telephone calls  *Intervention Length (weeks) =* 24  *Adverse Events =* Arm of the DXA scanner struck a participant's knee as it was returning to the start position but the physician detected no injury.  **The DVD provided verbal instructions and videos in which people with SCI demonstrated stretching, aerobic exercise, and strength training routines designed specifically for people with paraplegia or tetraplegia.* | *AV̇O_2peak_ (L/min)*   - *Pre:* NR - *Post:* NR   *RV̇O_2peak_ (mL/kg/min)*   - *Pre:* 13.4 ± 3.0 - *Post:* 13.8 ± 3.4   *PPO (W)*   - *Pre:* 69 ± 20 - *Post:* 73 ± 10 |
|  | Control | *N =* 8 (6 M/ 2 F)    *Age =* 49 ± 13 years  *TSI =* 18 ± 16 years (0 acute/ 8 chronic)  *Classification =* 2 T/ 6 P  *Severity =* NR  *CPET =* Graded maximal ACE exercise test | Advised to seek medical care for advice on lifestyle changes such as diet and exercise. | *AV̇O_2peak_ (L/min)*   - *Pre:* NR - *Post:* NR   *RV̇O_2peak_ (mL/kg/min)*   - *Pre:* 13.9 ± 3.9 - *Post:* 12.7 ± 3.3   *PPO (W)*   - *Pre:* 74 ± 25 - *Post:* 67 ± 21 |
| Froehlich-Grobe et al. (2022)  USA | Intervention | *N =* 87 (51 M/ 36 F)    *Age =* 48.6 ± 11.5 years  *TSI =* 14.8 ± 12.2 years (NR acute/ NR chronic)  *Classification =* NR  *Severity =* NR  *CPET =* Graded maximal ACE exercise test  *CPET same modality of intervention? =* No | *Type of Behaviour Change Counselling =* Mixture of support mechanisms including website content (exercise and disability information plus 16 weekly modules on self-management topics), weekly 60-minute group-based virtual meetings, Facebook group for participants, exercise equipment starter package (TheraBands, seated aerobics DVD, pedal table-top arm ergometer).  *Number of Contact Sessions =* At least 16  *Intervention Length (weeks) =* 16  *Adverse Events =* NR | *AV̇O_2peak_ (L/min)*   - *Pre:* NR - *Post:* NR   *RV̇O_2peak_ (mL/kg/min) *N=20*   - *Pre:* 10.7 ± 3.3 - *Post:* 11.7 ± 3.3   *PPO (W) *N=20*   - *Pre:* 39 ± 18 - *Post:* 41 ± 20 |
|  | Control | *N =* 81 (45 M/ 36 F)    *Age =* 50.6 ± 13.1 years  *TSI =* 16.2 ± 12.4 years (NR acute/ NR chronic)  *Classification =* NR  *Severity =* NR  *CPET =* Graded maximal ACE exercise test | Waitlist control. | *AV̇O_2peak_ (L/min)*   - *Pre:* NR - *Post:* NR   *RV̇O_2peak_ (mL/kg/min) *N=16*   - *Pre:* 11.7 ± 3.7 - *Post:* 11.9 ± 4.0   *PPO (W) *N=16*   - *Pre:* 41 ± 24 - *Post:* 43 ± 24 |
| Kooijmans et al. (2017)  The Netherlands | Intervention | *N =* 33 (21 M/ 12 F)    *Age =* 48 ± 10 years  *TSI =* 21 ± 8 years (0 acute/ 33 chronic)  *Classification =* 11 T/ 22 P  *Severity =* 24 comp./ 9 incomp.  *CPET =* Maximal wheelchair treadmill test  *CPET same modality of intervention? =* No | *Type of Behaviour Change Counselling =* Self-management intervention (group meetings, book and individual counselling)  *Number of Contact Sessions =* 12 (2 home visits + 5 individual sessions + 5 group sessions +)  *Intervention Length (weeks) =* 42  *Adverse Events =* NR | *AV̇O_2peak_ (L/min) *N=15*   - *Pre:* 3.50 ± 5.70 - *Post:* 3.00 ± 5.50   *RV̇O_2peak_ (mL/kg/min)*   - *Pre:* NR - *Post:* NR   *PPO (W) *N=15*   - *Pre:* 49 ± 23 - *Post:* 58 ± 26 |
|  | Control | *N =* 31 (24 M/ 7 F)    *Age =* 49 ± 11 years  *TSI =* 23 ± 10 years (0 acute/ 31 chronic)  *Classification =* 10 T/ 21 P  *Severity =* 26 comp./ 5 incomp.  *CPET =* Maximal wheelchair treadmill test | Only received information about active lifestyle by one group meeting and via a book. | *AV̇O_2peak_ (L/min) *N=15*   - *Pre:* 3.20 ± 5.30 - *Post:* 2.10 ± 2.70   *RV̇O_2peak_ (mL/kg/min)*   - *Pre:* NR - *Post:* NR   *PPO (W) *N=15*   - *Pre:* 42 ± 20 - *Post:* 48 ± 26 |
| Ma et al. (2019)  Canada | Intervention | *N =* 14 (9 M/ 5 F)    *Age =* 45.79 ± 13.63 years  *TSI =* 14.71 ± 13.63 years (0 acute/ 14 chronic)  *Classification =* 5 T/ 9 P  *Severity =* 8 comp./ 6 incomp.  *CPET =* Graded ACE exercise test  *CPET same modality of intervention? =* No | *Type of Behaviour Change Counselling =* Behavioural physical activity coaching  *Number of Contact Sessions =* 9 (1-h introductory session followed by eight once-weekly 10- to 15-min behavioural physical activity coaching sessions)  *Intervention Length (weeks) =* 8  *Adverse Events =* NR | *AV̇O_2peak_ (L/min)*   - *Pre:* 1.16 ± 0.38 - *Post:* 1.30 ± 0.43   *RV̇O_2peak_ (mL/kg/min)*   - *Pre:* 15.9 ± 4.2 - *Post:* 17.8 ± 4.9   *PPO (W)*   - *Pre:* 82 ± 27 - *Post:* 87 ± 30 |
|  | Control | *N =* 14 (8 M/ 6 F)    *Age =* 45.57 ± 10.49 years  *TSI =* 18.14 ± 10.85 years (0 acute/ 14 chronic)  *Classification =* 8 T/ 6 P  *Severity =* 7 comp./ 7 incomp.  *CPET =* Graded ACE exercise test | Waitlist controls. | *AV̇O_2peak_ (L/min)*   - *Pre:* 1.13 ± 0.46 - *Post:* 1.06 ± 0.40   *RV̇O_2peak_ (mL/kg/min)*   - *Pre:* 15.0 ± 5.7 - *Post:* 13.9 ± 5.5   *PPO (W)*   - *Pre:* 70 ± 37 - *Post:* 65 ± 36 |
| Nooijen et al. (2017)  The Netherlands | Intervention | *N =* 20 (17 M/ 3 F)    *Age =* 44 ± 15 years  *TSI =* 0.38 ± 0.18 years (20 acute/ 0 chronic)  *Classification =* 7 T/ 13 P  *Severity =* 13 comp./7 incomp.  *CPET =* Graded maximal handcycle test  *CPET same modality of intervention? =* No | *Type of Behaviour Change Counselling =* Face-to-face sessions  *Number of Contact Sessions =* 13  *Intervention Length (weeks) =* 52  *Adverse Events =* NR | *AV̇O_2peak_ (L/min)*   - *Pre:* 1.20 ± 0.43 - *Post:* 1.52 ± 0.60   *RV̇O_2peak_ (mL/kg/min)*   - *Pre:* NR - *Post:* NR   *PPO (W)*   - *Pre:* 44 ± 23 - *Post:* 80 ± 49 |
|  | Control | *N =* 19 (16 M/ 3 F)    *Age =* 44 ± 15 years  *TSI =* 0.44 ± 0.22 years (19 acute/ 0 chronic)  *Classification =* 6 T/ 13 P  *Severity =* 11 comp./ 8 incomp.  *CPET =* Graded maximal handcycle test | Regular rehabilitation without behaviour intervention. | *AV̇O_2peak_ (L/min)*   - *Pre:* 1.19 ± 0.40 - *Post:* 1.48 ± 0.62   *RV̇O_2peak_ (mL/kg/min)*   - *Pre:* NR - *Post:* NR   *PPO (W)*   - *Pre:* 47 ± 25 - *Post:* 73 ± 48 |
| Data is presented as mean ± standard deviation, unless otherwise stated. 1RM, one repetition maximum; ACE, arm crank ergometry; AV̇O2peak, absolute peak oxygen consumption; BWSTT, body weight supported treadmill training; CPET, cardiopulmonary exercise test; F, females; FES, functional electrical stimulation; HIIT, high-intensity interval training; HRmax, maximum heart rate; HRpeak, peak heart rate; HRR, heart rate reserve; M, male; NMES, neuromuscular electrical stimulation; NR, not reported; P, paraplegia; PPO, peak power output; RABWSTT, robotic-assisted body weight supported treadmill training; RCT, randomised-controlled trial; RPE, rating of perceived exertion; RT, resistance training; RV̇O2peak, relative peak oxygen consumption; T, tetraplegia; TSI, time since injury; V̇O2peak, peak oxygen consumption; W, watts. | | | | |

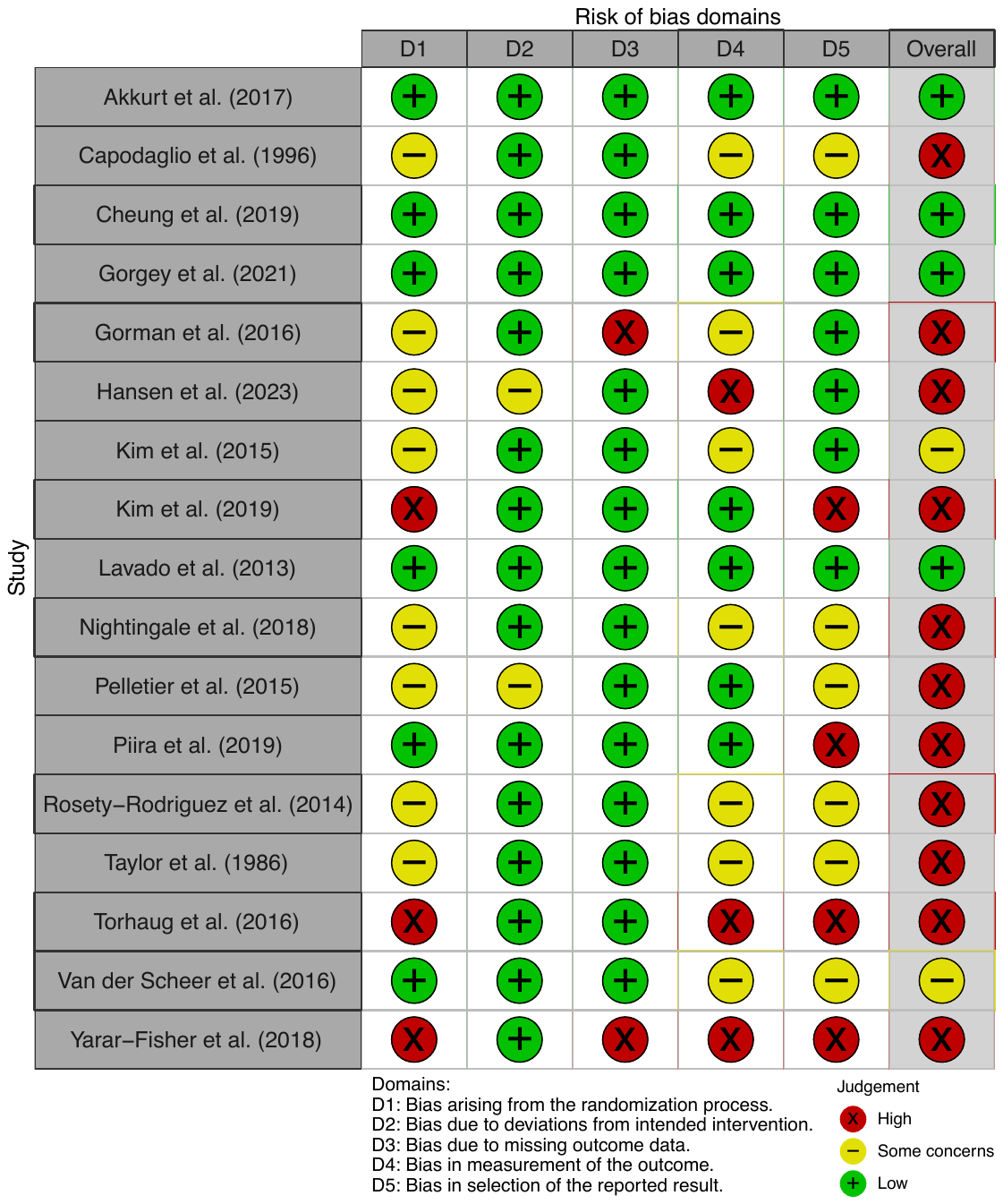

**Figure 1:** Traffic plot for risk of bias in the individual RCTs (exercise intervention vs. true-world control or standard of care), assessed via the Cochrane RoB 2 tool.

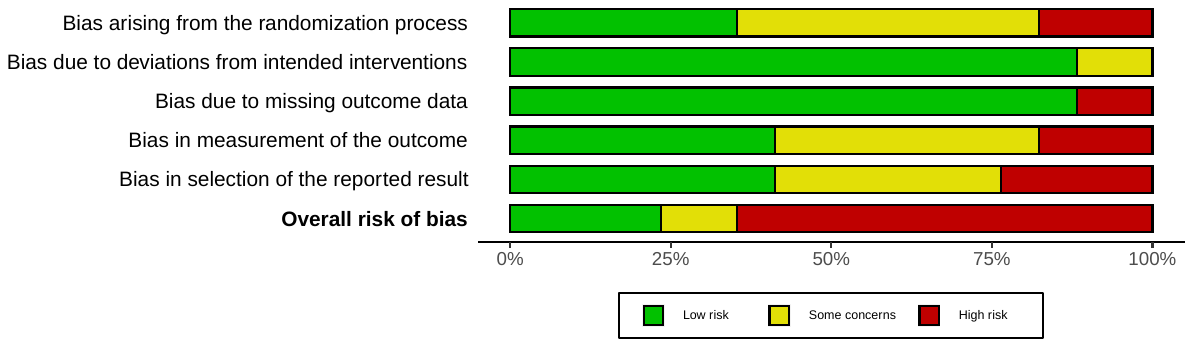

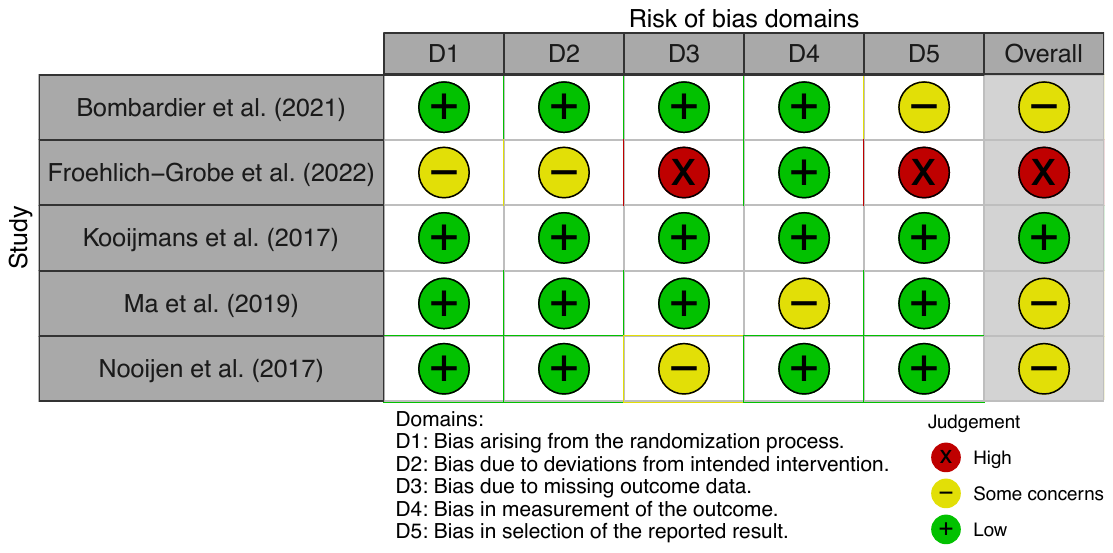
**Figure 2:** Summary plot for risk of bias in the individual RCTs (exercise intervention vs. true-world control or standard of care), assessed via the Cochrane RoB 2 tool.

**Figure 3:** Traffic light plot for risk of bias in the behaviour change RCTs, assessed via the Cochrane RoB 2 tool.

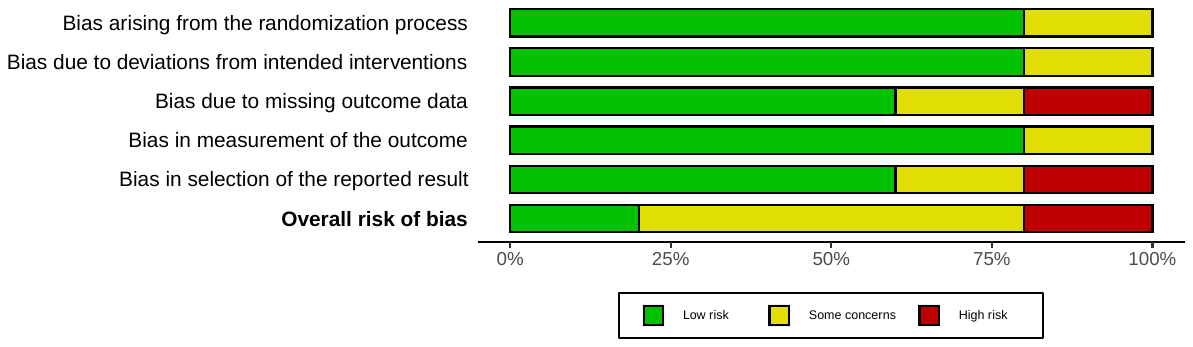
**Figure 4:** Summary plot for risk of bias in the behaviour change RCTs, assessed via the Cochrane RoB 2 tool.

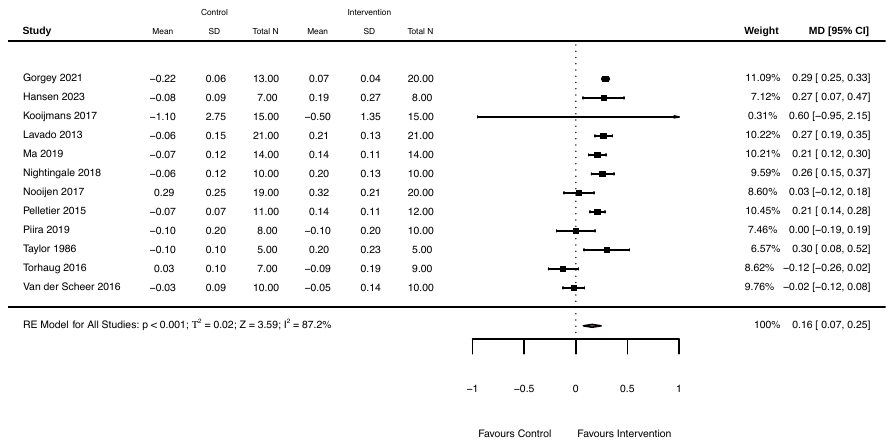

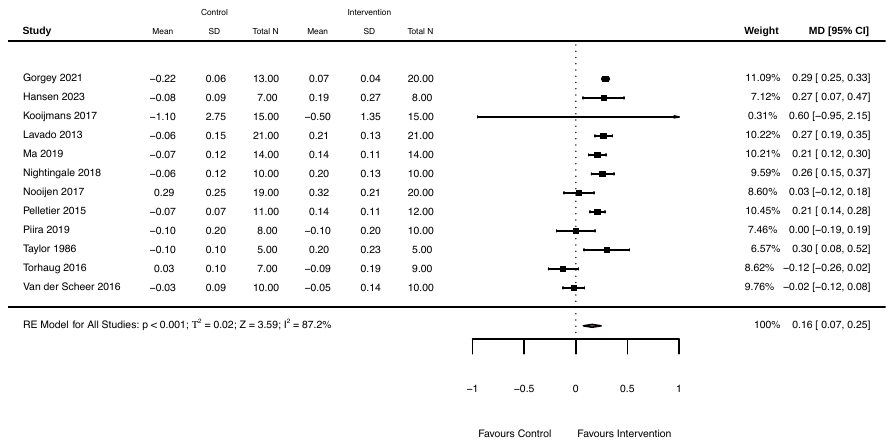

**Figure 5:** Changes in absolute peak oxygen consumption (L/min) following control and exercise interventions in randomised-controlled trial studies. MD, mean difference; N, number of participants; RE, random effects; SD, standard deviation.

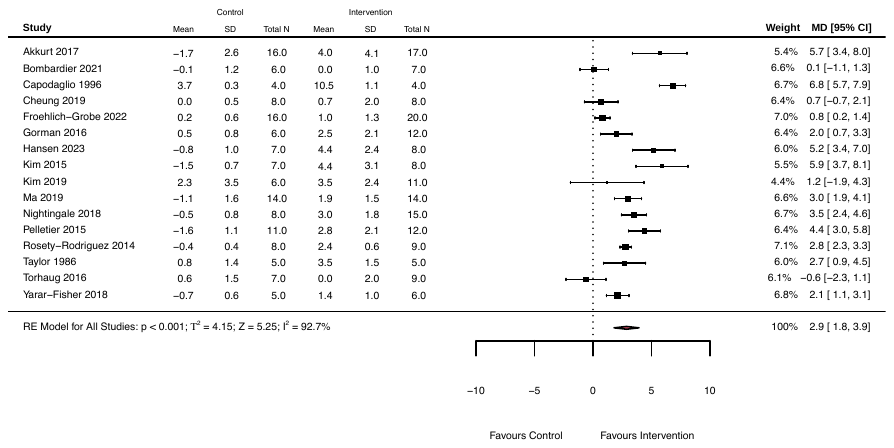

**Figure 6:** Changes in relative peak oxygen consumption (mL/kg/min) following control and exercise interventions in randomised-controlled trial studies. MD, mean difference; N, number of participants; RE, random effects; SD, standard deviation.

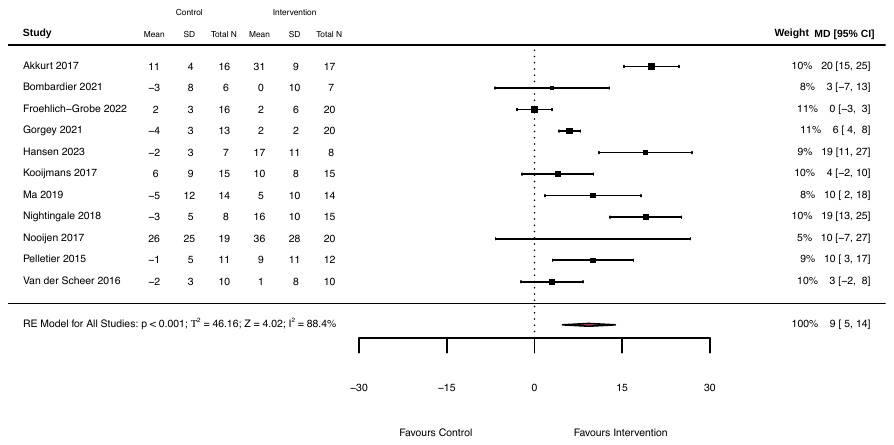
**Figure 7:** Changes in peak power output (W) following control and exercise interventions in randomised-controlled trial studies. MD, mean difference; N, number of participants; RE, random effects; SD, standard deviation.

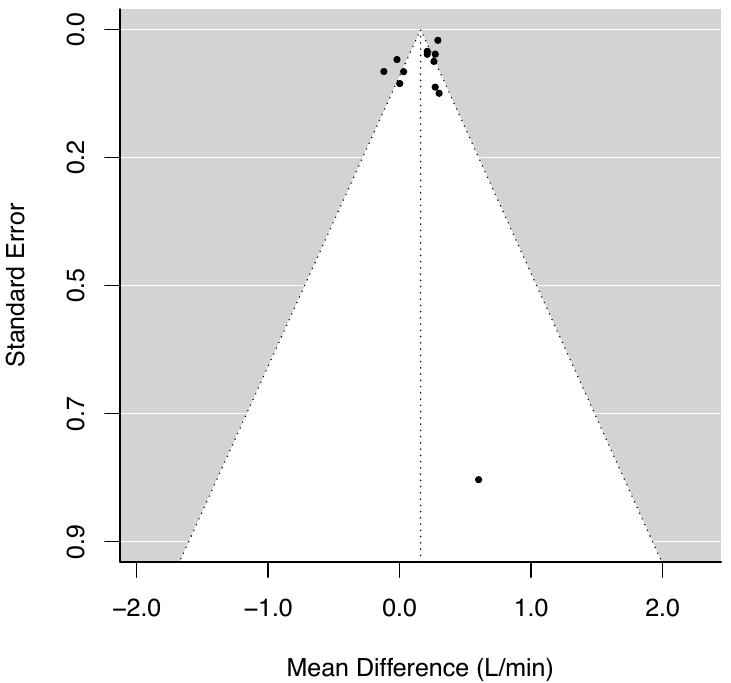

**Figure 8:** Funnel plot of absolute peak oxygen consumption in randomised-controlled trial studies. Egger’s test for funnel plot asymmetry was not significant (Z = 0.02, *p* = 0.99).

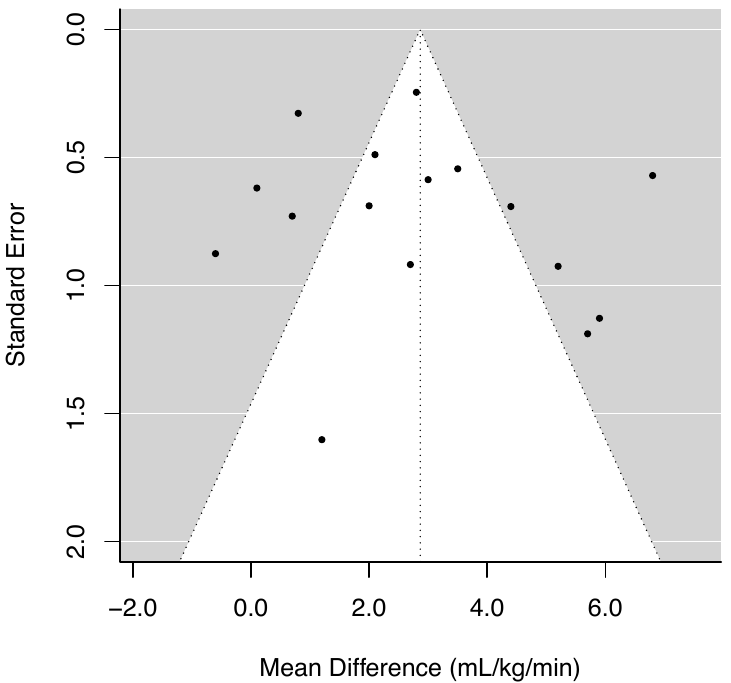

**Figure 9:** Funnel plot of relative peak oxygen consumption in randomised-controlled trial studies. Egger’s test for funnel plot asymmetry was not significant (Z = 0.62, *p* = 0.54).

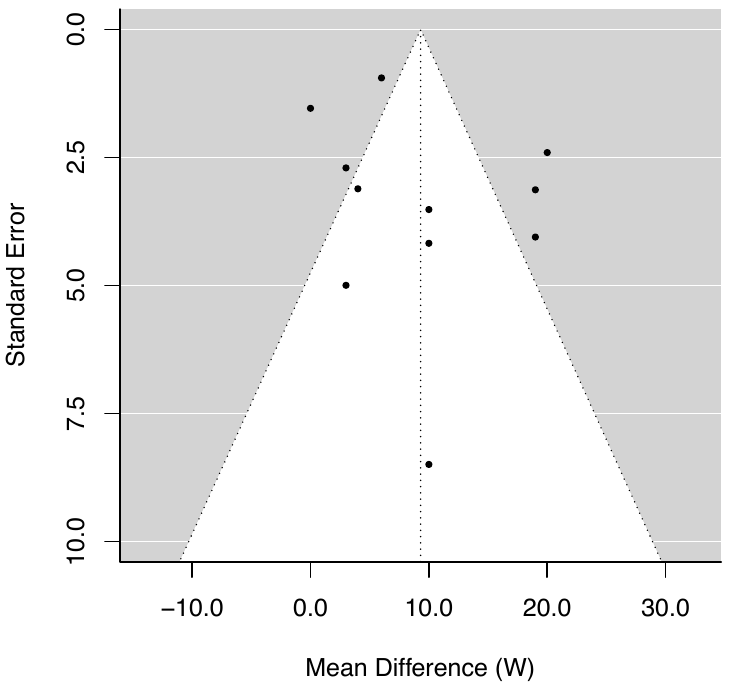

**Figure 10:** Funnel plot of peak power output in randomised-controlled trial studies. Egger’s test for funnel plot asymmetry was not significant (Z = 0.54, *p* = 0.59).

**References**

RCT (exercise intervention vs. true-world control or standard of care) studies included in the systematic review, sorted alphabetically:

Akkurt H, Karapolat HU, Kirazli Y, Kose T. The effects of upper extremity aerobic exercise in patients with spinal cord injury: a randomized controlled study. Eur J Phys Rehabil Med. 2017;53: 219–227.

Bombardier CH, Dyer JR, Burns P, Crane DA, Takahashi MM, Barber J, et al. A tele-health intervention to increase physical fitness in people with spinal cord injury and cardiometabolic disease or risk factors: a pilot randomized controlled trial. Spinal Cord. 2021;59: 63–73.

Capodaglio P, Grilli C, Bazzini G. Tolerable exercise intensity in the early rehabilitation of paraplegic patients. A preliminary study. Spinal Cord. 1996;34: 684–690.

Cheung EYY, Yu KKK, Kwan RLC, Ng CKM, Chau RMW, Cheing GLY. Effect of EMG-biofeedback robotic-assisted body weight supported treadmill training on walking ability and cardiopulmonary function on people with subacute spinal cord injuries - a randomized controlled trial. BMC Neurol. 2019;19: 140.

Froehlich-Grobe K, Lee J, Ochoa C, Lopez A, Sarker E, Driver S et al. Effectiveness and feasibility of the workout on wheels internet intervention (WOWii) for individuals with spinal cord injury: a randomized controlled trial. Spinal Cord. 2022 Oct;60(10):862-874.

Gorgey AS, Lai RE, Khalil RE, Rivers J, Cardozo C, Chen Q, et al. Neuromuscular electrical stimulation resistance training enhances oxygen uptake and ventilatory efficiency independent of mitochondrial complexes after spinal cord injury: a randomized clinical trial. J Appl Physiol. 2021;131: 265–276.

Gorman PH, Scott W, York H, Theyagaraj M, Price-Miller N, McQuaid J, et al. Robotically assisted treadmill exercise training for improving peak fitness in chronic motor incomplete spinal cord injury: A randomized controlled trial. J Spinal Cord Med. 2016;39: 32–44.

Hansen RK, Samani A, Laessoe U, Handberg A, Mellergaard M, Figlewski K et al. Rowing exercise increases cardiorespiratory fitness and brachial artery diameter but not traditional cardiometabolic risk factors in spinal cord-injured humans. Eur J Appl Physiol. 2023; 1-15. Epub ahead of print.

Kim D-I, Lee H, Lee B-S, Kim J, Jeon JY. Effects of a 6-Week Indoor Hand-Bike Exercise Program on Health and Fitness Levels in People With Spinal Cord Injury: A Randomized Controlled Trial Study. Arch Phys Med Rehabil. 2015;96: 2033–40.e1.

Kim D-I, Taylor JA, Tan CO, Park H, Kim JY, Park S-Y, et al. A pilot randomized controlled trial of 6-week combined exercise program on fasting insulin and fitness levels in individuals with spinal cord injury. Eur Spine J. 2019;28: 1082–1091.

Kooijmans H, Post MWM, Stam HJ, van der Woude LHV, Spijkerman DCM, Snoek GJ, et al. Effectiveness of a Self-Management Intervention to Promote an Active Lifestyle in Persons With Long-Term Spinal Cord Injury: The HABITS Randomized Clinical Trial. Neurorehabil Neural Repair. 2017;31: 991–1004.

Lavado EL, Cardoso JR, Silva LGA, Dela Bela LF, Atallah AN. Effectiveness of aerobic physical training for treatment of chronic asymptomatic bacteriuria in subjects with spinal cord injury: a randomized controlled trial. Clin Rehabil. 2013;27: 142–149.

Ma JK, West CR, Martin Ginis KA. The Effects of a Patient and Provider Co-Developed, Behavioral Physical Activity Intervention on Physical Activity, Psychosocial Predictors, and Fitness in Individuals with Spinal Cord Injury: A Randomized Controlled Trial. Sports Med. 2019;49: 1117–1131.

Nightingale TE, Rouse PC, Walhin J-P, Thompson D, Bilzon JLJ. Home-Based Exercise Enhances Health-Related Quality of Life in Persons With Spinal Cord Injury: A Randomized Controlled Trial. Arch Phys Med Rehabil. 2018;99: 1998–2006.e1.

Nooijen CF, Stam HJ, Sluis T, Valent L, Twisk J, van den Berg-Emons RJ. A behavioral intervention promoting physical activity in people with subacute spinal cord injury: secondary effects on health, social participation and quality of life. Clin Rehabil. 2017;31: 772–780.

Pelletier CA, Totosy de Zepetnek JO, MacDonald MJ, Hicks AL. A 16-week randomized controlled trial evaluating the physical activity guidelines for adults with spinal cord injury. Spinal Cord. 2015;53: 363–367.

Piira A, Lannem AM, Sørensen M, Glott T, Knutsen R, Jørgensen L, et al. Manually assisted body-weight supported locomotor training does not re-establish walking in non-walking subjects with chronic incomplete spinal cord injury: A randomized clinical trial. J Rehabil Med. 2019;51: 113–119.

Rosety-Rodriguez M, Camacho A, Rosety I, Fornieles G, Rosety MA, Diaz AJ, et al. Low-grade systemic inflammation and leptin levels were improved by arm cranking exercise in adults with chronic spinal cord injury. Arch Phys Med Rehabil. 2014;95: 297–302.

Taylor AW, McDonell E, Brassard L. The effects of an arm ergometer training programme on wheelchair subjects. Paraplegia. 1986;24: 105–114.

Tørhaug T, Brurok B, Hoff J, Helgerud J, Leivseth G. The effect from maximal bench press strength training on work economy during wheelchair propulsion in men with spinal cord injury. Spinal Cord. 2016;54: 838–842.

van der Scheer JW, de Groot S, Tepper M, Faber W, ALLRISC group, Veeger DH, et al. Low-intensity wheelchair training in inactive people with long-term spinal cord injury: A randomized controlled trial on fitness, wheelchair skill performance and physical activity levels. J Rehabil Med. 2016;48: 33–42.

Yarar-Fisher C, Polston KFL, Eraslan M, Henley KY, Kinikli GI, Bickel CS, et al. Paralytic and nonparalytic muscle adaptations to exercise training versus high-protein diet in individuals with long-standing spinal cord injury. J Appl Physiol. 2018;125: 64–72.
