## Supplementary material for "The effect of exercise on aerobic capacity in individuals with spinal cord injury: A systematic review with meta-analysis and meta-regression": S6 Secondary pooled meta-analysis (pre-post and RCT exercise interventions) (

**Electronic Supplementary Material 6**

**PLoS Medicine**

Hodgkiss, D.D^1^, Bhangu, G^2,3^, Lunny, C^4^, Jutzeler C.R^5,6^, Chiou S.Y^1,7,8,9^ Walter, M^2,10^, Lucas S.E^1,7^, Krassioukov, A.V.^2,11,12^, Nightingale, T.E.^1,2,9^

**^1^** School of Sport, Exercise and Rehabilitation Sciences, University of Birmingham, UK.

**^2^** International Collaboration on Repair Discoveries (ICORD), University of British Columbia, Vancouver, British Columbia, Canada. **^3^** MD Undergraduate Program, Faculty of Medicine, University of British Columbia, Vancouver, Canada. **^4^** Knowledge Translation Program, Li Ka Shing Knowledge Institute, St. Michael’s Hospital, Toronto, and the University of British Columbia, Vancouver, British Columbia, Canada.

**^5^** Department of Health Sciences and Technology, ETH Zurich, Zurich, Switzerland. **^6^** Schulthess Clinic, Zurich, Switzerland. **^7^** Centre for Human Brain Health, University of Birmingham, United Kingdom. **^8^** MRC Versus Arthritis Centre for Musculoskeletal Ageing Research, University of Birmingham, United Kingdom.

**^9^** Centre for Trauma Science Research, University of Birmingham, United Kingdom. **^10^** Department of Urology, University Hospital Basel, University of Basel, Basel, Switzerland. **^11^** Department of Medicine, Division of Physical Medicine and Rehabilitation, University of British Columbia, Vancouver, British Columbia, Canada. **^12^** GF Strong Rehabilitation Centre, Vancouver Coastal Health, Vancouver, British Columbia, Canada.

***Purpose:*** This supplementary file contains the study information, participant/injury characteristics, exercise intervention parameters, and cardiorespiratory fitness (CRF) outcome data for each exercise intervention included in the secondary, pooled meta-analysis. Risk of bias assessments are presented for both pre-post and randomised-controlled trial (RCT) studies. Forest and funnel plots are presented for each subgroup analysis to identify whether certain injury characteristics or exercise intervention parameters mediate CRF responses.

***Conclusion:*** Exercise interventions >2 weeks significantly improve absolute (AV̇O_2peak_) and relative peak oxygen consumption (RV̇O_2peak_), and peak power output (PPO). Subgroup analyses reveal no influence of injury characteristics or exercise intervention parameters on AV̇O_2peak_, yet there is evidence for greater changes in RV̇O_2peak_ with exercise interventions 12 weeks or less, relative to interventions longer than 12 weeks. Upper-body aerobic exercise or resistance training resulted in greater changes in RV̇O_2peak,_ relative to other exercise modalities. Furthermore, there is evidence for greater changes in PPO in individuals with an acute injury or with paraplegia. Aerobic, upper-body exercise or resistance training, with intensity prescribed via oxygen consumption or heart rate, may also result in greater improvements in PPO. The overall methodological quality of pre-post studies was either good or fair, whereas the majority of RCTs had a high risk of bias. Ultimately, more adequately powered RCTs are required to investigate the dose-response relationship between exercise and CRF in individuals with SCI.

| **Table 1.** Participant demographics and injury characteristics reported within the included studies of the secondary, pooled meta-analysis at baseline. | | | |
| --- | --- | --- | --- |
|  | **AV̇O_2peak_**  **(L/min)** | **RV̇O_2peak_ (mL/kg/min)** | **PPO**  **(W)** |
| **Baseline CRF** | | | |
| Total number of interventions [sum of participants]  Mean (range) | 74 [849]  1.28 (0.51 – 3.50) | 79 [897]  17.8 (7.3 – 36.9) | 65 [774]  49 (0 – 168) |
| **Participant demographics** | | | |
| *Age (years)* | 38 (23 - 59) | 40 (24 - 58) | 40 (23 - 57) |
| *Sex* | | | |
| Male | 23 [111] | 23 [170] | 21 [167] |
| Female | - | - | - |
| Mixed (% F) | 47 [632] (21%) | 47 [644] (25%) | 42 [594] (25%) |
| Not reported/cannot determine | 4 [26] | 9 [83] | 2 [13] |
| **Injury characteristics** | | | |
| *Time since injury (years)* | 8 (0 - 21) | 7 (0 - 24) | 8 (0 - 21) |
| Acute (<1-year) | 8 [121] | 8 [95] | 10 [127] |
| Chronic (>1-year) | 49 [487] | 52 [478] | 40 [382] |
| Mixed (% acute) | 7 [89] (13.5%) | 6 [64] (17%) | 7 [84] (24%) |
| Not reported/cannot determine | 10 [152] | 13 [260] | 8 [181] |
| *Neurological level of injury (TETRA/PARA)* | | | |
| TETRA | 2 [18] | 3 [23] | 3 [23] |
| PARA | 22 [204] | 29 [288] | 24 [237] |
| Mixed (% PARA) | 42 [535] (58%) | 36 [406] (51%) | 32 [382] (62%) |
| Not reported/cannot determine | 8 [92] | 8 [180] | 6 [132] |
| *Severity* | |  |  |
| Motor-complete (AIS A-B) | 27 [248] | 31 [273] | 24 [219] |
| Motor-incomplete (AIS C-D) | 8 [102] | 13 [142] | 2 [14] |
| Mixed (% motor-incomplete) | 25 [363] (31%) | 23 [310] (32%) | 27 [359] (36%) |
| Not reported/cannot determine | 14 [136] | 12 [172] | 12 [182] |
| Total number of studies (N) and participants (Σ), along with descriptive characteristics for the primary meta-analysis included in this systematic review that describes Δ in CRF outcomes in response to prospective, well-characterised exercise interventions lasting >2 weeks (e.g., combining exercise intervention-arms from RCTs and pre-post studies). Continuous variables are displayed as weighted means (range: lowest – highest mean values reported from studies). Categorical variables are displayed as n (%). Weighted means were calculated to account for differences in sample size between studies using the following formula: Σn*x̅ /Σn, where Σ = the sum of, n = number of participants in each study, and x̅ = mean CRF outcome of each study. AIS, American Spinal Injury Association Impairment Scale; F, females; M, males; NR, not reported; PARA, paraplegia; PPO, peak power output; TETRA, tetraplegia; V̇O_2peak_, peak oxygen consumption; W, watts. | | | |

| **Table 2.** Exercise intervention parameters reported within the included studies of the secondary, pooled meta-analysis at baseline. | | | |
| --- | --- | --- | --- |
|  | **AV̇O_2peak_**  **(L/min)** | **RV̇O_2peak_ (mL/kg/min)** | **PPO**  **(W)** |
| **Baseline CRF** | | | |
| Total number of interventions [sum of participants]  Mean (range) | 74 [849]  1.28 (0.51 – 3.50) | 78 [887]  17.8 (7.3 – 36.9) | 65 [774]  49 (0 – 168) |
| **Exercise intervention parameters** | | | |
| *Modality* | | | |
| Upper-body aerobic exercise | 26 [283] | 33 [299] | 26 [259] |
| Upper-body resistance training/circuits | 4 [33] | 5 [49] | 3 [25] |
| Functional electrical stimulation | 17 [170] | 8 [66] | 14 [140] |
| Gait/locomotor training | 10 [130] | 10 [126] | 2 [28] |
| Mixed/multimodal | 14 [166] | 20 [249] | 15 [161] |
| Behaviour change | 3 [67] | 3 [108] | 5 [161] |
| *Relative intensity* | | | |
| Light | 1 [14] | - | 1 [14] |
| Moderate | 8 [58] | 14 [114] | 10 [73] |
| Moderate-to-vigorous | 24 [332] | 26 [327] | 168 [198] |
| Vigorous | 15 [130] | 20 [194] | 11 [104] |
| Supramaximal | - | 1 [4] | 1 [10] |
| Mixed/cannot determine | 26 [315] | 18 [258] | 24 [375] |
| *Relative intensity prescription method* | | | |
| VO_2_ (%peak, %reserve) | 8 [61] | 12 [112] | 9 [93] |
| Heart rate (%HRR, %HR_peak_, _--- %HR_max_) | 18 [214] | 28 [312] | 14 [113] |
| RPE | 10 [152] | 9 [119] | 7 [98] |
| Workload (%PPO, %MTP, __  -_%1RM) | 10 [78] | 7 [56] | 8 [60] |
| Mixed/cannot determine | 28 [344] | 23 [298] | 27 [410] |
| *Session duration (min)* | 42 (20 - 90) | 42 (15 - 90) | 39 (5 - 90) |
| *Frequency (sessions/week)* | 3 (2 - 7) | 3 (2 - 7) | 3 (2 - 7) |
| < 3 | 19 [230] | 11 [108] | 13 [156] |
| ≥ 3 and < 5 | 39 [412] | 53 [582] | 40 [402] |
| ≥ 5 | 11 [116] | 11 [85] | 5 [31] |
| Not reported | 5 [91] | 4 [122] | 7 [185] |
| *Volume (min/week)* | 121 (40 - 450) | 128 (40 - 330) | 112 (15 - 330) |
| SCI-specific exercise guidelines [fitness (40 – 89 min/wk)] | 14 [147]  46 (40 - 84) | 15 [163]  48 (40 - 88) | 16 [153]  49 (15 - 88) |
| SCI-specific exercise guidelines [cardiometabolic (90 – 149 min/wk)] | 32 [328]  96 (90 - 135) | 31 [299]  100 (90 - 135) | 26 [253]  100 (90 - 135) |
| Achieving general population exercise guidelines (≥150 min/wk) | 14 [182]  209 (150 - 450) | 25 [309]  197 (150 - 330) | 14 [162]  190 (171 - 330) |
| Cannot classify | 14 [192] | 9 [166] | 9 [206] |
| *Length (weeks)* | 17 (6 - 52) | 12 (4 - 52) | 16 (4 - 52) |
| ≤ 6 weeks | 11 [95] | 23 [215] | 19 [185] |
| > 6 and ≤ 12 weeks | 36 [394] | 40 [406] | 23 [238] |
| > 12 weeks | 27 [360] | 16 [276] | 23 [351] |
| Total number of studies (N) and participants, (Σ) along with descriptive characteristics for the primary meta-analysis included in this systematic review that describes Δ in CRF outcomes in response to prospective, well-characterised exercise interventions lasting >2 weeks (e.g., combining exercise intervention-arms from RCTs and pre-post studies). Continuous variables are displayed as weighted means (range: lowest – highest mean values reported from studies). Categorical variables are displayed as n (%). Weighted means were calculated to account for differences in sample size between studies using the following formula: Σn*x̅ /Σn, where Σ = the sum of, n = number of participants in each study, and x̅ = mean CRF outcome of each study. F, females; HR _max_, maximal heart rate; HR _peak_, peak heart rate; HRR, heart rate reserve; 1RM, one repetition maximum; M, males; MTP, maximal tolerated power; NR, not reported; PPO, peak power output; V̇O_2 peak_, peak oxygen consumption; W, watts. | | | |

| **Table 3.** Summary statistics of the three subgroup analyses on injury characteristics describing Δ in CRF outcomes. | | | | | | |
| --- | --- | --- | --- | --- | --- | --- |
|  | **AV̇O_2peak_**  **(L/min)** | | **RV̇O_2peak_**  **(mL/kg/min)** | | **PPO**  **(W)** | |
|  | **N [Σ] (%)** | **WMD (95% CIs)**  ***p*-values** | **N [Σ] (%)** | **WMD (95% CIs)**  ***p-values*** | **N [Σ] (%)** | **WMD (95% CIs)**  ***p*-values** |
| **Main effect** | 74 [779] | 0.22 [0.17, 0.26]  ***p* < 0.001** | 79 [778] | 2.8 [2.3, 3.4]  ***p* < 0.001** | 65 [647] | 11 [9, 13]  ***p* < 0.001** |
| **Heterogeneity (I^2^)** | 72% (*p* < 0.001) | | 53% (*p* < 0.001) | | 78% (*p* < 0.001) | |
| ***Time since injury*** | | | | | | |
| Acute (<1-year) | 8 [96]  (12.3%) | 0.23 [0.12, 0.34]  ***p* < 0.001** | 8 [70]  (9.0%) | 3.8 [1.5, 6.1]  ***p* = 0.002** | 10 [105]  (16.2%) | 17 [12, 22]  ***p* < 0.001** |
| Chronic (≥1-year) | 49 [476]  (61.1%) | 0.20 [0.14, 0.26)  ***p* < 0.001** | 52 [466]  (59.8%) | 2.9 [2.1, 3.6]  ***p* < 0.003** | 40 [358]  (55.3%) | 10 [6, 13]  ***p* < 0.001** |
| Mixed | 8 [126]  (16.2%) | 0.23 [0.11, 0.36]  ***p* < 0.001** | 8 [121]  (15.6%) | 1.5 [0.3, 2.7]  ***p* = 0.01** | 8 [95]  (14.7%) | 6 [5, 7]  ***p* < 0.001** |
| Not reported/cannot determine | 9 [81]  (10.4%) | 0.24 [0.12, 0.36]  ***p* < 0.001** | 11 [121]  (15.6%) | 2.7 [2.0, 3.3]  ***p* < 0.003** | 7 [89]  (13.8%) | 16 [9, 23]  ***p* < 0.001** |
| Subgroup differences | - | *p* = 0.90 | - | *p* = 0.21 | - | ***p* < 0.001** |
| ***Neurological level of injury*** | | | | | | |
| Tetraplegia | 2 [18]  (2.3%) | 0.45 [-0.28, 1.19]  *p* = 0.23 | 3 [23]  (3%) | 5.9 [0.2, 11.7]  ***p* = 0.04** | 3 [23]  (3.6%) | 9 [6, 13]  ***p* < 0.002** |
| Paraplegia | 23 [202]  (25.9%) | 0.24 [0.18, 0.31]  ***p* < 0.002** | 31 [289]  (37.1%) | 3.0 [2.3, 3.6]  ***p* < 0.003** | 24 [233]  (36%) | 16 [12, 20]  ***p* < 0.002** |
| Mixed | 46 [525]  (67.4%) | 0.20 [0.15, 0.25]  ***p* < 0.002** | 43 [453]  (58.2%) | 2.0 [1.4, 2.6]  ***p* < 0.003** | 36 [378]  (58.4%) | 5 [4, 7]  ***p* < 0.002** |
| Not reported/cannot determine | 3 [34]  (4.4%) | 0.19 [0.11, 0.27]  ***p* < 0.002** | 2 [13]  (1.7%) | 2.8 [0.7, 4.8]  ***p* = 0.02** | 2 [13]  (2%) | 17 [7, 27]  ***p* = 0.001** |
| Subgroup differences | - | *p* = 0.60 | - | *p* = 0.12 |  | ***p* < 0.001** |
| ***Injury severity*** | | | | | | |
| Motor-complete  (AIS A-B) | 27 [235]  (30.2%) | 0.21 [0.14, 0.27]  ***p* < 0.002** | 31 [261]  (33.5%) | 2.9 [2.2, 3.6]  ***p* < 0.002** | 24 [210]  (32.4%) | 11 [8, 15]  ***p* < 0.002** |
| Motor-incomplete (AIS C-D) | 8 [103]  (13.2%) | 0.10 [-0.01, 0.21]  *p* = 0.08 | 13 [139]  (17.9%) | 1.6 [0.2, 2.9]  ***p* = 0.02** | 2 [14]  (2.2%) | 4 [-3, 12]  *p* = 0.25 |
| Mixed (AIS A-D) | 25 [309]  (39.7%) | 0.18 [0.13, 0.23]  ***p* < 0.002** | 23 [266]  (34.2%) | 2.6 [1.7, 3.5]  ***p* < 0.002** | 27 [311]  (48.1%) | 11 [7, 15]  ***p* < 0.002** |
| Not reported/cannot determine | 14 [132]  (16.9%) | 0.31 [0.20, 0.42]  ***p* < 0.002** | 12 [112]  (14.4%) | 3.6 [1.5, 5.6]  ***p* < 0.002** | 12 [112]  (17.3%) | 11 [6, 17]  ***p* < 0.002** |
| Subgroup differences | - | *p* = 0.06 | - | *p* = 0.27 | - | *p* = 0.41 |
| Total number of interventions (N), sum of participants analysed at post-intervention (Σ), weighting of subgroups (%). Thresholds for statistically significant subgroup differences were adjusted for the number of subgroup comparisons and are highlighted in bold: time since injury (*p*<0.0125), neurological level of injury (*p*<0.0125) and injury severity (*p*<0.0125). Individual subgroup p-values were adjusted for multiple comparisons via the Bonferroni correction method. AIS, American Spinal Injury Association Impairment Scale; AV̇O_2peak_, absolute peak oxygen consumption; CIs, confidence intervals; PPO, peak power output; RV̇O_2peak_, relative peak oxygen consumption; WMD, weighted mean difference. | | | | | | |

| **Table 4.** Summary statistics of the six subgroup analyses on exercise parameters describing Δ in CRF outcomes. | | | | | | |
| --- | --- | --- | --- | --- | --- | --- |
|  | **AV̇O_2peak_**  **(L/min)** | | **RV̇O_2peak_**  **(mL/kg/min)** | | **PPO**  **(W)** | |
|  | **N [Σ] (%)** | **WMD (95% CIs)**  ***p*-values** | **N [Σ] (%)** | **WMD (95% CIs)**  ***p-values*** | **N [Σ] (%)** | **WMD (95% CIs)**  ***p*-values** |
| **Main effect** | 74 [779] | 0.22 [0.17, 0.26]  ***p* < 0.001** | 79 [778] | 2.8 [2.3, 3.4]  ***p* < 0.001** | 65 [647] | 11 [9, 13]  ***p* < 0.001** |
| **Heterogeneity (I^2^)** | 72% (*p* < 0.001) | | 53% (*p* < 0.001) | | 78% (*p* < 0.001) | |
| ***Exercise modality*** | | | | | | |
| Aerobic, volitional upper-body | 26 [246]  (31.5%) | 0.25 [0.16, 0.33]  ***p* < 0.004** | 33 [264]  (33.9%) | 3.4 [2.4, 4.4]  ***p* < 0.004** | 26 [223]  (34.5%) | 15 [11, 19]  ***p* < 0.003** |
| Resistance training | 4 [31]  (4.0%) | 0.33 [0.13, 0.52]  ***p* = 0.003** | 5 [47]  (6%) | 5.0 [3.4, 6.7]  ***p* < 0.004** | 3 [25]  (3.9%) | 20 [12, 28]  ***p* < 0.003** |
| Functional electrical stimulation | 17 [168]  (21.6%) | 0.22 [0.15, 0.29]  ***p* < 0.004** | 8 [66]  (8.5%) | 2.4 [0.9, 3.9]  ***p* = 0.006** | 14 [138]  (21.3%) | 6 [3, 10]  ***p* < 0.003** |
| Gait training | 10 [127]  (16.3%) | 0.07 [-0.02, 0.17]  *p* = 0.14 | 10 [120]  (15.4%) | 1.0 [-0.5, 2.6]  *p* = 0.40 | 2 [24]  (3.7%) | 4 [-9, 18]  *p* = 0.54 |
| Behaviour change | 3 [49]  (6.3%) | 0.22 [0.00, 0.44]  *p* = 0.10 | 3 [41]  (5.3%) | 1.1 [-0.5, 2.6]  *p* = 0.36 | 5 [76]  (11.7%) | 8 [-1, 17]  *p* = 0.14 |
| Mixed | 14 [158]  (20.3%) | 0.20 [0.15, 0.26]  ***p* < 0.004** | 20 [240]  (30.9%) | 2.4 [1.7, 3.1]  ***p* < 0.004** | 15 [161]  (24.9%) | 12 [8, 17]  ***p* < 0.003** |
| Subgroup differences | - | *p* = 0.07 | - | ***p* = 0.002** | - | ***p* = 0.002** |
| ***Length of intervention*** | | | | | | |
| ≤6 weeks | 11 [89]  (11.4%) | 0.26 [0.20, 0.32]  ***p* < 0.001** | 23 [206]  (26.5%) | 2.9 [1.9, 3.9]  ***p* < 0.001** | 18 [169]  (26.1%) | 11 [7, 15]  ***p* < 0.001** |
| >6 – ≤12 weeks | 35 [353]  (45.3%) | 0.21 [0.14, 0.28]  ***p* < 0.001** | 40 [372]  (47.8%) | 3.3 [2.5, 4.1]  ***p* < 0.001** | 24 [217]  (33.5%) | 13 [9, 18]  ***p* < 0.001** |
| >12 weeks | 28 [337]  (43.3%) | 0.22 [0.15, 0.28]  ***p* < 0.001** | 16 [200]  (25.7%) | 1.7 [1.0, 2.4]  ***p* < 0.001** | 23 [261]  (40.4%) | 9 [5, 13]  ***p* < 0.001** |
| Subgroup differences | - | *p* = 0.54 | - | ***p* = 0.01** | - | *p* = 0.36 |
| ***Relative exercise intensity*** | | | | | | |
| Light | 1 [10]  (1.3%) | -0.05 [-0.57, 0.47]  *p* = 0.85 | - | - | 1 [10]  (1.5%) | -1 [-22, 20]  *p* = 0.92 |
| Moderate | 8 [58]  (7.4%) | 0.32 [0.09, 0.54]  ***p* = 0.01** | 14 [112]  (14.4%) | 3.5 [1.7, 5.4]  ***p* < 0.002** | 10 [71]  (11%) | 13 [4, 21]  ***p* = 0.009** |
| Moderate-to- vigorous | 24 [309]  (39.7%) | 0.21 [0.15, 0.27]  ***p* < 0.003** | 26 [301]  (38.7%) | 2.7 [2.0, 3.4]  ***p* < 0.002** | 18 [176]  (27.2%) | 17 [14, 21]  ***p* < 0.004** |
| Vigorous | 15 [120]  (15.4%) | 0.19 [0.14, 0.25]  ***p* < 0.003** | 20 [183]  (23.5%) | 2.2 [1.4, 3.0]  ***p* < 0.002** | 11 [96]  (14.9%) | 10 [5, 16]  ***p* < 0.004** |
| Supramaximal | - | - | 1 [4]  (0.5%) | 1.1 [-8.2, 10.4]  *p* = 0.82 | 1 [10]  (1.5%) | 17 [-12, 46]  *p* = 0.50 |
| Mixed/cannot determine | 26 [282]  (36.2%) | 0.22 [0.15, 0.28]  ***p* < 0.003** | 18 [178]  (22.9%) | 2.5 [1.4, 3.6]  ***p* < 0.002** | 24 [284]  (43.9%) | 8 [5, 11]  ***p* < 0.004** |
| Subgroup differences | - | *p* = 0.71 | - | *p* = 0.74 | - | ***p* = 0.004** |
| ***Exercise intensity prescription*** | | | | | | |
| Oxygen consumption | 8 [57]  (7.3%) | 0.19 [0.07, 0.32]  ***p* = 0.003** | 12 [107]  (13.8%) | 2.3 [1.5, 3.2]  ***p* < 0.002** | 9 [89]  (13.8%) | 20 [15, 25]  ***p* < 0.003** |
| Heart rate | 18 [214]  (27.5%) | 0.27 [0.15, 0.38]  ***p* < 0.002** | 28 [311]  (40%) | 3.3 [2.2, 4.4]  ***p* < 0.002** | 14 [113]  (17.5%) | 14 [8, 19]  ***p* < 0.003** |
| Rating of perceived exertion | 10 [129]  (16.6%) | 0.18 [0.09, 0.26]  ***p* < 0.002** | 9 [92]  (11.8%) | 3.4 [1.4, 5.4]  ***p* = 0.001** | 7 [74]  (11.4%) | 10 [2, 18]  ***p* = 0.02** |
| Workload | 10 [72]  (9.2%) | 0.23 [0.14, 0.33]  ***p* < 0.002** | 7 [50]  (6.4%) | 3.0 [1.3, 4.7]  ***p* < 0.002** | 8 [56]  (8.7%) | 12 [5, 19]  ***p* = 0.01** |
| Mixed/cannot determine | 28 [307]  (39.4%) | 0.21 [0.14, 0.27]  ***p* < 0.002** | 23 [218]  (28%) | 2.3 [1.4, 3.2]  ***p* < 0.002** | 27 [315]  (48.6%) | 8 [5, 11]  ***p* < 0.003** |
| Subgroup differences | - | *p* = 0.76 | - | *p* = 0.53 | - | ***p* = 0.001** |
| ***Frequency of exercise sessions*** | | | | | | |
| <3 sessions/wk | 19 [213]  (27.3%) | 0.18 [0.12, 0.25]  ***p* < 0.002** | 12 [115]  (14.8%) | 2.9 [1.5, 4.2]  ***p* < 0.002** | 13 [148]  (22.9%) | 4 [1, 6]  ***p* = 0.003** |
| ≥3 – <5 sessions/wk | 39 [379]  (48.7%) | 0.25 [0.19, 0.32]  ***p* < 0.002** | 52 [527]  (67.7%) | 2.8 [2.1, 3.5]  ***p* < 0.002** | 40 [372]  (57.5%) | 13 [10, 16]  ***p* < 0.004** |
| ≥5 sessions/wk | 11 [118]  (15.1%) | 0.15 [0.07, 0.23]  ***p* < 0.002** | 11 [85]  (10.9%) | 3.9 [2.3, 5.5]  ***p* < 0.002** | 5 [31]  (4.8%) | 10 [-2, 22]  *p* = 0.10 |
| Not reported/cannot determine | 5 [69]  (8.9%) | 0.16 [0.02, 0.31]  ***p* = 0.03** | 4 [51]  (6.6%) | 0.9 [-0.5, 2.3]  *p* = 0.21 | 7 [96]  (14.8%) | 11 [2, 19]  ***p* = 0.02** |
| Subgroup differences | - | *p* = 0.17 | - | *p* = 0.04 | - | ***p* < 0.001** |
| ***Exercise volume*** | | | | | | |
| SCI-specific exercise guidelines for fitness (40 - 89 min/wk) | 14 [139]  (17.8%) | 0.23 [0.13, 0.33]  ***p* < 0.001** | 14 [118]  (15.2%) | 3.4 [2.1, 4.7]  ***p* < 0.002** | 16 [145]  (22.4%) | 7 [3, 11]  ***p* < 0.002** |
| SCI-specific exercise guidelines for cardiometabolic health (90 - 149 min/wk) | 32 [288]  (37%) | 0.23 [0.16, 0.30]  ***p* < 0.001** | 32 [303]  (38.9%) | 2.8 [1.9, 3.7]  ***p* < 0.002** | 27 [268]  (41.4%) | 12 [9, 16]  ***p* < 0.002** |
| Achieving general population exercise guidelines (≥150 min/wk) | 14 [180]  (23.1%) | 0.18 [0.11, 0.24]  ***p* < 0.001** | 24 [262]  (33.7%) | 3.0 [2.0, 4.0]  ***p* < 0.002** | 13 [117]  (18.1%) | 11 [5, 17]  ***p* < 0.002** |
| Not reported/cannot determine | 14 [172]  (22.1%) | 0.24 [0.12, 0.36]  ***p* < 0.001** | 9 [95]  (12.2%) | 1.7 [0.6, 2.9]  ***p* = 0.003** | 9 [117]  (18.1%) | 11 [4, 18]  ***p* = 0.001** |
| Subgroup differences | - | *p* = 0.69 | - | *p* = 0.24 | - | *p* = 0.31 |
| Total number of interventions (N), sum of participants analysed at post-intervention (Σ), weighting of subgroups (%). Thresholds for statistically significant subgroup differences were adjusted for the number of subgroup comparisons and are highlighted in bold: exercise modality (*p*<0.008), length of intervention (*p*<0.017), relative exercise intensity [AV̇O_2peak_ and RV̇O_2peak_ (*p*<0.01), PPO (*p*<0.008)], exercise intensity prescription (*p*<0.01), frequency of exercise sessions (*p*<0.025), and exercise volume (*p*<0.025). Individual subgroup *p*-values were adjusted for multiple comparisons via the Bonferroni correction method. AV̇O_2peak_, absolute peak oxygen consumption; CIs, confidence intervals; PPO, peak power output; RV̇O_2peak_, relative peak oxygen consumption; WMD, weighted mean difference. | | | | | | |

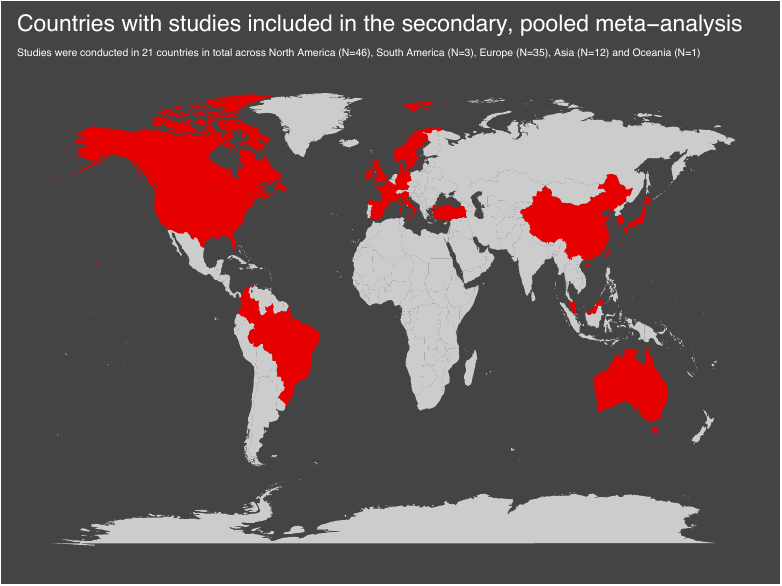

**Figure 1.** Visualisation of countries in which exercise interventions included in the secondary, pooled meta-analysis assessed were conducted. Exercise interventions were conducted worldwide across 6 continents, 21 countries.

**Table 5:** Summaries of individual pre-post studies and randomised controlled trial exercise interventions

| **Author/Year/Country/Design** | | | | **Population** | **Training Details** | **Cardiorespiratory Fitness Outcomes** |
| --- | --- | --- | --- | --- | --- | --- |
| Akkurt et al. (2017)  Turkey  RCT | | | | *N =* 17 (16M/1F)    *Age =* 31 ± 7.8 years  *TSI =* 3.7 ± 3.5 years (no. acute/chronic NR)  *Classification =* 0 T/ 17 P  *Severity =* 10 comp./ 7 incomp.  *CPET =* Progressive ACE  *CPET same modality of intervention? =* Yes  *Pre-registered? =* No  *Protocol manuscript? =* No | *Type of Exercise =* ACE  *Relative Intensity and Class =* Moderate-to-vigorous (50-70% VO_2peak_)  *Session Duration (min) =* 30  *Frequency (times/week) =* 3  *Intervention Length (weeks) =* 12  *Adverse Events =* NR | *AV̇O_2peak_ (L/min)*   - *Pre:* NR - *Post:* NR   *RV̇O_2peak_ (mL/kg/min)*   - *Pre:* 19.9 ± 8.8 - *Post:* 23.9 ± 5.1   *PPO (W)*   - *Pre:* 72 ±16 - *Post:* 103 ± 23 |
| Alrashidi et al. (2021)  Canada  Pre-post | ACET | | | *N =* 14 (10M/4F)    *Age =* 42 ± 10 years  *TSI =* 10 ± 18 years (0 acute/14 chronic)  *Classification =* 8 T/6 P  *Severity =* 14 comp./0 incomp.  *CPET =* Electronically braked ACE  *CPET same modality of intervention? =* Yes  *Pre-registered? =* Yes (NCT01718977)  *Protocol manuscript? =* Yes | *Type of Exercise =* ACE  *Relative Intensity and Class =* Moderate-to-vigorous (RPE 11 – 16)  *Session Duration (min) =* 30  *Frequency (times/week) =* 3  *Intervention Length (weeks) =* 24  *Adverse Events =* NR | *AV̇O_2peak_ (L/min)*   - *Pre:* 0.9 ± 0.4 - *Post:* 1.2 ± 0.6   *RV̇O_2peak_ (mL/kg/min)*   - *Pre:* 12.5 ± 6.7 - *Post:* 15.5 ± 8.5   *PPO (W)*   - *Pre:* 47 ± 30 - *Post:* 61 ± 46 |
|  | BWSTT | | | *N =* 14 (12M/2F)    *Age =* 39 ± 11 years  *TSI =* 6 ± 6 years (0 acute/14 chronic)  *Classification =* 7 T/7 P  *Severity =* 14 comp./0 incomp.  *CPET =* Electronically braked ACE  *CPET same modality of intervention? =* No  *Pre-registered? =* Yes (NCT01718977)  *Protocol manuscript? =* Yes | *Type of Exercise =* BWSTT  *Relative Intensity and Class =* Mixed/cannot determine  *Session Duration (min) =* 60  *Frequency (times/week) =* 3  *Intervention Length (weeks) =* 24  *Adverse Events =* NR | *AV̇O_2peak_ (L/min)*   - *Pre:* 1.1 ± 0.4 - *Post:* 1.1 ± 0.3   *RV̇O_2peak_ (mL/kg/min)*   - *Pre:* 13.3 ± 5.9 - *Post:* 13.3 ± 5.2   *PPO (W)*   - *Pre:* 61 ± 30 - *Post:* 69 ± 30 |
| Bakkum et al. (2015)  The Netherlands  Pre-post | HANDCYCLING | | | *N =* 10 (9M/1F)    *Age =* 47 ± 9 years  *TSI =* 16 ± 6 years (0 acute/10 chronic)  *Classification =* 3 T/7 P  *Severity =* 7 comp./3 incomp.  *CPET =* Handrim-propelled wheelchair  *CPET same modality of intervention? =* Yes  *Pre-registered? =* No  *Protocol manuscript? =* Yes | *Type of Exercise =* Handcycling  *Relative Intensity and Class =* Vigorous (65-75% HRR)  *Session Duration (min) =* 32  *Frequency (times/week) =* 2  *Intervention Length (weeks) =* 16  *Adverse Events =* NR | *AV̇O_2peak_ (L/min)*   - *Pre:* 1.43 ± 0.51 - *Post:* 1.48 ± 0.44   *RV̇O_2peak_ (mL/kg/min)*   - *Pre:* NR - *Post:* NR   *PPO (W)*   - *Pre:* 50.7 ± 25 - *Post:* 52.5 ± 23 |
|  | HYBRID | | | *N =* 10 (10M/0F)    *Age =* 48 ± 10 years  *TSI =* 20 ± 8 years (0 acute/10 chronic)  *Classification =* 5 T/5 P  *Severity =* 8 comp./2 incomp.  *CPET =* Wheelchair on motor driven treadmill  *CPET same modality of intervention? =* No  *Pre-registered? =* No  *Protocol manuscript? =* Yes | *Type of Exercise =* Hybrid cycling  *Relative Intensity and Class =* Vigorous (65-75% HRR)  *Session Duration (min) =* 32  *Frequency (times/week) =* 2  *Intervention Length (weeks) =* 16  *Adverse Events =* NR | *AV̇O_2peak_ (L/min)*   - *Pre:* 1.19 ± 0.38 - *Post:* 1.33 ± 0.66   *RV̇O_2peak_ (mL/kg/min)*   - *Pre:* NR - *Post:* NR   *PPO (W)*   - *Pre:* 39.5 ± 30 - *Post:* 45.4 ± 26.9 |
| Barstow et al. (1996)  USA  Pre-post | | | | *N =* 9 (9M/0F)    *Age =* 34 ± 6 years  *TSI =* 10 ± 4 years (0 acute/9 chronic)  *Classification =* 2 T/7 P  *Severity =* 9 comp./0 incomp.  *CPET =* ACE  *CPET same modality of intervention? =* Yes  *Pre-registered? =* No  *Protocol manuscript? =* No | *Type of Exercise =* FES leg cycle ergometer  *Relative Intensity and Class =* Mixed/cannot determine (Increase by 1/8 kp after 3 consecutive sessions at a given work rate)  *Session Duration (min) =* 30  *Frequency (times/week) =* 3  *Intervention Length (weeks) =* 8  *Adverse Events =* NR | *AV̇O_2peak_ (L/min)*   - *Pre:* 1.28 ± 0.31 - *Post:* 1.42 ± 0.34   *RV̇O_2peak_ (mL/kg/min)*   - *Pre:* NR - *Post:* NR   *PPO (W)*   - *Pre:* 9.9 ± 5.6 - *Post:* 14.5 ± 5.6 |
| Beillot et al. (1996)  France  Pre-post | | | | *N =* 14 (13M/1F)    *Age =* 29 ± 7 years  *TSI =* 3 ± 2 years (3 acute/11 chronic)  *Classification =* 0 T/14 P  *Severity =* 14 comp./0 incomp.  *CPET =* ACE  *CPET same modality of intervention? =* No  *Pre-registered? =* No  *Protocol manuscript? =* No | *Type of Exercise =* Walking using a reciprocating gait orthosis with FES  *Relative Intensity and Class =* Mixed/cannot determine (Walking range of at least 40m, various exercises at different speeds)  *Session Duration (min) =* NR  *Frequency (times/week) =* NR  *Intervention Length (weeks) =* 12  *Adverse Events =* 10 participants stopped due to spontaneous fracture of lower limbs, occurrence of a syringomyelia and pressure sores. | *AV̇O_2peak_ (L/min)*   - *Pre:* 1.18 ± 0.28 - *Post:* 1.18 ± 0.32   *RV̇O_2peak_ (mL/kg/min)*   - *Pre:* 20 ± 4.2 - *Post:* 20 ± 4.5   *PPO (W)*   - *Pre:* 129 ± 20 - *Post:* 131 ± 20 |
| Berry et al. (2008)  UK  Pre-post | | | | *N =* 11 (8M/3F)    *Age =* 42 ± 8 years  *TSI =* 11 ± 7 years (0 acute/11 chronic)  *Classification =* 0 T/11 P  *Severity =* 11 comp./0 incomp.  *CPET =* Electrically stimulated cycle ergometer  *CPET same modality of intervention? =* Yes  *Pre-registered? =* No  *Protocol manuscript? =* No | *Type of Exercise =* Electrically stimulated cycle training  *Relative Intensity and Class =* Mixed/cannot determine (Highest resistance level subjects could pedal against at 50 rpm until cadence dropped to 30-35 rpm)  *Session Duration (min) =* 60  *Frequency (times/week) =* 4.5  *Intervention Length (weeks) =* 52  *Adverse Events =* 1 participant had an adverse autonomic response | *AV̇O_2peak_ (L/min)*   - *Pre:* 0.54 ± 0.15 - *Post:* 0.82 ± 0.23   *RV̇O_2peak_ (mL/kg/min)*   - *Pre:* NR - *Post:* NR   *PPO (W)*   - *Pre:* 8.4 ± 3.2 - *Post:* 18.4 ± 8.7 |
| Bombardier et al. (2021)  USA  RCT behaviour change | | | | *N =* 7 (5M/2F)    *Age =* 56 ± 13 years  *TSI =* 12 ± 8 years (0 acute/ 7 chronic)  *Classification =* 0 T/ 7 P  *Severity =* NR  *CPET =* Graded maximal ACE exercise test  *CPET same modality of intervention? =* No  *Pre-registered? =* Yes (NCT02225028)  *Protocol manuscript? =* No | *Type of Behaviour Change Counselling =* Tele-health intervention (target of 150 min of MVPA per week) plus DVD*  *Number of Contact Sessions =* 16 telephone calls  *Intervention Length (weeks) =* 24  *Adverse Events =* Arm of the DXA scanner struck a participant's knee as it was returning to the start position but the physician detected no injury.  **The DVD provided verbal instructions and videos in which people with SCI demonstrated stretching, aerobic exercise, and strength training routines designed specifically for people with paraplegia or tetraplegia.* | *AV̇O_2peak_ (L/min)*   - *Pre:* NR - *Post:* NR   *RV̇O_2peak_ (mL/kg/min)*   - *Pre:* 13.4 ± 3.0 - *Post:* 13.8 ± 3.4   *PPO (W)*   - *Pre:* 69 ± 20 - *Post:* 73 ± 10 |
| Brazg et al. (2017)  USA  RCT intensity comparison | MODERATE | | | *N =* 15 (11M/4F)    *Age =* 49 ± 8.1 years  *TSI =* 7.7 ± 7.9 years (0 acute/ 15 chronic)  *Classification =* 10 T/ 5 P  *Severity =* 0 comp./ 15 incomp.  *CPET =* Modified graded peak treadmill test  *CPET same modality of intervention? =* Yes  *Pre-registered? =* Yes (NCT02115685)  *Protocol manuscript? =* No | *Type of Exercise =* Gait (locomotor) training  *Relative Intensity =* Moderate [50-60% HR_max_ (RPE 11-13)]  *Session Duration (min) =* 60  *Frequency (times/week) =* 4  *Intervention Length (weeks) =* 6  *Adverse Events =* 1 individual terminated participation due to an increase in his back pain | *AV̇O_2peak_ (L/min)*   - *Pre:* NR - *Post:* NR   *RV̇O_2peak_ (mL/kg/min)*   - *Pre:* 18 ± 6.8 - *Post:* 18 ± 6.1   *PPO (W)*   - *Pre:* NR - *Post:* NR |
|  | VIGOROUS | | | *See above: cross-over of participants following a 4-week washout period.* | *Type of Exercise =* Gait (locomotor) training  *Relative Intensity =* Vigorous [70-85% HR_max_ (RPE 15-17)]  *Session Duration (min) =* 60  *Frequency (times/week) =* 4  *Intervention Length (weeks) =* 6  *Adverse Events =* None | *AV̇O_2peak_ (L/min)*   - *Pre:* NR - *Post:* NR   RV̇O_2peak_ (mL/kg/min)   - *Pre:* 20 ± 7.9 - *Post:* 20 ± 7.5   *PPO (W)*   - *Pre:* NR - *Post:* NR |
| Bresnahan et al. (2019)  USA  Pre-post | | | | *N =* 10 (8M/2F)    *Age =* 36 ± 13 years  *TSI =* 12 ± 14 years (1 acute/9 chronic)  *Classification =* 3 T/7 P  *Severity =* 10 comp./0 incomp.  *CPET =* ACE  *CPET same modality of intervention? =* Yes  *Pre-registered? =* No  *Protocol manuscript? =* No | *Type of Exercise =* ACE  *Relative Intensity and Class =* Vigorous (70% V̇O_2peak_)  *Session Duration (min) =* 30  *Frequency (times/week) =* 3  *Intervention Length (weeks) =* 10  *Adverse Events =* NR | *AV̇O_2peak_ (L/min)*   - *Pre:* 0.78 ± 0.28 - *Post:* 0.92 ± 0.31   *RV̇O_2peak_ (mL/kg/min)*   - *Pre:* 10.8 ± 3.6 - *Post:* 12.8 ± 4   *PPO (W)*   - *Pre:* 40 ± 16 - *Post:* 54 ± 17 |
| Brurok et al. (2011)  Norway  Pre-post | | | | *N =* 6 (6M/0F)    *Age =* 40 ± 11 years  *TSI =* 18 ± 8 years (0 acute/6 chronic)  *Classification =* 1 T/5 P  *Severity =* 6 comp./0 incomp.  *CPET =* Hybrid cycling  *CPET same modality of intervention? =* Yes  *Pre-registered? =* No  *Protocol manuscript? =* No | *Type of Exercise =* Hybrid cycling and FES cycling  *Relative Intensity and Class =* Vigorous (Hybrid cycling at 80-85% of peak workload from ACE, FES cycling at 80% of default current of 140 mA.  *Session Duration (min) =* NR  *Frequency (times/week) =* 3  *Intervention Length (weeks) =* 8  *Adverse Events =* Two participants exacerbated latent shoulder dysfunction during training. | *AV̇O_2peak_ (L/min)*   - *Pre:* 1.92 ± 0.35 - *Post:* 2.43 ± 0.52   *RV̇O_2peak_ (mL/kg/min)*   - *Pre:* 24.6 ± 3.9 - *Post:* 30.6 ± 5.2   *PPO (W) *from ACE CPET*   - *Pre:* 103 ± 23 - *Post:* 118 ± 36 |
| Capodaglio et al. (1996)  Italy  RCT | | | | *N =* 4 (4M/0F)    *Age =* 31.8 ± 11.0 years  *TSI =* CD (4 acute/ 0 chronic)  *Classification =* 0 T/ 4 P  *Severity =* 4 comp./ 0 incomp.  *CPET =* Progressive ACE  *CPET same modality of intervention? =* Yes  *Pre-registered? =* No  *Protocol manuscript? =* No | *Type of Exercise =* ACE in addition to conventional rehabilitation  *Relative Intensity and Class =* Mixed/cannot determine (‘moderate subjective perception of effort’)  *Session Duration (min) =* 20-30  *Frequency (times/week) =* 5  *Intervention Length (weeks) =* 6  *Adverse Events =* NR | *AV̇O_2peak_ (L/min)*   - *Pre:* NR - *Post:* NR   *RV̇O_2peak_ (mL/kg/min)*   - *Pre:* 11.0 ± 1.6 - *Post:* 14.7 ± 1.8   *PPO (W)*   - *Pre:* NR - *Post:* NR |
| Carty et al. (2012)  Ireland  Pre-post | | | | *N =* 14 (11M/3F)    *Age =* 45 ± 8 years  *TSI =* 11 ± 11 years (3 acute/11 chronic)  *Classification =* 0 T/14 P  *Severity =* 14 comp./0 incomp.  *CPET =* Wheelchair exercise  *CPET same modality of intervention? =* No  *Pre-registered? =* No  *Protocol manuscript? =* No | *Type of Exercise =* NMES  *Relative Intensity and Class =* Moderate-to-vigorous (80-200mA to target an RPE of 13-15)  *Session Duration (min) =* 60  *Frequency (times/week) =* 5  *Intervention Length (weeks) =* 8  *Adverse Events =* NR | *AV̇O_2peak_ (L/min)*   - *Pre:* 1.09 ± 0.16 - *Post:* 1.32 ± 0.20   *RV̇O_2peak_ (mL/kg/min)*   - *Pre:* NR - *Post:* NR   *PPO (W)*   - *Pre:* NR - *Post:* NR |
| Cheung et al. (2019)  China  RCT | | | | *N =* 8 (7M/1F)    *Age =* 55.6 ± 4.98 years  *TSI =* 1.42 ± 0.58 years (CD acute/chronic)  *Classification =* CD  *Severity =* 0 comp./ 8 incomp.  *CPET =* Submaximal ACE to predict maximal outcomes  *CPET same modality of intervention? =* No  *Pre-registered? =* Yes (NCT01989806)  *Protocol manuscript? =* No | *Type of Exercise =* Physiotherapy programme plus RABWSTT  *Relative Intensity and Class =* Mixed/cannot determine  *Session Duration (min) =* 30  *Frequency (times/week) =* 3  *Intervention Length (weeks) =* 8  *Adverse Events =* None | *AV̇O_2peak_ (L/min)*   - *Pre:* NR - *Post:* NR   *RV̇O_2peak_ (mL/kg/min)*   - *Pre:* 25.7 ± 7.16 - *Post:* 26.4 ± 6.99   *PPO (W)*   - *Pre:* NR - *Post:* NR |
| Cooney and Walker (1986)  USA  Pre-post | | | | *N =* 10 (6M/4F)    *Age =* 29 ± 6 years  *TSI =* 5 ± 3 years (0 acute/10 chronic)  *Classification =* NR  *Severity =* NR  *CPET =* ACE  *CPET same modality of intervention? =* No  *Pre-registered? =* No  *Protocol manuscript? =* No | *Type of Exercise =* Hydraulic resistance exercise  *Relative Intensity and Class =* Moderate-to-vigorous (60-90% of HR_max_)  *Session Duration (min) =* 29  *Frequency (times/week) =* 3  *Intervention Length (weeks) =* 9  *Adverse Events =* NR | *AV̇O_2peak_ (L/min)*   - *Pre:* NR - *Post:* NR   *RV̇O_2peak_ (mL/kg/min)*   - *Pre:* 19.2 ± 2.3 - *Post:* 24.7 ± 3.2   *PPO (W)*   - *Pre:* 54.2 ± 10.5 - *Post:* 73.9 ± 13.0 |
| Davis et al. (1987)  Canada  Pre-post | | | | *N =* 9 (9M/0F)    *Age =* 30 ± 3 years  *TSI =* 19 ± 3 years (NR acute/NR chronic)  *Classification =* 0 T/9 P  *Severity =* NR comp./NR incomp.  *CPET =* ACE  *CPET same modality of intervention? =* Yes  *Pre-registered? =* No  *Protocol manuscript? =* No | *Type of Exercise =* ACE  *Relative Intensity and Class =* Moderate-to-vigorous (50 or 70% V̇O_2peak_)  *Session Duration (min) =* 40  *Frequency (times/week) =* 3  *Intervention Length (weeks) =* 16  *Adverse Events =* NR | *AV̇O_2peak_ (L/min)*   - *Pre:* 1.48 ± 0.10 - *Post:* 1.94 ± 0.12   *RV̇O_2peak_ (mL/kg/min)*   - *Pre:* NR - *Post:* NR   *PPO (W)*   - *Pre:* NR - *Post:* NR |
| de Groot et al. (2003)  The Netherlands  RCT intensity comparison | MODERATE | | | *N =* 3 (1M/2F)    *Age =* 52 ± 2 years  *TSI =* 0.32 ± 0.26 years (3 acute/ 0 chronic)  *Classification =* 0 T/ 3 P  *Severity =* 0 comp./ 3 incomp.  *CPET =* Graded ACE  *CPET same modality of intervention? =* Yes  *Pre-registered? =* No  *Protocol manuscript? =* No | *Type of Exercise =* ACE interspersed with boxing, push-ups and ball throwing  *Relative Intensity =* Moderate (40-55% HRR)  *Session Duration (min) =* 60  *Frequency (times/week) =* 3  *Intervention Length (weeks) =* 8  *Adverse Events =* NR | *AV̇O_2peak_ (L/min)*   - *Pre:* NR - *Post:* NR   *RV̇O_2peak_ (mL/kg/min)*   - *Pre:* 14.1 ± 2.5 - *Post:* 16.4 ± 2.9   *PPO (W)*   - *Pre:* 52 ± 20 - *Post:* 65 ± 22 |
|  | VIGOROUS | | | *N =* 3 (3M/0F)    *Age =* 39 ± 2 years  *TSI =* 0.26 ± 0.20 years (3 acute/ 0 chronic)  *Classification =* 1 T/ 2 P  *Severity =* 2 comp./ 1 incomp.  *CPET =* Graded ACE  *CPET same modality of intervention? =* Yes  *Pre-registered? =* No  *Protocol manuscript? =* No | *Type of Exercise =* ACE interspersed with boxing, push-ups and ball throwing  *Relative Intensity =* Vigorous (70-80% HRR)  *Session Duration (min) =* 60  *Frequency (times/week) =* 3  *Intervention Length (weeks) =* 8  *Adverse Events =* NR | *AV̇O_2peak_ (L/min)*   - *Pre:* NR - *Post:* NR   *RV̇O_2peak_ (mL/kg/min)*   - *Pre:* 15.1 ± 8.4 - *Post:* 21.3 ± 10.5   *PPO (W)*   - *Pre:* 68 ± 52 - *Post:* 94 ± 70 |
| DiCarlo (1988)  USA  Pre-post | | | | *N =* 8 (8M/0F)    *Age =* 24 ± 4 years  *TSI =* 9 ± 6 years (0 acute/8 chronic)  *Classification =* 8 T/0 P  *Severity =* NR comp./NR incomp.  *CPET =* Handycycling  *CPET same modality of intervention? =* Yes  *Pre-registered? =* No  *Protocol manuscript? =* No | *Type of Exercise =* ACE  *Relative Intensity and Class =* Moderate (50-60% HRR)  *Session Duration (min) =* 30  *Frequency (times/week) =* 3  *Intervention Length (weeks) =* 8  *Adverse Events =* NR | *AV̇O_2peak_ (L/min)*   - *Pre:* 0.92 ± 0.04 - *Post:* 1.75 ± 0.23   *RV̇O_2peak_ (mL/kg/min)*   - *Pre:* 12.1 ± 0.54 - *Post:* 23.5 ± 3.1   *PPO (W)*   - *Pre:* NR - *Post:* NR |
| DiPiro et al. (2016)  USA  Pre-post | | | | *N =* 10 (5M/5F)    *Age =* 58 ± 9 years  *TSI =* 11 ± 10 years (1 acute/9 chronic)  *Classification =* 9 T/1 P  *Severity =* 0 comp./10 incomp.  *CPET =* NuStep recumbent cross-trainer  *CPET same modality of intervention? =* Yes  *Pre-registered? =* No  *Protocol manuscript? =* No | *Type of Exercise =* NuStep recumbent cross-trainer  *Relative Intensity and Class =* Moderate-to-vigorous (Start at V̇O_2reserve_ and increased by 5% per week to target 60-70%)  *Session Duration (min) =* 20  *Frequency (times/week) =* 3  *Intervention Length (weeks) =* 6  *Adverse Events =* NR | *AV̇O_2peak_ (L/min)*   - *Pre:* NR - *Post:* NR   *RV̇O_2peak_ (mL/kg/min)*   - *Pre:* 20.9 ± 5.5 - *Post:* 23.9 ± 4.5   *PPO (W)*   - *Pre:* NR - *Post:* NR |
| Duffell et al. (2019)  UK  Pre-post | | | | *N =* 11 (10M/1F)    *Age =* 57 ± 9 years  *TSI =* 17 ± 7 years (5 acute/6 chronic)  *Classification =* 4 T/7 P  *Severity =* 0 comp./11 incomp.  *CPET =* FES Cycling  *CPET same modality of intervention? =* Yes  *Pre-registered? =* Yes (NCT03834324)  *Protocol manuscript? =* No | *Type of Exercise =* FES cycling  *Relative Intensity and Class =* Mixed/cannot determine  *Session Duration (min) =* 57  *Frequency (times/week) =* 3  *Intervention Length (weeks) =* 4  *Adverse Events =* NR | *AV̇O_2peak_ (L/min)*   - *Pre:* NR - *Post:* NR   *RV̇O_2peak_ (mL/kg/min)*   - *Pre:* NR - *Post:* NR   *PPO (W)*   - *Pre:* 7.2 ± 7.3 - *Post:* 10.8 ± 10.5 |
| Duran et al. (2001)  Colombia  Pre-post | | | | *N =* 13 (12M/1F)    *Age =* 26 ± 8 years  *TSI =* 2 ± 3 years (8 acute/5 chronic)  *Classification =* 0 T/13 P  *Severity =* 12 comp./1 incomp.  *CPET =* ACE  *CPET same modality of intervention? =* No  *Pre-registered? =* No  *Protocol manuscript? =* No | *Type of Exercise =* Directed physical exercise  *Relative Intensity and Class =* Moderate-to-vigorous (40-80% HR_max_)  *Session Duration (min) =* 15-40  *Frequency (times/week) =* 3  *Intervention Length (weeks) =* 16  *Adverse Events =* One patient developed sinus bradycardia and hypotension after the arm crank test | *AV̇O_2peak_ (L/min)*   - *Pre:* NR - *Post:* NR   *RV̇O_2peak_ (mL/kg/min)*   - *Pre:* NR - *Post:* NR   *PPO (W)*   - *Pre:* 90 ± 24 - *Post:* 110 ± 26 |
| El-Sayed and Younesian (2005)  UK  Pre-post | | | | *N =* 5 (NR M/NR F)    *Age =* 31 ± 3 years  *TSI =* NR  *Classification =* NR  *Severity =* NR  *CPET =* ACE  *CPET same modality of intervention? =* Yes  *Pre-registered? =* No  *Protocol manuscript? =* No | *Type of Exercise =* ACE  *Relative Intensity and Class =* Moderate-to-vigorous (60-65% V̇O_2peak_)  *Session Duration (min) =* 30  *Frequency (times/week) =* 3  *Intervention Length (weeks) =* 12  *Adverse Events = NR* | *AV̇O_2peak_ (L/min)*   - *Pre:* 1.81 ± 0.10 - *Post:* 1.94 ± 0.05   *RV̇O_2peak_ (mL/kg/min)*   - *Pre:* 24.1 ± 2.6 - *Post:* 26.2 ± 1.4   *PPO (W)*   - *Pre:* 168 ± 37.6 - *Post:* 185 ± 24.1 |
| Farkas et al. (2021)  USA  Pre-post | | ACE | | *N = 7* (5M/2F)    *Age =* 42 ± 11 years  *TSI =* NR years (0 acute/7 chronic)  *Classification =* 0 T/7 P  *Severity =* 7 comp./0 incomp.  *CPET =* ACE  *CPET same modality of intervention? =* Yes  *Pre-registered? =* Yes (NCT00270855)  *Protocol manuscript? =* No | *Type of Exercise =* ACE  *Relative Intensity and Class =* Moderate (50 rpm with progression from 20-40W to maintain 75% HR_max_)  *Session Duration (min) =* 40  *Frequency (times/week) =* 5  *Intervention Length (weeks) =* 16  *Adverse Events =* NR | *AV̇O_2peak_ (L/min)*   - *Pre:* 1.3 ± 0.3 - *Post:* 1.5 ± 0.4   *RV̇O_2peak_ (mL/kg/min)*   - *Pre:* 15.9 ± 2.6 - *Post:* 19.5 ± 4.3   *PPO (W)*   - *Pre:* 12.1 ± 6.5 - *Post:* 43.8 ± 8.9 |
|  |  | FES-LCE | | *N =* 6 (4M/2F)    *Age =* 39 ± 20 years  *TSI =* NR years (0 acute/6 chronic)  *Classification =* 0 T/6 P  *Severity =* 6 comp./0 incomp.  *CPET =* ACE  *CPET same modality of intervention? =* No  *Pre-registered? =* Yes (NCT00270855)  *Protocol manuscript? =* No | *Type of Exercise =* FES leg cycle ergometry  *Relative Intensity and Class =* Moderate (50 rpm with progression from 0.8 kp to 0.125kp to maintain 75% HR_max_)  *Session Duration (min) =* 40  *Frequency (times/week) =* 5  *Intervention Length (weeks) =* 16  *Adverse Events =* NR | *AV̇O_2peak_ (L/min)*   - *Pre:* 1.3 ± 0.3 - *Post:* 1.2 ± 0.3   *RV̇O_2peak_ (mL/kg/min)*   - *Pre:* 14.0 ± 1.3 - *Post:* 14.3 ± 2.4   *PPO (W)*   - *Pre:* 0.3 ± 0.1 - *Post:* 4.1 ± 3.8 |
| Froehlich-Grobe et al. (2022)  USA  RCT behaviour change | | | | *N =* 87 (51 M/ 36 F)    *Age =* 48.6 ± 11.5 years  *TSI =* 14.8 ± 12.2 years (NR acute/ NR chronic)  *Classification =* NR  *Severity =* NR  *CPET =* Graded maximal ACE exercise test  *CPET same modality of intervention? =* No  *Pre-registered?* = Yes (NCT03189095)  *Protocol manuscript?* = Yes | *Type of Behaviour Change Counselling =* Mixture of support mechanisms including website content (exercise and disability information plus 16 weekly modules on self-management topics), weekly 60-minute group-based virtual meetings, Facebook group for participants, exercise equipment starter package (TheraBands, seated aerobics DVD, pedal table-top arm ergometer).  *Number of Contact Sessions =* At least 16  *Intervention Length (weeks) =* 16  *Adverse Events =* NR | *AV̇O_2peak_ (L/min)*   - *Pre:* NR - *Post:* NR   *RV̇O_2peak_ (mL/kg/min) *N=20*   - *Pre:* 10.7 ± 3.3 - *Post:* 11.7 ± 3.3   *PPO (W) *N=20*   - *Pre:* 39 ± 18 - *Post:* 41 ± 20 |
| Fukuoka et al. (2006)  Japan  Pre-post | | | | *N =* 8 (7M/1F)    *Age =* 47 ± 8 years  *TSI =* NR  *Classification =* 0 T/8 P  *Severity =* 8 comp./0 incomp.  *CPET =* ACE  *CPET same modality of intervention? =* No  *Pre-registered? =* No  *Protocol manuscript? =* No | *Type of Exercise =* Wheelchair exercise  *Relative Intensity and Class =* Moderate (50% HRR)  *Session Duration (min) =* 30  *Frequency (times/week) =* 3  *Intervention Length (weeks) =* 6  *Adverse Events =* NR | *AV̇O_2peak_ (L/min)*   - *Pre:* NR - *Post:* NR   *RV̇O_2peak_ (mL/kg/min)*   - *Pre:* 17.2 ± 0.9 - *Post:* 19.9 ± 1.1   *PPO (W)*   - *Pre:* NR - *Post:* NR |
| Gant et al. (2018)  USA  Pre-post | | | | *N =* 8 (6M/2F)    *Age =* 31 ± 12 years  *TSI =* 10 ± 11 years (0 acute/8 chronic)  *Classification =* 0 T/8 P  *Severity =* 8 comp./0 incomp.  *CPET =* ACE  *CPET same modality of intervention? =* No  *Pre-registered? =* No  *Protocol manuscript? =* No | *Type of Exercise =* Multimodal training (treadmill, circuit, resistance, FES cycling)  *Relative Intensity and Class =* Mixed/cannot determine (1.2 km/hr (increase by 2.5%/wk), 35 Hz, 35 rpm)  *Session Duration (min) =* Circuit RT: 30-45, FES: 15, Wheelchair: 20-40  *Frequency (times/week) =* Circuit RT: 3, FES: 3, Wheelchair: 2, Treadmill: 2  *Intervention Length (weeks) =* 12  *Adverse Events =* NR | *AV̇O_2peak_ (L/min)*   - *Pre:* NR - *Post:* NR   *RV̇O_2peak_ (mL/kg/min)*   - *Pre:* 15.6 ± 4.1 - *Post:* 15.8 ± 4.0   *PPO (W)*   - *Pre:* NR - *Post:* NR |
| Gass et al. (1980)  Australia  Pre-post | | | | *N =* 7 (NR M/F)    *Age =* 35 ± 11 years  *TSI =* 12 ± 5 years (0 acute/7 chronic)  *Classification =* 4 T/3 P  *Severity =* 6 comp./1 incomp.  *CPET =* Wheelchair test on motor treadmill  *CPET same modality of intervention? =* Yes  *Pre-registered? =* No  *Protocol manuscript? =* No | *Type of Exercise =* Wheelchair propulsion on motor driven treadmill  *Relative Intensity and Class =* Vigorous (Progressive increase in gradient of wheelchair and speed every 5 min until exhaustion)  *Session Duration (min) =* Until exhaustion  *Frequency (times/week) =* 5  *Intervention Length (weeks) =* 7  *Adverse Events =* NR | *AV̇O_2peak_ (L/min)*   - *Pre:* 0.76 ± 0.34 - *Post:* 1.03 ± 0.42   *RV̇O_2peak_ (mL/kg/min)*   - *Pre:* 9.5 ± 4.6 - *Post:* 12.7 ± 5.9   *PPO (W)*   - *Pre:* NR - *Post:* NR |
| Gauthier et al. (2018)  Canada  RCT intensity comparison | MODERATE | | | *N =* 5 (5M/0F)    *Age =* 43.2 ± 18.5 years  *TSI =* 11.5 ± 10.3 years (0 acute/ 5 chronic)  *Classification =* 1 T/ 4 P  *Severity =* 4 comp./ 1 incomp.  *CPET =* Progressive ACE  *CPET same modality of intervention? =* No  *Pre-registered? =* No  *Protocol manuscript? =* No | *Type of Exercise =* Wheelchair propulsion  *Relative Intensity =* Moderate [4-5 RPE (CR10 scale)]  *Session Duration (min) =* 30  *Frequency (times/week) =* 3  *Intervention Length (weeks) =* 6  *Adverse Events =* NR | *AV̇O_2peak_ (L/min)*   - *Pre:* 1.65 ± 0.59 - *Post:* 1.74 ± 0.66   *RV̇O_2peak_ (mL/kg/min)*   - *Pre:* 18.5 ± 6.8 - *Post:* 18.9 ± 8.4   *PPO (W)*   - *Pre:* 80 ± 26 - *Post:* 82 ± 28 |
|  | VIGOROUS | | | *N =* 4 (3M/1F)    *Age =* 33.9 ± 3 years  *TSI =* 6 ± 3.6 years (0 acute/ 4 chronic)  *Classification =* 1 T/ 3 P  *Severity =* 3 comp./ 1 incomp.  *CPET =* Progressive ACE  *CPET same modality of intervention? =* No  *Pre-registered? =* No  *Protocol manuscript? =* No | *Type of Exercise =* Wheelchair propulsion  *Relative Intensity =* Vigorous [High intensity 6-8 RPE, low intensity 1-2 RPE (CR10 scale)]  *Session Duration (min) =* 30 (30s high intensity, 60s low intensity bouts)  *Frequency (times/week) =* 3  *Intervention Length (weeks) =* 6  *Adverse Events =* One dropout due to development of significant shoulder pain | *AV̇O_2peak_ (L/min)*   - *Pre:* 1.33 ± 0.27 - *Post:* 1.46 ± 0.32   *RV̇O_2peak_ (mL/kg/min)*   - *Pre:* 19.5 ± 0.70 - *Post:* 20.4 ± 3.9   *PPO (W)*   - *Pre:* 80 ± 18 - *Post:* 80 ± 14 |
| Gibbons et al. (2014)  UK  Pre-post | | | | *N =* 8 (4M/4F)    *Age =* 31 ± 11 years  *TSI =* 8 ± 4 years (0 acute/8 chronic)  *Classification =* 8 T/0 P  *Severity =* 6 comp./2 incomp.  *CPET =* FES rowing  *CPET same modality of intervention? =* Yes  *Pre-registered? =* No  *Protocol manuscript? =* No | *Type of Exercise =* FES rowing and leg conditioning  *Relative Intensity and Class =* Mixed/cannot determine (60-80% PPO, RPE 6-8)  *Session Duration (min) =* Rowing: 30, Leg conditioning: 60  *Frequency (times/week) =* 7  *Intervention Length (weeks) =* 52  *Adverse Events =* NR | *AV̇O_2peak_ (L/min)*   - *Pre:* NR - *Post:* NR   *RV̇O_2peak_ (mL/kg/min)*   - *Pre:* NR - *Post:* NR   *PPO (W)*   - *Pre:* 14.9 ± 10.3 - *Post:* 23.2 ± 15.6 |
| Gibbons et al. (2016)  UK  Pre-post | | | | *N =* 5 (1M/4F)    *Age =* 32 ± 5 years  *TSI =* 7 ± 7 years (0 acute/5 chronic)  *Classification =* 4 T/1 P  *Severity =* 5 comp./0 incomp.  *CPET =* FES rowing  *CPET same modality of intervention? =* Yes  *Pre-registered? =* No  *Protocol manuscript? =* No | *Type of Exercise =* FES rowing and leg conditioning  *Relative Intensity and Class =* Moderate-to-vigorous (60-80% V̇O_2peak_)  *Session Duration (min) =* 90  *Frequency (times/week) =* 7  *Intervention Length (weeks) =* 8  *Adverse Events =* Autonomic dysreflexia | *AV̇O_2peak_ (L/min)*   - *Pre:* 0.97 ± 0.22 - *Post:* 1.08 ± 0.26   *RV̇O_2peak_ (mL/kg/min)*   - *Pre:* 17.8 ± 3.5 - *Post:* 19.7 ± 4.1   *PPO (W)*   - *Pre:* 17.6 ± 11.6 - *Post:* 22.4 ± 12.9 |
| Gorgey et al. (2021)  USA  RCT | | | | *N =* 20 (18M/2F)    *Age =* 39 ± 11 years  *TSI =* 13 ± 11 years (0 acute/ 20 chronic)  *Classification =* 6 T/ 14 P  *Severity =* 17 comp./ 3 incomp.  *CPET =* FES-LCE  *CPET same modality of intervention? =* No  *Pre-registered? =* Yes (NCT01652040 and NCT02660073)  *Protocol manuscript? =* No | *Type of Exercise =* NMES-RT  *Relative Intensity and Class =* Mixed/cannot determine (4 sets of 10 reps per leg knee extension)  *Session Duration (min) =* 45-60  *Frequency (times/week) =* 2  *Intervention Length (weeks) =* 12  *Adverse Events =* NR | *AV̇O_2peak_ (L/min)*   - *Pre:* 0.513 ± 0.144 - *Post:* 0.579 ± 0.169   *RV̇O_2peak_ (mL/kg/min)*   - *Pre:* NR - *Post:* NR   *PPO (W)*   - *Pre:* 6.6 ± 3.3 - *Post:* 9 ± 4.4 |
| Gorman et al. (2016)  USA  RCT | | | | *N =* 12 (M/F NR)    *Age =* 51.5 ± 12.7 years  *TSI =* NR (0 acute/ 12 chronic)  *Classification =* 8 T/ 4 P  *Severity =* 0 comp./ 12 incomp.  *CPET* =* Peak robotic treadmill testing  *CPET same modality of intervention? =* Yes  *Pre-registered? =* Yes (NCT00385918)  *Protocol manuscript? =* No  **ACE CPET data also available* | *Type of Exercise =* RABWSTT  *Relative Intensity and Class =* Vigorous (80-85% HRR)  *Session Duration (min) =* 38.75 (First week 20-min and increased by 5-min each week thereafter)  *Frequency (times/week) =* 3  *Intervention Length (weeks) =* 12  *Adverse Events =* Two subjects developed skin irritation or abrasion in hip, groin, penis, back, wrist, glutei, and scapula. | *AV̇O_2peak_ (L/min)*   - *Pre:* NR - *Post:* NR   *RV̇O_2peak_ (mL/kg/min): RABWSTT*   - *Pre:* 20.2 ± 7.4 - *Post:* 22.7 ± 7.5   *RV̇O_2peak_ (mL/kg/min): ACE*   - *Pre:* 20.0 ± 6.5 - *Post:* 21.7 ± 7.5   *PPO (W)*   - *Pre:* NR - *Post:* NR |
| Gorman et al. (2019)  USA  Pre-post | | AQUATIC | | *N =* 15 (NR)    *Age =* 47 ± 10 years  *TSI =* 12 ± 13 years (0 acute/15 chronic)  *Classification =* 11 T/4 P  *Severity =* 0 comp./15 incomp.  *CPET =* ACE  *CPET same modality of intervention? =* No  *Pre-registered? =* Yes (NCT01407354)  *Protocol manuscript? =* No | *Type of Exercise =* Aquatic therapy  *Relative Intensity and Class =* Moderate-to-vigorous (50-75% HRR)  *Session Duration (min) =* 43  *Frequency (times/week) =* 3  *Intervention Length (weeks) =* 129  *Adverse Events =* NR | *AV̇O_2peak_ (L/min)*   - *Pre:* NR - *Post:* NR   *RV̇O_2peak_ (mL/kg/min)*   - *Pre:* 13.3 ± 3.1 - *Post:* 14.3 ± 3.9   *PPO (W)*   - *Pre:* NR - *Post:* NR |
|  |  | ROBOTIC/  LOKOMAT | | *N =* 18 (NR)    *Age =* 45 ± 13 years  *TSI =* 7 ± 4 years (0 acute/18 chronic)  *Classification =* 12 T/6 P  *Severity =* 0 comp./18 incomp.  *CPET =* Robotic treadmill walking  *CPET same modality of intervention? =* Yes  *Pre-registered? =* Yes (NCT01407354)  *Protocol manuscript? =* No | *Type of Exercise =* Robotic training  *Relative Intensity and Class =* Vigorous (65-75% HRR)  *Session Duration (min) =* 43  *Frequency (times/week) =* 3  *Intervention Length (weeks) =* 129  *Adverse Events =* NR | *AV̇O_2peak_ (L/min)*   - *Pre:* NR - *Post:* NR   *RV̇O_2peak_ (mL/kg/min)*   - *Pre:* 14.9 ± 4.3 - *Post:* 17.0 ± 6.1   *PPO (W)*   - *Pre:* NR - *Post:* NR |
| Goss et al. (1992)  USA  Pre-post | | | | *N =* 5 (3M/2F)    *Age =* 30 ± 7 years  *TSI =* 8 ± 10 years (acute/ chronic)  *Classification =* 0 T/5 P  *Severity =* 2 comp./3 incomp.  *CPET =* Computerised FES  *CPET same modality of intervention? =* Yes  *Pre-registered? =* No  *Protocol manuscript? =* No | *Type of Exercise =* Computerised FES cycle ergometry  *Relative Intensity and Class =* Mixed/cannot determine (Incremental 6W increases to level where subject could maintain pedalling speed of greater than 35 rev/min)  *Session Duration (min) =* 30  *Frequency (times/week) =* 3  *Intervention Length (weeks) =* 24  *Adverse Events =* NR | *AV̇O_2peak_ (L/min)*   - *Pre:* 0.8 ± 0.2 - *Post:* 1.0 ± 0.3   *RV̇O_2peak_ (mL/kg/min)*   - *Pre:* NR - *Post:* NR   *PPO (W)*   - *Pre:* NR - *Post:* NR |
| Graham et al. (2019)  USA  RCT intensity comparison | MODERATE | | | *N =* 3 (sex NR)    *Age =* 51.3 ± 1.2 years  *TSI =* NR (0 acute/ 3 chronic)  *Classification =* 1 T/ 2 P  *Severity =* 2 comp./ 1 incomp.  *CPET =* ACE  *CPET same modality of intervention? =* Yes  *Pre-registered? =* No  *Protocol manuscript? =* No | *Type of Exercise =* ACE  *Relative Intensity =* Moderate (55% V̇O_2peak_)  *Session Duration (min) =* 30  *Frequency (times/week) =* 3  *Intervention Length (weeks) =* 6  *Adverse Events =* NR | *AV̇O_2peak_ (L/min)*   - *Pre:* NR - *Post:* NR   *RV̇O_2peak_ (mL/kg/min)*   - *Pre:* 11.5 ± 2.6 - *Post:* 13.9 ± 1.3   *PPO (W)*   - *Pre:* NR - *Post:* NR |
|  | VIGOROUS | | | *N =* 4 (sex NR)    *Age =* 49.4 ± 13 years  *TSI =* NR (0 acute/ 4 chronic)  *Classification =* 2 T/ 2 P  *Severity =* 4 comp./ 0 incomp.  *CPET =* ACE  *CPET same modality of intervention? =* Yes  *Pre-registered? =* No  *Protocol manuscript? =* No | *Type of Exercise =* ACE  *Relative Intensity =* Supramaximal (25% HRR and 50% PPO)  *Session Duration (min) =* 20 (4-min at 25% HRR, 30-s at 50% PPO; 2-min recovery to finish at 25% HRR)  *Frequency (times/week) =* 2  *Intervention Length (weeks) =* 6  *Adverse Events =* NR | *AV̇O_2peak_ (L/min)*   - *Pre:* NR - *Post:* NR   *RV̇O_2peak_ (mL/kg/min)*   - *Pre:* 14.2 ± 6.0 - *Post:* 15.3 ± 7.3   *PPO (W)*   - *Pre:* NR - *Post:* NR |
| Grange et al. (2002)  France  Pre-post | | | | *N =* 7 (7M/0F)    *Age =* 35 ± 16 years  *TSI =* 12 ± 10 years (0 acute/14 chronic)  *Classification =* 0 T/7 P  *Severity =* 7 comp./0 incomp.  *CPET =* Graded wheelchair exercise  *CPET same modality of intervention? =* Yes  *Pre-registered? =* No  *Protocol manuscript? =* No | *Type of Exercise =* Wheelchair propulsion interval training  *Relative Intensity and Class =* Moderate-to-vigorous (4 minutes at ventilatory threshold alternated by 1 minute at maximal tolerated power)  *Session Duration (min) =* 45  *Frequency (times/week) =* 3  *Intervention Length (weeks) =* 6  *Adverse Events =* NR | *AV̇O_2peak_ (L/min)*   - *Pre:* NR - *Post:* NR   *RV̇O_2peak_ (mL/kg/min)*   - *Pre:* 27.7 ± 6.1 - *Post:* 32.0 ± 7.0   *PPO (W)*   - *Pre:* 73.7 ± 12.6 - *Post:* 88.1 ± 18.2 |
| Gurney et al. (1998)  USA  Pre-post | | | | *N =* 6 (6M/0F)    *Age =* 30 ± 7 years  *TSI =* 11 ± 8 years (0 acute/6 chronic)  *Classification =* 2 T/4 P  *Severity =* NR  *CPET =* Intermittent graded computerised FES test  *CPET same modality of intervention? =* Yes  *Pre-registered? =* No  *Protocol manuscript? =* No | *Type of Exercise =* Computerised FES leg ergometry and hybrid cycling (leg + arm cycling)  *Relative Intensity and Class =* Mixed/cannot determine (Incremental increasing time intervals and resistance in response to increasing strength of the individual)  *Session Duration (min) =* NR  *Frequency (times/week) =* 3  *Intervention Length (weeks) =* 12  *Adverse Events =* NR | *AV̇O_2peak_ (L/min)*   - *Pre:* 0.6 ± 0.1 - *Post:* 1.0 ± 0.3   *RV̇O_2peak_ (mL/kg/min)*   - *Pre:* NR - *Post:* NR   *PPO (W)*   - *Pre:* NR - *Post:* NR |
| Haddad et al. (1997)  Brazil  Pre-post | | | | *N =* 11 (7M/4F)    *Age =* 59 years (SD not reported)  *TSI =* NR  *Classification =* 0 T/11 P  *Severity =* NR  *CPET =* Incremental ACE test  *CPET same modality of intervention? =* Yes  *Pre-registered? =* No  *Protocol manuscript? =* No | *Type of Exercise =* Arm-crank ergometry  *Relative Intensity and Class =* 65 – 85% HRR  *Session Duration (min) =* 30  *Frequency (times/week) =* 3  *Intervention Length (weeks) =* 12  *Adverse Events =* NR | *AV̇O_2peak_ (L/min)*   - *Pre:* 0.93 ± 0.35 - *Post:* 1.14 ± 0.29   *RV̇O_2peak_ (mL/kg/min)*   - *Pre:* NR - *Post:* NR   *PPO (W)*   - *Pre:* NR - *Post:* NR |
| Han et al. (2016)  Taiwan  Pre-post | | | | *N =* 5 (5M/0F)    *Age =* 40 ± 7 years  *TSI =* 14 ± 5 years (0 acute/5 chronic)  *Classification =* 3 T/2 P  *Severity =* 4 comp./1 incomp.  *CPET =* ACE ramp protocol  *CPET same modality of intervention? =* Yes  *Pre-registered? =* Yes (NCT02850133)  *Protocol manuscript? =* No | *Type of Exercise =* ACE  *Relative Intensity and Class =* Moderate-to-vigorous (‘Anaerobic threshold’)  *Session Duration (min) =* 30  *Frequency (times/week) =* 3  *Intervention Length (weeks) =* 14  *Adverse Events =* NR | *AV̇O_2peak_ (L/min)*   - *Pre:* 0.9 ± 0.4 - *Post:* 0.9 ± 0.4   *RV̇O_2peak_ (mL/kg/min)*   - *Pre:* 12.3 ± 5.5 - *Post:* 13.9 ± 5.5   *PPO (W)*   - *Pre:* 39.6 ± 27.9 - *Post:* 59.6 ± 32.1 |
| Hansen et al. (2023)  Denmark  RCT | | | | *N =* 8 (5M/3F)    *Age =* 50 ± 4 years  *TSI =* NR (0 acute/ 8 chronic)  *Classification =* NR  *Severity =* 4 comp./ 4 incomp.  *CPET =* Graded arm-crank ergometry exercise test  *CPET same modality of intervention? =* No  *Pre-registered? =* No  *Protocol manuscript? =* Yes | *Type of Exercise =* Hybrid, ski ergometry  *Relative Intensity and Class =* Moderate-to-vigorous (RPE 12 – 17).  *Session Duration (min) =* 30  *Frequency (times/week) =* 3  *Intervention Length (weeks) =* 12  *Adverse Events =* NR | *AV̇O_2peak_ (L/min)*   - *Pre:* 1.24 ± 0.34 - *Post:* 1.16 ± 0.28   *RV̇O_2peak_ (mL/kg/min)*   - *Pre:* 16.8 ± 6.1 - *Post:* 16.0 ± 6.0   *PPO (W)*   - *Pre:* 64 ± 21 - *Post:* 62 ± 23 |
| Hasnan et al. (2013)  Malaysia  Pre-post | | | | *N =* 8 (NR)    *Age =* NR  *TSI =* NR (0 acute/8 chronic)  *Classification =* NR  *Severity =* NR  *CPET =* Hybrid ergometry  *CPET same modality of intervention? =* Yes  *Pre-registered? =* No  *Protocol manuscript? =* No | *Type of Exercise =* Hybrid high-intensity interval training using an arm + FES leg tricycle  *Relative Intensity and Class =* Moderate-to-vigorous (4x8 minute exercise intervals at 80-90% predicted HR_max_ with interspersed 4x8 minute intervals at 40% HR_max_)  *Session Duration (min) =* 38  *Frequency (times/week) =* 2.5  *Intervention Length (weeks) =* 6  *Adverse Events =* NR | *AV̇O_2peak_ (L/min)*   - *Pre:* 1.1 ± 0.1 - *Post:* 1.3 ± 0.1   *RV̇O_2peak_ (mL/kg/min)*   - *Pre:* 19.3 ± 3.4 - *Post:* 23.2 ± 3.4   *PPO (W)*   - *Pre:* 52.5 ± 10.4 - *Post:* 70 ± 12 |
| Heesterbeek et al. (2005)  The Netherlands  Pre-post | | | | *N =* 10 (9M/1F)    *Age =* 39 ± 9 years  *TSI =* 11 ± 6 years (NR)  *Classification =* 0 T/10 P  *Severity =* 9 comp./1 incomp.  *CPET =* Graded hybrid FES-cycle test  *CPET same modality of intervention? =* Yes  *Pre-registered? =* No  *Protocol manuscript? =* No | *Type of Exercise =* Hybrid FES cycling  *Relative Intensity and Class =* Moderate-to-vigorous (60-70% estimated V̇O_2peak_)  *Session Duration (min) =* 30  *Frequency (times/week) =* 2.5  *Intervention Length (weeks) =* 4  *Adverse Events =* NR | *AV̇O_2peak_ (L/min)*   - *Pre:* NR - *Post:* NR   *RV̇O_2peak_ (mL/kg/min)*   - *Pre:* 25.7 ± 5.8 - *Post:* 28.1 ± 7.5   *PPO (W)*   - *Pre:* 86 ± 23 - *Post:* 96 ± 26 |
| Hjeltnes et al. (1997)  Norway  Pre-post | | | | *N =* 5 (5M/0F)    *Age =* 35 ± 7 years  *TSI =* 10 ± 8 years (0 acute/5 chronic)  *Classification =* 5 T/0 P  *Severity =* 5 comp./0 incomp.  *CPET =* Graded electrically stimulated leg cycling  *CPET same modality of intervention? =* Yes  *Pre-registered? =* No  *Protocol manuscript? =* No | *Type of Exercise =* Electrically stimulated leg cycling  *Relative Intensity and Class =* Mixed/cannot determine  *Session Duration (min) =* Up to 30  *Frequency (times/week) =* 7  *Intervention Length (weeks) =* 8  *Adverse Events =* NR | *AV̇O_2peak_ (L/min)*   - *Pre:* NR - *Post:* NR   *RV̇O_2peak_ (mL/kg/min)*   - *Pre:* 7.3 ± 1.5 - *Post:* 12.5 ± 1.2   *PPO (W)*   - *Pre:* 41.7 ± 7.9 - *Post:* 42 ± 13.5 |
| Hjeltnes and Wallbeg-  Henriksson (1998)  Norway  Pre-post | | PARA | | *N =* 10 (10M/0F)    *Age =* 31 ± 4 years  *TSI =* 0.2 ± 0.01 years (10 acute/0 chronic)  *Classification =* 0 T/10 P  *Severity =* 10 comp./0 incomp.  *CPET =* ACE  *CPET same modality of intervention? =* Yes  *Pre-registered? =* No  *Protocol manuscript? =* No | *Type of Exercise =* ACE/wheelchair ergometry  *Relative Intensity and Class =* Mixed/cannot determine  *Session Duration (min) =* 30  *Frequency (times/week) =* 3  *Intervention Length (weeks) =* 8  *Adverse Events =* NR | *AV̇O_2peak_ (L/min)*   - *Pre:* 1.37 ± 0.08 - *Post:* 1.64 ± 0.10   *RV̇O_2peak_ (mL/kg/min)*   - *Pre:* 19.9 ± 1.2 - *Post:* 23.8 ± 1.4   *PPO (W)*   - *Pre:* 75 ± 6 - *Post:* 97 ± 6 |
|  |  | TETRA | | *N =* 10 (10M/0F)    *Age =* 25 ± 2 years  *TSI =* 0.3 ± 0.02 years (10 acute/0 chronic)  *Classification =* 10 T/0 P  *Severity =* 10 comp./0 incomp.  *CPET =* ACE  *CPET same modality of intervention? =* Yes  *Pre-registered? =* No  *Protocol manuscript? =* No | *Type of Exercise =* ACE/wheelchair ergometry  *Relative Intensity and Class =* Mixed/cannot determine  *Session Duration (min) =* 30  *Frequency (times/week) =* 3  *Intervention Length (weeks) =* 8  *Adverse Events =* NR | *AV̇O_2peak_ (L/min)*   - *Pre:* 0.78 ± 0.07 - *Post:* 0.86 ± 0.08   *RV̇O_2peak_ (mL/kg/min)*   - *Pre:* 11.6 ± 1.2 - *Post:* 12.9 ± 1.2   *PPO (W)*   - *Pre:* 22 ± 2 - *Post:* 32 ± 5 |
| Hoekstra et al. (2013)  The Netherlands  Pre-post | | | | *N =* 10 (4M/6F)    *Age =* 49 ± 14 years  *TSI =* NR (2 acute/8 chronic)  *Classification =* 3 T/7 P  *Severity =* 0 comp./10 incomp.  *CPET =* Discontinuous progressive ACE  *CPET same modality of intervention? =* No  *Pre-registered? =* Yes (ISRCTN67827069)  *Protocol manuscript? =* No | *Type of Exercise =* Robotic gait training  *Relative Intensity and Class =* Mixed/cannot determine (Individually adapted walking speed to walk comfortably for 30 minutes)  *Session Duration (min) =* 40  *Frequency (times/week) =* 2.5  *Intervention Length (weeks) =* 13  *Adverse Events =* NR | *AV̇O_2peak_ (L/min)*   - *Pre:* 1.16 ± 0.41 - *Post:* 1.21 ± 0.4   *RV̇O_2peak_ (mL/kg/min)*   - *Pre:* 15.7 ± 5.1 - *Post:* 16.5 ± 5.7   *PPO (W)*   - *Pre:* NR - *Post:* NR |
| Hooker and Wells (1989)  USA  RCT intensity comparison | MODERATE | | | *N =* 6 (3M/3F)    *Age =* 31.3 ± 4.2 years  *TSI =* 12.88 ± 6.8 years (1 acute/ 5 chronic)  *Classification =* 1 T/ 5 P  *Severity =* NR  *CPET =* Incremental discontinuous wheelchair ergometer exercise  *CPET same modality of intervention? =* Yes  *Pre-registered? =* No  *Protocol manuscript? =* No | *Type of Exercise =* Wheelchair ergometry  *Relative Intensity =* Moderate (50-60% HRR)  *Session Duration (min) =* 20  *Frequency (times/week) =* 3  *Intervention Length (weeks) =* 8  *Adverse Events =* NR | *AV̇O_2peak_ (L/min)*   - *Pre:* NR - *Post:* NR   *RV̇O_2peak_ (mL/kg/min)*   - *Pre:* 19.4 ± 8.1 - *Post:* 21.4 ± 6.5   *PPO (W)*   - *Pre:* 33 ± 29 - *Post:* 41 ± 30 |
|  | VIGOROUS | | | *N =* 5 (3M/2F)    *Age =* 30.4 ± 5 years  *TSI =* 10.2 ± 7.9 years (0 acute/ 5 chronic)  *Classification =* 2 T/ 3 P  *Severity =* NR  *CPET =* Incremental discontinuous wheelchair ergometer exercise  *CPET same modality of intervention? =* Yes  *Pre-registered? =* No  *Protocol manuscript? =* No | *Type of Exercise =* Wheelchair ergometry  *Relative Intensity =* Vigorous (70-80% HRR)  *Session Duration (min) =* 20  *Frequency (times/week) =* 3  *Intervention Length (weeks) =* 8  *Adverse Events =* NR | *AV̇O_2peak_ (L/min)*   - *Pre:* NR - *Post:* NR   *RV̇O_2peak_ (mL/kg/min)*   - *Pre:* 19.2 ± 9.8 - *Post:* 21.5 ± 9.2   *PPO (W)*   - *Pre:* 33 ± 33 - *Post:* 38 ± 36 |
| Hooker et al. (1992)  USA  Pre-post | | | | *N =* 18 (17M/1F)    *Age =* 31 ± 2 years  *TSI =* 6 ± 1 years (1 acute/17 chronic)  *Classification =* 10 T/8 P  *Severity =* 11 comp./7 incomp.  *CPET =* Discontinuous graded FES-leg ergometry  *CPET same modality of intervention? =* Yes  *Pre-registered? =* No  *Protocol manuscript? =* No | *Type of Exercise =* FES leg cycling  *Relative Intensity and Class =* Mixed/cannot determine (30 continuous minutes with 6.1W increase after completion of three consecutive training sessions)  *Session Duration (min) =* 30  *Frequency (times/week) =* 3  *Intervention Length (weeks) =* 14  *Adverse Events =* NR | *AV̇O_2peak_ (L/min)*   - *Pre:* 0.88 ± 0.01 - *Post:* 0.95 ± 0.01   *RV̇O_2peak_ (mL/kg/min)*   - *Pre:* NR - *Post:* NR   *PPO (W)*   - *Pre:* 13.6 ± 0.4 - *Post:* 19.7 ± 0.4 |
| Hooker et al. (1995)  USA  Pre-post | | | | *N =* 8 (8M/0F)    *Age =* 36 ± 5 years  *TSI =* 10 ± 4 years (0 acute/8 chronic)  *Classification =* 2 T/6 P  *Severity =* 8 comp./0 incomp.  *CPET =* Graded NMES leg cycling and ACE  *CPET same modality of intervention? =* Yes  *Pre-registered? =* No  *Protocol manuscript? =* No | *Type of Exercise =* NMES leg cycling  *Relative Intensity and Class =* Mixed/cannot determine (30 continuous minutes with 6.1W increase after completion of three consecutive training sessions)  *Session Duration (min) =* 30  *Frequency (times/week) =* 2  *Intervention Length (weeks) =* 27  *Adverse Events =* NR | *AV̇O_2peak_ (L/min)*   - *Pre:* 1.29 ± 0.30 - *Post:* 1.42 ± 0.39   *RV̇O_2peak_ (mL/kg/min)*   - *Pre:* 17.7 ± 5.8 - *Post:* 19.4 ± 7.5   *PPO (W)*   - *Pre:* 12.2 ± 5.6 - *Post:* 15.3 ± 4.6 |
| Horiuchi and Okita (2017)  Japan  Pre-post | | | | *N =* 9 (9M/0F)    *Age =* 38 ± 10 years  *TSI =* 16 ± 7 years (0 acute/9 chronic)  *Classification =* 0 T/9 P  *Severity =* 9 comp./0 incomp.  *CPET =* Incremental ACE  *CPET same modality of intervention? =* Yes  *Pre-registered? =* No  *Protocol manuscript? =* No | *Type of Exercise =* ACE  *Relative Intensity and Class =* Moderate-to-vigorous (50-70% HRR)  *Session Duration (min) =* 60  *Frequency (times/week) =* 4  *Intervention Length (weeks) =* 10  *Adverse Events =* NR | *AV̇O_2peak_ (L/min)*   - *Pre:* 1.8 ± 0.3 - *Post:* 1.9 ± 0.3   *RV̇O_2peak_ (mL/kg/min)*   - *Pre:* 28.9 ± 4.1 - *Post:* 32.7 ± 4.4   *PPO (W)*   - *Pre:* NR - *Post:* NR |
| Jacobs et al. (1997)  USA  Pre-post | | | | *N =* 15 (12M/3F)    *Age =* 28 ± 7 years  *TSI =* 4 ± 3 years (NR)  *Classification =* 0 T/15 P  *Severity =* 15 comp./0 incomp.  *CPET =* ACE  *CPET same modality of intervention? =* No  *Pre-registered? =* No  *Protocol manuscript? =* No | *Type of Exercise =* FNS ambulation training  *Relative Intensity and Class =* Mixed/cannot determine (Subjects selected their own ambulation pace and duration)  *Session Duration (min) =* NR  *Frequency (times/week) =* 3  *Intervention Length (weeks) =* 11  *Adverse Events =* NR | *AV̇O_2peak_ (L/min)*   - *Pre:* NR - *Post:* NR   *RV̇O_2peak_ (mL/kg/min)*   - *Pre:* 20.0 ± 3.3 - *Post:* 23.0 ± 3.6   *PPO (W)*   - *Pre:* 48.1 ± 13.4 - *Post:* 60.0 ± 18.1 |
| Jacobs et al. (2001)  USA  Pre-post | | | | *N =* 10 (10M/0F)    *Age =* 39 ± 6 years  *TSI =* 7 ± 6 years (NR)  *Classification =* 0 T/10 P  *Severity =* 10 comp./0 incomp.  *CPET =* Hydraulically braked discontinuous ACE  *CPET same modality of intervention? =* Yes  *Pre-registered? =* No  *Protocol manuscript? =* No | *Type of Exercise =* Circuit RT  *Relative Intensity and Class =* Mixed/cannot determine  *Session Duration (min) =* 45  *Frequency (times/week) =* 3  *Intervention Length (weeks) =* 12  *Adverse Events =* NR | *AV̇O_2peak_ (L/min)*   - *Pre:* 1.45 ± 0.22 - *Post:* 1.88 ± 0.31   *RV̇O_2peak_ (mL/kg/min)*   - *Pre:* 18.7 ± 4.2 - *Post:* 23.8 ± 6.0   *PPO (W)*   - *Pre:* 109 ± 17 - *Post:* 127 ± 18 |
| Jacobs (2009)  USA  Pre-post | | ACE | | *N =* 9 (6M/3F)    *Age =* 29 ± 10 years  *TSI =* NR  *Classification =* 0 T/9 P  *Severity =* 9 comp./0 incomp.  *CPET =* Discontinuous, multistage ACE  *CPET same modality of intervention? =* Yes  *Pre-registered? =* No  *Protocol manuscript? =* No | *Type of Exercise =* ACE  *Relative Intensity and Class =* Moderate-to-vigorous (70-85% HR_peak_)  *Session Duration (min) =* 30  *Frequency (times/week) =* 3  *Intervention Length (weeks) =* 12  *Adverse Events =* NR | *AV̇O_2peak_ (L/min)*   - *Pre:* 1.27 ± 0.54 - *Post:* 1.42 ± 0.64   *RV̇O_2peak_ (mL/kg/min)*   - *Pre:* NR - *Post:* NR   *PPO (W)*   - *Pre:* NR - *Post:* NR |
|  |  | RT | | *N =* 9 (6M/3F)    *Age =* 34 ± 8 years  *TSI =* NR  *Classification =* 0 T/9 P  *Severity =* 9 comp./0 incomp.  *CPET =* Discontinuous, multistage ACE  *CPET same modality of intervention? =* Yes  *Pre-registered? =* No  *Protocol manuscript? =* No | *Type of Exercise =* Resistance training  *Relative Intensity and Class =* Mixed/cannot determine  *Session Duration (min) =* NR  *Frequency (times/week) =* 3  *Intervention Length (weeks) =* 12  *Adverse Events =Type of Exercise =* NR | *AV̇O_2peak_ (L/min)*   - *Pre:* 1.19 ± 0.52 - *Post:* 1.37 ± 0.67   *RV̇O_2peak_ (mL/kg/min)*   - *Pre:* NR - *Post:* NR   *PPO (W)*   - *Pre:* NR - *Post:* NR |
| Janssen and Pringle (2008)  The Netherlands  Pre-post | | | | *N =* 12 (12M/0F)    *Age =* 36 ± 16 years  *TSI =* 16 ± 11 years (1 acute/11 chronic)  *Classification =* 6 T/6 P  *Severity =* 9 comp./3 incomp.  *CPET =* Graded leg cycle ergometry with and without modified electrical stimulation  *CPET same modality of intervention? =* Yes  *Pre-registered? =* No  *Protocol manuscript? =* No | *Type of Exercise =* Modified electrical stimulation-induced leg cycle ergometry  *Relative Intensity and Class =* Mixed/cannot determine  *Session Duration (min) =* 30  *Frequency (times/week) =* 2.5  *Intervention Length (weeks) =* 6  *Adverse Events =Type of Exercise =* A few subjects’ experiences lightheadedness | *AV̇O_2peak_ (L/min)*   - *Pre:* 0.82 ± 0.29 - *Post:* 1.07 ± 0.26   *RV̇O_2peak_ (mL/kg/min)*   - *Pre:* NR - *Post:* NR   *PPO (W)*   - *Pre:* 8.6 ± 9.9 - *Post:* 13.5 ± 9.7 |
| Jeon et al. (2010)  South Korea  Pre-post | | | | *N =* 6 (6M/0F)    *Age =* 46 ± 5 years  *TSI =* 21 ± 5 years (0 acute/6 chronic)  *Classification =* 0 T/6 P  *Severity =* 6 comp./0 incomp.  *CPET =* FES rowing  *CPET same modality of intervention? =* Yes  *Pre-registered? =* No  *Protocol manuscript? =* No | *Type of Exercise =* FES rowing  *Relative Intensity and Class =* Moderate-to-vigorous (5-minute sets at 80% V̇O_2peak_)  *Session Duration (min) =* 27.5  *Frequency (times/week) =* 3.5  *Intervention Length (weeks) =* 12  *Adverse Events =* NR | *AV̇O_2peak_ (L/min)*   - *Pre:* 1.5 ± 0.1 - *Post:* 1.6 ± 0.1   *RV̇O_2peak_ (mL/kg/min)*   - *Pre:* 21.4 ± 1.2 - *Post:* 23.1 ± 0.8   *PPO (W)*   - *Pre:* 33 ± 2.7 - *Post:* 55 ± 3.5 |
| Kesiktas et al. (2021)  Turkey  Pre-post | | | Home-based, unsupervised | *N =* 10 (10M/0F)    *Age =* 36 ± 6.7 years  *TSI =* 7.8 ± 1.7 years (0 acute/10 chronic)  *Classification =* 0 T/10 P  *Severity =* 10 comp./0 incomp.  *CPET =* Multistep arm crank ergometry exercise test  *CPET same modality of intervention? =* No  *Pre-registered? =* No  *Protocol manuscript? =* No | *Type of Exercise =* Circuit training consisting of upper-extremity stretching and strengthening with elastic bands. Home-based exercise sessions, unsupervised.  *Relative Intensity and Class =* Moderate (70% HR_peak_).  *Session Duration (min) =* 60  *Frequency (times/week) =* 5  *Intervention Length (weeks) =* 8  *Adverse Events =* NR | *AV̇O_2peak_ (L/min)*   - *Pre:* NR - *Post:* NR   *RV̇O_2peak_ (mL/kg/min)*   - *Pre:* 15.5 ± 2.6 - *Post:* 19.0 ± 3.7   *PPO (W)*   - *Pre:* NR - *Post:* NR |
|  |  |  | Hospital-based, supervised | *N =* 10 (10M/0F)    *Age =* 35 ± 9.6 years  *TSI =* 8 ± 1.6 years (0 acute/10 chronic)  *Classification =* 0 T/10 P  *Severity =* 10 comp./0 incomp.  *CPET =* Multistep arm crank ergometry exercise test  *CPET same modality of intervention? =* No  *Pre-registered? =* No  *Protocol manuscript? =* No | *Type of Exercise =* Circuit training consisting of upper-extremity stretching and strengthening with elastic bands. Home-based exercise sessions, unsupervised.  *Relative Intensity and Class =* Moderate (70% HR_peak_).  *Session Duration (min) =* 60  *Frequency (times/week) =* 5  *Intervention Length (weeks) =* 8  *Adverse Events =* NR | *AV̇O_2peak_ (L/min)*   - *Pre:* NR - *Post:* NR   *RV̇O_2peak_ (mL/kg/min)*   - *Pre:* 16.5 ± 3.0 - *Post:* 24.0 ± 4.4   *PPO (W)*   - *Pre:* NR   *Post:* NR |
| Kim et al. (2014)  South Korea  Pre-post | | | | *N =* 12 (10M/2F)    *Age =* 36 ± 12 years  *TSI =* 11 ± 6 years (0 acute/12 chronic)  *Classification =* 2 T/10 P  *Severity =* 8 comp./4 incomp.  *CPET =* ACE  *CPET same modality of intervention? =* No  *Pre-registered? =* No  *Protocol manuscript? =* No | *Type of Exercise =* FES rowing  *Relative Intensity and Class =* Vigorous (>70% HR_max_)  *Session Duration (min) =* 30  *Frequency (times/week) =* 5  *Intervention Length (weeks) =* 6  *Adverse Events =* NR | *AV̇O_2peak_ (L/min)*   - *Pre:* NR - *Post:* NR   *RV̇O_2peak_ (mL/kg/min)*   - *Pre:* 17.2 ± 3.8 - *Post:* 19.4 ± 4.7   *PPO (W)*   - *Pre:* NR - *Post:* NR |
| Kim et al. (2015)  South Korea  RCT | | | | *N =* 8 (6M/2F)    *Age =* 31.5 ± 5.5 years  *TSI =* 5 ± 3.2 years (0 acute/ 8 chronic)  *Classification =* 6 T/ 2 P  *Severity =* 8 comp./ 0 incomp.  *CPET =* Maximal graded handbike exercise test  *CPET same modality of intervention? =* Yes  *Pre-registered? =* No  *Protocol manuscript? =* No | *Type of Exercise =* Handbike exercise  *Relative Intensity and Class =* Moderate-to-vigorous (Wk1-2: RPE 5, 70% HR_max_; Wk2-4: RPE 6, 75% HR_max_; Wk4-6: RPE 7, 80% HR_max_)  *Session Duration (min) =* 60  *Frequency (times/week) =* 3  *Intervention Length (weeks) =* 6  *Adverse Events =* NR | *AV̇O_2peak_ (L/min)*   - *Pre:* NR - *Post:* NR   *RV̇O_2peak_ (ml/kg/min)*   - *Pre:* 16.8 ± 7.2 - *Post:* 21.2 ± 9.1   *PPO (W)*   - *Pre:* NR - *Post:* NR |
| Kim et al. (2019)  South Korea  RCT | | | | *N =* 11 (M/F NR)    *Age =* NR  *TSI =* NR (0 acute/ 11 chronic)  *Classification =* NR  *Severity =* NR  *CPET =* Maximal ACE  *CPET same modality of intervention? =* Yes  *Pre-registered? =* No  *Protocol manuscript? =* No | *Type of Exercise =* Mixed (ACE and resistance exercises)  *Relative Intensity and Class =* Moderate-to-vigorous (6-8 RPE)  *Session Duration (min) =* 60  *Frequency (times/week) =* 3  *Intervention Length (weeks) =* 6  *Adverse Events =* NR | *AV̇O_2peak_ (L/min)*   - *Pre:* NR - *Post:* NR   *RV̇O_2peak_ (mL/kg/min)*   - *Pre:* 11.7 ± 8.1 - *Post:* 15.2 ± 9.6   *PPO (W)*   - *Pre:* NR - *Post:* NR |
| Kjaer et al. (2001)  Denmark  Pre-post | | | | *N =* 10 (8M/2F)    *Age =* 35 ± 5 years  *TSI =* 12 ± 6 years (0 acute/10 chronic)  *Classification =* 10 T/0 P  *Severity =* 10 comp./0 incomp.  *CPET =* Incremental workload test  *CPET same modality of intervention? =* Yes  *Pre-registered? =* No  *Protocol manuscript? =* No | *Type of Exercise =* FES leg cycle ergometry  *Relative Intensity and Class =* Vigorous (Workload as high as possible with addition of 0.125kPa resistance if able to cycle for 30 min for >2-3 weeks)  *Session Duration (min) =* 30  *Frequency (times/week) =* 2.3  *Intervention Length (weeks) =* 52  *Adverse Events =* NR | *AV̇O_2peak_ (L/min)*   - *Pre:* 1.20 ± 0.30 - *Post:* 1.38 ± 0.30   *RV̇O_2peak_ (mL/kg/min)*   - *Pre:* NR - *Post:* NR   *PPO (W)*   - *Pre:* NR - *Post:* NR |
| Koch et al. (1983)  Germany  Pre-post | | | | *N =* 10 (10M/0F)    *Age =* 22.6 ± 5.5 years  *TSI =* 0.39 ± 0.12 years (144 ± 44 days) (10 acute)  *Classification =* 0 T/10 P  *Severity =* NR  *CPET =* Incremental arm-crank test  *CPET same modality of intervention? =* No  *Pre-registered? =* No  *Protocol manuscript? =* No | *Type of Exercise =* Multimodal [“to develop endurance, strength, speed and coordinative skills (agility, flexibility and responsiveness)”]  *Relative Intensity and Class =* NR  *Session Duration (min) =* NR  *Frequency (times/week) =* NR  *Intervention Length (weeks) =* 6  *Adverse Events =* NR | *AV̇O_2peak_ (L/min)*   - *Pre:* 1.36 ± 0.33 - *Post:* 1.61 ± 0.30   *RV̇O_2peak_ (mL/kg/min)*   - *Pre:* NR - *Post:* NR   *PPO (W)*   - *Pre:* 42 ± 9 - *Post:* 64 ± 10 |
| Kooijmans et al. (2017)  The Netherlands  RCT behaviour change | | | | *N =* 33 (21M/12F)    *Age =* 48 ± 10 years  *TSI =* 21 ± 8 years (0 acute/ 33 chronic)  *Classification =* 11 T/ 22 P  *Severity =* 24 comp./ 9 incomp.  *CPET =* Maximal wheelchair treadmill test  *CPET same modality of intervention? =* No  *Pre-registered? =* Yes (ISRCTN11233847)  *Protocol manuscript? =* Yes | *Type of Behaviour Change Counselling =* Self-management intervention (group meetings, book and individual counselling)  *Number of Contact Sessions =* 12 (2 home visits + 5 individual sessions + 5 group sessions +)  *Intervention Length (weeks) =* 42  *Adverse Events =* NR | *AV̇O_2peak_ (L/min) *N=15*   - *Pre:* 3.50 ± 5.70 - *Post:* 3.00 ± 5.50   *RV̇O_2peak_ (mL/kg/min)*   - *Pre:* NR - *Post:* NR   *PPO (W) *N=15*   - *Pre:* 49 ± 23 - *Post:* 58 ± 26 |
| Koontz et al. (2021)  USA  Pre-post | | | | *N =* 10 (7M/3F)    *Age =* 39 ± 14 years  *TSI =* 12 ± 11 years (0 acute/10 chronic)  *Classification =* 7 T/3 P  *Severity =* 2 comp./8 incomp.  *CPET =* ACE  *CPET same modality of intervention? =* Yes  *Pre-registered? =* Yes (NCT03152110)  *Protocol manuscript? =* No | *Type of Exercise =* Handcycling  *Relative Intensity and Class =* Vigorous (10x1 min intervals of 90% PPO separated by 1 min intervals of 0-20% PPO)  *Session Duration (min) =* 25  *Frequency (times/week) =* 2  *Intervention Length (weeks) =* 6  *Adverse Events =* One participant experienced bad spasms during a session | *AV̇O_2peak_ (L/min)*   - *Pre:* 1.09 ± 0.48 - *Post:* 1.09 ± 0.44   *RV̇O_2peak_ (mL/kg/min)*   - *Pre:* 14.3 ± 5.0 - *Post:* 14.3 ± 4.8   *PPO (W)*   - *Pre:* 60 ± 33.3 - *Post:* 65 ± 38.5 |
| Krauss et al. (1993)  USA  Pre-post | | CFES | | *N =* 8 (7M/1F)    *Age =* 32 ± 2 years  *TSI =* 13 ± 2 years (0 acute/8 chronic)  *Classification =* 1 T/7 P  *Severity =* NR  *CPET =* Computerised FES  *CPET same modality of intervention? =* Yes  *Pre-registered? =* No  *Protocol manuscript? =* No | *Type of Exercise =* Computerised FES leg cycle ergometry  *Relative Intensity and Class =* Mixed/cannot determine  *Session Duration (min) =* 30  *Frequency (times/week) =* 3  *Intervention Length (weeks) =* 6  *Adverse Events =* NR | *AV̇O_2peak_ (L/min)*   - *Pre:* 0.5 ± 0.04 - *Post:* 0.8 ± 0.2   *RV̇O_2peak_ (mL/kg/min)*   - *Pre:* NR - *Post:* NR   *PPO (W)*   - *Pre:* NR - *Post:* NR |
|  |  | HYBRID | | *N =* 8 (7M/1F)    *Age =* 32 ± 2 years  *TSI =* 13 ± 2 years (0 acute/8 chronic)  *Classification =* 1 T/7 P  *Severity =* NR  *CPET =* ACE, computerised FES, and intermittent hybrid ergometry  *CPET same modality of intervention? =* Yes  *Pre-registered? =* No  *Protocol manuscript? =* No | *Type of Exercise =* Hybrid ergometry with computerised FES + ACE  *Relative Intensity and Class =* Moderate (50% of PPO attained during CPET)  *Session Duration (min) =* 30  *Frequency (times/week) =* 3  *Intervention Length (weeks) =* 6  *Adverse Events =* NR | *AV̇O_2peak_ (L/min)*   - *Pre:* 1.3 ± 0.2 - *Post:* 1.5 ± 0.2   *RV̇O_2peak_ (mL/kg/min)*   - *Pre:* NR - *Post:* NR   *PPO (W)*   - *Pre:* 45.6 ± 4.5 - *Post:* 46.2 ± 4.6 |
| Kressler et al. (2013)  USA  Pre-post | | OVERGROUND WALKING | | *N =* 14 (12M/2F)    *Age =* 41 ± 17 years  *TSI =* NR (0 acute/14 chronic)  *Classification =* 11 T/3 P  *Severity =* 0 comp./14 incomp.  *CPET =* Maximal treadmill walking  *CPET same modality of intervention? =* Yes  *Pre-registered? =* Yes (part of R01HD41487)  *Protocol manuscript? =* No | *Type of Exercise =* Walking with transcutaneous FES  *Relative Intensity and Class =* Moderate (Equal to or above 13 RPE)  *Session Duration (min) =* NR  *Frequency (times/week) =* 5  *Intervention Length (weeks) =* 12  *Adverse Events =* NR | *AV̇O_2peak_ (L/min)*   - *Pre:* 1.00 ± 0.39 - *Post:* 1.13 ± 0.45   *RV̇O_2peak_ (mL/kg/min)*   - *Pre:* NR - *Post:* NR   *PPO (W)*   - *Pre:* NR - *Post:* NR |
|  |  | TREADMILL + FES | | *N =* 17 (13M/4F)    *Age =* 38 ± 11 years  *TSI =* NR (0 acute/17 chronic)  *Classification =* 10 T/7 P  *Severity =* 0 comp./17 incomp.  *CPET =* Maximal treadmill walking  *CPET same modality of intervention? =* Yes  *Pre-registered? =* Yes (part of R01HD41487)  *Protocol manuscript? =* No | *Type of Exercise =* Treadmill-based locomotor training with transcutaneous FES  *Relative Intensity and Class =* Moderate (Equal to or above 13 RPE)  *Session Duration (min) =* NR  *Frequency (times/week) =* 5  *Intervention Length (weeks) =* 12  *Adverse Events =* Increase in spasticity, increase in chronic back pain in a participant | *AV̇O_2peak_ (L/min)*   - *Pre:* 1.07 ± 0.36 - *Post:* 1.17 ± 0.44   *RV̇O_2peak_ (mL/kg/min)*   - *Pre:* NR - *Post:* NR   *PPO (W)*   - *Pre:* NR - *Post:* NR |
|  |  | TREADMILL | | *N =* 17 (12M/5F)    *Age =* 45 ± 13 years  *TSI =* NR (0 acute/17 chronic)  *Classification =* 10 T/7 P  *Severity =* 0 comp./17 incomp.  *CPET =* Maximal treadmill walking  *CPET same modality of intervention? =* Yes  *Pre-registered? =* Yes (part of R01HD41487)  *Protocol manuscript? =* No | *Type of Exercise =* Treadmill-based locomotor training with manual assistance  *Relative Intensity and Class =* Moderate (Equal to or above 13 RPE)  *Session Duration (min) =* NR  *Frequency (times/week) =* 5  *Intervention Length (weeks) =* 12  *Adverse Events =* NR | *AV̇O_2peak_ (L/min)*   - *Pre:* 0.97 ± 0.25 - *Post:* 1.17 ± 0.35   *RV̇O_2peak_ (mL/kg/min)*   - *Pre:* NR - *Post:* NR   *PPO (W)*   - *Pre:* NR - *Post:* NR |
|  |  | DRIVEN GAIT ORTHOSIS | | *N =* 14 (12M/2F)    *Age =* 44 ± 8 years  *TSI =* NR (0 acute/14 chronic)  *Classification =* 5 T/9 P  *Severity =* 0 comp./14 incomp.  *CPET =* Maximal treadmill walking  *CPET same modality of intervention? =* Yes  *Pre-registered? =* Yes (part of R01HD41487)  *Protocol manuscript? =* No | *Type of Exercise =* Driven gait orthosis  *Relative Intensity and Class =* Moderate (Equal to or above 13 RPE)  *Session Duration (min) =* NR  *Frequency (times/week) =* 5  *Intervention Length (weeks) =* 12  *Adverse Events =* NR | *AV̇O_2peak_ (L/min)*   - *Pre:* 1.32 ± 0.40 - *Post:* 1.28 ± 0.40   *RV̇O_2peak_ (mL/kg/min)*   - *Pre:* NR - *Post:* NR   *PPO (W)*   - *Pre:* NR - *Post:* NR |
| Lavado et al. (2013)  Brazil  RCT | | | | *N =* 21 (18M/3F)    *Age =* 34.1 ± 11.1 years  *TSI =* 4.8 ± 2.2 years (0 acute/ 21 chronic)  *Classification =* NR  *Severity =* NR  *CPET =* Maximal ACE  *CPET same modality of intervention? =* Yes  *Pre-registered? =* No  *Protocol manuscript? =* No | *Type of Exercise =* Physiotherapy sessions plus aerobic training (cycloergometer of upper limbs, performed distance with a wheelchair, and general exercises to gain muscle power)  *Relative Intensity and Class =* Moderate-to-vigorous (70-80% HR_max_ or VO_2peak_)  *Session Duration (min) =* 60  *Frequency (times/week) =* 2-3  *Intervention Length (weeks) =* 16  *Adverse Events =* None | *AV̇O_2peak_ (L/min)*   - *Pre:* 0.96 ± 0.37 - *Post:* 1.17 ± 0.26   *RV̇O_2peak_ (mL/kg/min)*   - *Pre:* NR - *Post:* NR   *PPO (W)*   - *Pre:* NR - *Post:* NR |
| Le Foll-de Moro et al. (2005)  France  Pre-post | | | | *N =* 6 (5M/1F)    *Age =* 29 ± 14 years  *TSI =* 14 ± 0.3 years (6 acute/0 chronic)  *Classification =* 0 T/6 P  *Severity =* NR  *CPET =* Maximal wheelchair test  *CPET same modality of intervention? =* Yes  *Pre-registered? =* No  *Protocol manuscript? =* No | *Type of Exercise =* Wheelchair exercise  *Relative Intensity and Class =* Moderate-to-vigorous (Up to 80% HR_max_)  *Session Duration (min) =* 30  *Frequency (times/week) =* 3  *Intervention Length (weeks) =* 6  *Adverse Events =* NR | *AV̇O_2peak_ (L/min)*   - *Pre:* 1.21 ± 0.3 - *Post:* 1.64 ± 0.3   *RV̇O_2peak_ (mL/kg/min)*   - *Pre:* NR - *Post:* NR   *PPO (W)*   - *Pre:* 49 ± 4 - *Post:* 65 ± 9 |
| Lindberg et al. (2012)  Sweden  Pre-post | | | | *N =* 13 (8M/5F)    *Age =* 47 ± 12 years  *TSI =* 20 ± 10 years (0 acute/13 chronic)  *Classification =* 0 T/13 P  *Severity =* 10 comp./3 incomp.  *CPET =* Maximal double poling exercises on ergometer  *CPET same modality of intervention? =* Yes  *Pre-registered? =* No  *Protocol manuscript? =* No | *Type of Exercise =* Interval training on seated double-poling ergometer  *Relative Intensity and Class =* Moderate-to-vigorous (70-100% HR_peak_)  *Session Duration (min) =* 28  *Frequency (times/week) =* 3  *Intervention Length (weeks) =* 10  *Adverse Events =* NR | *AV̇O_2peak_ (L/min)*   - *Pre:* 1.27 ± 0.39 - *Post:* 1.56 ± 0.48   *RV̇O_2peak_ (mL/kg/min)*   - *Pre:* 18.5 ± 4.8 - *Post:* 23.0 ± 6.3   *PPO (W)*   - *Pre:* NR - *Post:* NR |
| Lotter et al. (2020)  USA  Pre-post | | IMPAIRMENT | | *N =* 8 (6M/2F)    *Age =* 51 ± 17 years  *TSI =* 4 ± 2 years (0 acute/8 chronic)  *Classification =* 6 T/2 P  *Severity =* 0 comp./8 incomp.  *CPET =* Treadmill graded exercise test  *CPET same modality of intervention? =* No  *Pre-registered? =* Yes (NCT02115685)  *Protocol manuscript? =* No | *Type of Exercise =* Impairment-based training including rehabilitation strategies directed toward specific impairments underlying walking dysfunction with strength and aerobic exercises  *Relative Intensity and Class =* Vigorous (70-80% HRR and RPE >14)  *Session Duration (min) =* 40  *Frequency (times/week) =* 3.3  *Intervention Length (weeks) =* 6  *Adverse Events =* No serious events but minor events included 11 falls, 10 incidents of soreness/ankle foot orthoses-related abrasions, one incident of nausea, hypertension, and anxiety | *AV̇O_2peak_ (L/min)*   - *Pre:* NR - *Post:* NR   *RV̇O_2peak_ (mL/kg/min)*   - *Pre:* 17.0 ± 7.8 - *Post:* 18.0 ± 9.4   *PPO (W)*   - *Pre:* NR - *Post:* NR |
|  |  | TASK | | *N =* 8 (4M/4F)    *Age =* 46 ± 13 years  *TSI =* 13 ± 4 years (0 acute/8 chronic)  *Classification =* 4 T/4 P  *Severity =* 0 comp./8 incomp.  *CPET =* Treadmill graded exercise test  *CPET same modality of intervention? =* No  *Pre-registered? =* Yes (NCT02115685)  *Protocol manuscript? =* No | *Type of Exercise =* Task specific training including 40 minutes of stepping practice including speed-dependent treadmill training, over-ground training, skill-dependent treadmill training, and stair climbing  *Relative Intensity and Class =* Vigorous (70-80% HRR and RPE >14)  *Session Duration (min) =* 40  *Frequency (times/week) =* 3.3  *Intervention Length (weeks) =* 6  *Adverse Events =* No serious events but minor events included 11 falls, 10 incidents of soreness/ankle foot orthoses-related abrasions, one incident of nausea, hypertension, and anxiety | *AV̇O_2peak_ (L/min)*   - *Pre:* NR - *Post:* NR   *RV̇O_2peak_ (mL/kg/min)*   - *Pre:* 17.0 ± 5.2 - *Post:* 19.0 ± 8.2   *PPO (W)*   - *Pre:* NR - *Post:* NR |
| Ma et al. (2019)  Canada  RCT behaviour change | | | | *N =* 14 (9M/5F)    *Age =* 45.79 ± 13.63 years  *TSI =* 14.71 ± 13.63 years (0 acute/ 14 chronic)  *Classification =* 5 T/ 9 P  *Severity =* 8 comp./ 6 incomp.  *CPET =* Graded ACE exercise test  *CPET same modality of intervention? =* No  *Pre-registered? =* Yes (NCT03111030)  *Protocol manuscript? =* No | *Type of Behaviour Change Counselling =* Behavioural physical activity coaching  *Number of Contact Sessions =* 9 (1-h introductory session followed by eight once-weekly 10- to 15-min behavioural physical activity coaching sessions)  *Intervention Length (weeks) =* 8  *Adverse Events =* NR | *AV̇O_2peak_ (L/min)*   - *Pre:* 1.16 ± 0.38 - *Post:* 1.30 ± 0.43   *RV̇O_2peak_ (mL/kg/min)*   - *Pre:* 15.9 ± 4.2 - *Post:* 17.8 ± 4.9   *PPO (W)*   - *Pre:* 82 ± 27 - *Post:* 87 ± 30 |
| Mcleod et al. (2020)  Canada  RCT intensity comparison | | MODERATE | | *N =* 10 (5M/5F)    *Age =* 45 ± 17 years  *TSI =* 0.15 ± 0.12 years (10 acute/ 0 chronic)  *Classification =* 5 T/ 5 P  *Severity =* 1 comp./ 9 incomp.  *CPET =* ACE  *CPET same modality of intervention? =* Yes  *Pre-registered? =* Yes (NCT03709095)  *Protocol manuscript? =* No | *Type of Exercise =* ACE  *Relative Intensity =* Moderate [12 RPE (45% PPO)]  *Session Duration (min) =* 20  *Frequency (times/week) =* 3  *Intervention Length (weeks) =* 5  *Adverse Events =* NR | *AV̇O_2peak_ (L/min)*   - *Pre:* NR - *Post:* NR   *RV̇O_2peak_ (mL/kg/min)*   - *Pre:* NR - *Post:* NR   *PPO (W)*   - *Pre:* 45 ± 20 - *Post:* 58 ± 21 |
|  |  | VIGOROUS | | *N =* 10 (10M/0F)    *Age =* 47 ± 15 years  *TSI =* 0.20 ± 0.19 years (10 acute/ 0 chronic)  *Classification =* 4 T/ 6 P  *Severity =* 1 comp./ 9 incomp.  *CPET =* ACE  *CPET same modality of intervention? =* Yes  *Pre-registered? =* Yes (NCT03709095)  *Protocol manuscript? =* No | *Type of Exercise =* ACE  *Relative Intensity =* Supramaximal (‘All out efforts’ and 10% PPO active recovery)  *Session Duration (min) =* 5 (3 x 20-s supramaximal bouts interspersed by 120-s active recovery)  *Frequency (times/week) =* 3  *Intervention Length (weeks) =* 5  *Adverse Events =* 1 individual experienced post-exercise hypotension | *AV̇O_2peak_ (L/min)*   - *Pre:* NR - *Post:* NR   *RV̇O_2peak_ (mL/kg/min)*   - *Pre:* NR - *Post:* NR   *PPO (W)*   - *Pre:* 52 ± 29 - *Post:* 69 ± 37 |
| Midha et al. (1999)  USA  Pre-post | | | | *N =* 12 (11M/1F)    *Age =* 38 ± 11 years  *TSI =* 15 ± 8 years (0 acute/10 chronic)  *Classification =* 3 T/7 P  *Severity =* 3 comp./7 incomp.  *CPET =* Wheelchair aerobic fitness trainer  *CPET same modality of intervention? =* Yes  *Pre-registered? =* No  *Protocol manuscript? =* No | *Type of Exercise =* Wheelchair aerobic fitness trainer  *Relative Intensity and Class =* Moderate-to-vigorous (55-90% age-predicted HR_max_)  *Session Duration (min) =* 22.5  *Frequency (times/week) =* 2.5  *Intervention Length (weeks) =* 10  *Adverse Events =* NR | *AV̇O_2peak_ (L/min)*   - *Pre:* 1.4 ± 0.4 - *Post:* 1.8 ± 0.5   *RV̇O_2peak_ (mL/kg/min)*   - *Pre:* 19 ± 6 - *Post:* 24 ± 6   *PPO (W)*   - *Pre:* NR - *Post:* NR |
| Milia et al. (2014)  Italy  Pre-post | | | | *N =* 9 (7M/2F)    *Age =* 41 ± 11 years  *TSI =* NR (0 acute/9 chronic)  *Classification =* 0 T/9 P  *Severity =* 9 comp./0 incomp.  *CPET =* Electromagnetically braked ACE  *CPET same modality of intervention? =* Yes  *Pre-registered? =* No  *Protocol manuscript? =* No | *Type of Exercise =* ACE  *Relative Intensity and Class =* Moderate (60% of W_max_)  *Session Duration (min) =* 60  *Frequency (times/week) =* 4  *Intervention Length (weeks) =* 52  *Adverse Events =* NR | *AV̇O_2peak_ (L/min)*   - *Pre:* 1.38 ± 0.18 - *Post:* 1.57 ± 0.32   *RV̇O_2peak_ (mL/kg/min)*   - *Pre:* 20.1 ± 3.1 - *Post:* 22.6 ± 2.7   *PPO (W)*   - *Pre:* 97.1 ± 8.8 - *Post:* 111 ± 7.8 |
| Mohr et al. (1997)  Denmark  Pre-post | | | | *N =* 10 (8M/2F)    *Age =* 35 ± 2 years  *TSI =* 13 ± 3 years (0 acute/10 chronic)  *Classification =* 6 T/4 P  *Severity =* 9 comp./1 incomp.  *CPET =* Progressive exercise test by adding load to ergometer  *CPET same modality of intervention? =* Yes  *Pre-registered? =* No  *Protocol manuscript? =* No | *Type of Exercise =* FES  *Relative Intensity and Class =* Vigorous [Highest possible resistance compatible with pedalling frequency of 50 rpm (up to 7/8kp)]  *Session Duration (min) =* 30  *Frequency (times/week) =* 3  *Intervention Length (weeks) =* 46  *Adverse Events =* Three subjects experienced post-exercise hypotension, one experienced a small hematoma in the medial portion of the quadriceps due to a fibrillar tear | *AV̇O_2peak_ (L/min)*   - *Pre:* 1.20 ± 0.30 - *Post:* 1.43 ± 0.20   *RV̇O_2peak_ (mL/kg/min)*   - *Pre:* NR - *Post:* NR   *PPO (W)*   - *Pre:* NR - *Post:* NR |
| Mutton et al. (1997)  USA  Pre-post | | PHASE I & II | | *N =* 11 (11M/0F)    *Age =* 36 ± 7 years  *TSI =* 10 ± 4 years (0 acute/11 chronic)  *Classification =* 2 T/9 P  *Severity =* 11 comp./0 incomp.  *CPET =* FES leg cycling  *CPET same modality of intervention? =* Yes  *Pre-registered? =* No  *Protocol manuscript? =* No | *Type of Exercise =* FES leg cycle ergometry and hybrid exercise  *Relative Intensity and Class =* Mixed/cannot determine  *Session Duration (min) =* 30  *Frequency (times/week) =* 2  *Intervention Length (weeks) =* 18  *Adverse Events =* NR | *AV̇O_2peak_ (L/min)*   - *Pre:* 1.30 ± 0.27 - *Post:* 1.42 ± 0.3   *RV̇O_2peak_ (mL/kg/min)*   - *Pre:* NR - *Post:* NR   *PPO (W)*   - *Pre:* 10.5 ± 4.8 - *Post:* 14.4 ± 4.9 |
|  |  | PHASE III | | *N =* 8 (8M/0F)    *Age =* 36 ± 8 years  *TSI =* 9 ± 4 years (0 acute/8 chronic)  *Classification =* 2 T/6 P  *Severity =* 8 comp./0 incomp.  *CPET =* Hybrid cycling  *CPET same modality of intervention? =* Yes  *Pre-registered? =* No  *Protocol manuscript? =* No | *Type of Exercise =* Hybrid exercise with FES leg cycle ergometry and ACE  *Relative Intensity and Class =* Mixed/cannot determine  *Session Duration (min) =* 30  *Frequency (times/week) =* 2  *Intervention Length (weeks) =* 24  *Adverse Events =* NR | *AV̇O_2peak_ (L/min)*   - *Pre:* 1.69 ± 0.64 - *Post:* 1.91 ± 0.49   *RV̇O_2peak_ (mL/kg/min)*   - *Pre:* NR - *Post:* NR   *PPO (W)*   - *Pre:* 36.6 ± 23.4 - *Post:* 45.4 ± 24.2 |
| Nash et al. (2001)  USA  Pre-post | | | | *N =* 5 (5M/0F)    *Age =* 38 ± 4 years  *TSI =* 5 ± 1 years (0 acute/5 chronic)  *Classification =* 0 T/5 P  *Severity =* 5 comp./0 incomp.  *CPET =* Hydraulically braked ACE  *CPET same modality of intervention? =* Yes  *Pre-registered? =* No  *Protocol manuscript? =* No | *Type of Exercise =* Circuit resistance exercise training  *Relative Intensity and Class =* Moderate (50-60% 1RM)  *Session Duration (min) =* 45  *Frequency (times/week) =* 3  *Intervention Length (weeks) =* 12  *Adverse Events =* NR | *AV̇O_2peak_ (L/min)*   - *Pre:* 1.3 ± 0.40 - *Post:* 1.68 ± 0.30   *RV̇O_2peak_ (mL/kg/min)*   - *Pre:* NR - *Post:* NR   *PPO (W)*   - *Pre:* 85 ± 16 - *Post:* 111 ± 17 |
| Nightingale et al. (2018)  UK  RCT | | | | *N =* 15 (11M/4F)    *Age =* 46.7 ± 5.6 years  *TSI =* NR (0 acute/ 15 chronic)  *Classification =* 0 T/ 15 P  *Severity =* NR  *CPET =* Discontinuous incremental submaximal ACE  *CPET same modality of intervention? =* Yes  *Pre-registered? =* Yes (ISRCTN57096451)  *Protocol manuscript? =* Yes | *Type of Exercise =* ACE  *Relative Intensity and Class =* Moderate (60-65% V̇O_2peak_ )  *Session Duration (min) =* 45  *Frequency (times/week) =* 4  *Intervention Length (weeks) =* 6  *Adverse Events =* One participant did not complete the trial due to illness, not related to intervention. | *AV̇O_2peak_ (L/min)*   - *Pre:* 1.49 ± 0.50 - *Post:* 1.69 ± 0.54   *RV̇O_2peak_ (mL/kg/min)*   - *Pre:* 17.78 ± 4.89 - *Post:* 20.73 ± 5.53   *PPO (W)*   - *Pre:* 77 ± 27 - *Post:* 93 ± 32 |
| Nooijen et al. (2015)  The Netherlands  Pre-post | | | | *N =* 45 (39M/6F)    *Age =* 43 ± 19 years  *TSI =* 0.4 ± 0.2 years (45 acute/0 chronic)  *Classification =* 15 T/30 P  *Severity =* 29 comp./16 incomp.  *CPET =* Maximal hand cycling  *CPET same modality of intervention? =* Yes  *Pre-registered? =* Yes (NTR2424)  *Protocol manuscript? =* No | *Type of Exercise =* Handcycle interval training program  *Relative Intensity and Class =* Moderate-to-vigorous (4-7 on Borg CR10 scale)  *Session Duration (min) =* 60  *Frequency (times/week) =* 3  *Intervention Length (weeks) =* 8  *Adverse Events =* Two participants did not complete training due to severe pressure ulcers | *AV̇O_2peak_ (L/min)*   - *Pre:* 1.1 ± 0.4 - *Post:* 1.3 ± 0.5   *RV̇O_2peak_ (mL/kg/min)*   - *Pre:* 17.0 ± 7.5 - *Post:* 19.4 ± 8.8   *PPO (W)*   - *Pre:* 40.7 ± 23.9 - *Post:* 55.7 ± 29.5 |
| Nooijen et al. (2017)  The Netherlands  RCT behaviour change | | | | *N =* 20 (17M/3F)    *Age =* 44 ± 15 years  *TSI =* 0.38 ± 0.18 years (20 acute/ 0 chronic)  *Classification =* 7 T/ 13 P  *Severity =* 13 comp./7 incomp.  *CPET =* Graded maximal handcycle test  *CPET same modality of intervention? =* No  *Pre-registered? =* Yes (NTR2424)  *Protocol manuscript? =* No | *Type of Behaviour Change Counselling =* Face-to-face sessions  *Number of Contact Sessions =* 13  *Intervention Length (weeks) =* 52  *Adverse Events =* NR | *AV̇O_2peak_ (L/min)*   - *Pre:* 1.20 ± 0.43 - *Post:* 1.52 ± 0.60   *RV̇O_2peak_ (mL/kg/min)*   - *Pre:* NR - *Post:* NR   *PPO (W)*   - *Pre:* 44 ± 23 - *Post:* 80 ± 49 |
| Ozturk et al. (2021)  USA  Pre-post | | PARA | | *N =* 8 (8M/0F)    *Age =* 30 ± 6 years  *TSI =* 2 ± 2 years (NR)  *Classification =* NR  *Severity =* 8 comp./0 incomp.  *CPET =* FES-assisted exercise tolerance test  *CPET same modality of intervention? =* Yes  *Pre-registered? =* No  *Protocol manuscript? =* No | *Type of Exercise =* FES-assisted full body rowing  *Relative Intensity and Class =* Mixed/cannot determine  *Session Duration (min) =* 45  *Frequency (times/week) =* 2.5  *Intervention Length (weeks) =* 24  *Adverse Events =* NR | *AV̇O_2peak_ (L/min)*   - *Pre:* 1.73 ± 0.27 - *Post:* 1.91 ± 0.48   *RV̇O_2peak_ (mL/kg/min)*   - *Pre:* 22.8 ± 2.5 - *Post:* 26.9 ± 2.2   *PPO (W)*   - *Pre:* NR - *Post:* NR |
|  |  | MIXED | | *N =* 16 (15M/1F)    *Age =* 28 ± 7 years  *TSI =* 2 ± 1 years (NR)  *Classification =* NR  *Severity =* 13 comp./3 incomp.  *CPET =* FES-assisted exercise tolerance test  *CPET same modality of intervention? =* Yes  *Pre-registered? =* No  *Protocol manuscript? =* No | *Type of Exercise =* FES-assisted full body rowing  *Relative Intensity and Class =* Mixed/cannot determine  *Session Duration (min) =* 45  *Frequency (times/week) =* 2.5  *Intervention Length (weeks) =* 24  *Adverse Events =* NR | *AV̇O_2peak_ (L/min)*   - *Pre:* 1.39 ± 0.35 - *Post:* 1.52 ± 0.41   *RV̇O_2peak_ (mL/kg/min)*   - *Pre:* 17.7 ± 5.2 - *Post:* 19.2 ± 5.7   *PPO (W)*   - *Pre:* NR - *Post:* NR |
| Pelletier et al. (2015)  Canada  RCT | | | | *N =* 12 (12M/0F)    *Age =* 40 ± 12.3 years  *TSI =* 15 ± 8.52 years (0 acute/ 12 chronic)  *Classification =* NR  *Severity =* 4 comp./ 8 incomp.  *CPET =* Symptom-limited graded ACE exercise test  *CPET same modality of intervention? =* Yes  *Pre-registered? =* No  *Protocol manuscript? =* No | *Type of Exercise =* Mixed: aerobic ACE and RT  *Relative Intensity and Class =* Moderate-to-vigorous (3-6 RPE; 50-70% 1RM RT)  *Session Duration (min) =* 20  *Frequency (times/week) =* 2  *Intervention Length (weeks) =* 16  *Adverse Events =* NR | *AV̇O_2peak_ (L/min)*   - *Pre:* 1.42 ± 0.48 - *Post:* 1.56 ± 0.48   *RV̇O_2peak_ (mL/kg/min)*   - *Pre:* 16.3 ± 5.5 - *Post:* 19.1 ± 6.7   *PPO (W)*   - *Pre:* 67 ± 29 - *Post:* 76 ± 34 |
| Piira et al. (2019)  Norway  RCT | | | | *N =* 10 (6M/4F)    *Age =* 46 ± 14 years  *TSI =* 11.3 ± 9 years (0 acute/ 10 chronic)  *Classification =* 3 T/ 7 P  *Severity =* 0 comp./ 10 incomp.  *CPET =* Progressive ACE  *CPET same modality of intervention? =* No  *Pre-registered? =* No  *Protocol manuscript? =* No | *Type of Exercise =* Gait training (body-weight supported locomotor training)  *Relative Intensity and Class =* Mixed/cannot determine (“increase towards 3-5 km/h”)  *Session Duration (min) =* 90  *Frequency (times/week) =* 5  *Intervention Length (weeks) =* 12  *Adverse Events =* NR | *AV̇O_2peak_ (L/min)*   - *Pre:* 1.40 ± 0.50 - *Change:* -0.10 ± 0.20   *RV̇O_2peak_ (mL/kg/min)*   - *Pre:* NR - *Post:* NR   *PPO (W)*   - *Pre:* NR - *Post:* NR |
| Pollack et al. (1989)  USA  Pre-post | | | | *N =* 11 (7M/4F)    *Age =* 29 ± 9 years  *TSI =* 6 ± 3 years (1 acute/10 chronic)  *Classification =* 7 T/4 P  *Severity =* 11 comp./0 incomp.  *CPET =* Incremental pedalling on ergometer  *CPET same modality of intervention? =* Yes  *Pre-registered? =* No  *Protocol manuscript? =* No | *Type of Exercise =* FES cycle ergometry  *Relative Intensity and Class =* Mixed/cannot determine  *Session Duration (min) =* NR  *Frequency (times/week) =* 3  *Intervention Length (weeks) =* 20.5  *Adverse Events =* NR | *AV̇O_2peak_ (L/min)*   - *Pre:* 0.5 ± 0.1 - *Post:* 1.0 ± 0.1   *RV̇O_2peak_ (mL/kg/min)*   - *Pre:* NR - *Post:* NR   *PPO (W)*   - *Pre:* NR - *Post:* NR |
| Qiu et al. (2016)  USA  Pre-post | | | | *N =* 12 (11M/1F)    *Age =* 33 ± 4 years  *TSI =* 8 ± 3 years (NR)  *Classification =* NR  *Severity =* NR  *CPET =* FES rowing  *CPET same modality of intervention? =* Yes  *Pre-registered? =* No  *Protocol manuscript? =* No | *Type of Exercise =* Hybrid FES rowing  *Relative Intensity and Class =* Vigorous (75-85% HR_max_)  *Session Duration (min) =* 30  *Frequency (times/week) =* 3  *Intervention Length (weeks) =* 24  *Adverse Events =* NR | *AV̇O_2peak_ (L/min)*   - *Pre:* 1.1 ± 0.1 - *Post:* 1.3 ± 0.1   *RV̇O_2peak_ (mL/kg/min)*   - *Pre:* 15.3 ± 1.5 - *Post:* 17.1 ± 1.6   *PPO (W)*   - *Pre:* 34.6 ± 4.4 - *Post:* 44.4 ± 5.7 |
| Rosety-Rodriguez et al. (2014)  Spain  RCT | | | | *N =* 9 (9M/0F)    *Age =* 29.6 ± 3.6 years  *TSI =* 4.6 ± 0.3 years (0 acute/ 9 chronic)  *Classification =* 0 T/ 9 P  *Severity =* 9 comp./ 0 incomp.  *CPET =* Continuous incremental workload test on an ACE  *CPET same modality of intervention? =* Yes  *Pre-registered? =* No  *Protocol manuscript? =* No | *Type of Exercise =* Upper-body, aerobic ACE  *Relative Intensity and Class =* Moderate-to-vigorous (50-65% HRR)  *Session Duration (min) =* 20-30 (increase of 2.5 min every 3 weeks)  *Frequency (times/week) =* 3  *Intervention Length (weeks) =* 12  *Adverse Events =* NR | *AV̇O_2peak_ (L/min)*   - *Pre:* NR - *Post:* NR   *RV̇O_2peak_ (mL/kg/min)*   - *Pre:* 23.2 ± 2.1 - *Post:* 25.6 ± 1.9   *PPO (W)*   - *Pre:* NR - *Post:* NR |
| Schleifer et al. (2022)  USA  Pre-post | | | | *N =* 47 (44M/3F)    *Age =* 33.4 ± 11.6 years  *TSI =* 5.63 ± 9.75 years (NR)  *Classification =* 22 T/25 P  *Severity =* 38 comp./7 incomp.  *CPET =* Hybrid FES rowing  *CPET same modality of intervention? =* Yes  *Pre-registered? =* Yes (NCT02139436)  *Protocol manuscript? =* No | *Type of Exercise =* FES with volitional arm pull  *Relative Intensity and Class =* Moderate-to-vigorous (60-85% HRR)  *Session Duration (min) =* 60  *Frequency (times/week) =* 3  *Intervention Length (weeks) =* 24  *Adverse Events =* NR | *AV̇O_2peak_ (L/min)*   - *Pre:* 1.57 ± 0.52 - *Post:* 1.71 ± 0.61   *RV̇O_2peak_ (mL/kg/min)*   - *Pre:* 19.5 ± 6.0 - *Post:* 21.1 ± 7.1   *PPO (W)*   - *Pre:* NR - *Post:* NR |
| Sutbeyaz et al. (2005)  Turkey  Pre-post | | | | *N =* 20 (12M/8F)    *Age =* 31 ± 8 years  *TSI =* 0.3 ± 0.4 years (NR)  *Classification =* 0 T/20 P  *Severity =* 14 comp./6 incomp.  *CPET =* ACE  *CPET same modality of intervention? =* Yes  *Pre-registered? =* No  *Protocol manuscript? =* No | *Type of Exercise =* ACE  *Relative Intensity and Class =* Vigorous (75% of baseline V̇O_2peak_ and gradually increased to maximal exercise)  *Session Duration (min) =* 60  *Frequency (times/week) =* 3  *Intervention Length (weeks) =* 6  *Adverse Events =* NR | *AV̇O_2peak_ (L/min)*   - *Pre:* NR - *Post:* NR   *RV̇O_2peak_ (mL/kg/min)*   - *Pre:* 9.9 ± 4.2 - *Post:* 14.6 ± 4.0   *PPO (W)*   - *Pre:* 31.2 ± 12.8 - *Post:* 51.6 ± 16.6 |
| Taylor et al. (1986)  Canada  RCT | | | | *N =* 5 (5M/0F)    *Age =* 27 ± 3 years  *TSI =* 7.2 ± 4.4 years (0 acute/ 5 chronic)  *Classification =* 0 T/ 5 P  *Severity =* NR  *CPET =* Maximal ACE exercise test  *CPET same modality of intervention? =* Yes  *Pre-registered? =* No  *Protocol manuscript? =* No | *Type of Exercise =* Upper-body aerobic ACE  *Relative Intensity and Class =* Vigorous (80% HR_peak_)  *Session Duration (min) =* 30  *Frequency (times/week) =* 5  *Intervention Length (weeks) =* 8  *Adverse Events =* NR | *AV̇O_2peak_ (L/min)*   - *Pre:* 1.9 ± 0.6 - *Post:* 2.1 ± 0.4   *RV̇O_2peak_ (mL/kg/min)*   - *Pre:* 22.8 ± 4.6 - *Post:* 26.3 ± 3.6   *PPO (W)*   - *Pre:* NR - *Post:* NR |
| Taylor et al. (2014)  USA  Pre-post | | | | *N =* 14 (13M/1F)    *Age =* 39 ± 3 years  *TSI =* 10 ± 3 years (0 acute/14 chronic)  *Classification =* 0 T/14 P  *Severity =* 14 comp./0 incomp.  *CPET =* Graded FES rowing  *CPET same modality of intervention? =* Yes  *Pre-registered? =* No  *Protocol manuscript? =* No | *Type of Exercise =* Hybrid FES rowing  *Relative Intensity and Class =* Vigorous (75-85% HR_max_)  *Session Duration (min) =* 30  *Frequency (times/week) =* 3  *Intervention Length (weeks) =* 24  *Adverse Events =* NR | *AV̇O_2peak_ (L/min)*   - *Pre:* NR - *Post:* NR   *RV̇O_2peak_ (mL/kg/min)*   - *Pre:* 19.6 ± 6.0 - *Post:* 21.4 ± 6.6   *PPO (W)*   - *Pre:* 54 ± 20 - *Post:* 60 ± 20 |
| Tordi et al. (2001)  France  Pre-post | | | | *N =* 5 (5M/0F)    *Age =* 27 ± 8 years  *TSI =* >2 years (0 acute/5 chronic)  *Classification =* 0 T/5 P  *Severity =* 5 comp./0 incomp.  *CPET =* Wheelchair test  *CPET same modality of intervention? =* Yes  *Pre-registered? =* No  *Protocol manuscript? =* No | *Type of Exercise =* Wheelchair ergometry  *Relative Intensity and Class =* Moderate-to-vigorous (50-80% maximal tolerated power)  *Session Duration (min) =* 30  *Frequency (times/week) =* 3  *Intervention Length (weeks) =* 4  *Adverse Events =* NR | *AV̇O_2peak_ (L/min)*   - *Pre:* NR - *Post:* NR   *RV̇O_2peak_ (mL/kg/min)*   - *Pre:* 23 ± 4.6 - *Post:* 26.5 ± 4.1   *PPO (W)*   - *Pre:* 42.5 ± 2.8 - *Post:* 55 ± 5.8 |
| Torhaug et al. (2016)  Norway  RCT | | | | *N =* 9 (9M/0F)    *Age =* 42 ± 12.4 years  *TSI =* 13.2 ± 7.2 years (0 acute/ 9 chronic)  *Classification =* 0 T/ 9 P  *Severity =* 6 comp./ 3 incomp.  *CPET =* Maximal wheelchair ergometry test  *CPET same modality of intervention? =* No  *Pre-registered? =* No  *Protocol manuscript? =* No | *Type of Exercise =* RT (bench press)  *Relative Intensity and Class =* Vigorous (85-95% 1RM)  *Session Duration (min) =* 60  *Frequency (times/week) =* 3  *Intervention Length (weeks) =* 6  *Adverse Events =* NR | *AV̇O_2peak_ (L/min)*   - *Pre:* 2.15 ± 0.47 - *Post:* 2.06 ± 0.61   *RV̇O_2peak_ (mL/kg/min)*   - *Pre:* 28.5 ± 6.5 - *Post:* 28.5 ± 7.0   *PPO (W)*   - *Pre:* NR - *Post:* NR |
| van der Scheer et al. (2016)  The Netherlands  RCT | | | | *N =* 14 (12M/2F)    *Age =* 53.6 ± 16.3 years  *TSI =* 19.3 ± 11.9 years (0 acute/ 14 chronic)  *Classification =* 5 T/ 9 P  *Severity =* 10 comp./ 4 incomp.  *CPET =* Peak incremental treadmill test  *CPET same modality of intervention? =* Yes  *Pre-registered? =* Yes (NTR3037)  *Protocol manuscript? =* Yes | *Type of Exercise =* Wheelchair propulsion*  *Relative Intensity and Class =* Light (1-3 RPE; 30-40% HRR)  *Session Duration (min) =* 30  *Frequency (times/week) =* 2  *Intervention Length (weeks) =* 16  *Adverse Events =* NR  **Exercise was continuous in participants with the highest baseline fitness levels, while those with lower levels followed protocols with intermittent exercise (4 × 7.5 or 10 × 3 min with 1–2 min rest intervals).* | *AV̇O_2peak_ (L/min)*   - *Pre:* 1.11 ± 0.60 - *Post:* 1.06 ± 0.58   *RV̇O_2peak_ (mL/kg/min)*   - *Pre:* NR - *Post:* NR   *PPO (W)*   - *Pre:* 35 ± 25 - *Post:* 36 ± 27 |
| Vestergaard et al. (2022)  Denmark  Pre-post | | | | *N =* 7 (6M/1F)    *Age =* 44.0 ± 15.9 years  *TSI =* 14.4 ± 16.9 years (0 acute/ 7 chronic)  *Classification =* 0 T/ 7 P  *Severity =* 3 comp./ 4 incomp.  *CPET =* Ski ergometry maximal exercise test  *CPET same modality of intervention? =* Yes  *Pre-registered? =* Yes (NCT04211311)  *Protocol manuscript? =* No | *Type of Exercise =* FES-assisted cycling with volitional arm ski ergometry  *Relative Intensity and Class =* Moderate-to-vigorous (90% PPO intervals with 2-min active rest; yet majority of participants could not reach this intensity, thus the intensity was categorised as moderate-to-vigorous)  *Session Duration (min) =* 22 (4x4 min intervals, interspersed with 2-min active rest periods)  *Frequency (times/week) =* 3  *Intervention Length (weeks) =* 8  *Adverse Events =* Slight non-persisting pain in neck (n = 1), arms and shoulders (n = 4) during and between training sessions, dizziness that disappeared after 5 min (n = 1), feeling tired in the head/dizziness that disappeared after training with no other signs of autonomic hyperreflexia (n = 2), increased spasms (n = 2), and vomiting just after training (n = 2). | *AV̇O_2peak_ (L/min)*   - *Pre:* 1.64 ± 0.39 - *Post:* 1.91 ± 0.61   *RV̇O_2peak_ (mL/kg/min)*   - *Pre:* 18.3 ± 2.8 - *Post:* 21.2 ± 5.6   *PPO (W)*   - *Pre:* 55 ± 22 - *Post:* 76 ± 17 |
| Wheeler et al. (2002)  Canada  Pre-post | | | | *N =* 6 (NR)    *Age =* 43 ± 18 years  *TSI =* 14 ± 12 years (0 acute/6 chronic)  *Classification =* 1 T/5 P  *Severity =* 5 comp./1 incomp.  *CPET =* Hybrid (FES-row)  *CPET same modality of intervention? =* Yes  *Pre-registered? =* No  *Protocol manuscript? =* No | *Type of Exercise =* FES-assisted rowing  *Relative Intensity and Class =* Vigorous (70-75% V̇O_2peak_)  *Session Duration (min) =* 30  *Frequency (times/week) =* 3  *Intervention Length (weeks) =* 12  *Adverse Events =* NR | *AV̇O_2peak_ (L/min)*   - *Pre:* 1.81 ± 0.41 - *Post:* 2.01 ± 0.38   *RV̇O_2peak_ (mL/kg/min)*   - *Pre:* NR - *Post:* NR   *PPO (W)*   - *Pre:* NR - *Post:* NR |
| Wilbanks et al. (2016)  USA  Pre-post | | | | *N =* 10 (8M/2F)    *Age =* 47 ± 18 years  *TSI =* 18 ± 14 years (0 acute/10 chronic)  *Classification =* 0 T/10 P  *Severity =* 7 comp./3 incomp.  *CPET =* FES-row training  *CPET same modality of intervention? =* Yes  *Pre-registered? =* No  *Protocol manuscript? =* No | *Type of Exercise =* FES-assisted rowing  *Relative Intensity and Class =* Mixed/cannot determine (Self-selected stroke rate but instructed to exceed power output from previous session)  *Session Duration (min) =* 30  *Frequency (times/week) =* 3  *Intervention Length (weeks) =* 6  *Adverse Events =* NR | *AV̇O_2peak_ (L/min)*   - *Pre:* NR - *Post:* NR   *RV̇O_2peak_ (mL/kg/min)*   - *Pre:* 18.5 ± 6.4 - *Post:* 20 ± 6.7   *PPO (W)*   - *Pre:* 36.1 ± 19.0 - *Post:* 42.8 ± 24.7 |
| Williams et al. (2020)  Canada  Pre-post | | | | *N =* 14 (8M/6F)    *Age =* 44 ± 10 years  *TSI =* 22 ± 13 years (0 acute/14 chronic)  *Classification =* 5 T/9 P  *Severity =* 8 comp./3 incomp.  *CPET =* ACE  *CPET same modality of intervention? =* Yes  *Pre-registered? =* No  *Protocol manuscript? =* No | *Type of Exercise =* ACE  *Relative Intensity and Class =* Moderate (Borg CR10)  *Session Duration (min) =* 40  *Frequency (times/week) =* 3  *Intervention Length (weeks) =* 5  *Adverse Events =* NR | *AV̇O_2peak_ (L/min)*   - *Pre:* NR - *Post:* NR   *RV̇O_2peak_ (mL/kg/min)*   - *Pre:* 17.1 ± 4.9 - *Post:* 19.8 ± 4.8   *PPO (W)*   - *Pre:* 84.8 ± 38.9 - *Post:* 96.8 ± 41.4 |
| Wouda et al. (2018)  Norway  RCT intensity comparison | | MODERATE | | *N =* 10 (8M/2F)    *Age =* 34 ± 15 years  *TSI =* 0.18 ± 0.09 years (10 acute/ 0 chronic)  *Classification =* 4 T/ 6 P  *Severity =* 0 comp./ 10 incomp.  *CPET =* Maximal graded treadmill test  *CPET same modality of intervention? =* Yes  *Pre-registered? =* Yes (NCT01903226)  *Protocol manuscript? =* No | *Type of Exercise =* Gait training (walking or running)  *Relative Intensity =* Moderate (70% HR_peak_)  *Session Duration (min) =* 45  *Frequency (times/week) =* 3  *Intervention Length (weeks) =* 12  *Adverse Events =* NR | *AV̇O_2peak_ (L/min)*   - *Pre:* 2.79 ± 0.79 - *Post:* 3.23 ± 0.94   *RV̇O_2peak_ (mL/kg/min)*   - *Pre:* 36.9 ± 11.8 - *Post:* 42.3 ± 12.0   *PPO (W)*   - *Pre:* NR - *Post:* NR |
|  |  | VIGOROUS | | *N =* 10 (8M/2F)    *Age =* 50 ± 15 years  *TSI =* 0.19 ± 0.08 years (10 acute/ 0 chronic)  *Classification =* 7 T/ 3 P  *Severity =* 0 comp./ 10 incomp.  *CPET =* Maximal graded treadmill test  *CPET same modality of intervention? =* Yes  *Pre-registered? =* Yes (NCT01903226)  *Protocol manuscript? =* No | *Type of Exercise =* Gait training (walking or running) HIIT session  *Relative Intensity =* Vigorous (85-95% HR_peak_ and 70% HR_peak_)  *Session Duration (min) =* 25 (4 x 4-min intervals at 85-95% HR_peak_ interspersed with 3 x 3-min recovery at 70% HR_peak_)  *Frequency (times/week) =* 2  *Intervention Length (weeks) =* 12  *Adverse Events =* NR | *AV̇O_2peak_ (L/min)*   - *Pre:* 2.70 ± 0.81 - *Post:* 3.00 ± 0.62   *RV̇O_2peak_ (mL/kg/min)*   - *Pre:* 32.1 ± 9.1 - *Post:* 35.7 ± 5.3   *PPO (W)*   - *Pre:* NR - *Post:* NR |
| Yarar-Fisher et al. (2018)  USA  RCT | | | | *N =* 6 (6M/0F)    *Age =* 48.5 ± 6.6 years  *TSI =* 23.8 ± 7.5 years (0 acute/ 6 chronic)  *Classification =* 2 T/ 4 P  *Severity =* 6 comp./ 0 incomp.  *CPET =* Progressive ACE test  *CPET same modality of intervention? =* Yes  *Pre-registered? =* No  *Protocol manuscript? =* No | *Type of Exercise =* Mixed (NMES, RT and ACE)  *Relative Intensity and Class =* Vigorous (80-90% VO_2peak_)  *Session Duration (min) =* 15  *Frequency (times/week) =* 3  *Intervention Length (weeks) =* 8  *Adverse Events =* NR | *AV̇O_2peak_ (L/min)*   - *Pre:* NR - *Post:* NR   *RV̇O_2peak_ (mL/kg/min)*   - *Pre:* 13.2 ± 3.4 - *Post:* 14.6 ± 3.3   *PPO (W)*   - *Pre:* NR - *Post:* NR |

Data is presented as mean ± standard deviation, unless otherwise stated. 1RM, one repetition maximum; ACE, arm crank ergometry; AV̇O_2peak_, absolute peak oxygen consumption; BWSTT, body weight supported treadmill training; CPET, cardiopulmonary exercise test; F, females; FES, functional electrical stimulation; HIIT, high-intensity interval training; HR_max_, maximum heart rate; HR_peak_, peak heart rate; HRR, heart rate reserve; M, male; NMES, neuromuscular electrical stimulation; NR, not reported; P, paraplegia; PPO, peak power output; RABWSTT, robotic-assisted body weight supported treadmill training; RCT, randomised-controlled trial; RPE, rating of perceived exertion; RT, resistance training; RV̇O_2peak_, relative peak oxygen consumption; T, tetraplegia; TSI, time since injury; V̇O_2peak_, peak oxygen consumption; W, watts.

**TIME SINCE INJURY**

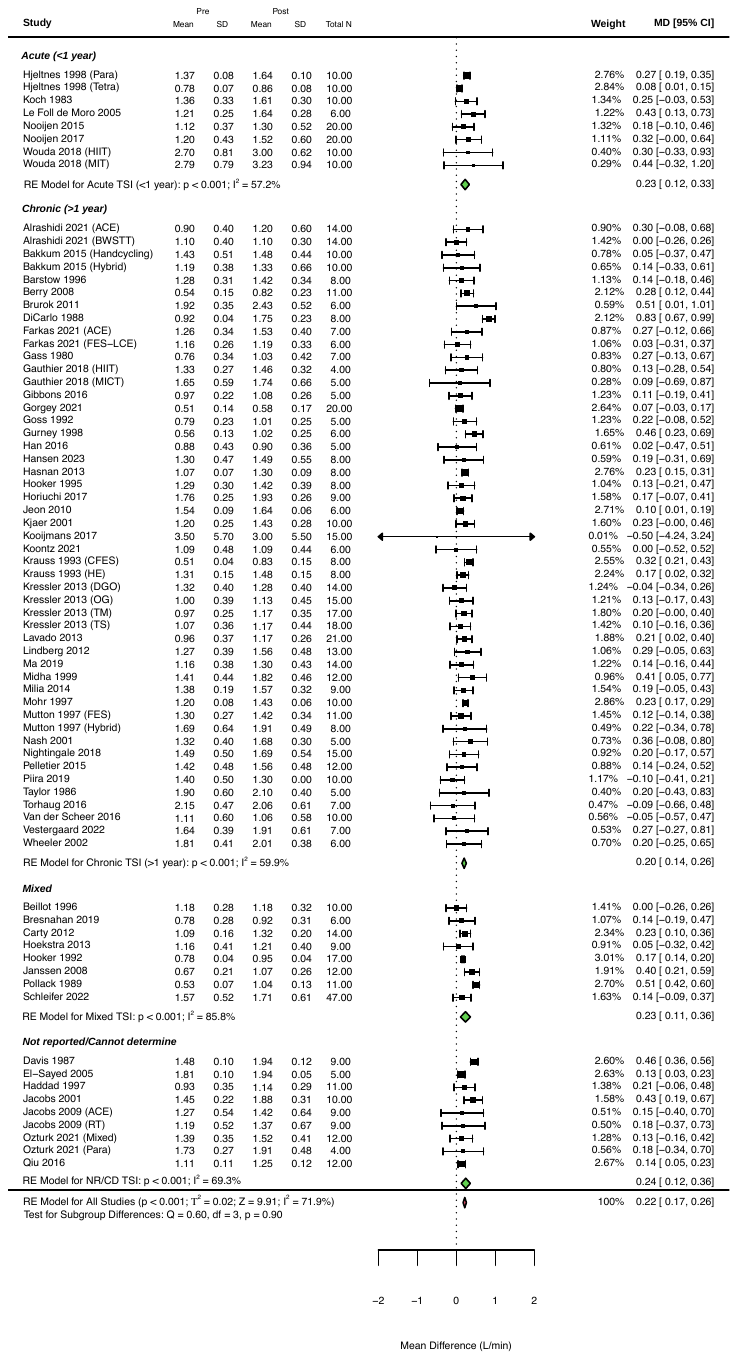

**Figure 2:** Changes in absolute peak oxygen consumption following exercise interventions > 2 weeks, sub-grouped based on time since injury.

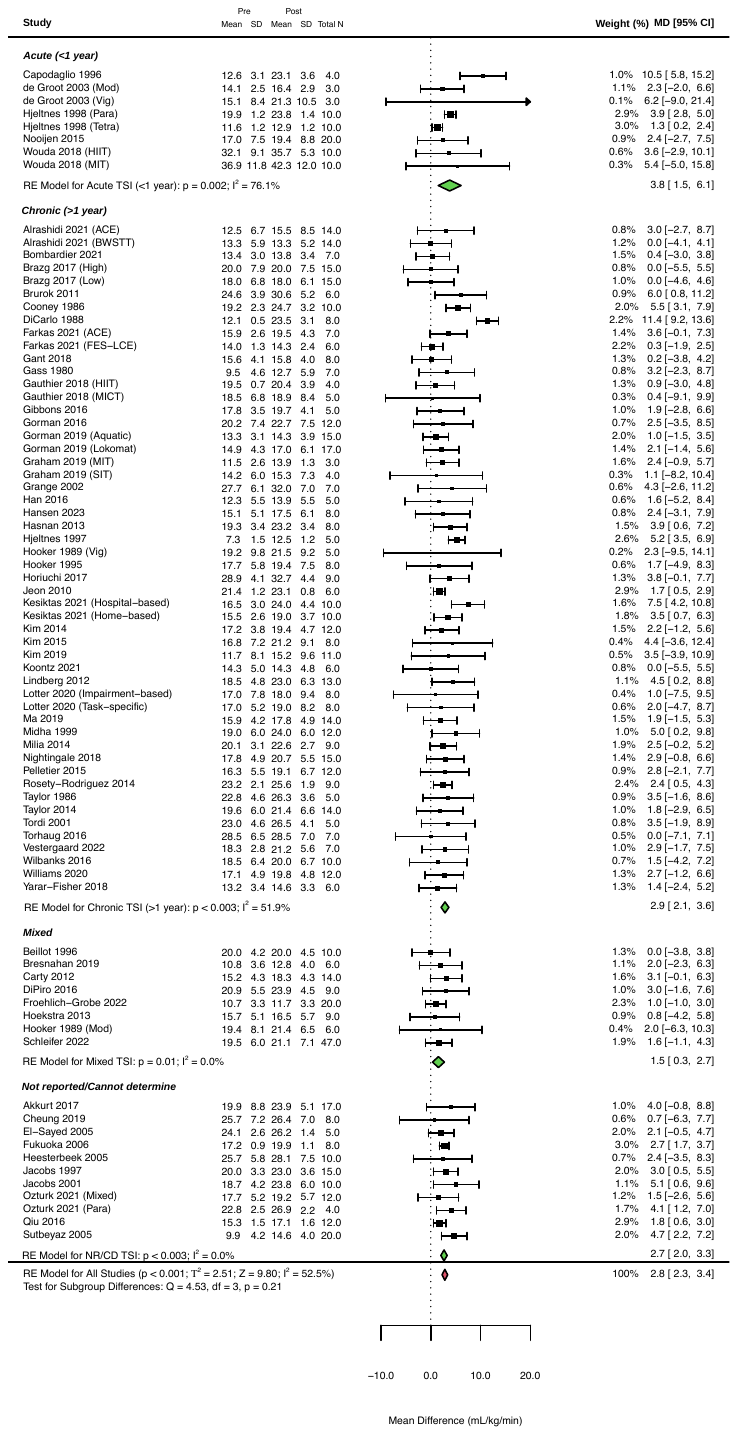

**Figure 3:** Changes in relative peak oxygen consumption following exercise interventions > 2 weeks, sub-grouped based on time since injury.

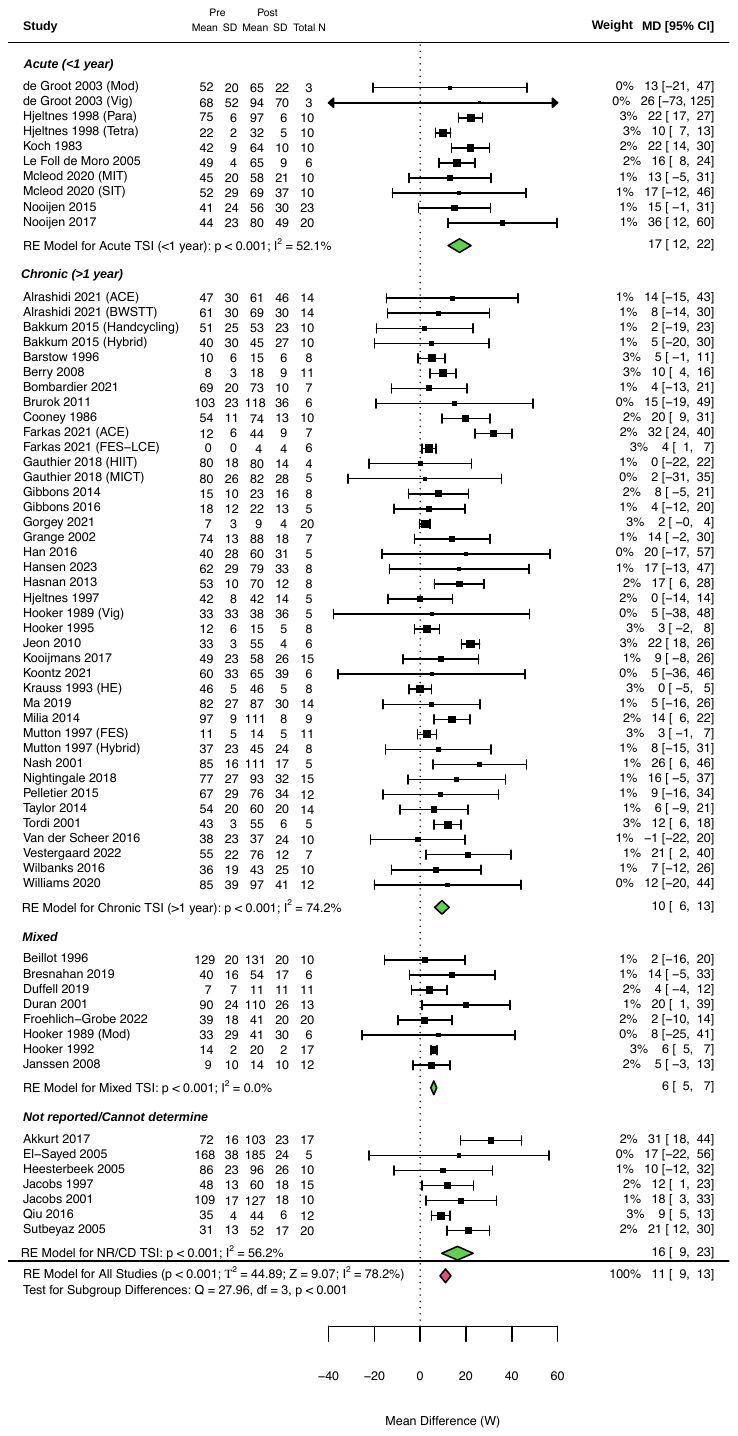

**Figure 4:** Changes in peak power output following exercise interventions > 2 weeks, sub-grouped based on time since injury.

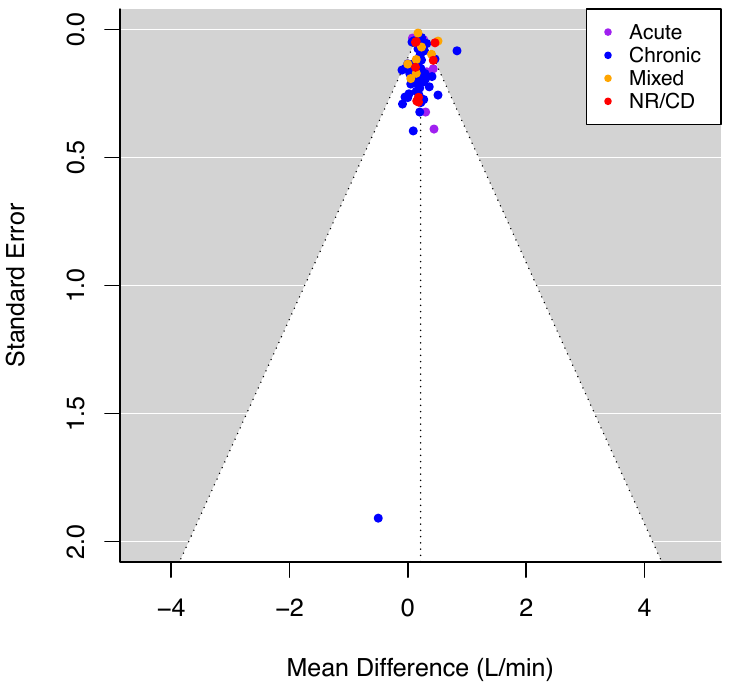

**Figure 5:** Funnel plot of absolute peak oxygen consumption with studies sub-grouped based on time since injury.

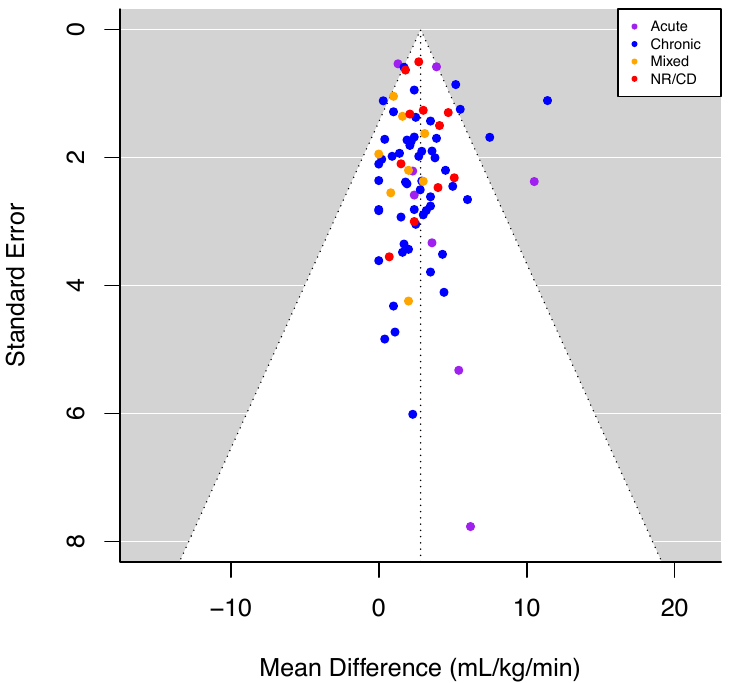

**Figure 6:** Funnel plot of relative peak oxygen consumption with studies sub-grouped based on time since injury.

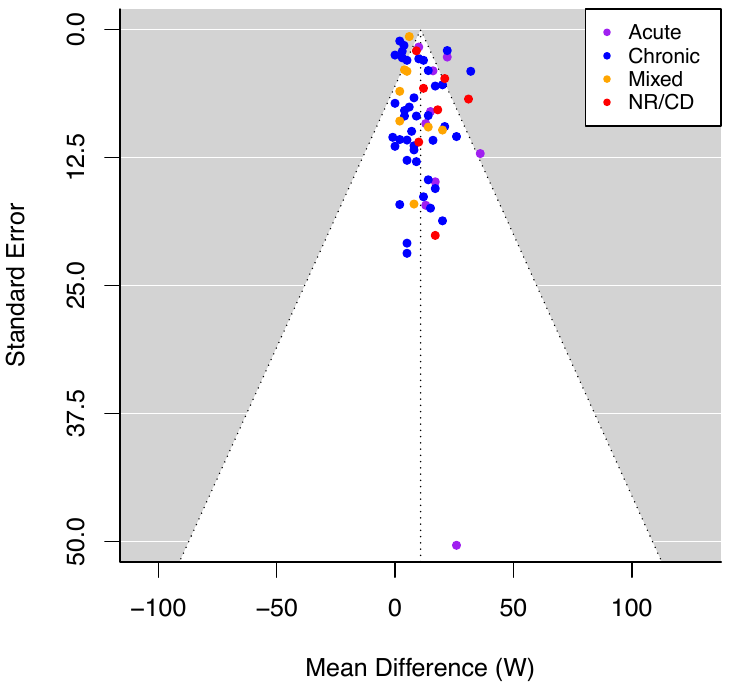

**Figure 7:** Funnel plot of peak power output with studies sub-grouped based on time since injury.

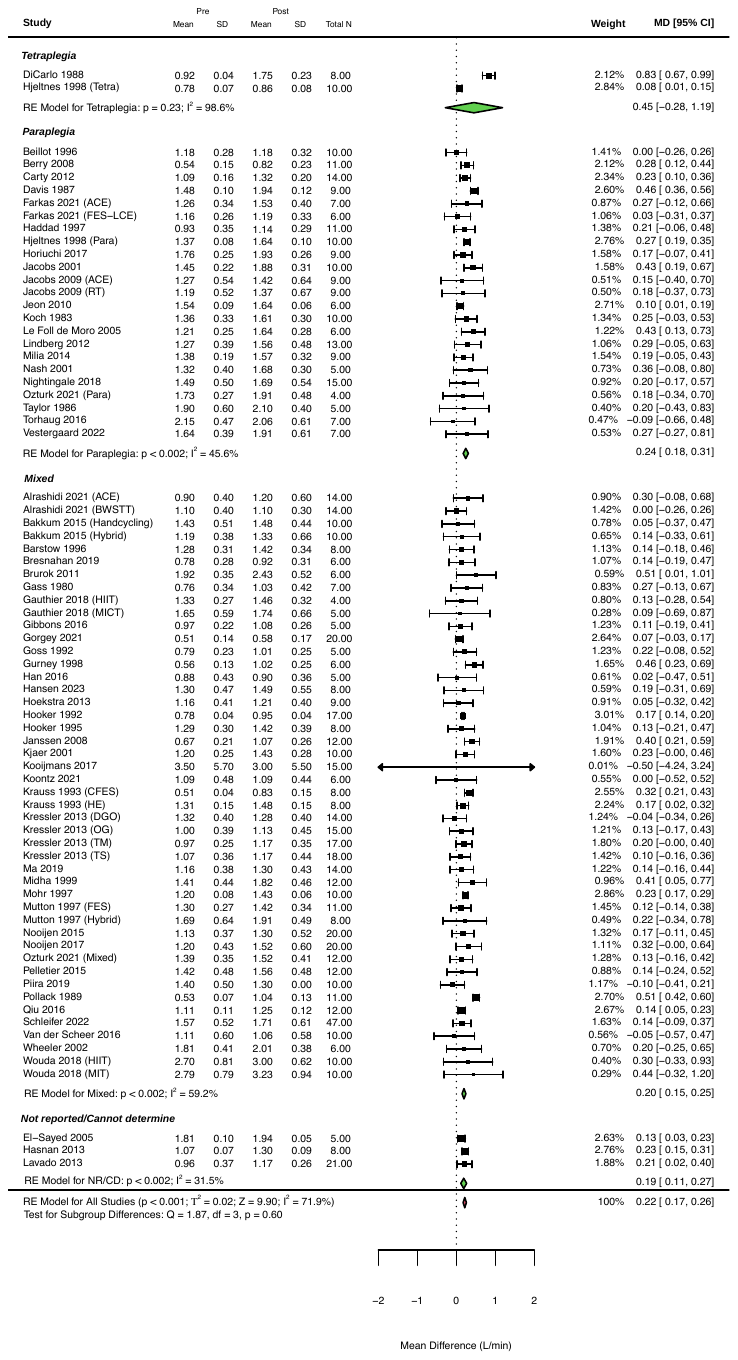
**NEUROLOGICAL LEVEL OF INJURY**

**Figure 8:** Changes in absolute peak oxygen consumption following exercise interventions > 2 weeks, sub-grouped based on neurological level of injury.

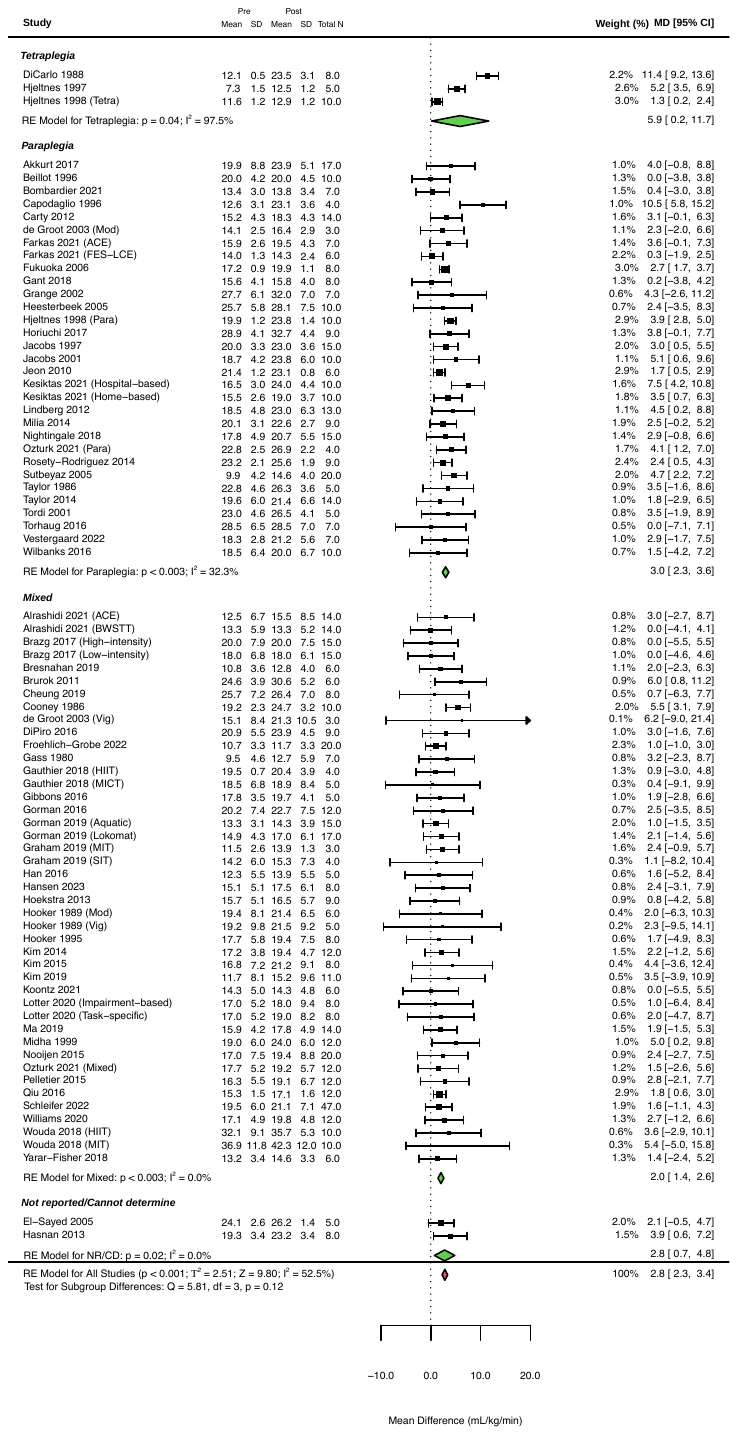

**Figure 9:** Changes in relative peak oxygen consumption following exercise interventions > 2 weeks, sub-grouped based on neurological level of injury.

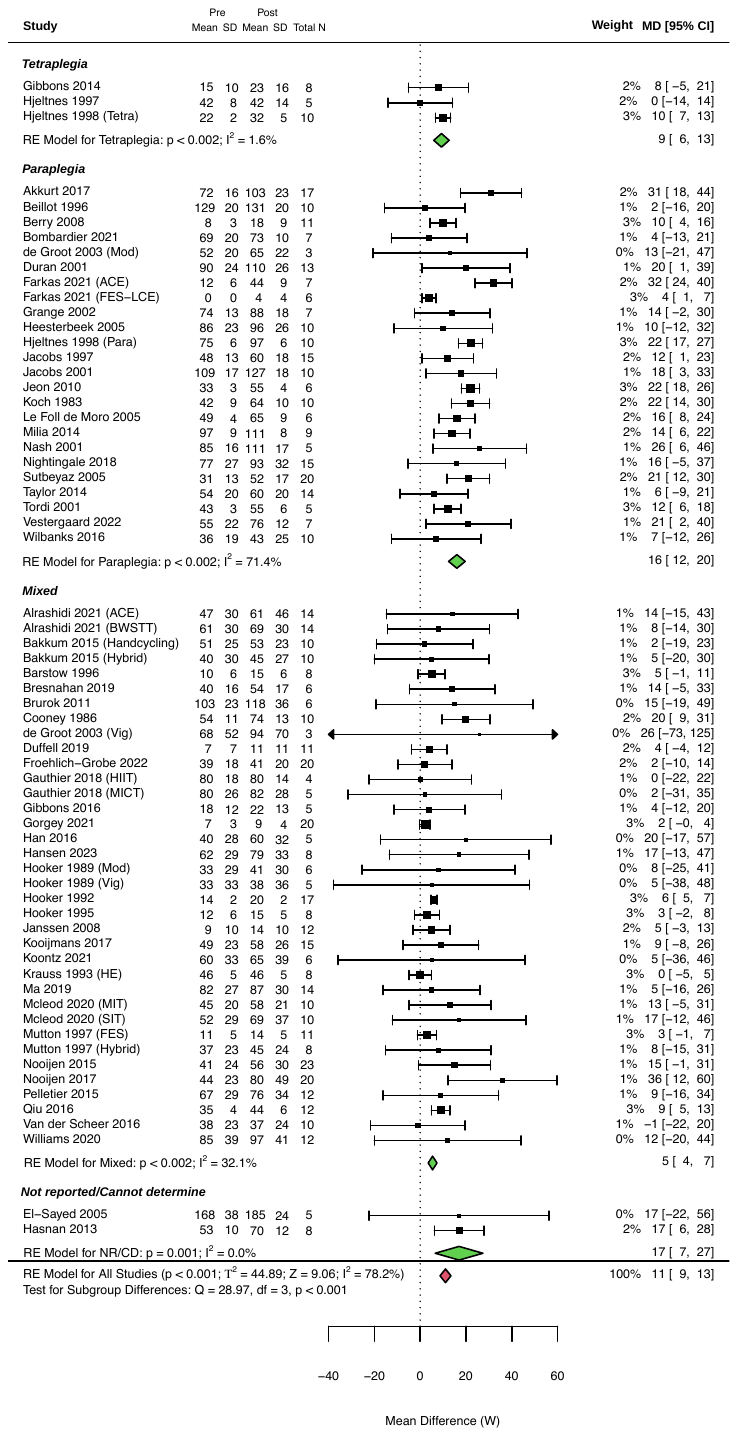

**Figure 10:** Changes in peak power output following exercise interventions > 2 weeks, sub-grouped based on neurological level of injury.

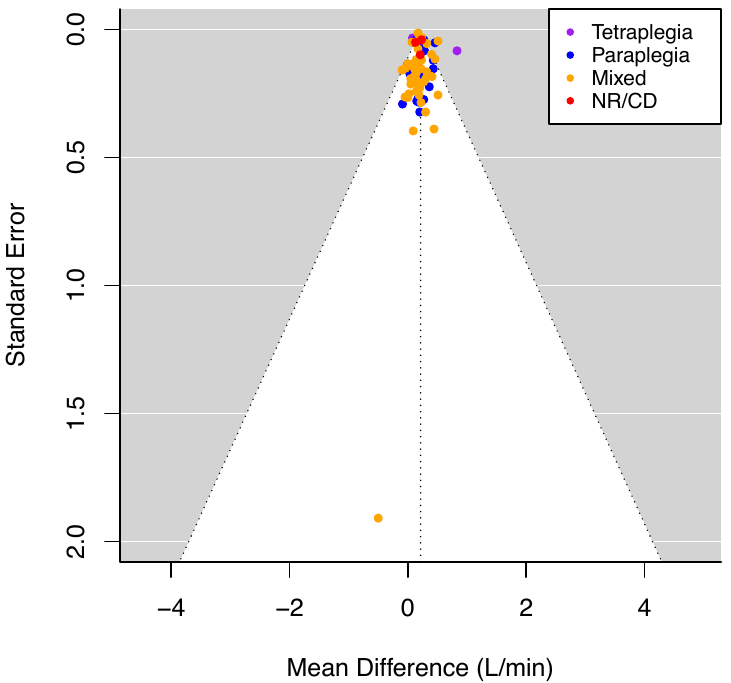

**Figure 11:** Funnel plot of absolute peak oxygen consumption with studies sub-grouped based on neurological level of injury.

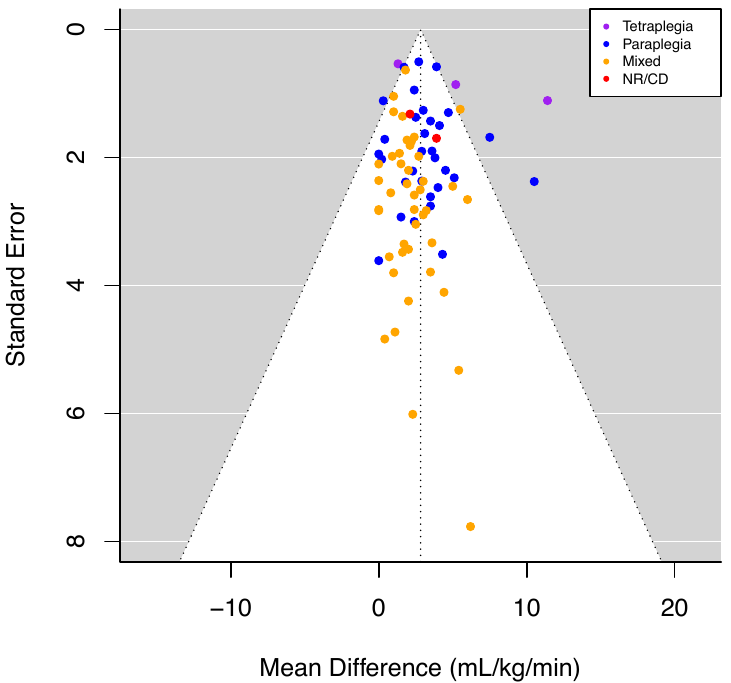

**Figure 12:** Funnel plot of relative peak oxygen consumption with studies sub-grouped based on neurological level of injury.

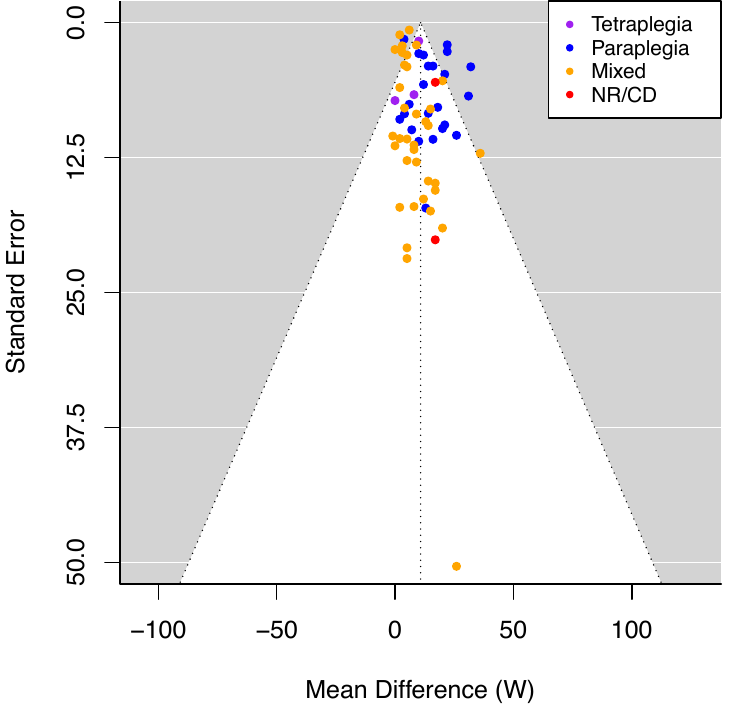

**Figure 13:** Funnel plot of peak power output with studies sub-grouped based on neurological level of injury.

**INJURY SEVERITY**

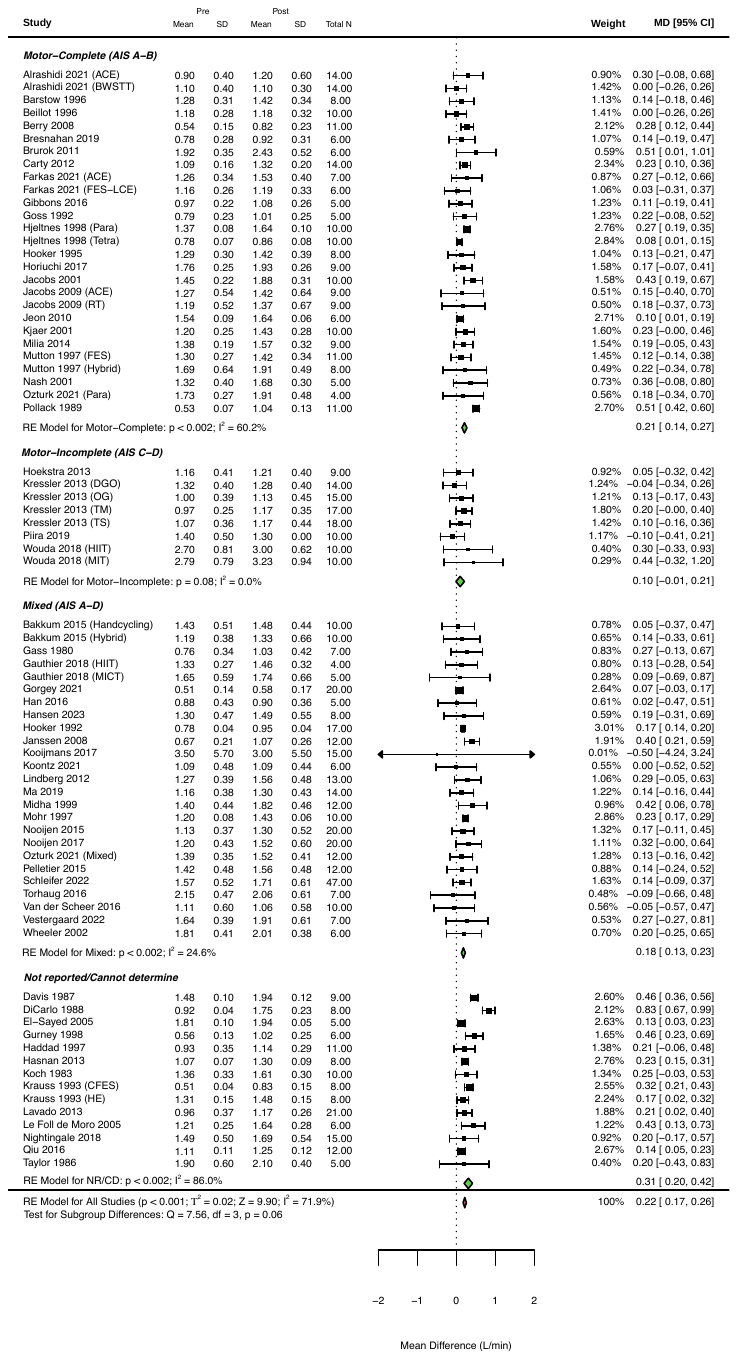

**Figure 14:** Changes in absolute peak oxygen consumption following exercise interventions > 2 weeks, sub-grouped based on injury severity.

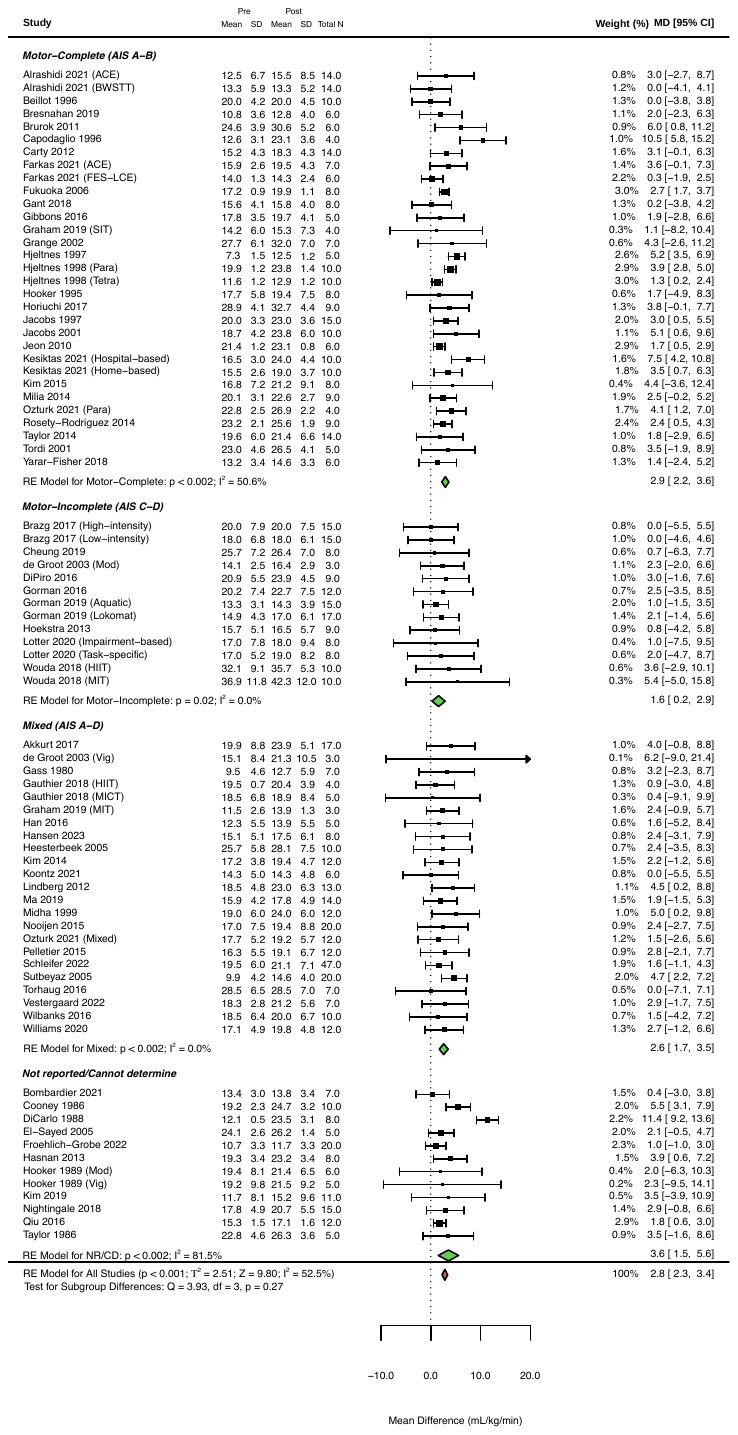

**Figure 15:** Changes in relative peak oxygen consumption following exercise interventions > 2 weeks, sub-grouped based on injury severity.

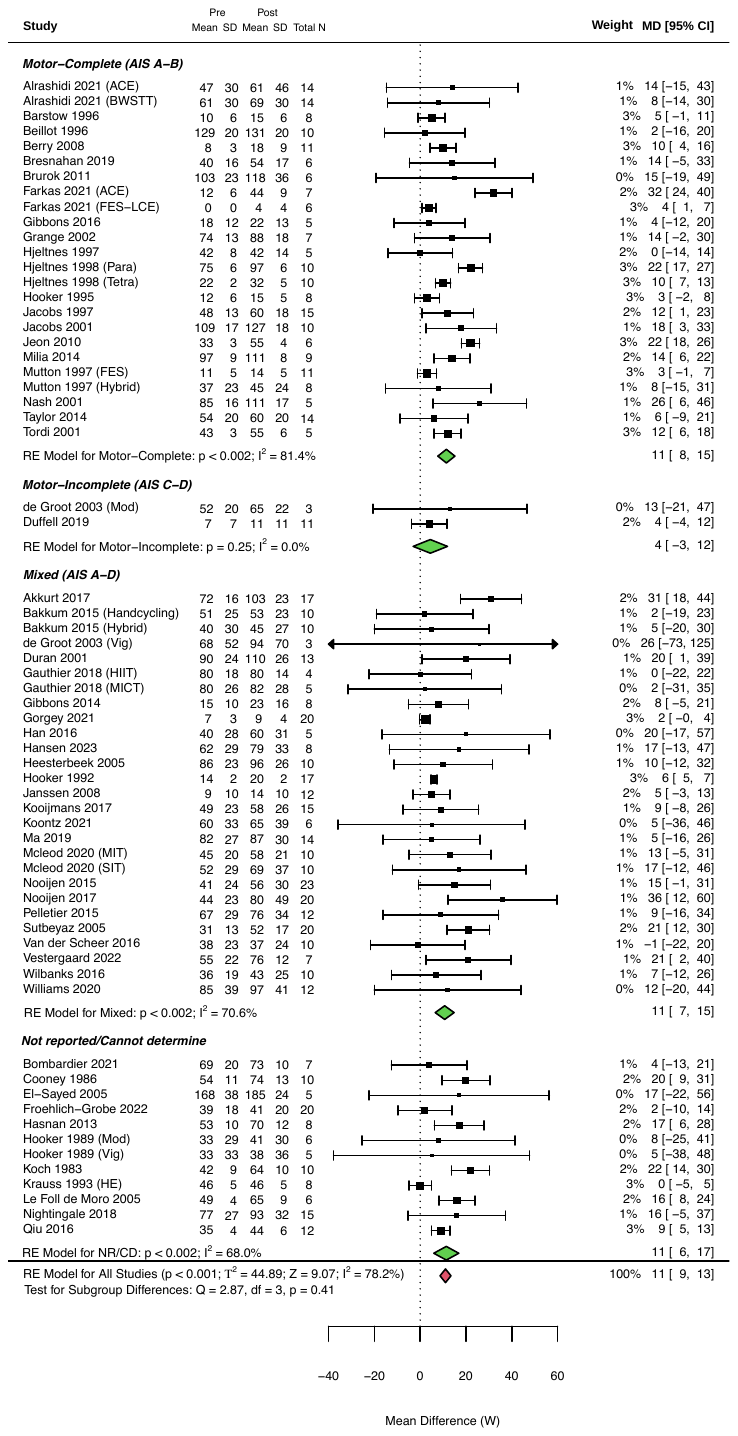

**Figure 16:** Changes in peak power output following exercise interventions > 2 weeks, sub-grouped based on injury severity.

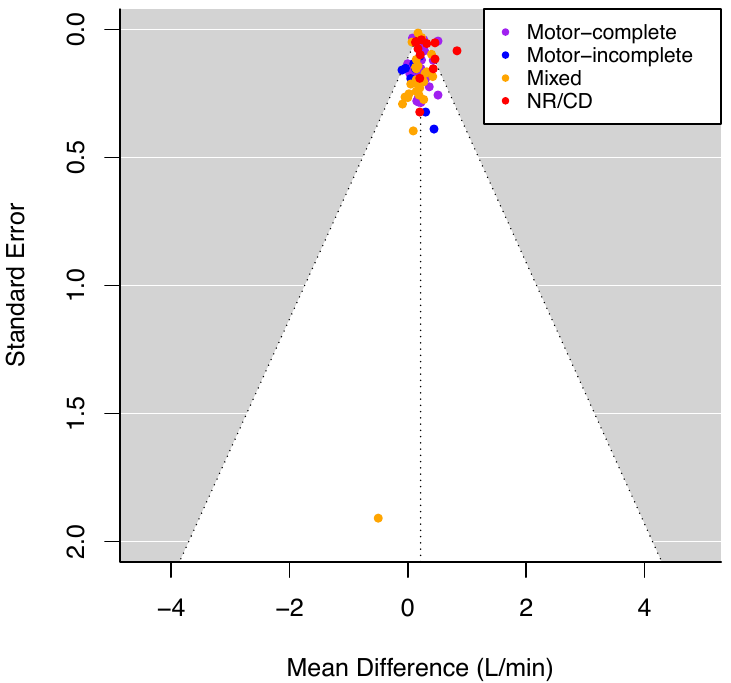

**Figure 17:** Funnel plot of absolute peak oxygen consumption with studies sub-grouped based on injury severity.

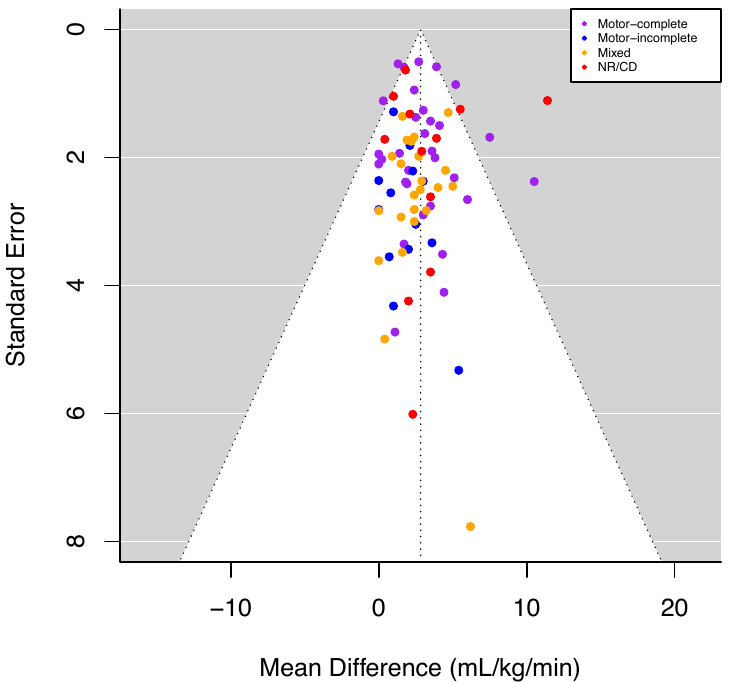

**Figure 18:** Funnel plot of relative peak oxygen consumption with studies sub-grouped based on injury severity.

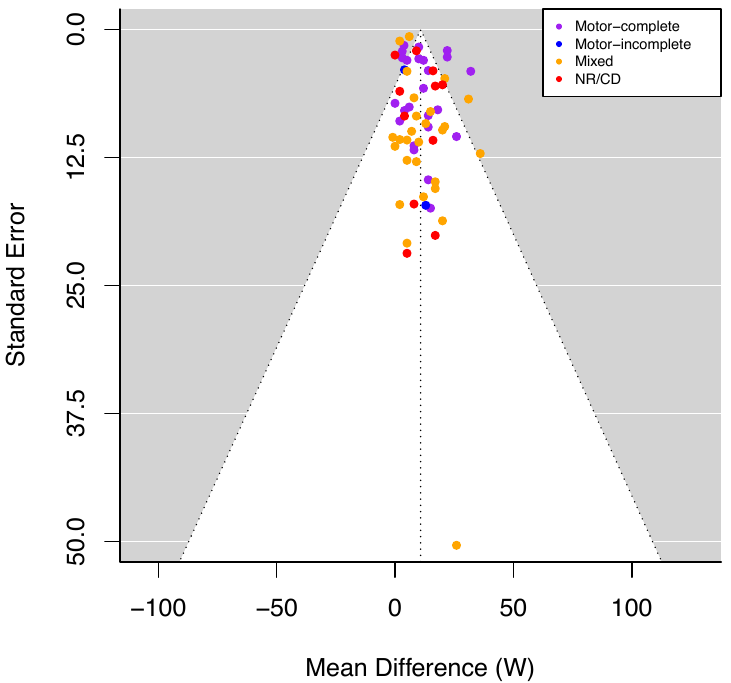

**Figure 19:** Funnel plot of peak power output with studies sub-grouped based on injury severity.

**EXERCISE MODALITY**

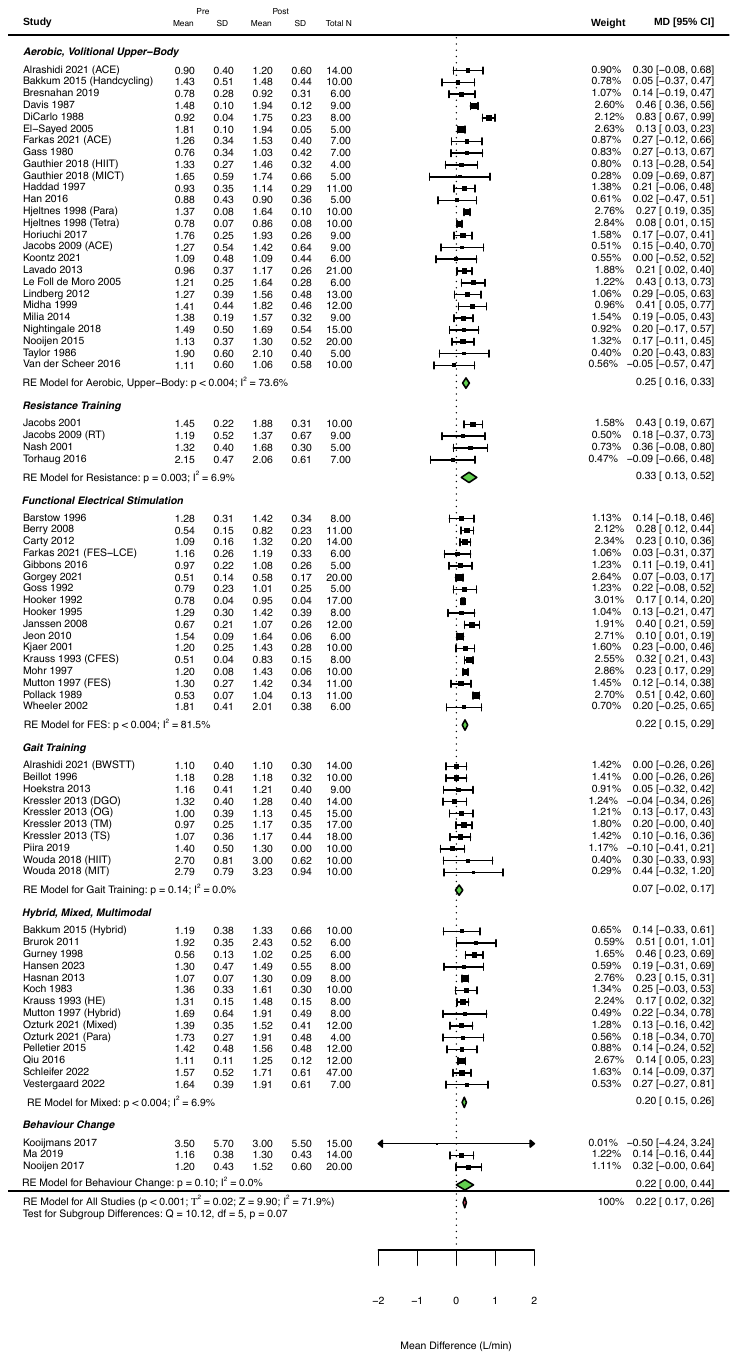

**Figure 20:** Changes in absolute peak oxygen consumption following exercise interventions > 2 weeks, sub-grouped based on exercise modality.

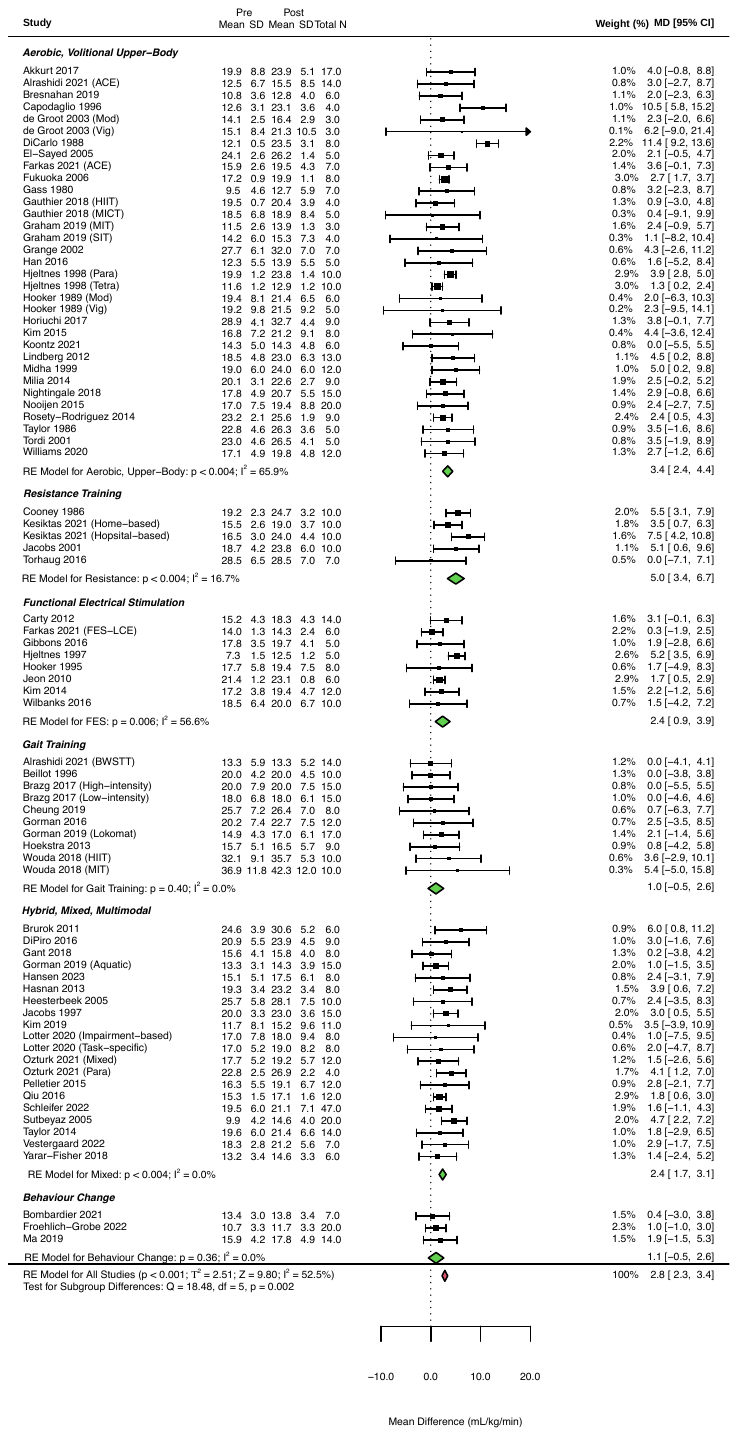

**Figure 21:** Changes in relative peak oxygen consumption following exercise interventions > 2 weeks, sub-grouped based on exercise modality.

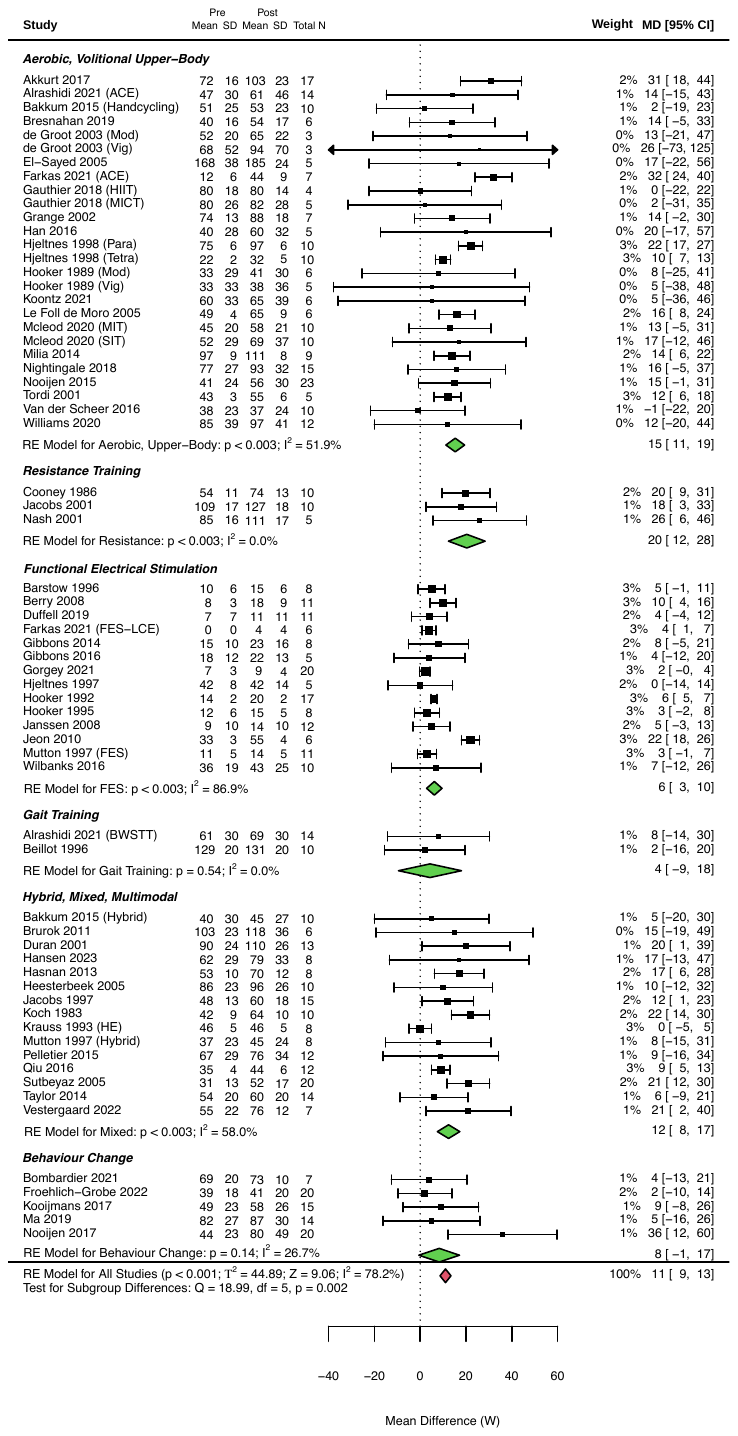

**Figure 22:** Changes in peak power output following exercise interventions > 2 weeks, sub-grouped based on exercise modality.

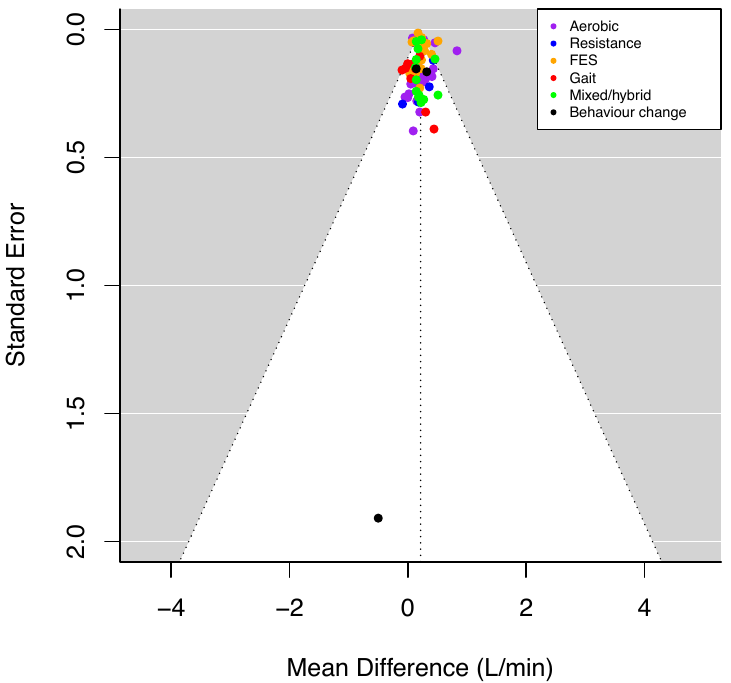

**Figure 23:** Funnel plot of absolute peak oxygen consumption with studies sub-grouped based on exercise modality.

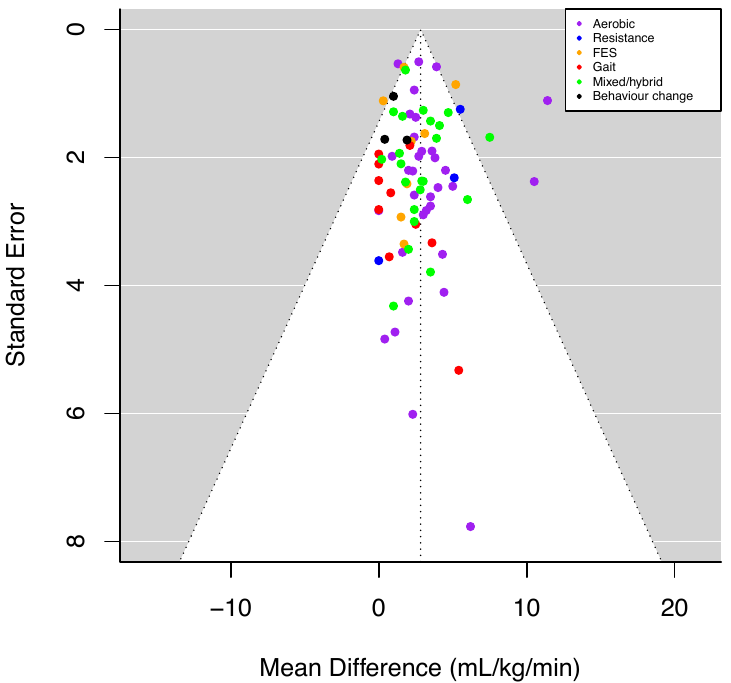

**Figure 24:** Funnel plot of relative peak oxygen consumption with studies sub-grouped based on exercise modality.

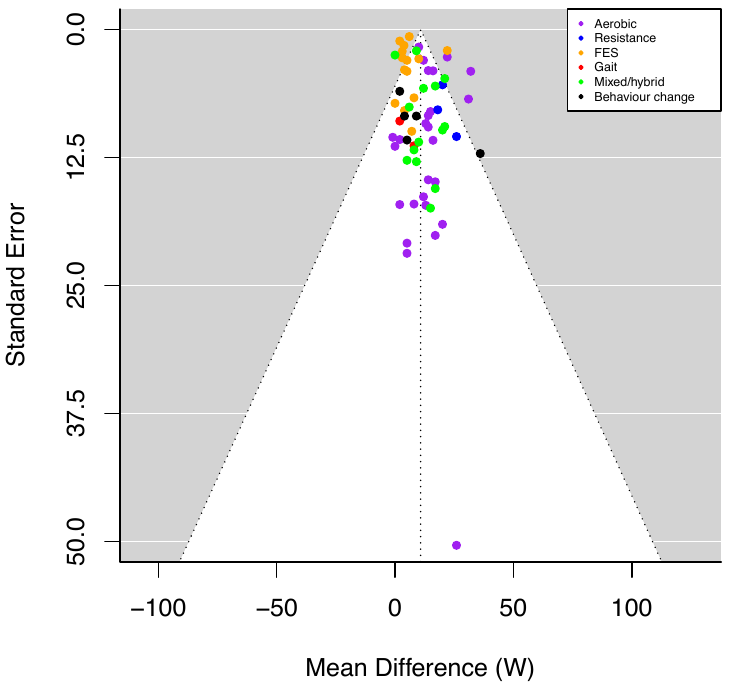

**Figure 25:** Funnel plot of peak power output with studies sub-grouped based on exercise modality.

**LENGTH OF INTERVENTION**

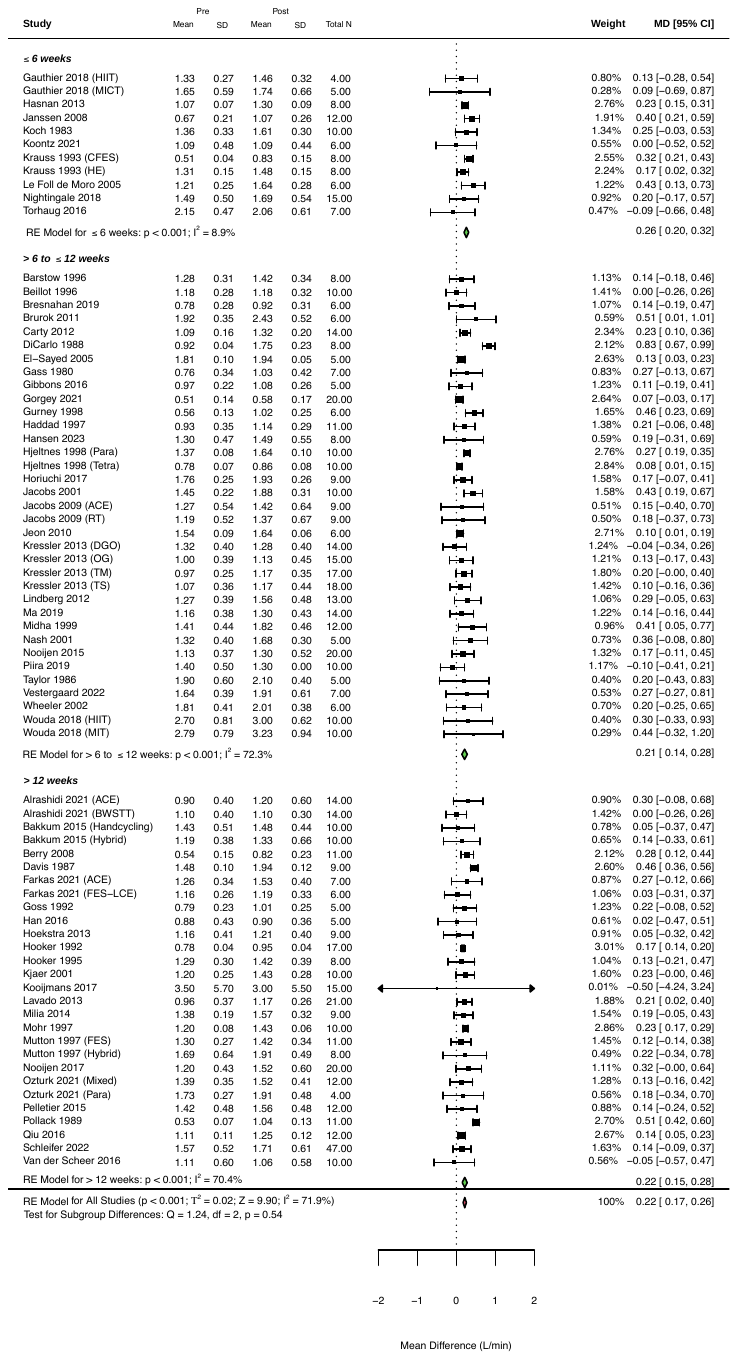

**Figure 26:** Changes in absolute peak oxygen consumption following exercise interventions > 2 weeks, sub-grouped based on length of intervention.

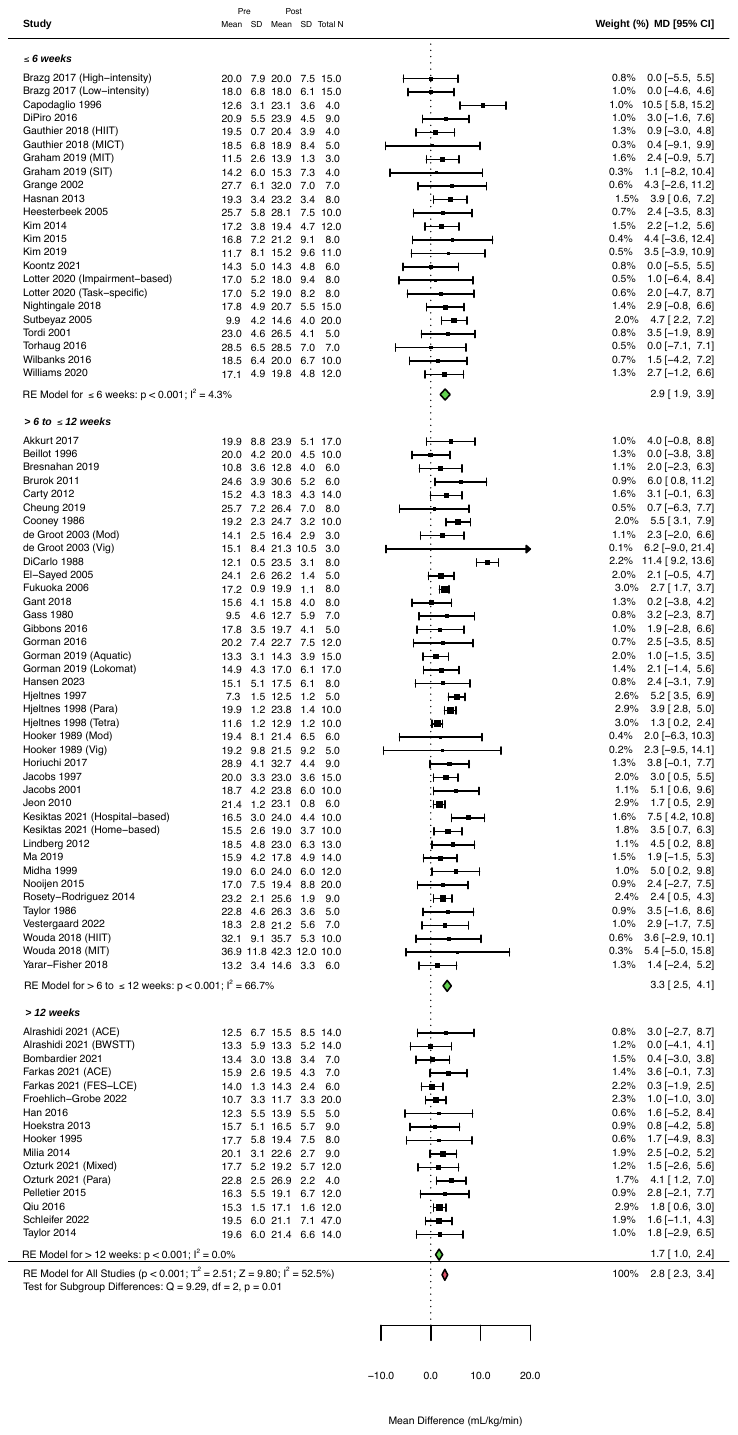

**Figure 27:** Changes in relative peak oxygen consumption following exercise interventions > 2 weeks, sub-grouped based on length of intervention.

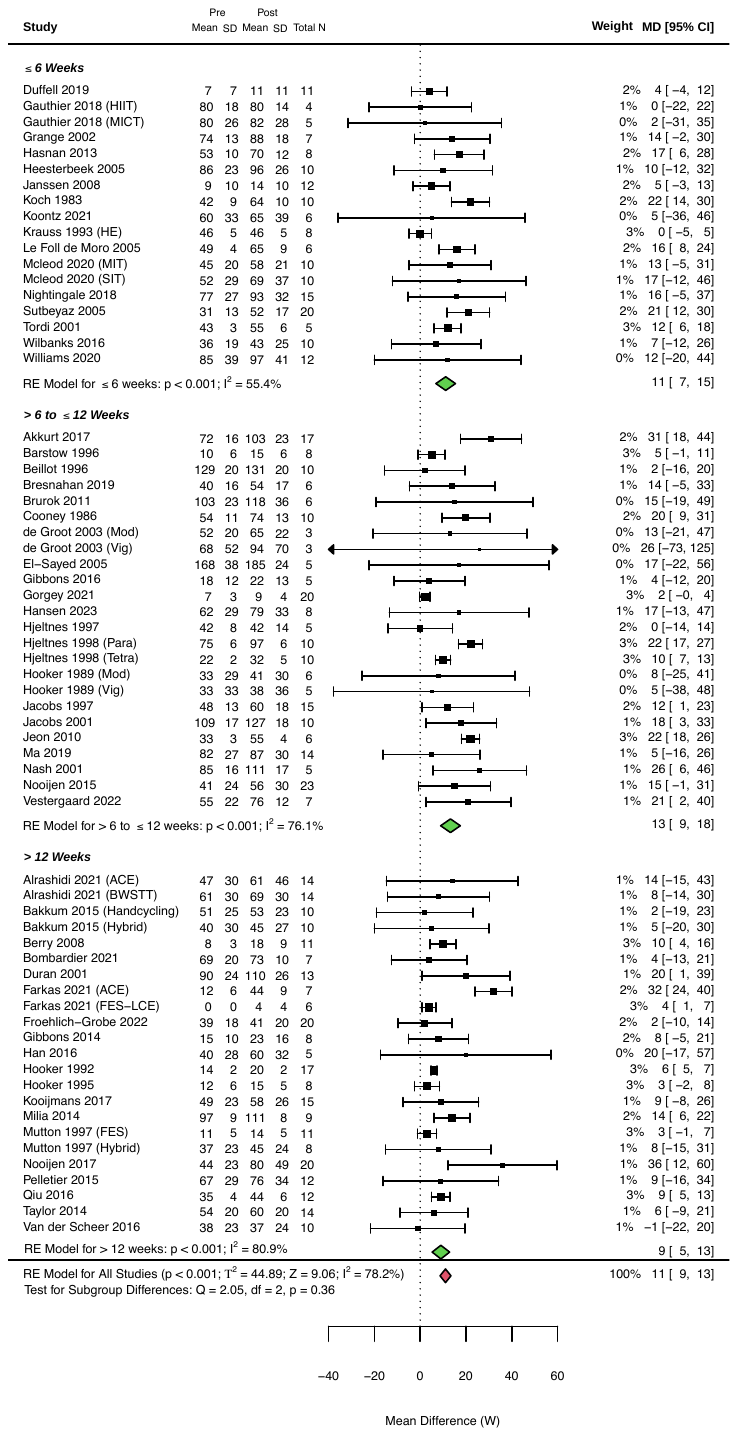

**Figure 28:** Changes in peak power output following exercise interventions > 2 weeks, sub-grouped based on length of intervention.

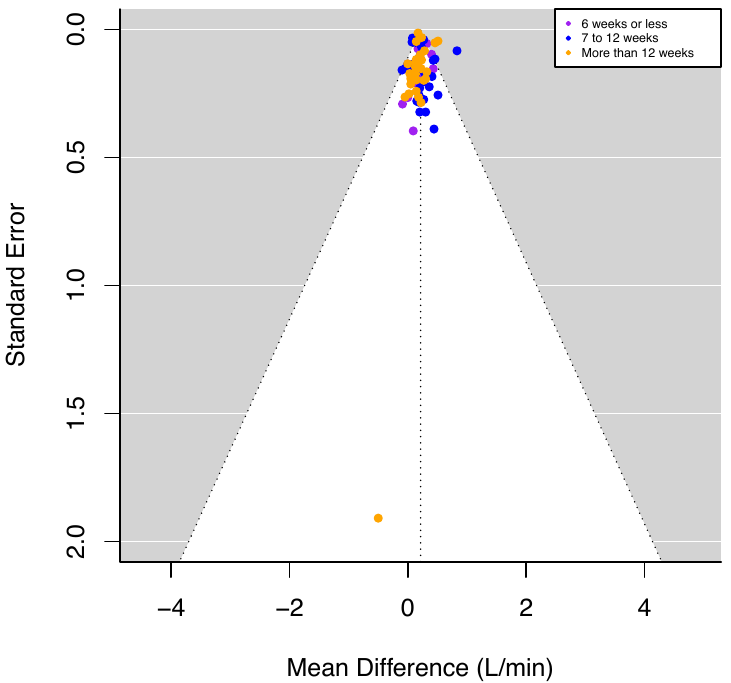

**Figure 29:** Funnel plot of absolute peak oxygen consumption with studies sub-grouped based on length of intervention.

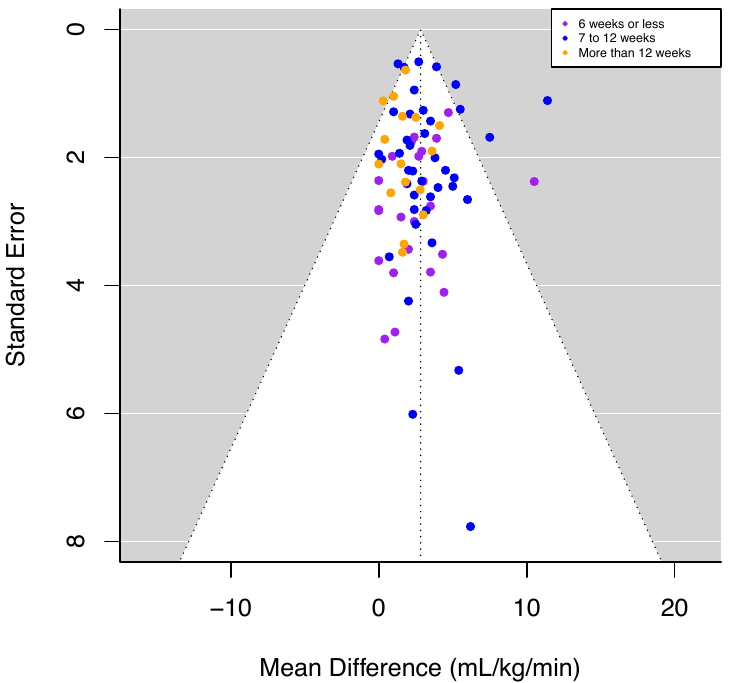

**Figure 30:** Funnel plot of relative peak oxygen consumption with studies sub-grouped based on length of intervention.

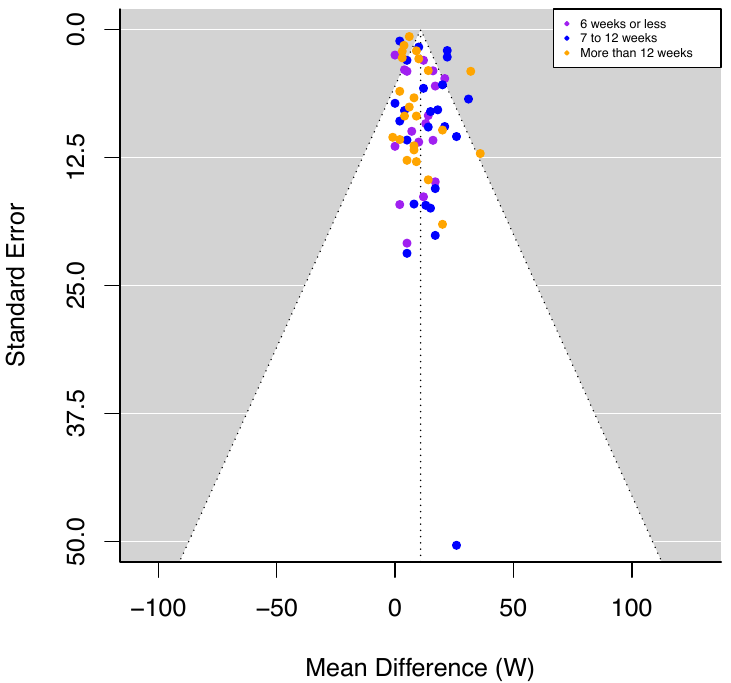

**Figure 31:** Funnel plot of peak power output with studies sub-grouped based on length of intervention.

**RELATIVE EXERCISE INTENSITY**

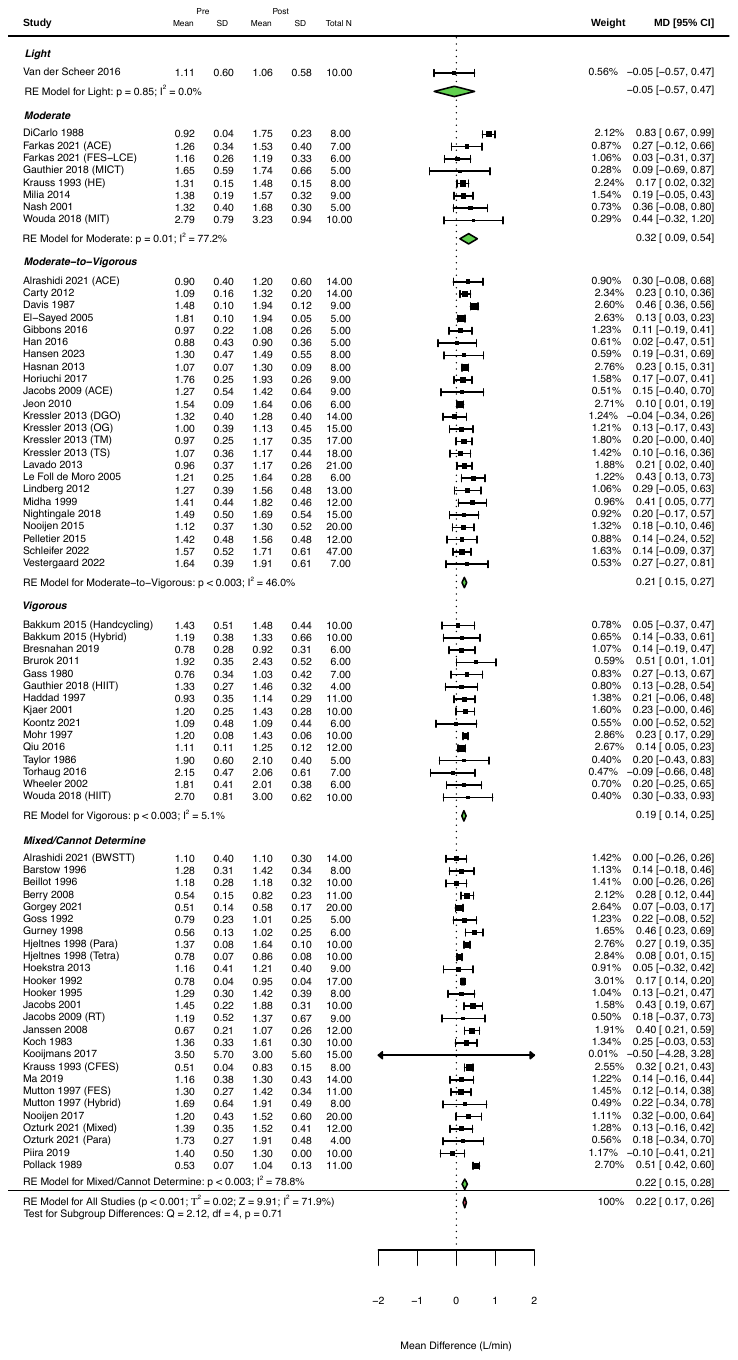

**Figure 32:** Changes in absolute peak oxygen consumption following exercise interventions > 2 weeks, sub-grouped based on relative exercise intensity.

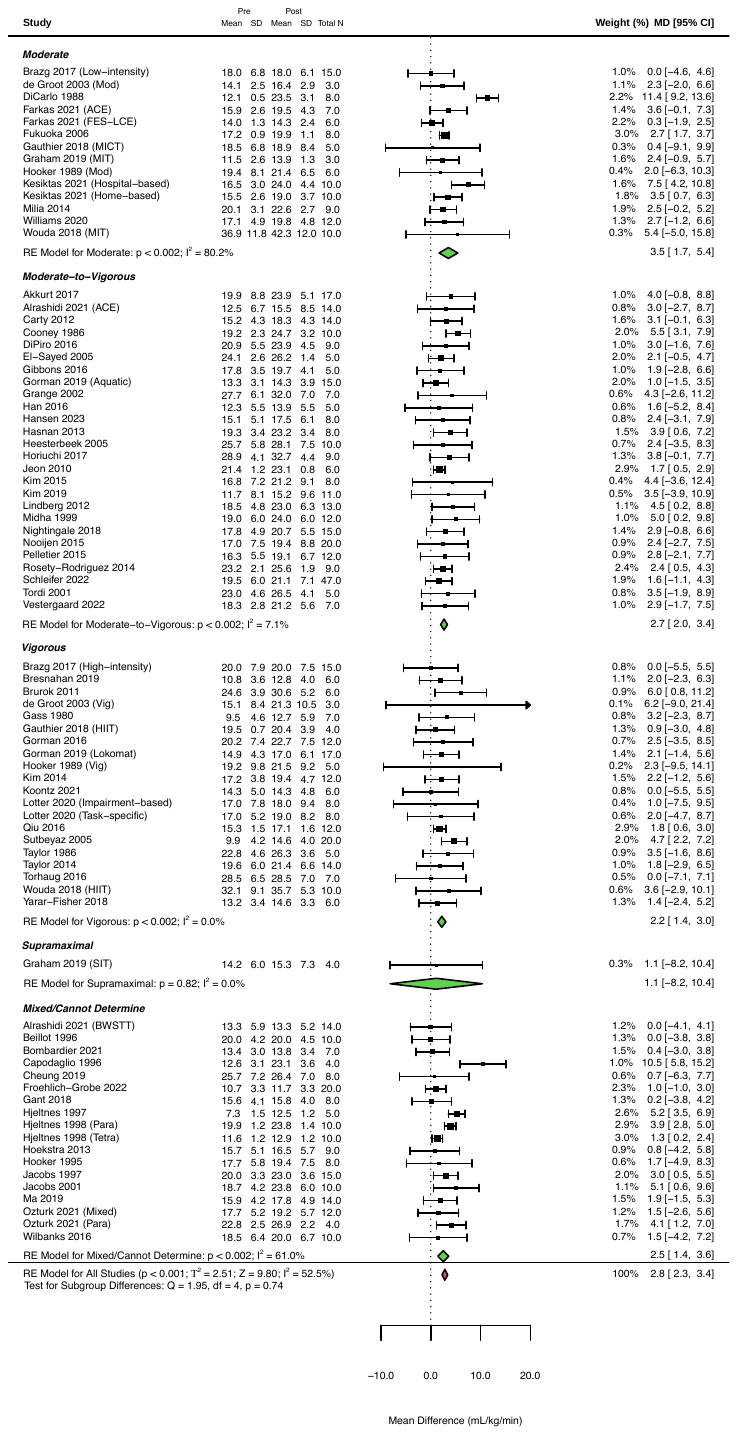

**Figure 33:** Changes in absolute peak oxygen consumption following exercise interventions > 2 weeks, sub-grouped based on relative exercise intensity.

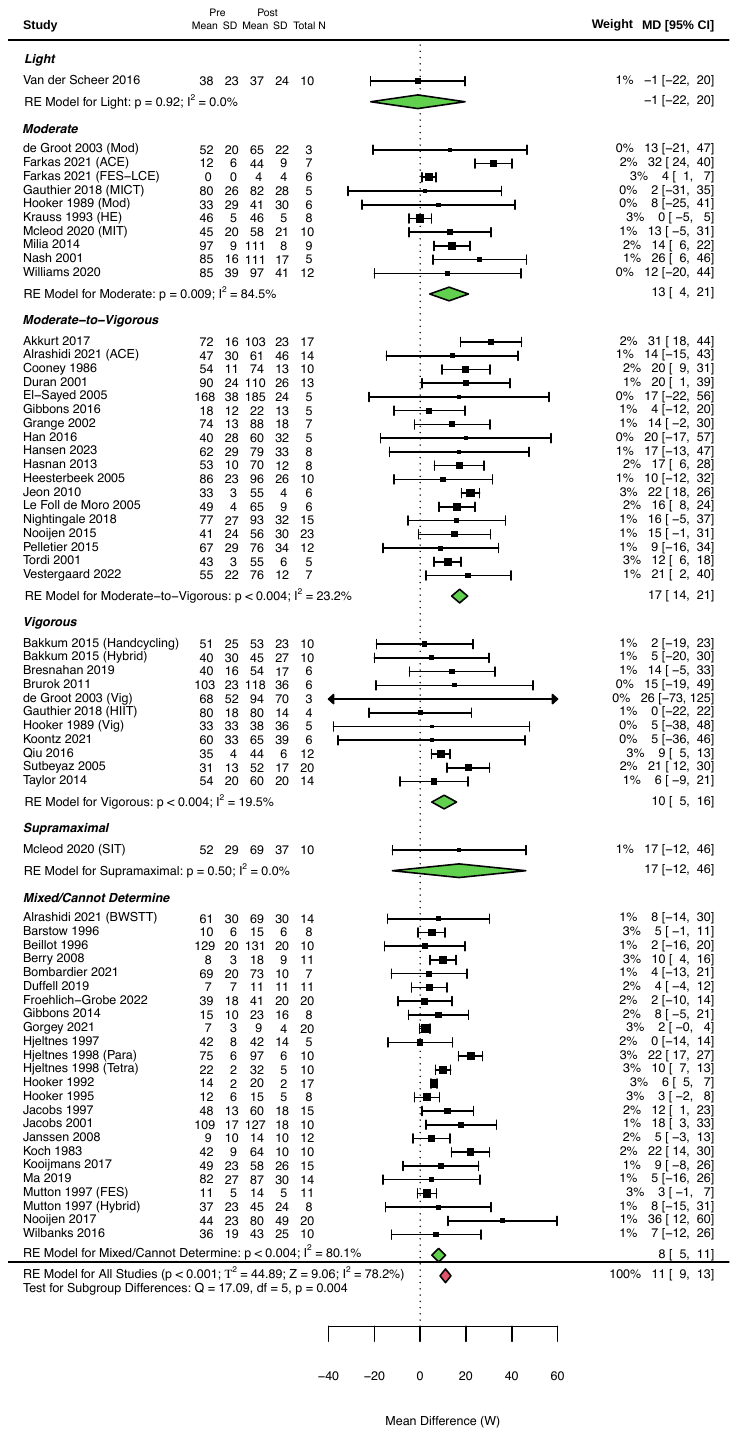

**Figure 34:** Changes in peak power output following exercise interventions > 2 weeks, sub-grouped based on relative exercise intensity.

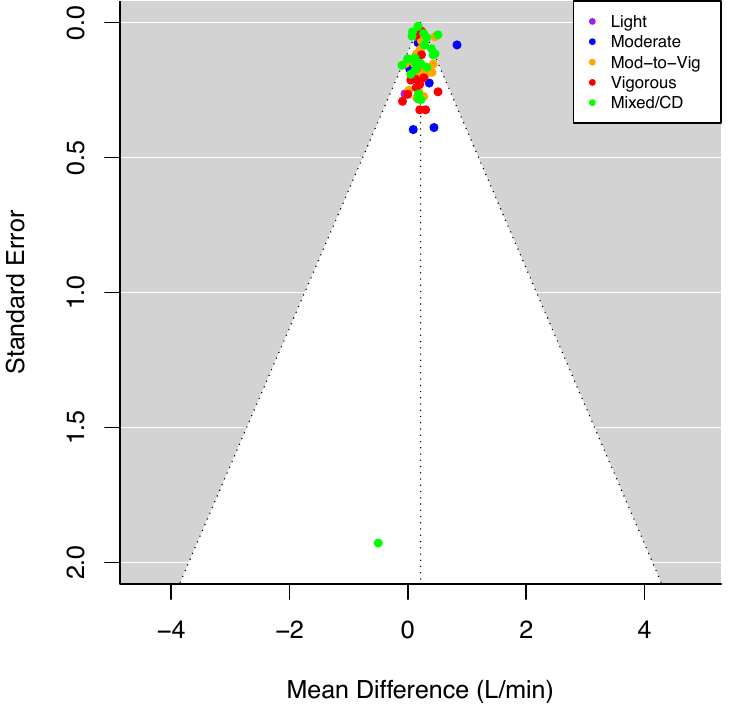

**Figure 35:** Funnel plot of absolute peak oxygen consumption with studies sub-grouped based on relative exercise intensity.

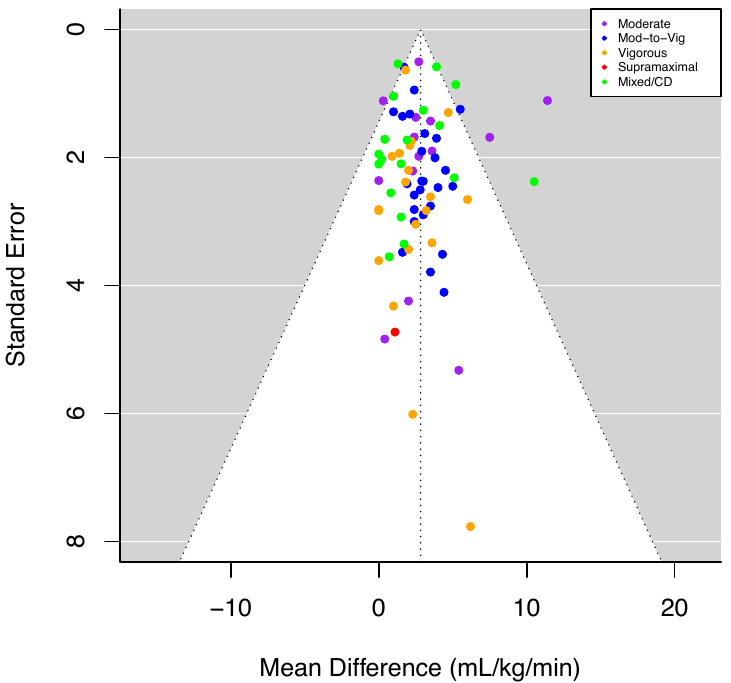

**Figure 36:** Funnel plot of relative peak oxygen consumption with studies sub-grouped based on relative exercise intensity.

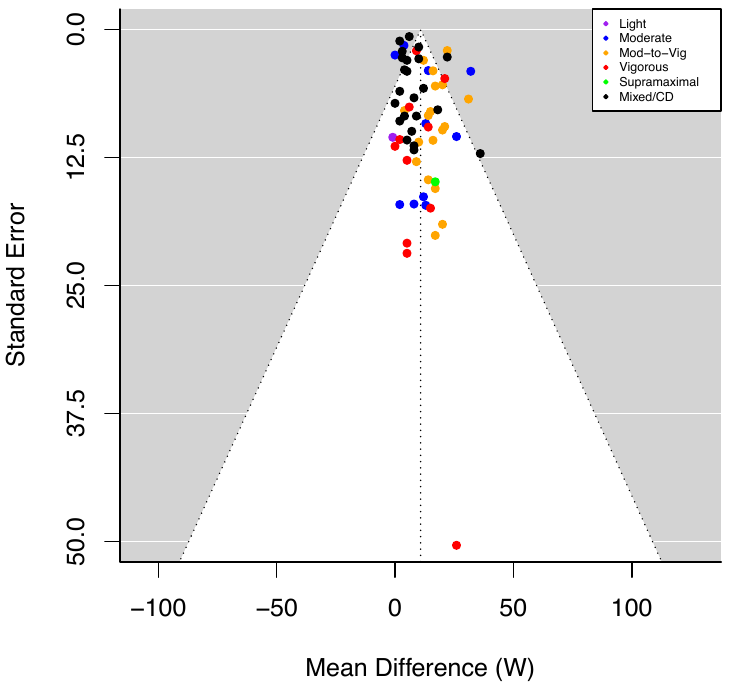

**Figure 37:** Funnel plot of peak power output with studies sub-grouped based on relative exercise intensity.

**METHOD OF EXERCISE INTENSITY PRESCRIPTION**

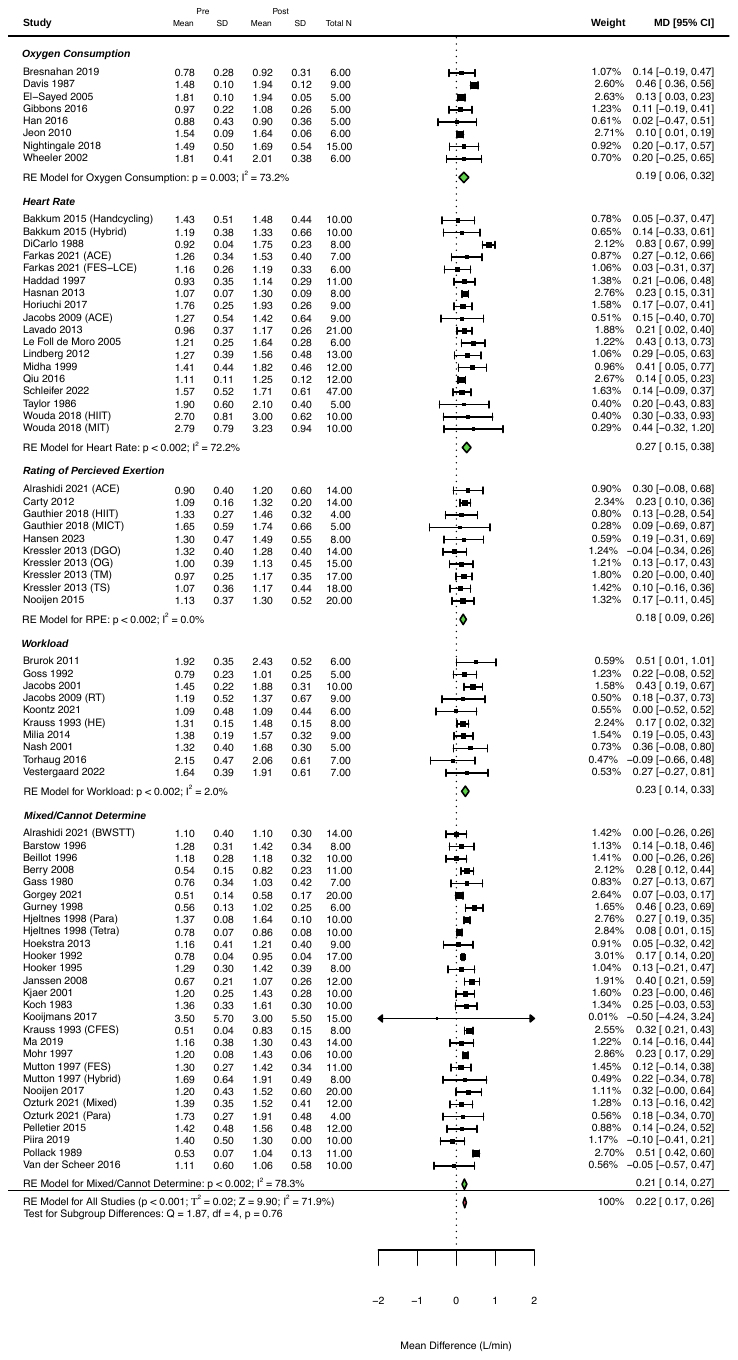
**Figure 38:** Changes in absolute peak oxygen consumption following exercise interventions > 2 weeks, sub-grouped based on method of exercise intensity prescription.

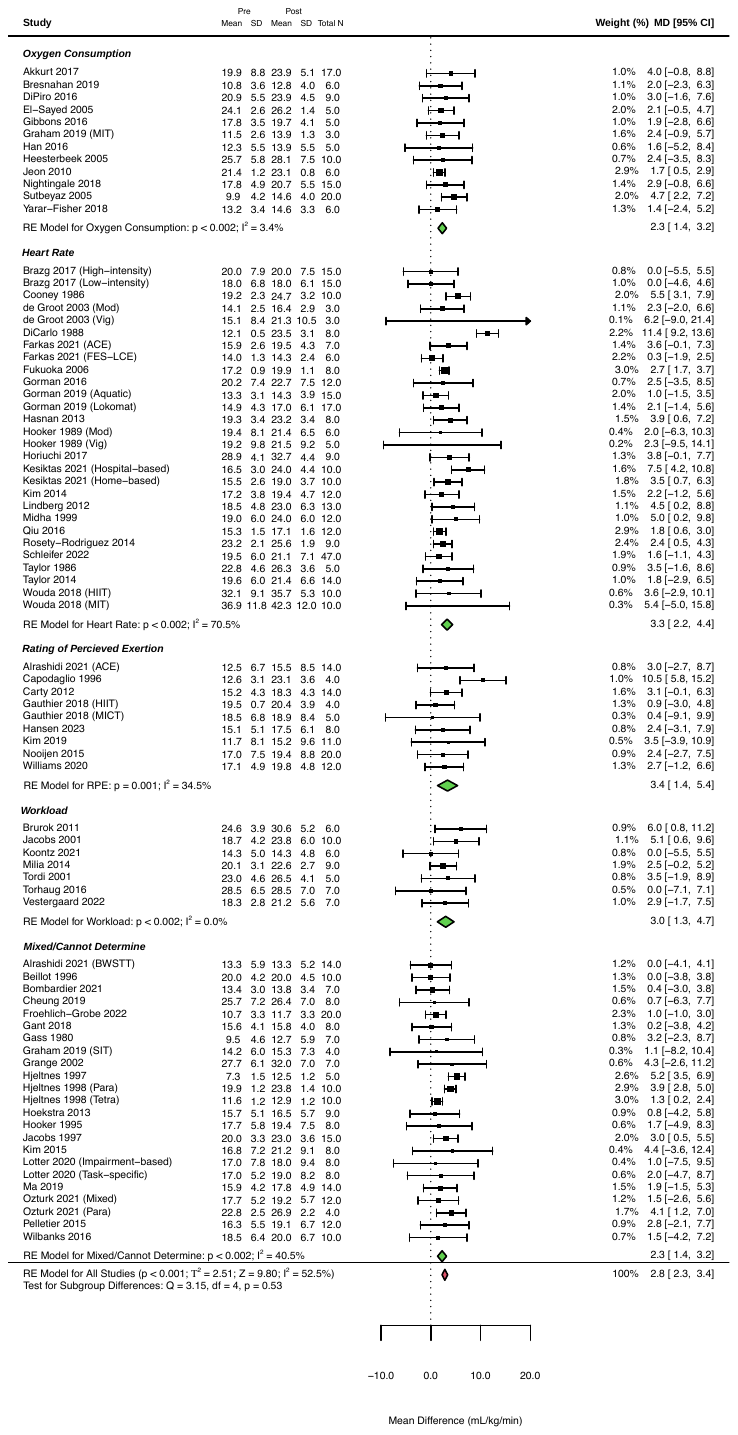

**Figure 39:** Changes in relative peak oxygen consumption following exercise interventions > 2 weeks, sub-grouped based on method of exercise intensity prescription.

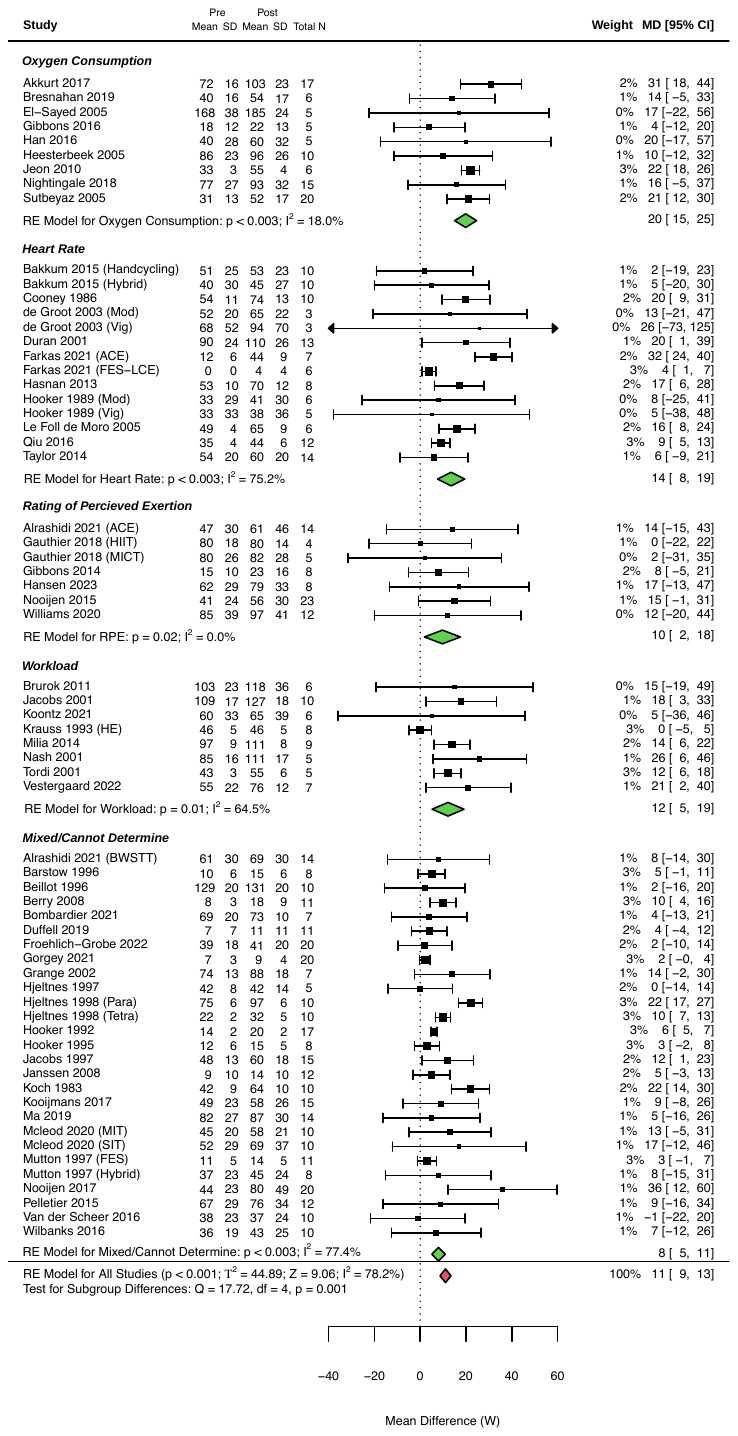

**Figure 40:** Changes in peak power output following exercise interventions > 2 weeks, sub-grouped based on method of exercise intensity prescription.

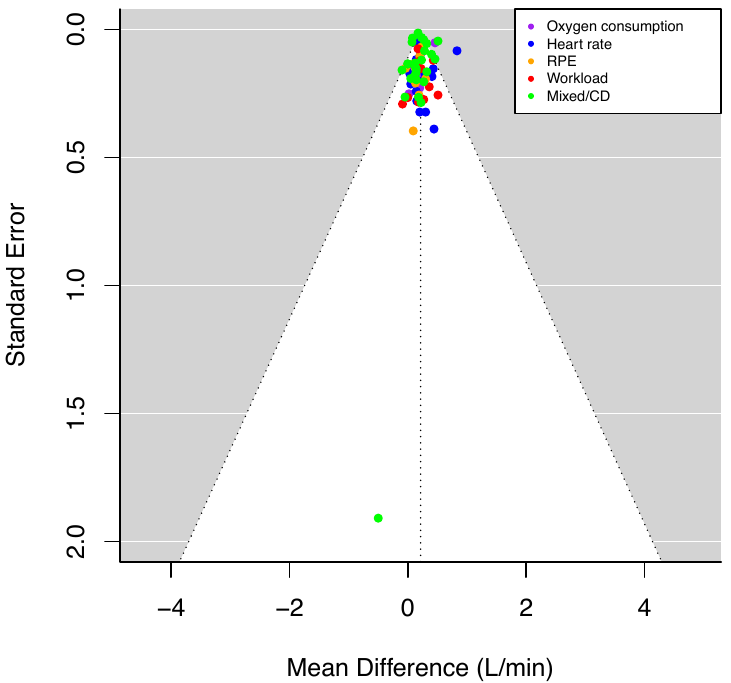

**Figure 41:** Funnel plot of absolute peak oxygen consumption with studies sub-grouped based on method of exercise intensity prescription.

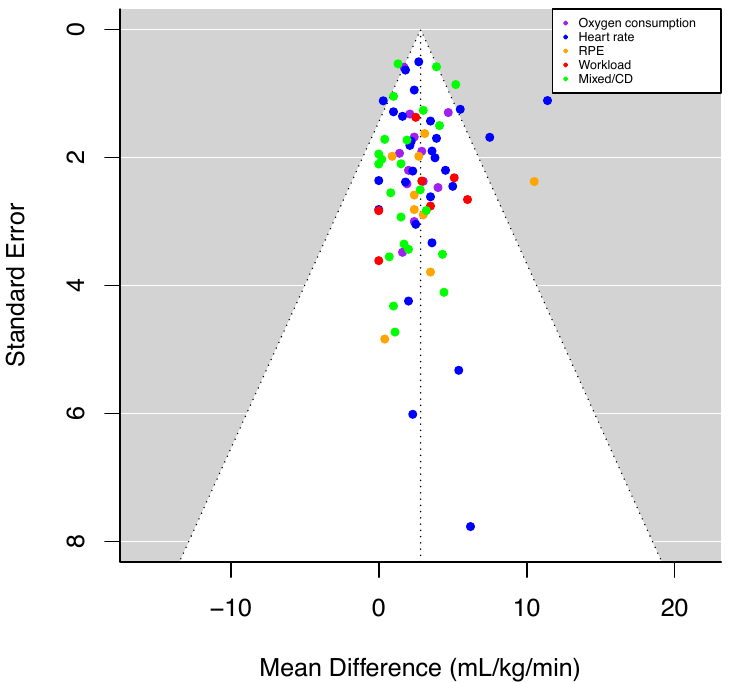

**Figure 42:** Funnel plot of relative peak oxygen consumption with studies sub-grouped based on method of exercise intensity prescription.

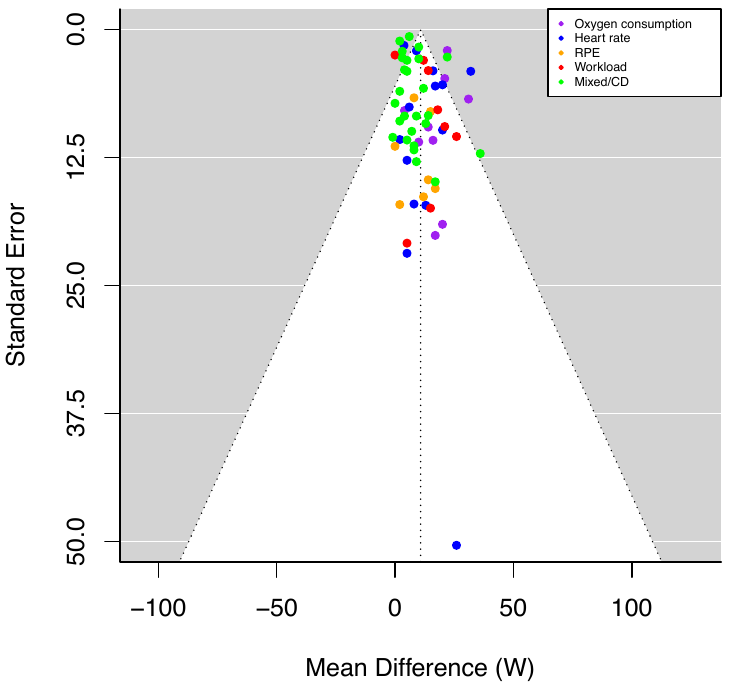

**Figure 43:** Funnel plot of peak power output with studies sub-grouped based on method of exercise intensity prescription.

**FREQUENCY OF EXERCISE SESSIONS**

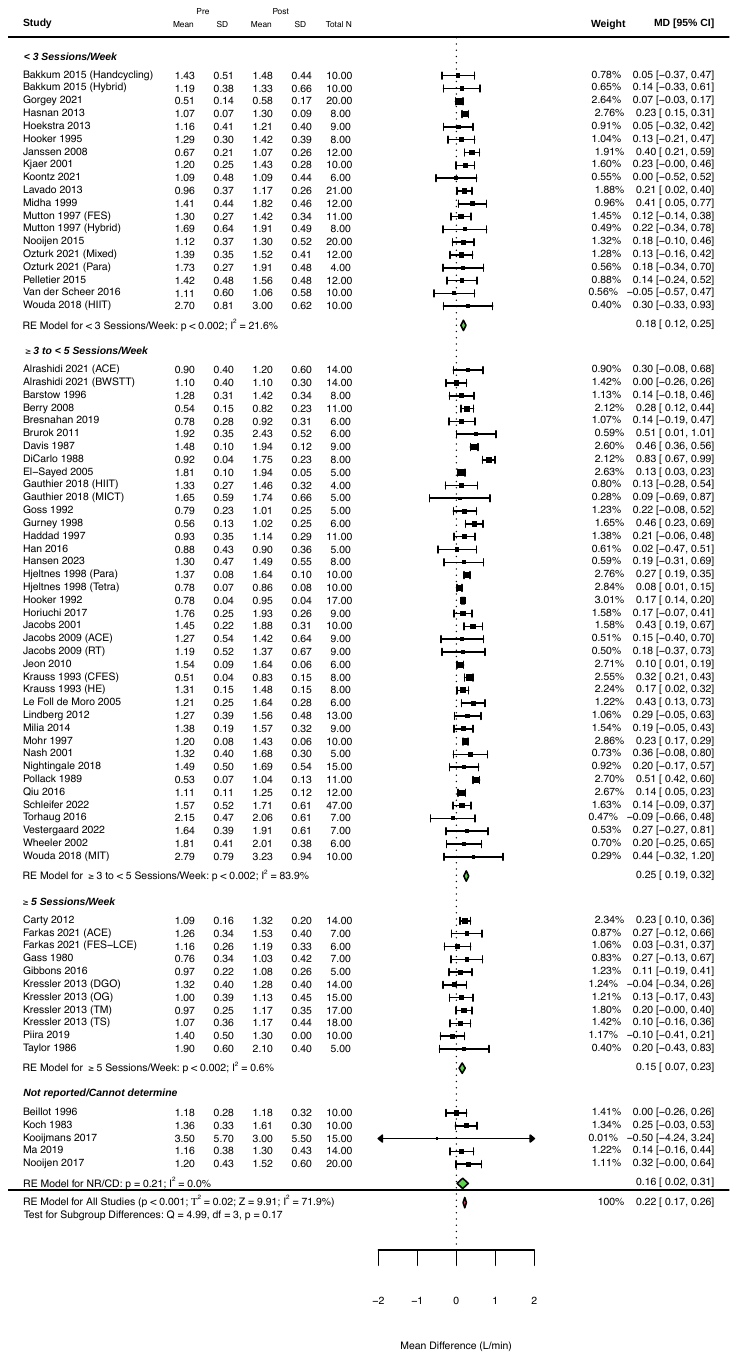

**Figure 44:** Changes in absolute peak oxygen consumption following exercise interventions > 2 weeks, sub-grouped based on frequency of exercise sessions.

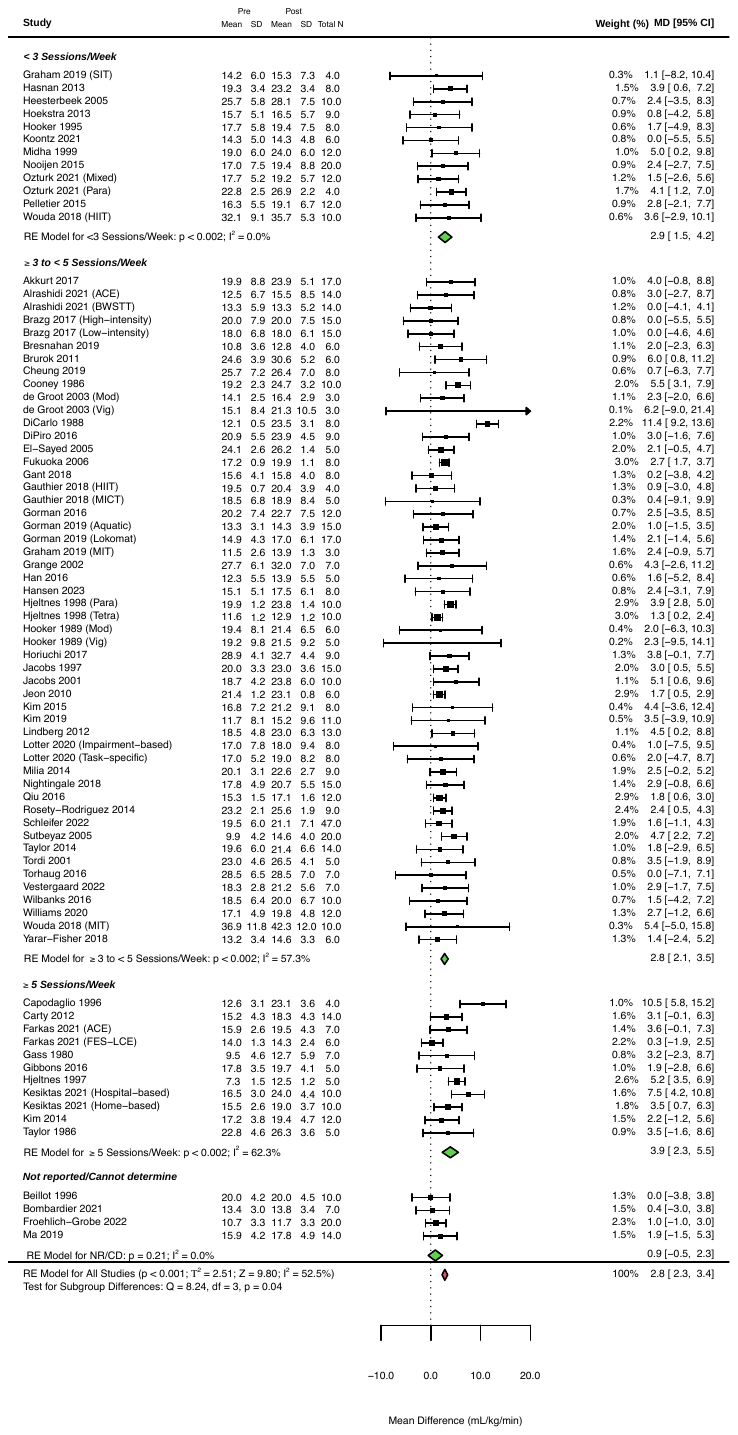

**Figure 45:** Changes in relative peak oxygen consumption following exercise interventions > 2 weeks, sub-grouped based on frequency of exercise sessions.

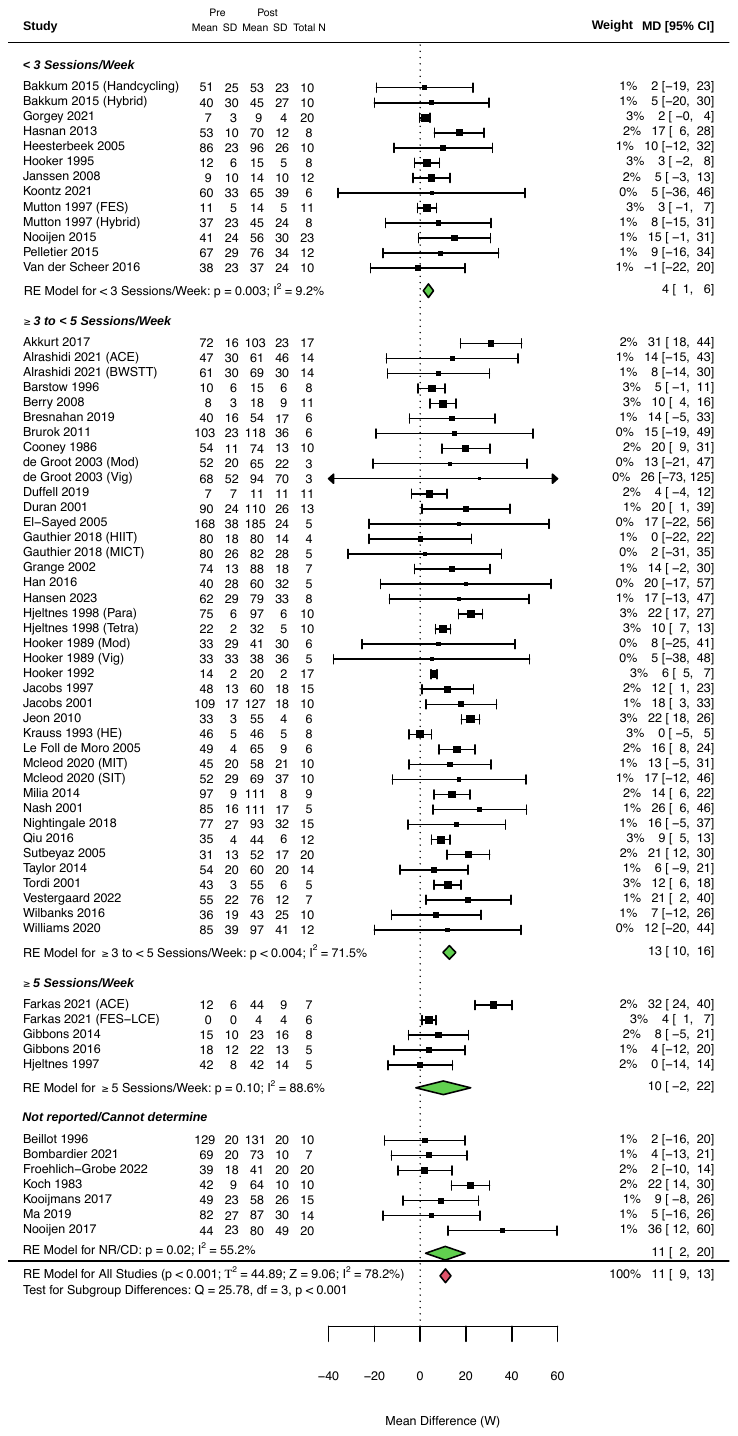

**Figure 46:** Changes in peak power output following exercise interventions > 2 weeks, sub-grouped based on frequency of exercise sessions.

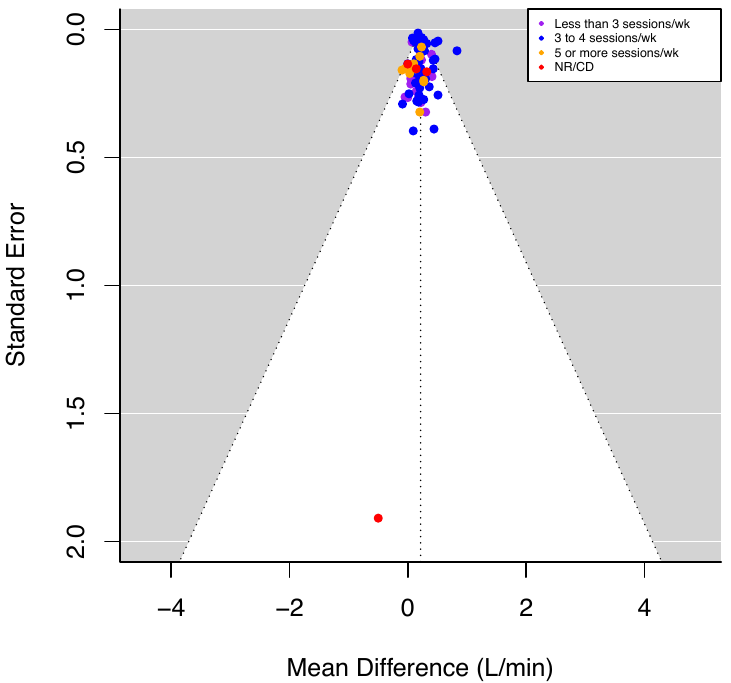

**Figure 47:** Funnel plot of absolute peak oxygen consumption with studies sub-grouped based on frequency of exercise sessions.

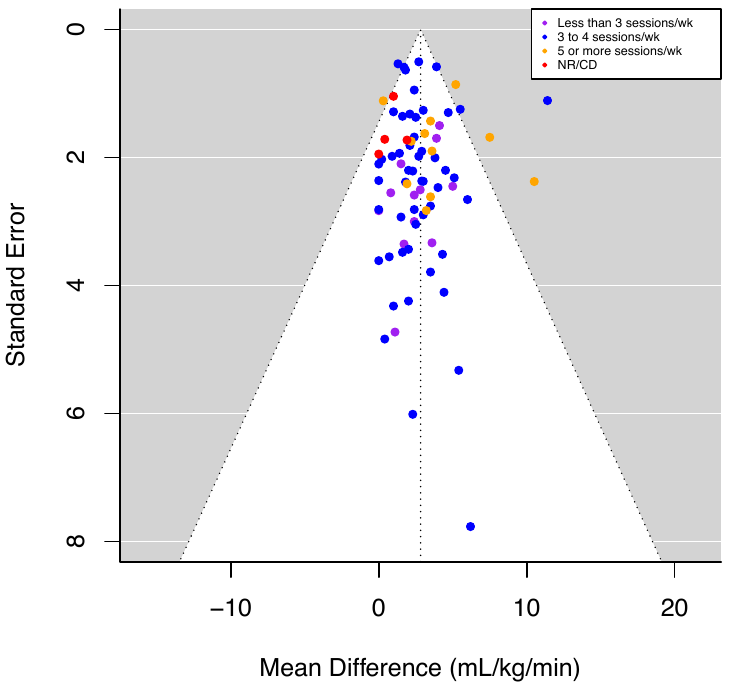

**Figure 48:** Funnel plot of relative peak oxygen consumption with studies sub-grouped based on frequency of exercise sessions.

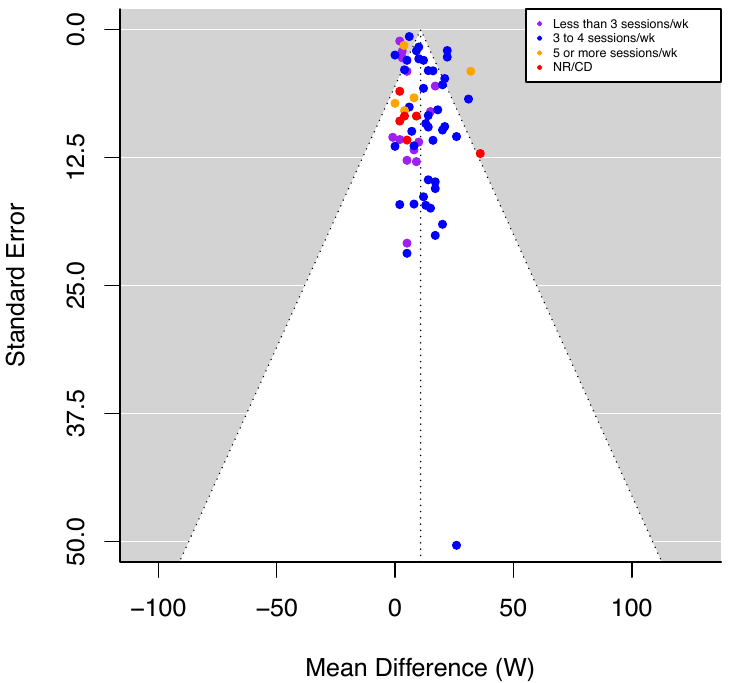

**Figure 49:** Funnel plot of peak power output with studies sub-grouped based on frequency of exercise sessions.

**EXERCISE VOLUME**

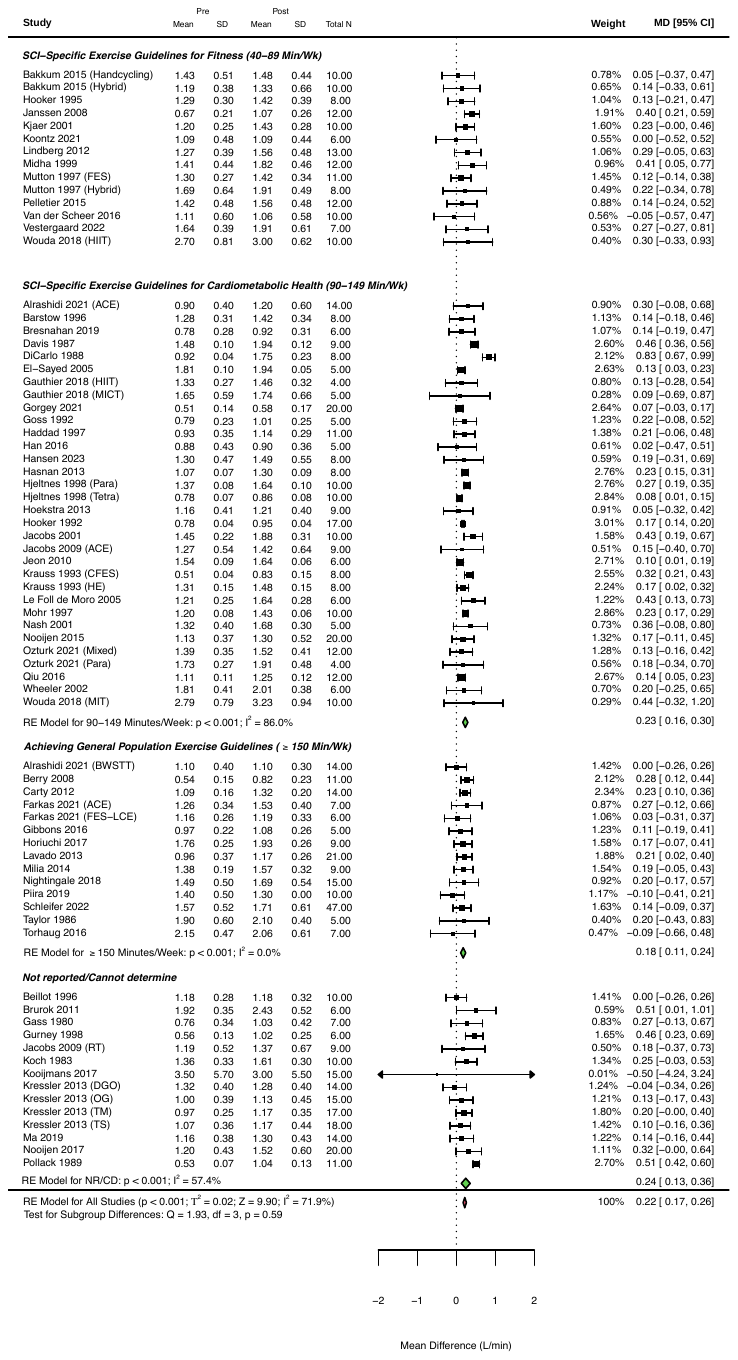
**Figure 50:** Changes in absolute peak oxygen consumption following exercise interventions > 2 weeks, sub-grouped based on volume of exercise.

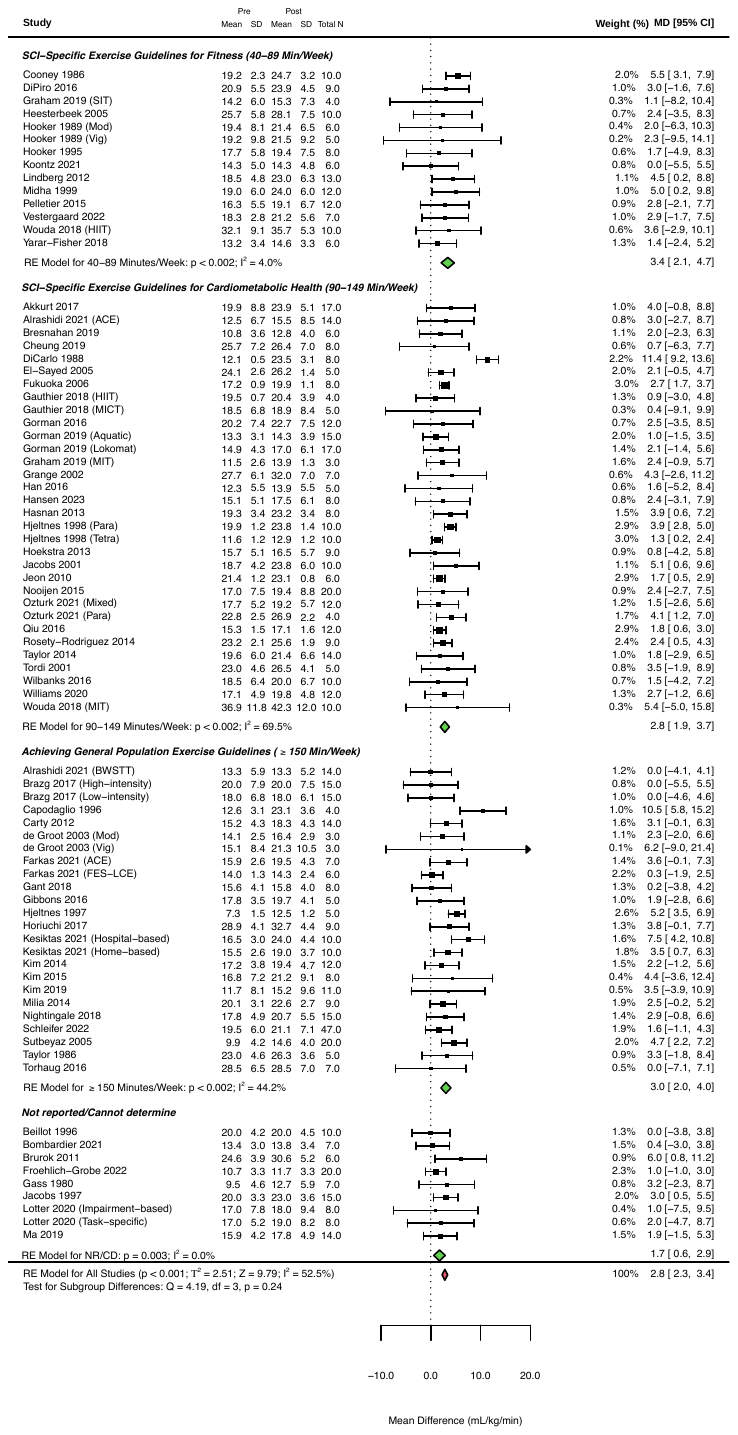

**Figure 51:** Changes in relative peak oxygen consumption following exercise interventions > 2 weeks, sub-grouped based on volume of exercise.

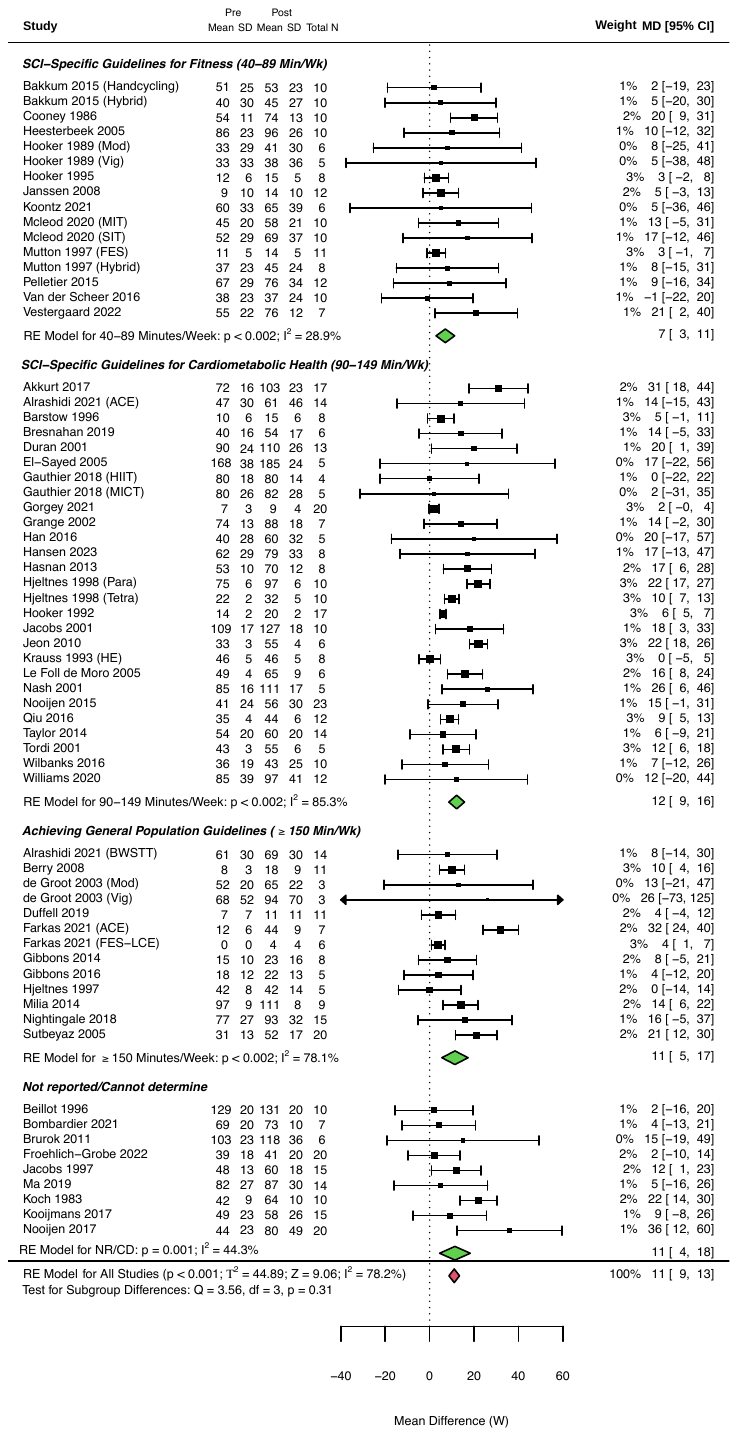

**Figure 52:** Changes in peak power output following exercise interventions > 2 weeks, sub-grouped based on volume of exercise.

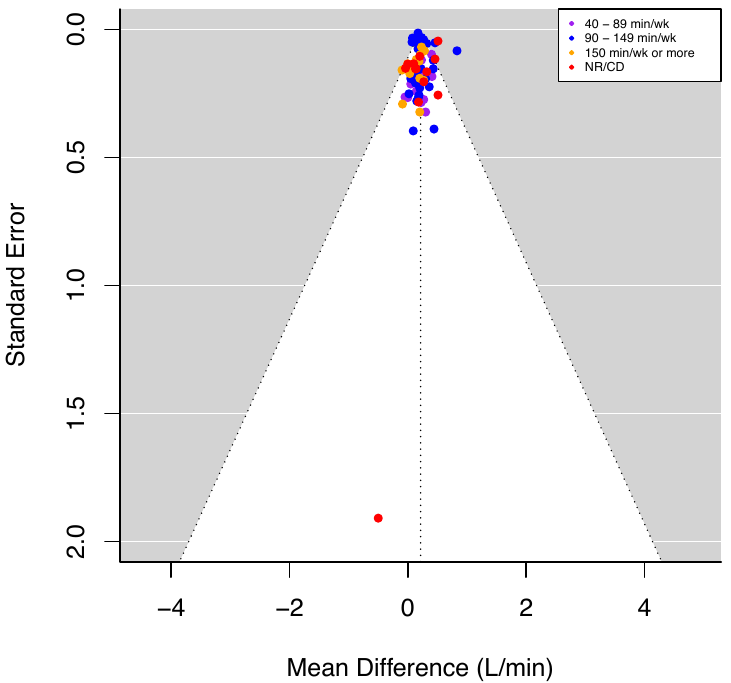

**Figure 53:** Funnel plot of absolute peak oxygen consumption with studies sub-grouped based on volume of exercise.

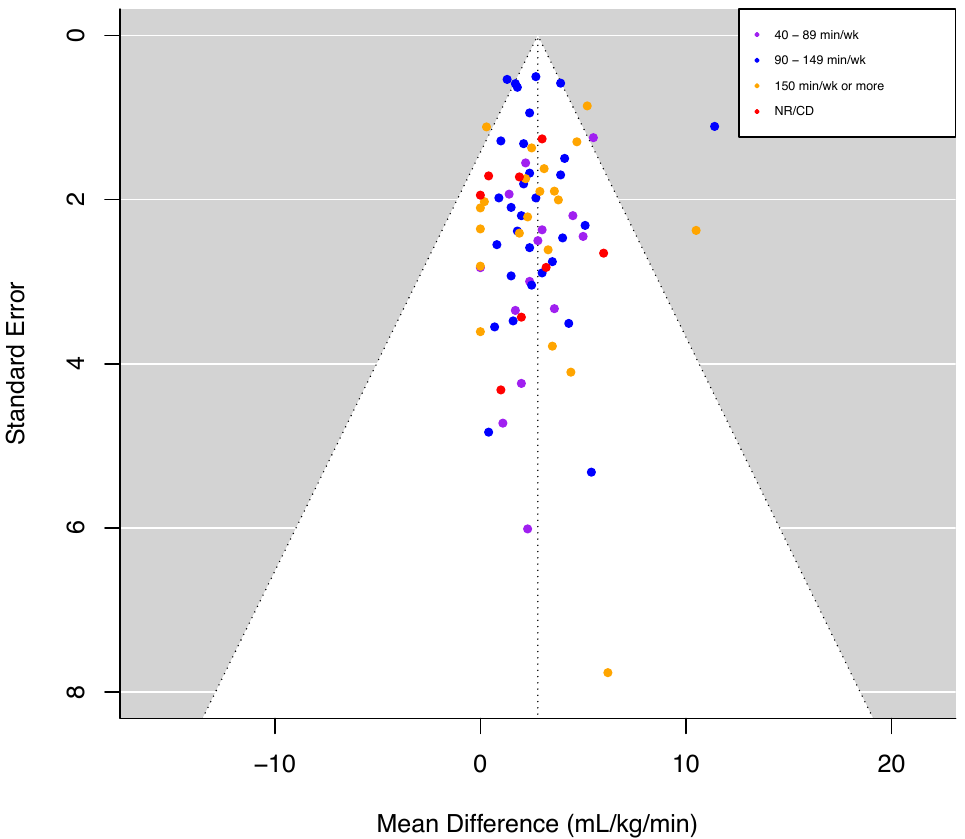

**Figure 54:** Funnel plot of relative peak oxygen consumption with studies sub-grouped based on volume of exercise.

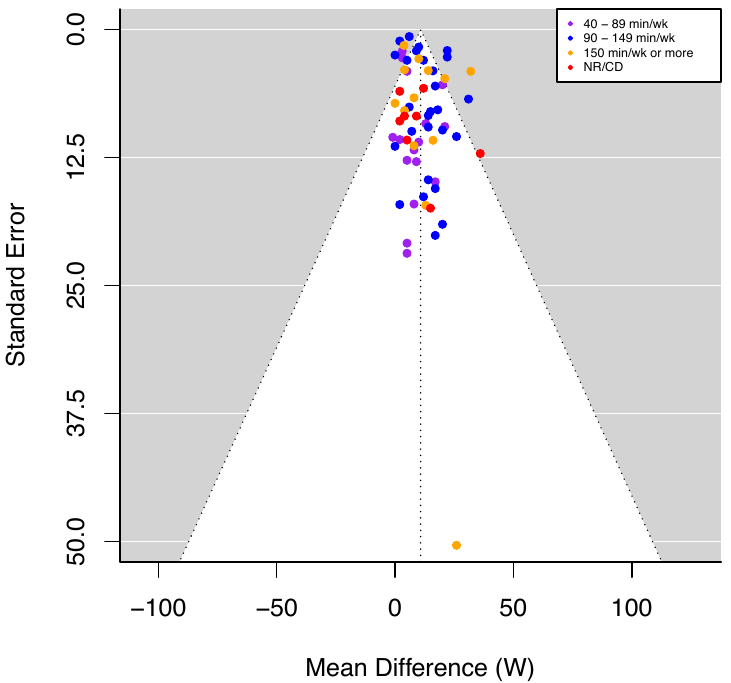

**Figure 55:** Funnel plot of peak power output with studies sub-grouped based on volume of exercise.

**PRE-POST STUDIES QUALITY ASSESSMENT**

| **Table 6.** Risk of bias of pre-post studies assessed via the NIH Quality Assessment Tool for Before-After (Pre-Post) Studies with No Control Group (12 items). | | | | | | | | | | | | | |
| --- | --- | --- | --- | --- | --- | --- | --- | --- | --- | --- | --- | --- | --- |
| **Author (Year)** | **Q1** | **Q2** | **Q3** | **Q4** | **Q5** | **Q6** | **Q7** | **Q8** | **Q9** | **Q10** | **Q11** | **Q12** | **Overall Quality** |
| Alrashidi et al. (2021) | **✓** | **✓** | **✓** | ❌ | **✓** | **✓** | **✓** | ❌ | ❌ | **✓** | ❌ | **N/A** | **GOOD** |
| Bakkum et al. (2015) | **✓** | **✓** | **✓** | **✓** | ❌ | **✓** | **✓** | **?** | ❌ | **✓** | ❌ | **N/A** | **FAIR** |
| Barstow et al. (1996) | **✓** | **✓** | **✓** | **?** | ❌ | **✓** | **✓** | **?** | **✓** | **✓** | ❌ | **N/A** | **FAIR** |
| Beillot et al. (1996) | **✓** | **✓** | **✓** | **?** | ❌ | **✓** | **✓** | **?** | ❌ | **✓** | ❌ | **N/A** | **FAIR** |
| Berry et al. (2008) | **✓** | **✓** | **✓** | **?** | ❌ | **✓** | **✓** | **?** | **✓** | **✓** | ❌ | **N/A** | **GOOD** |
| Bresnahan et al. (2019) | **✓** | **✓** | **✓** | **?** | ❌ | **✓** | **✓** | ❌ | ❌ | **✓** | ❌ | **N/A** | **FAIR** |
| Brurok et al. (2011) | **✓** | **✓** | **✓** | **?** | ❌ | ❌ | **✓** | ❌ | **✓** | **✓** | ❌ | **N/A** | **FAIR** |
| Carty et al. (2012) | **✓** | **✓** | **✓** | **?** | **✓** | **✓** | **✓** | **?** | **✓** | **✓** | ❌ | **N/A** | **GOOD** |
| Cooney and Walker  (1986) | **✓** | **✓** | **✓** | **?** | ❌ | **✓** | **✓** | **?** | **✓** | **✓** | ❌ | **N/A** | **GOOD** |
| Davis et al. (1987) | **✓** | ❌ | ❌ | **?** | ❌ | **?** | **✓** | **?** | **?** | **✓** | ❌ | **N/A** | **POOR** |
| DiCarlo  (1988) | **✓** | **✓** | ❌ | **?** | ❌ | **✓** | **✓** | ❌ | **?** | **✓** | ❌ | **N/A** | **FAIR** |
| DiPiro et al. (2016) | **✓** | **✓** | **✓** | ❌ | ❌ | **✓** | **✓** | ❌ | **✓** | **✓** | ❌ | **N/A** | **GOOD** |
| Duffell et al. (2019) | **✓** | ❌ | **✓** | **✓** | ❌ | **✓** | **✓** | ❌ | **✓** | **✓** | ❌ | **N/A** | **FAIR** |
| Duran et al. (2001) | **✓** | **✓** | **✓** | **?** | **✓** | **✓** | **✓** | ❌ | **✓** | **✓** | ❌ | **N/A** | **GOOD** |
| El-Sayed and Younesian (2005) | **✓** | ❌ | **?** | **?** | ❌ | **✓** | **✓** | ❌ | **?** | **✓** | ❌ | **N/A** | **POOR** |
| Farkas et al. (2021) | **✓** | **✓** | ❌ | **?** | ❌ | **✓** | **✓** | **✓** | **?** | **✓** | ❌ | **N/A** | **FAIR** |
| Fukuoka et al. (2006) | **✓** | ❌ | **✓** | **?** | ❌ | **✓** | **✓** | ❌ | **✓** | **✓** | ❌ | **N/A** | **FAIR** |
| Gant et al. (2018) | **✓** | **✓** | **✓** | **?** | ❌ | **✓** | **✓** | **?** | **✓** | **✓** | ❌ | **N/A** | **GOOD** |
| Gass et al. (1980) | **✓** | ❌ | **✓** | **?** | ❌ | **✓** | **✓** | ❌ | **✓** | **✓** | ❌ | **N/A** | **FAIR** |
| Gibbons et al. (2014) | **✓** | ❌ | **✓** | **?** | ❌ | **✓** | **✓** | ❌ | **✓** | **✓** | ❌ | **N/A** | **FAIR** |
| Gibbons et al. (2016) | **✓** | ❌ | **✓** | **?** | ❌ | **✓** | **✓** | ❌ | **✓** | **✓** | ❌ | **N/A** | **FAIR** |
| Gorman et al. (2019) | **✓** | **✓** | **✓** | **✓** | **✓** | **✓** | **✓** | **?** | **✓** | **✓** | ❌ | **N/A** | **GOOD** |
| Goss et al. (1992) | **✓** | **✓** | **✓** | **?** | ❌ | **✓** | **✓** | ❌ | **✓** | **✓** | ❌ | **N/A** | **GOOD** |
| Grange et al. (2002) | **✓** | ❌ | **✓** | **?** | ❌ | **✓** | **✓** | ❌ | **✓** | **✓** | ❌ | **N/A** | **FAIR** |
| Gurney et al. (1998) | **✓** | ❌ | **✓** | ❌ | ❌ | **?** | **✓** | ❌ | **✓** | **✓** | ❌ | **N/A** | **POOR** |
| Haddad et al. (1997) | **✓** | ❌ | ❌ | **?** | ❌ | **✓** | **✓** | **?** | **?** | **✓** | ❌ | **N/A** | **POOR** |
| Han et al. (2016) | **✓** | **✓** | **✓** | ❌ | ❌ | **✓** | **✓** | ❌ | ❌ | **✓** | ❌ | **N/A** | **FAIR** |
| Hasnan et al. (2013) | **✓** | ❌ | **?** | **?** | ❌ | **✓** | ❌ | ❌ | **?** | **✓** | ❌ | **N/A** | **POOR** |
| Heesterbeek et al. (2005) | **✓** | ❌ | ❌ | **?** | ❌ | **✓** | **✓** | ❌ | **✓** | **✓** | ❌ | **N/A** | **POOR** |
| Hjeltnes et al. (1997) | **✓** | ❌ | **?** | **?** | ❌ | **✓** | **✓** | **?** | **✓** | **✓** | ❌ | **N/A** | **POOR** |
| Hjeltnes and Wallberg-  Henriksson  (1998) | **✓** | **✓** | **✓** | **?** | **✓** | **✓** | **✓** | **?** | **?** | **✓** | ❌ | **N/A** | **GOOD** |
| Hoekstra et al. (2013) | **✓** | **✓** | **✓** | **?** | ❌ | **✓** | **✓** | **?** | **✓** | **✓** | ❌ | **N/A** | **GOOD** |
| Hooker et al. (1992) | **✓** | **?** | **✓** | **?** | **✓** | **✓** | **✓** | **?** | **?** | **✓** | ❌ | **N/A** | **FAIR** |
| Hooker et al. (1995) | **✓** | ❌ | **✓** | **?** | ❌ | **✓** | **✓** | **?** | **✓** | **✓** | ❌ | **N/A** | **FAIR** |
| Horiuchi et al. (2017) | **✓** | **✓** | **✓** | **?** | **✓** | **✓** | **✓** | **?** | **✓** | **✓** | ❌ | **N/A** | **GOOD** |
| Jacobs et al. (1997) | **✓** | **?** | **✓** | **?** | **✓** | **✓** | **✓** | **?** | **?** | **✓** | ❌ | **N/A** | **FAIR** |
| Jacobs et al. (2001) | **✓** | **✓** | **?** | **?** | ❌ | **✓** | **✓** | **?** | **✓** | **✓** | ❌ | **N/A** | **GOOD** |
| Jacobs  (2009) | **✓** | ❌ | ❌ | **?** | ❌ | **✓** | **✓** | **?** | **?** | **✓** | ❌ | **N/A** | **POOR** |
| Janssen et al. (2008) | **✓** | ❌ | **✓** | **?** | **✓** | **✓** | **✓** | **?** | **✓** | **✓** | ❌ | **N/A** | **FAIR** |
| Jeon et al. (2010) | **✓** | ❌ | ❌ | **?** | ❌ | **✓** | **✓** | **?** | ❌ | **✓** | ❌ | **N/A** | **POOR** |
| Kesiktas et al. (2021) | **✓** | **✓** | ❌ | ❌ | **✓** | ❌ | **✓** | **?** | **✓** | **✓** | ❌ | **N/A** | **FAIR** |
| Kim et al. (2014) | **✓** | **✓** | **✓** | **?** | **✓** | **✓** | **✓** | **?** | **✓** | **✓** | ❌ | **N/A** | **GOOD** |
| Kjaer et al. (2001) | **✓** | ❌ | **✓** | **?** | ❌ | **✓** | **✓** | **?** | **✓** | **✓** | ❌ | **N/A** | **FAIR** |
| Koch et al. (1983) | ❌ | ❌ | **✓** | **?** | ❌ | ❌ | **✓** | **?** | **?** | ❌ | ❌ | **N/A** | **POOR** |
| Koontz et al. (2021) | **✓** | **✓** | **✓** | **✓** | ❌ | **✓** | **✓** | **?** | ❌ | **✓** | ❌ | **N/A** | **FAIR** |
| Krauss et al. (1993) | **✓** | **✓** | **✓** | **✓** | ❌ | **✓** | **✓** | **?** | **✓** | **✓** | ❌ | **N/A** | **GOOD** |
| Kressler et al. (2013) | **✓** | **✓** | **✓** | ❌ | **✓** | **✓** | **✓** | **?** | **✓** | **✓** | ❌ | **N/A** | **GOOD** |
| Le Foll-de Moro et al. (2005) | **✓** | ❌ | **✓** | **?** | ❌ | **✓** | **✓** | **?** | **✓** | **✓** | ❌ | **N/A** | **FAIR** |
| Lindberg et al. (2012) | **✓** | **✓** | **?** | **?** | **✓** | **✓** | **✓** | **?** | **✓** | **✓** | ❌ | **N/A** | **GOOD** |
| Lotter et al. (2020) | **✓** | **✓** | **✓** | **?** | **✓** | **✓** | **✓** | **?** | **✓** | **✓** | ❌ | **N/A** | **GOOD** |
| Midha et al. (1999) | **✓** | ❌ | **?** | **?** | **✓** | **✓** | **✓** | **?** | **✓** | **✓** | ❌ | **N/A** | **GOOD** |
| Milia et al. (2014) | **✓** | ❌ | **✓** | **?** | ❌ | **✓** | **✓** | **?** | **✓** | **✓** | ❌ | **N/A** | **FAIR** |
| Mohr et al. (1997) | **✓** | ❌ | **✓** | **?** | ❌ | **✓** | **✓** | **?** | **✓** | **✓** | ❌ | **N/A** | **FAIR** |
| Mutton et al. (1997) | **✓** | ❌ | **✓** | **?** | ❌ | **✓** | **✓** | **?** | ❌ | **✓** | ❌ | **N/A** | **POOR** |
| Nash et al. (2001) | **✓** | **✓** | ❌ | **?** | ❌ | **✓** | **✓** | **?** | **?** | **✓** | ❌ | **N/A** | **POOR** |
| Nooijen et al. (2015) | **✓** | **✓** | **✓** | **?** | **✓** | **✓** | **✓** | ❌ | ❌ | **✓** | ❌ | **N/A** | **GOOD** |
| Ozturk et al. (2021) | **✓** | **✓** | **✓** | **?** | **✓** | **✓** | **✓** | ❌ | ❌ | **✓** | ❌ | **N/A** | **GOOD** |
| Pollack et al. (1989) | ❌ | **✓** | **✓** | **?** | ❌ | **?** | **✓** | **?** | **✓** | **✓** | ❌ | **N/A** | **POOR** |
| Qiu et al. (2016) | **✓** | **✓** | **✓** | **?** | **✓** | **✓** | **✓** | **?** | **✓** | **✓** | ❌ | **N/A** | **GOOD** |
| Schleifer et al. (2022) | **✓** | ❌ | **✓** | **?** | **✓** | **✓** | **✓** | **?** | **?** | **✓** | ❌ | **N/A** | **FAIR** |
| Sutbeyaz et al. (2005) | **✓** | **✓** | **✓** | **?** | **✓** | ❌ | **✓** | **?** | **✓** | **✓** | ❌ | **N/A** | **GOOD** |
| Taylor et al. (2014) | **✓** | ❌ | **✓** | **?** | **✓** | **✓** | **✓** | **?** | **✓** | **✓** | ❌ | **N/A** | **GOOD** |
| Tordi et al. (2001) | **✓** | ❌ | **?** | **?** | ❌ | **✓** | **✓** | **?** | **✓** | **✓** | ❌ | **N/A** | **POOR** |
| Vestergaard et al. (2022) | **✓** | **✓** | **✓** | **?** | ❌ | **✓** | **✓** | ❌ | **✓** | ❌ | ❌ | **N/A** | **FAIR** |
| Wheeler et al. (2002) | **✓** | ❌ | **✓** | **?** | ❌ | **✓** | **✓** | **?** | **✓** | **✓** | ❌ | **N/A** | **FAIR** |
| Wilbanks et al. (2016) | **✓** | **✓** | **✓** | **?** | ❌ | **✓** | **✓** | **?** | **✓** | **✓** | ❌ | **N/A** | **GOOD** |
| Williams et al. (2020) | **✓** | **✓** | **✓** | **?** | **✓** | **✓** | **✓** | **?** | **✓** | **✓** | ❌ | **N/A** | **GOOD** |
| **Total ✓** | **65** | **37** | **51** | **5** | **22** | **60** | **66** | **1** | **44** | **65** | **0** | **-** | **26 GOOD**  **27 FAIR**  **14 POOR** |
| **Total ❌** | **2** | **28** | **9** | **6** | **45** | **4** | **1** | **21** | **10** | **2** | **67** | **-** |  |
| **Total ?** | **0** | **2** | **7** | **56** | **0** | **3** | **0** | **45** | **13** | **0** | **0** | **-** |  |
| **✓** = yes, ❌ = no, **?** = cannot determine/not reported | | | | | | | | | | | | | |

**Quality Assessment Questions**

1. Was the study question or objective clearly stated?
2. Were eligibility/selection criteria for the study population prespecified and clearly described?
3. Were the participants in the study representative of those who would be eligible for the test/service/intervention in the general or clinical population of interest?
4. Were all eligible participants that met the prespecified entry criteria enrolled?
5. Was the sample size sufficiently large to provide confidence in the findings?
6. Was the test/service/intervention clearly described and delivered consistently across the study population?
7. Were the outcome measures prespecified, clearly defined, valid, reliable, and assessed consistently across all study participants?
8. Were the people assessing the outcomes blinded to the participants' exposures/interventions?
9. Was the loss to follow-up after baseline 20% or less? Were those lost to follow-up accounted for in the analysis?
10. Did the statistical methods examine changes in outcome measures from before to after the intervention? Were statistical tests done that provided p values for the pre-to-post changes?
11. Were outcome measures of interest taken multiple times before the intervention and multiple times after the intervention (i.e., did they use an interrupted time-series design)?

**RCTs RISK OF BIAS**

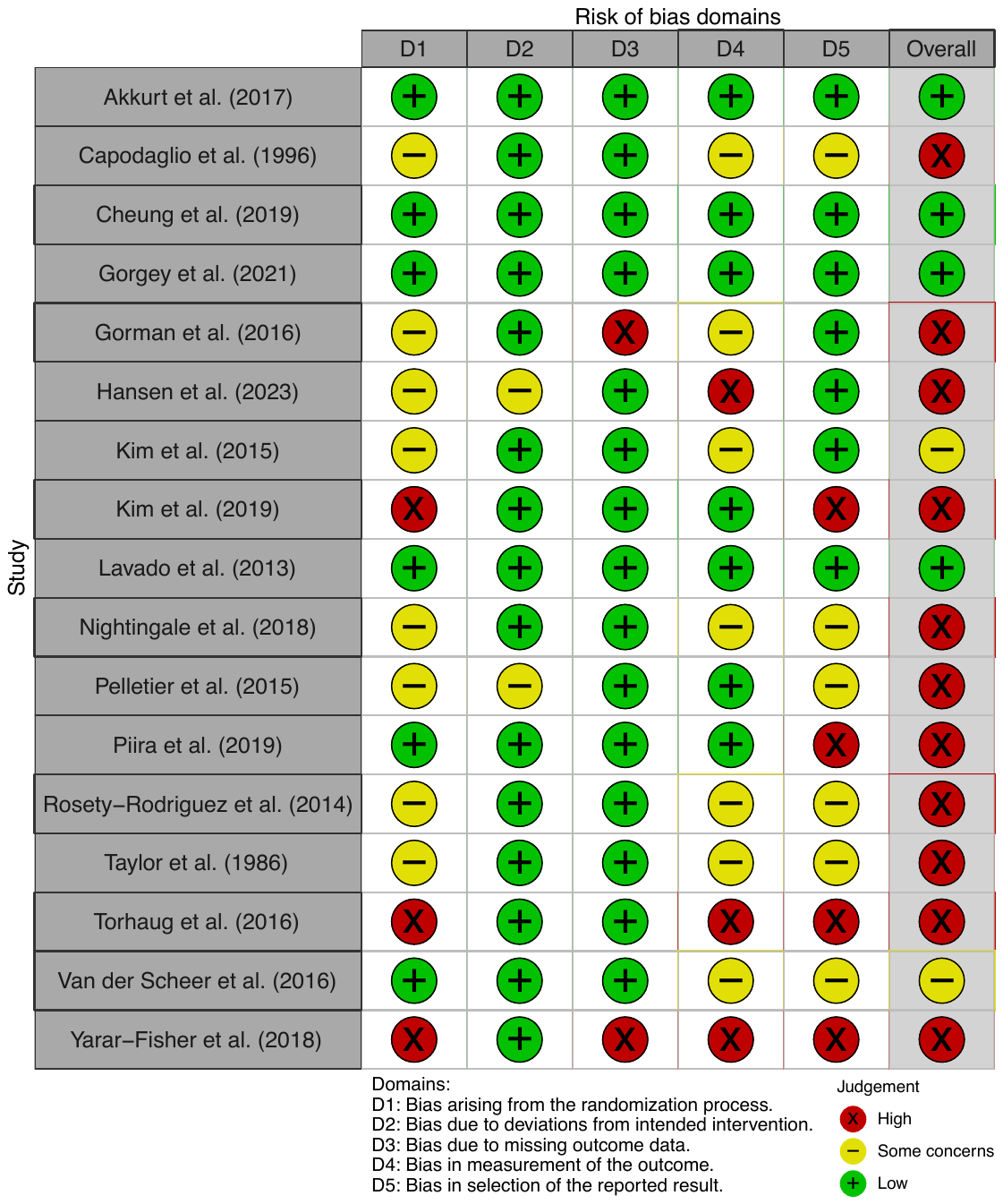

**Figure 56:** Traffic plot for risk of bias in the individual RCTs, assessed via the Cochrane RoB 2 tool.

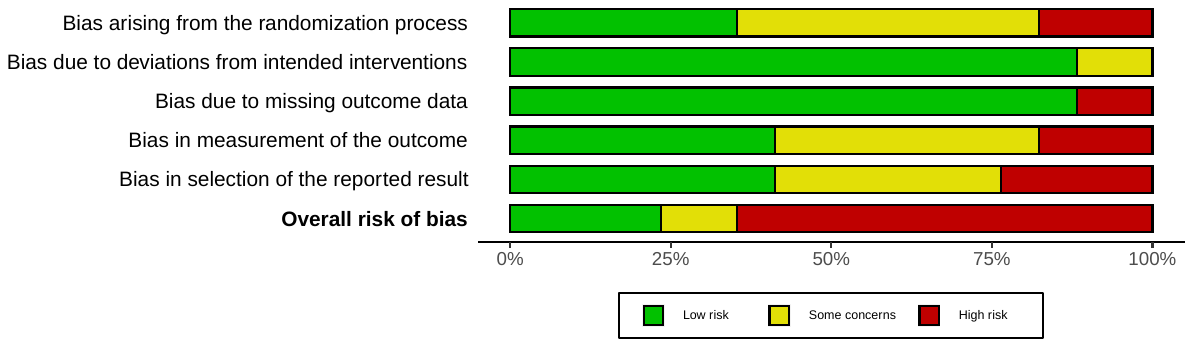

**Figure 57:** Summary plot for risk of bias in the individual RCTs, assessed via the Cochrane RoB 2 tool.

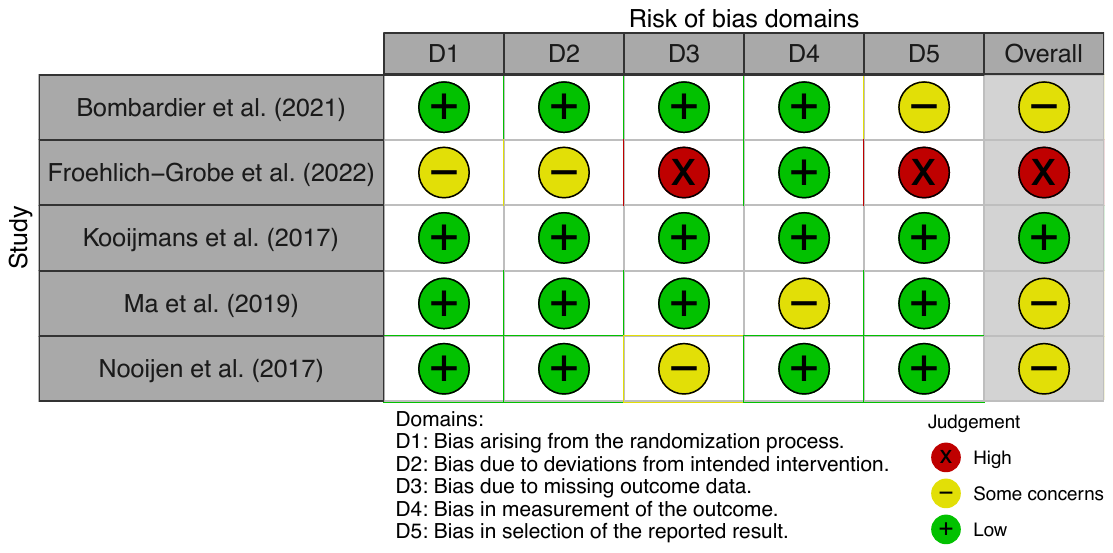

**Figure 58:** Traffic plot for risk of bias in the behaviour change RCTs, assessed via the Cochrane RoB 2 tool.

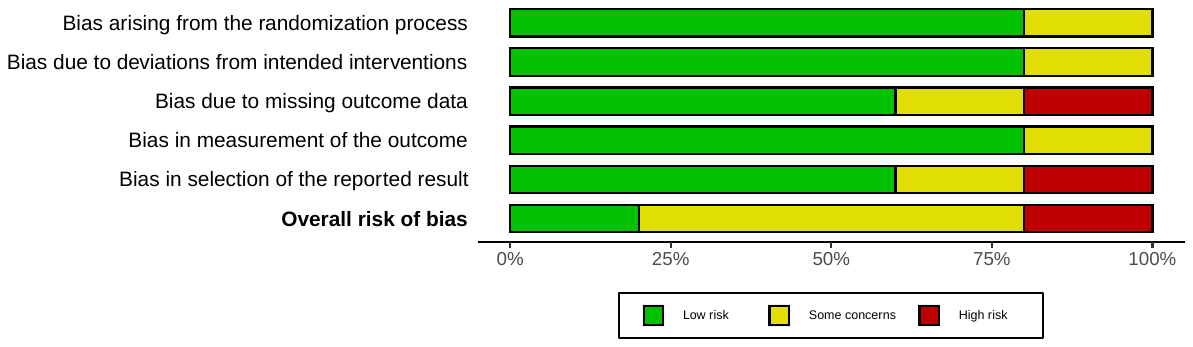

**Figure 59:** Summary plot for risk of bias in the behaviour change RCTs, assessed via the Cochrane RoB 2 tool.

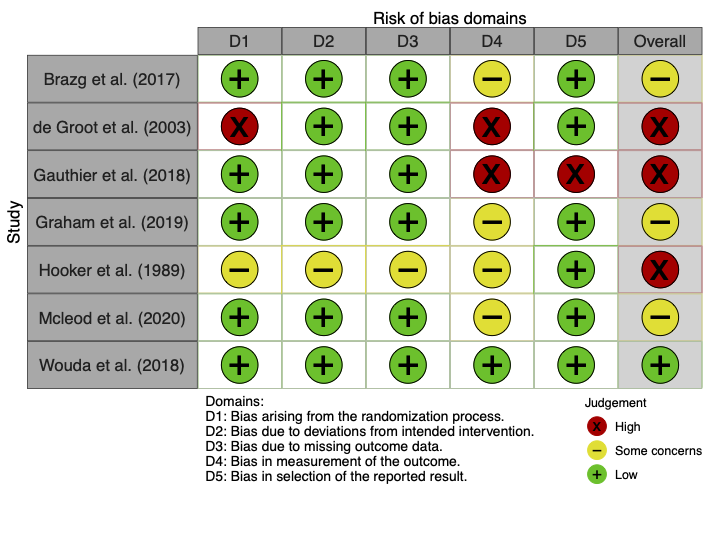

**Figure 60:** Traffic plot for risk of bias in the intensity comparison RCTs, assessed via the Cochrane RoB 2 tool.

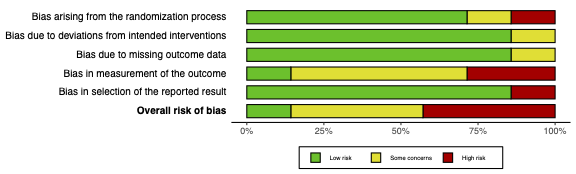

**Figure 61:** Summary plot for risk of bias in the intensity comparison RCTs, assessed via the Cochrane RoB 2 tool.

**References**

Pre-post and RCT studies included in the primary meta-analysis, sorted alphabetically:

Akkurt H, Karapolat HU, Kirazli Y, Kose T. The effects of upper extremity aerobic exercise in patients with spinal cord injury: a randomized controlled study. Eur J Phys Rehabil Med. 2017;53: 219–227.

Alrashidi AA, Nightingale TE, Currie KD, Hubli M, MacDonald MJ, Hicks AL, et al. Exercise Improves Cardiorespiratory Fitness, but Not Arterial Health, after Spinal Cord Injury: The CHOICES Trial. J Neurotrauma. 2021;38: 3020–3029.

Bakkum AJT, de Groot S, Stolwijk-Swüste JM, van Kuppevelt DJ, ALLRISC, van der Woude LHV, et al. Effects of hybrid cycling versus handcycling on wheelchair-specific fitness and physical activity in people with long-term spinal cord injury: a 16-week randomized controlled trial. Spinal Cord. 2015;53: 395–401.

Barstow TJ, Scremin AM, Mutton DL, Kunkel CF, Cagle TG, Whipp BJ. Changes in gas exchange kinetics with training in patients with spinal cord injury. Med Sci Sports Exerc. 1996;28: 1221–1228.

Beillot J, Carré F, Le Claire G, Thoumie P, Perruoin-Verbe B, Cormerais A, et al. Energy consumption of paraplegic locomotion using reciprocating gait orthosis. Eur J Appl Physiol Occup Physiol. 1996;73: 376–381.

Berry HR, Perret C, Saunders BA, Kakebeeke TH, Donaldson NDN, Allan DB, et al. Cardiorespiratory and power adaptations to stimulated cycle training in paraplegia. Med Sci Sports Exerc. 2008;40: 1573–1580.

Bombardier CH, Dyer JR, Burns P, Crane DA, Takahashi MM, Barber J, et al. A tele-health intervention to increase physical fitness in people with spinal cord injury and cardiometabolic disease or risk factors: a pilot randomized controlled trial. Spinal Cord. 2021;59: 63–73.

Brazg G, Fahey M, Holleran CL, Connolly M, Woodward J, Hennessy PW, et al. Effects of Training Intensity on Locomotor Performance in Individuals With Chronic Spinal Cord Injury: A Randomized Crossover Study. Neurorehabil Neural Repair. 2017;31: 944–954.

Bresnahan JJ, Farkas GJ, Clasey JL, Yates JW, Gater DR. Arm crank ergometry improves cardiovascular disease risk factors and community mobility independent of body composition in high motor complete spinal cord injury. J Spinal Cord Med. 2019;42: 272–280.

Brurok B, Helgerud J, Karlsen T, Leivseth G, Hoff J. Effect of aerobic high-intensity hybrid training on stroke volume and peak oxygen consumption in men with spinal cord injury. Am J Phys Med Rehabil. 2011;90: 407–414.

Capodaglio P, Grilli C, Bazzini G. Tolerable exercise intensity in the early rehabilitation of paraplegic patients. A preliminary study. Spinal Cord. 1996;34: 684–690.

Carty A, McCormack K, Coughlan GF, Crowe L, Caulfield B. Increased aerobic fitness after neuromuscular electrical stimulation training in adults with spinal cord injury. Arch Phys Med Rehabil. 2012;93: 790–795.

Cheung EYY, Yu KKK, Kwan RLC, Ng CKM, Chau RMW, Cheing GLY. Effect of EMG-biofeedback robotic-assisted body weight supported treadmill training on walking ability and cardiopulmonary function on people with subacute spinal cord injuries - a randomized controlled trial. BMC Neurol. 2019;19: 140.

Cooney MM, Walker JB. Hydraulic resistance exercise benefits cardiovascular fitness of spinal cord injured. Med Sci Sports Exerc. 1986;18: 522–525.

Davis GM, Shephard RJ, Leenen FH. Cardiac effects of short term arm crank training in paraplegics: echocardiographic evidence. Eur J Appl Physiol Occup Physiol. 1987;56: 90–96.

de Groot PCE, Hjeltnes N, Heijboer AC, Stal W, Birkeland K. Effect of training intensity on physical capacity, lipid profile and insulin sensitivity in early rehabilitation of spinal cord injured individuals. Spinal Cord. 2003;41: 673–679.

DiCarlo SE. Effect of arm ergometry training on wheelchair propulsion endurance of individuals with quadriplegia. Phys Ther. 1988;68: 40–44.

DiPiro ND, Embry AE, Fritz SL, Middleton A, Krause JS, Gregory CM. Effects of aerobic exercise training on fitness and walking-related outcomes in ambulatory individuals with chronic incomplete spinal cord injury. Spinal Cord. 2016;54: 675–681.

Duffell LD, Paddison S, Alahmary AF, Donaldson N, Burridge J. The effects of FES cycling combined with virtual reality racing biofeedback on voluntary function after incomplete SCI: a pilot study. J Neuroeng Rehabil. 2019;16: 149.

Durán FS, Lugo L, Ramírez L, Eusse E. Effects of an exercise program on the rehabilitation of patients with spinal cord injury. Arch Phys Med Rehabil. 2001;82: 1349–1354.

El-Sayed MS, Younesian A. Lipid profiles are influenced by arm cranking exercise and training in individuals with spinal cord injury. Spinal Cord. 2005;43: 299–305.

Farkas GJ, Gorgey AS, Dolbow DR, Berg AS, Gater DR Jr. Energy Expenditure, Cardiorespiratory Fitness, and Body Composition Following Arm Cycling or Functional Electrical Stimulation Exercises in Spinal Cord Injury: A 16-Week Randomized Controlled Trial. Top Spinal Cord Inj Rehabil. 2021;27: 121–134.

Froehlich-Grobe K, Lee J, Ochoa C, Lopez A, Sarker E, Driver S et al. Effectiveness and feasibility of the workout on wheels internet intervention (WOWii) for individuals with spinal cord injury: a randomized controlled trial. Spinal Cord. 2022 Oct;60(10):862-874.

Fukuoka Y, Nakanishi R, Ueoka H, Kitano A, Takeshita K, Itoh M. Effects of wheelchair training on VO2 kinetics in the participants with spinal-cord injury. Disabil Rehabil Assist Technol. 2006;1: 167–174.

Gant KL, Nagle KG, Cowan RE, Field-Fote EC, Nash MS, Kressler J, et al. Body System Effects of a Multi-Modal Training Program Targeting Chronic, Motor Complete Thoracic Spinal Cord Injury. J Neurotrauma. 2018;35: 411–423.

Gass GC, Watson J, Camp EM, Court HJ, McPherson LM, Redhead P. The effects of physical training on high level spinal lesion patients. Scand J Rehabil Med. 1980;12: 61–65.

Gauthier C, Brosseau R, Hicks AL, Gagnon DH. Feasibility, Safety, and Preliminary Effectiveness of a Home-Based Self-Managed High-Intensity Interval Training Program Offered to Long-Term Manual Wheelchair Users. Rehabil Res Pract. 2018;2018: 8209360.

Gibbons RS, Shave RE, Gall A, Andrews BJ. FES-rowing in tetraplegia: a preliminary report. Spinal Cord. 2014;52: 880–886.

Gibbons RS, Stock CG, Andrews BJ, Gall A, Shave RE. The effect of FES-rowing training on cardiac structure and function: pilot studies in people with spinal cord injury. Spinal Cord. 2016;54: 822–829.

Gorgey AS, Lai RE, Khalil RE, Rivers J, Cardozo C, Chen Q, et al. Neuromuscular electrical stimulation resistance training enhances oxygen uptake and ventilatory efficiency independent of mitochondrial complexes after spinal cord injury: a randomized clinical trial. J Appl Physiol. 2021;131: 265–276.

Gorman PH, Scott W, York H, Theyagaraj M, Price-Miller N, McQuaid J, et al. Robotically assisted treadmill exercise training for improving peak fitness in chronic motor incomplete spinal cord injury: A randomized controlled trial. J Spinal Cord Med. 2016;39: 32–44.

Gorman PH, Scott W, VanHiel L, Tansey KE, Sweatman WM, Geigle PR. Comparison of peak oxygen consumption response to aquatic and robotic therapy in individuals with chronic motor incomplete spinal cord injury: a randomized controlled trial. Spinal Cord. 2019;57: 471–481.

Goss FL, McDermott A, Robertson RJ. Changes in peak oxygen uptake following computerized functional electrical stimulation in the spinal cord injured. Res Q Exerc Sport. 1992;63: 76–79.

Graham K, Yarar-Fisher C, Li J, McCully KM, Rimmer JH, Powell D, et al. Effects of High-Intensity Interval Training Versus Moderate-Intensity Training on Cardiometabolic Health Markers in Individuals With Spinal Cord Injury: A Pilot Study. Top Spinal Cord Inj Rehabil. 2019;25: 248–259.

Grange CC, Bougenot MP, Groslambert A, Tordi N, Rouillon JD. Perceived exertion and rehabilitation with wheelchair ergometer: comparison between patients with spinal cord injury and healthy subjects. Spinal Cord. 2002;40: 513–518.

Gurney AB, Robergs RA, Aisenbrey J, Cordova JC, McClanahan L. Detraining from total body exercise ergometry in individuals with spinal cord injury. Spinal Cord. 1998;36: 782–789.

Haddad S, Silva PR, Barretto AC, Ferraretto I. [The effects of aerobic physical training of short duration using upper limbs in paraplegic persons with mild hypertension]. Arq Bras Cardiol. 1997;69(3):169-173.

Han D-S, Hsiao M-Y, Wang T-G, Chen S-Y, Yang W-S. Association of serum myokines and aerobic exercise training in patients with spinal cord injury: an observational study. BMC Neurol. 2016;16: 142.

Hansen RK, Samani A, Laessoe U, Handberg A, Mellergaard M, Figlewski K et al. Rowing exercise increases cardiorespiratory fitness and brachial artery diameter but not traditional cardiometabolic risk factors in spinal cord-injured humans. Eur J Appl Physiol. 2023; 1-15. Epub ahead of print.

Hasnan N, Engkasan JP, Husain R, Davis GM. High-Intensity Virtual-reality Arm plus FES-leg Interval Training in Individuals with Spinal Cord Injury. Biomed Tech. 2013;58. Suppl 1

Heesterbeek PJC, Berkelmans HWA, Thijssen DHJ, van Kuppevelt HJM, Hopman MTE, Duysens J. Increased physical fitness after 4-week training on a new hybrid FES-cycle in persons with spinal cord injury. Technol Disabil. 2005;17: 103–110.

Hjeltnes N, Aksnes AK, Birkeland KI, Johansen J, Lannem A, Wallberg-Henriksson H. Improved body composition after 8 wk of electrically stimulated leg cycling in tetraplegic patients. Am J Physiol. 1997;273: R1072–9.

Hjeltnes N, Wallberg-Henriksson H. Improved work capacity but unchanged peak oxygen uptake during primary rehabilitation in tetraplegic patients. Spinal Cord. 1998;36: 691–698.

Hoekstra F, van Nunen MPM, Gerrits KHL, Stolwijk-Swüste JM, Crins MHP, Janssen TWJ. Effect of robotic gait training on cardiorespiratory system in incomplete spinal cord injury. J Rehabil Res Dev. 2013;50: 1411–1422.

Hooker SP, Wells CL. Effects of low- and moderate-intensity training in spinal cord-injured persons. Med Sci Sports Exerc. 1989;21: 18–22.

Hooker SP, Figoni SF, Rodgers MM, Glaser RM, Mathews T, Suryaprasad AG, et al. Physiologic effects of electrical stimulation leg cycle exercise training in spinal cord injured persons. Arch Phys Med Rehabil. 1992;73: 470–476.

Hooker SP, Scremin AM, Mutton DL, Kunkel CF, Cagle G. Peak and submaximal physiologic responses following electrical stimulation leg cycle ergometer training. J Rehabil Res Dev. 1995;32: 361–366.

Horiuchi M, Okita K. Arm-Cranking Exercise Training Reduces Plasminogen Activator Inhibitor 1 in People With Spinal Cord Injury. Arch Phys Med Rehabil. 2017;98: 2174–2180.

Jacobs PL, Nash MS, Klose KJ, Guest RS, Needham-Shropshire BM, Green BA. Evaluation of a training program for persons with SCI paraplegia using the Parastep®1 ambulation system: Part 2. Effects on physiological responses to peak arm ergometry. Arch Phys Med Rehabil. 1997;78: 794–798.

Jacobs PL, Nash MS, Rusinowski JW. Circuit training provides cardiorespiratory and strength benefits in persons with paraplegia. Med Sci Sports Exerc. 2001;33: 711–717.

Jacobs PL. Effects of resistance and endurance training in persons with paraplegia. Med Sci Sports Exerc. 2009;41: 992–997.

Janssen TWJ, Pringle DD. Effects of modified electrical stimulation-induced leg cycle ergometer training for individuals with spinal cord injury. J Rehabil Res Dev. 2008;45: 819–830.

Jeon JY, Hettinga D, Steadward RD, Wheeler GD, Bell G, Harber V. Reduced plasma glucose and leptin after 12 weeks of functional electrical stimulation-rowing exercise training in spinal cord injury patients. Arch Phys Med Rehabil. 2010;91: 1957–1959.

Kesiktaş FN, Kaşıkçıoğlu E, Paker N, Bayraktar B, Karan A, Ketenci A, et al. Comparison of the functional and cardiovascular effects of home-based versus supervised hospital circuit training exercises in male wheelchair users with chronic paraplegia. Turk J Phys Med Rehabil. 2021;67(3):275-282.

Kim D-I, Park D-S, Lee BS, Jeon JY. A six-week motor-driven functional electronic stimulation rowing program improves muscle strength and body composition in people with spinal cord injury: a pilot study. Spinal Cord. 2014;52: 621–624.

Kim D-I, Lee H, Lee B-S, Kim J, Jeon JY. Effects of a 6-Week Indoor Hand-Bike Exercise Program on Health and Fitness Levels in People With Spinal Cord Injury: A Randomized Controlled Trial Study. Arch Phys Med Rehabil. 2015;96: 2033–40.e1.

Kim D-I, Taylor JA, Tan CO, Park H, Kim JY, Park S-Y, et al. A pilot randomized controlled trial of 6-week combined exercise program on fasting insulin and fitness levels in individuals with spinal cord injury. Eur Spine J. 2019;28: 1082–1091.

Kjaer M, Mohr T, Biering-Sørensen F, Bangsbo J. Muscle enzyme adaptation to training and tapering-off in spinal-cord-injured humans. Eur J Appl Physiol. 2001;84: 482–486.

Koch I, Schlegel M, Pirrwitz A, Jaschke B, Schlegel K. [Objectification of the training effect of sports therapy for wheelchair users]. Int J Rehabil Res. 1983;6(4):439-448.

Kooijmans H, Post MWM, Stam HJ, van der Woude LHV, Spijkerman DCM, Snoek GJ, et al. Effectiveness of a Self-Management Intervention to Promote an Active Lifestyle in Persons With Long-Term Spinal Cord Injury: The HABITS Randomized Clinical Trial. Neurorehabil Neural Repair. 2017;31: 991–1004.

Koontz AM, Garfunkel CE, Crytzer TM, Anthony SJ, Nindl BC. Feasibility, acceptability, and preliminary efficacy of a handcycling high-intensity interval training program for individuals with spinal cord injury. Spinal Cord. 2021;59: 34–43.

Krauss JC, Robergs RA, Depaepe JL, Kopriva LM, Aisenbury JA, Anderson MA, et al. Effects of electrical stimulation and upper body training after spinal cord injury. Med Sci Sports Exerc. 1993;25: 1054–1061.

Kressler J, Nash MS, Burns PA, Field-Fote EC. Metabolic responses to 4 different body weight-supported locomotor training approaches in persons with incomplete spinal cord injury. Arch Phys Med Rehabil. 2013;94: 1436–1442.

Lavado EL, Cardoso JR, Silva LGA, Dela Bela LF, Atallah AN. Effectiveness of aerobic physical training for treatment of chronic asymptomatic bacteriuria in subjects with spinal cord injury: a randomized controlled trial. Clin Rehabil. 2013;27: 142–149.

Le Foll-de Moro D, Tordi N, Lonsdorfer E, Lonsdorfer J. Ventilation efficiency and pulmonary function after a wheelchair interval-training program in subjects with recent spinal cord injury. Arch Phys Med Rehabil. 2005;86: 1582–1586.

Lindberg T, Arndt A, Norrbrink C, Wahman K, Bjerkefors A. Effects of seated double-poling ergometer training on aerobic and mechanical power in individuals with spinal cord injury. J Rehabil Med. 2012;44: 893–898.

Lotter JK, Henderson CE, Plawecki A, Holthus ME, Lucas EH, Ardestani MM, et al. Task-Specific Versus Impairment-Based Training on Locomotor Performance in Individuals With Chronic Spinal Cord Injury: A Randomized Crossover Study. Neurorehabil Neural Repair. 2020;34: 627–639.

Ma JK, West CR, Martin Ginis KA. The Effects of a Patient and Provider Co-Developed, Behavioral Physical Activity Intervention on Physical Activity, Psychosocial Predictors, and Fitness in Individuals with Spinal Cord Injury: A Randomized Controlled Trial. Sports Med. 2019;49: 1117–1131.

Mcleod JC, Diana H, Hicks AL. Sprint interval training versus moderate-intensity continuous training during inpatient rehabilitation after spinal cord injury: a randomized trial. Spinal Cord. 2020;58: 106–115.

Midha M, Schmitt JK, Sclater M. Exercise effect with the wheelchair aerobic fitness trainer on conditioning and metabolic function in disabled persons: a pilot study. Arch Phys Med Rehabil. 1999;80: 258–261.

Milia R, Roberto S, Marongiu E, Olla S, Sanna I, Angius L, et al. Improvement in hemodynamic responses to metaboreflex activation after one year of training in spinal cord injured humans. Biomed Res Int. 2014;2014: 893468.

Mohr T, Andersen JL, Biering-Sørensen F, Galbo H, Bangsbo J, Wagner A, et al. Long-term adaptation to electrically induced cycle training in severe spinal cord injured individuals. Spinal Cord. 1997;35: 1–16.

Mutton DL, Scremin AM, Barstow TJ, Scott MD, Kunkel CF, Cagle TG. Physiologic responses during functional electrical stimulation leg cycling and hybrid exercise in spinal cord injured subjects. Arch Phys Med Rehabil. 1997;78: 712–718.

Nash MS, Jacobs PL, Mendez AJ, Goldberg RB. Circuit resistance training improves the atherogenic lipid profiles of persons with chronic paraplegia. J Spinal Cord Med. 2001;24: 2–9.

Nightingale TE, Rouse PC, Walhin J-P, Thompson D, Bilzon JLJ. Home-Based Exercise Enhances Health-Related Quality of Life in Persons With Spinal Cord Injury: A Randomized Controlled Trial. Arch Phys Med Rehabil. 2018;99: 1998–2006.e1.

Nooijen CF, van den Brand IL, Ter Horst P, Wynants M, Valent LJ, Stam HJ, et al. Feasibility of Handcycle Training During Inpatient Rehabilitation in Persons With Spinal Cord Injury. Arch Phys Med Rehabil. 2015;96: 1654–1657.

Nooijen CF, Stam HJ, Sluis T, Valent L, Twisk J, van den Berg-Emons RJ. A behavioral intervention promoting physical activity in people with subacute spinal cord injury: secondary effects on health, social participation and quality of life. Clin Rehabil. 2017;31: 772–780.

Ozturk ED, Lapointe MS, Kim D-I, Hamner JW, Tan CO. Effect of 6-Month Exercise Training on Neurovascular Function in Spinal Cord Injury. Med Sci Sports Exerc. 2021;53: 38–46.

Pelletier CA, Totosy de Zepetnek JO, MacDonald MJ, Hicks AL. A 16-week randomized controlled trial evaluating the physical activity guidelines for adults with spinal cord injury. Spinal Cord. 2015;53: 363–367.

Piira A, Lannem AM, Sørensen M, Glott T, Knutsen R, Jørgensen L, et al. Manually assisted body-weight supported locomotor training does not re-establish walking in non-walking subjects with chronic incomplete spinal cord injury: A randomized clinical trial. J Rehabil Med. 2019;51: 113–119.

Pollack SF, Axen K, Spielholz N, Levin N, Haas F, Ragnarsson KT. Aerobic training effects of electrically induced lower extremity exercises in spinal cord injured people. Arch Phys Med Rehabil. 1989;70: 214–219.

Qiu S, Alzhab S, Picard G, Taylor JA. Ventilation Limits Aerobic Capacity after Functional Electrical Stimulation Row Training in High Spinal Cord Injury. Med Sci Sports Exerc. 2016;48: 1111–1118.

Rosety-Rodriguez M, Camacho A, Rosety I, Fornieles G, Rosety MA, Diaz AJ, et al. Low-grade systemic inflammation and leptin levels were improved by arm cranking exercise in adults with chronic spinal cord injury. Arch Phys Med Rehabil. 2014;95: 297–302.

Schleifer G, Solinsky R, Hamner JW, Picard G, Taylor JA. Hybrid Functional Electrical Stimulation Improves Anaerobic Threshold in First Three Years after Spinal Cord Injury. J Neurotrauma. 2022;39:1050-1056.

Sutbeyaz ST, Koseoglu BF, Gokkaya NKO. The combined effects of controlled breathing techniques and ventilatory and upper extremity muscle exercise on cardiopulmonary responses in patients with spinal cord injury. Int J Rehabil Res. 2005;28: 273–276.

Taylor AW, McDonell E, Brassard L. The effects of an arm ergometer training programme on wheelchair subjects. Paraplegia. 1986;24: 105–114.

Taylor JA, Picard G, Porter A, Morse LR, Pronovost MF, Deley G. Hybrid functional electrical stimulation exercise training alters the relationship between spinal cord injury level and aerobic capacity. Arch Phys Med Rehabil. 2014;95: 2172–2179.

Tordi N, Dugue B, Klupzinski D, Rasseneur L, Rouillon JD, Lonsdorfer J. Interval training program on a wheelchair ergometer for paraplegic subjects. Spinal Cord. 2001;39: 532–537.

Tørhaug T, Brurok B, Hoff J, Helgerud J, Leivseth G. The effect from maximal bench press strength training on work economy during wheelchair propulsion in men with spinal cord injury. Spinal Cord. 2016;54: 838–842.

van der Scheer JW, de Groot S, Tepper M, Faber W, ALLRISC group, Veeger DH, et al. Low-intensity wheelchair training in inactive people with long-term spinal cord injury: A randomized controlled trial on fitness, wheelchair skill performance and physical activity levels. J Rehabil Med. 2016;48: 33–42.

Vestergaard M, Jensen K, Juul-Kristensen B. Hybrid high-intensity interval training using functional electrical stimulation leg cycling and arm ski ergometer for people with spinal cord injuries: a feasibility study. Pilot Feasibility Stud. 2022;8(1):43.

Wheeler GD, Andrews B, Lederer R, Davoodi R, Natho K, Weiss C, et al. Functional electric stimulation-assisted rowing: Increasing cardiovascular fitness through functional electric stimulation rowing training in persons with spinal cord injury. Arch Phys Med Rehabil. 2002;83: 1093–1099.

Wilbanks SR, Rogers R, Pool S, Bickel CS. Effects of functional electrical stimulation assisted rowing on aerobic fitness and shoulder pain in manual wheelchair users with spinal cord injury. J Spinal Cord Med. 2016;39: 645–654.

Williams AMM, Chisholm AE, Lynn A, Malik RN, Eginyan G, Lam T. Arm crank ergometer “spin” training improves seated balance and aerobic capacity in people with spinal cord injury. Scand J Med Sci Sports. 2020;30: 361–369.

Wouda MF, Lundgaard E, Becker F, Strøm V. Effects of moderate- and high-intensity aerobic training program in ambulatory subjects with incomplete spinal cord injury-a randomized controlled trial. Spinal Cord. 2018;56: 955–963.

Yarar-Fisher C, Polston KFL, Eraslan M, Henley KY, Kinikli GI, Bickel CS, et al. Paralytic and nonparalytic muscle adaptations to exercise training versus high-protein diet in individuals with long-standing spinal cord injury. J Appl Physiol. 2018;125: 64–72.
