## Supplementary material for "The effect of exercise on aerobic capacity in individuals with spinal cord injury: A systematic review with meta-analysis and meta-regression": S7 Adverse events

**Electronic Supplementary Material 7**

**PLoS Medicine**

Hodgkiss, D.D^1^, Bhangu, G^2,3^, Lunny, C^4^, Jutzeler C.R^5,6^, Chiou S.Y^1,7,8,9^ Walter, M^2,10^, Lucas S.E^1,7^, Krassioukov, A.V.^2,11,12^, Nightingale, T.E.^1,2,9^

**^1^** School of Sport, Exercise and Rehabilitation Sciences, University of Birmingham, UK.

**^2^** International Collaboration on Repair Discoveries (ICORD), University of British Columbia, Vancouver, British Columbia, Canada. **^3^** MD Undergraduate Program, Faculty of Medicine, University of British Columbia, Vancouver, Canada. **^4^** Knowledge Translation Program, Li Ka Shing Knowledge Institute, St. Michael’s Hospital, Toronto, and the University of British Columbia, Vancouver, British Columbia, Canada.

**^5^** Department of Health Sciences and Technology, ETH Zurich, Zurich, Switzerland. **^6^** Schulthess Clinic, Zurich, Switzerland. **^7^** Centre for Human Brain Health, University of Birmingham, United Kingdom. **^8^** MRC Versus Arthritis Centre for Musculoskeletal Ageing Research, University of Birmingham, United Kingdom.

**^9^** Centre for Trauma Science Research, University of Birmingham, United Kingdom. **^10^** Department of Urology, University Hospital Basel, University of Basel, Basel, Switzerland. **^11^** Department of Medicine, Division of Physical Medicine and Rehabilitation, University of British Columbia, Vancouver, British Columbia, Canada. **^12^** GF Strong Rehabilitation Centre, Vancouver Coastal Health, Vancouver, British Columbia, Canada.

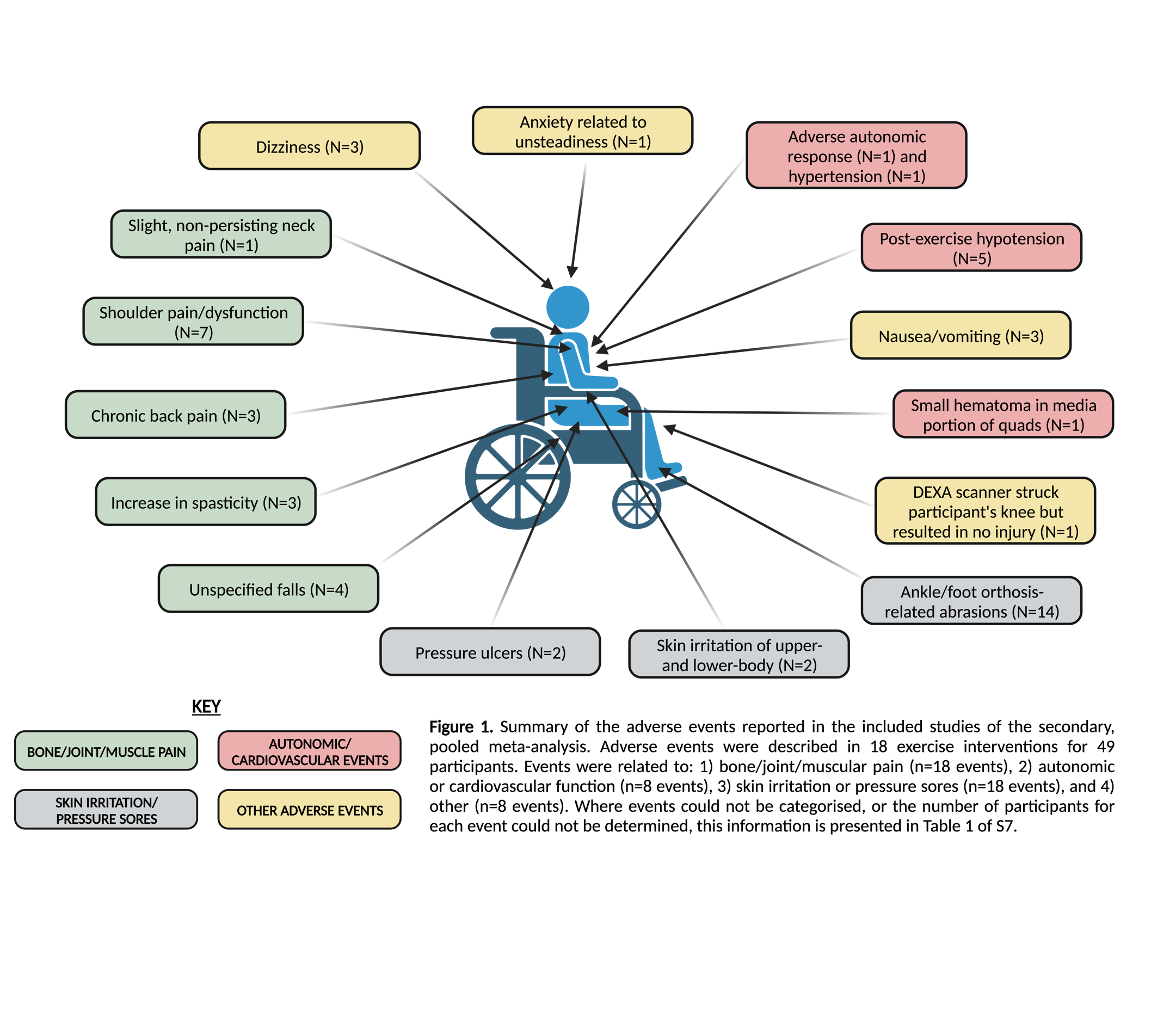

| **Table 1.** Adverse events described in the exercise intervention studies included in the primary meta-analysis. | | | | | | | |
| --- | --- | --- | --- | --- | --- | --- | --- |
| **Author/**  **Year** | **Exercise Modality** | **Total N** | **Adverse Event**s | | | | |
|  |  |  | **Total Adverse Events (%N)** | **Bone, joint or muscular pain** | **Autonomic or CV function** | **Skin irritation/ pressure sores** | **Other** |
| Beillot et al. (1996) | Walking with reciprocating gait orthosis | **14** | **10 (71%)**  *(“10 participants had to stop the training programme, the major complications included spontaneous fractures of lower limbs, occurrence of a syringomyelia and pressure sores at the foot and ankle”)* | **NS** | **0** | **NS** | **NS** |
| Berry et al. (2008) | Home-based FES-cycling | **11** | **1 (9%)**  *(“one subject dropped out of the study after baseline testing due to an adverse autonomic response to stimulation, data not included”)* | **0** | **1** | **0** | **0** |
| Bombardier et al. (2021) | Behaviour change (with tele-health intervention) | **7** | **1 (14%)**  *(“arm of the DXA scanner struck a participant's knee as it was returning to the start position but the physician detected no injury”)* | **0** | **0** | **0** | **1** |
| Brazg et al. (2017) | Low-intensity locomotor training | **15** | **1 (7%)**  *(“one individual terminated participation due to an increase in his back pain”)* | **1** | **0** | **0** | **0** |
| Brurok et al. (2011) | Aerobic HIIT hybrid cycling (arm crank ergometry and FES cycling) | **6** | **2 (33%)**  *(“two persons exacerbated a latent shoulder dysfunction during training but commenced shortly after a period of rest and therapy”)* | **2** | **0** | **0** | **0** |
| Duran et al. (2001) | Mixed exercise programme (mobility, strength, coordination, aerobic resistance,  and relaxation activities) | **13** | **1 (8%)**  *(“one patient developed transient sinus bradycardia and hypotension after the arm crank exercise test”)* | **0** | **1** | **0** | **0** |
| Gauthier et al. (2018) | Home-based, high-intensity wheelchair ergometry (HIIT group) | **4** | **1 (25%)**  *(“one dropout in the HIIT group was due to development of significant shoulder pain over the course of the training program”)* | **1** | **0** | **0** | **0** |
| Gibbons et al. (2016) | FES-rowing | **5** | **NS**  *(“a number of participants showed some level of autonomic dysreflexia during the FES response test”)* | **0** | **NS** | **0** | **0** |
| Gorman et al. (2016) | Robotically assisted BWSTT | **12** | **2 (17%)**  *(“skin irritation in hip, groin, penis, back, wrist, glutei and scapula”)* | **0** | **0** | **2** | **0** |
| Janssen et al. (2008) | FES-cycling | **12** | **NS**  *(“lightheadedness in some subjects”)* | **0** | **0** | **0** | **NS** |
| Koontz et al. (2021) | High-intensity handcycling | **10** | **1 (10%)**  *(“one participant experienced bad spasms during a session - but repeated this session on another day”)* | **1** | **0** | **0** | **0** |
| Kressler et al. (2013) | Treadmill- based locomotor training with transcutaneous FES (TS) | **17** | **1 (6%)**  *(“increase in spasticity and increase in chronic back pain in a patient”)* | **1** | **0** | **0** | **0** |
| Lotter et al. (2020) | Impairment- based training (e.g., recumbent cycling or stepping) | **16** | **5 (31%)**  *(“4 incidents of soreness/ankle foot orthoses– related abrasions and one incident of nausea”)* | **0** | **0** | **4** | **1** |
|  | Task-specific training (e.g., stepping practice including speed- dependent treadmill training, over-ground training, skill-dependent treadmill training, and stair climbing) | **16** | **16 (100%)**  *(“11 falls without significant injury outside of task-specific training, with subsequent discomfort in 4 incidences; one incident of hypertension; 10 incidents of soreness/ankle foot orthoses– related abrasions; one incident of anxiety related to the perception of unsteadiness such that assistance was required”)* | **4** | **1** | **10** | **1** |
| Mcleod et al. (2020) | Sprint interval training (high-intensity) using arm crank ergometry  (HIIT group) | **10** | **1 (10%)**  *(“post-exercise hypotension”)* | **0** | **1** | **0** | **0** |
| Mohr et al. (1997) | FES-cycling | **10** | **4 (40%)**  *(“three subjects experienced post-exercise hypotension…one subject experienced a small hematoma in the media portion of the quads after 15 min of exercise in the 2nd exercise bout”)* | **0** | **4** | **0** | **0** |
| Nooijen et al. (2015) | Handcycling | **45** | **2 (4%)**  *(“2 participants did not complete training due to severe pressure ulcers”)* | **0** | **0** | **2** | **0** |
| Vestergaard et al. (2022) | FES-  skiergometry | **7** | **NS - It cannot be determined whether these adverse events were in one participant or multiple.**  *(“Only a few minor adverse effects occurred: slight non-persisting pain in neck (n = 1), arms and shoulders (n = 4) during and between training sessions, dizziness that disappeared after 5 min (n = 1), feeling tired in the head/dizziness that disappeared after training with no other signs of autonomic hyperreflexia (n = 2), increased spasms (n = 2), and vomiting just after training (n = 2)”* | **7** | **0** | **0** | **5** |
| Number of participants in each intervention (N), and the percentage of those who experienced an adverse event (%N). BWSTT, body-weight supported treadmill training; CV, cardiovascular; FES, functional electrical stimulation; HIIT, high-intensity interval training NS, not specified. | | | | | | | |
