## Supplementary material for "The effect of exercise on aerobic capacity in individuals with spinal cord injury: A systematic review with meta-analysis and meta-regression": S8 Sensitivity analyses

**Electronic Supplementary Material 8**

**PLoS Medicine**

Hodgkiss, D.D^1^, Bhangu, G^2,3^, Lunny, C^4^, Jutzeler C.R^5,6^, Chiou S.Y^1,7,8,9^ Walter, M^2,10^, Lucas S.E^1,7^, Krassioukov, A.V.^2,11,12^, Nightingale, T.E.^1,2,9^

**^1^** School of Sport, Exercise and Rehabilitation Sciences, University of Birmingham, UK.

**^2^** International Collaboration on Repair Discoveries (ICORD), University of British Columbia, Vancouver, British Columbia, Canada. **^3^** MD Undergraduate Program, Faculty of Medicine, University of British Columbia, Vancouver, Canada. **^4^** Knowledge Translation Program, Li Ka Shing Knowledge Institute, St. Michael’s Hospital, Toronto, and the University of British Columbia, Vancouver, British Columbia, Canada.

**^5^** Department of Health Sciences and Technology, ETH Zurich, Zurich, Switzerland. **^6^** Schulthess Clinic, Zurich, Switzerland. **^7^** Centre for Human Brain Health, University of Birmingham, United Kingdom. **^8^** MRC Versus Arthritis Centre for Musculoskeletal Ageing Research, University of Birmingham, United Kingdom.

**^9^** Centre for Trauma Science Research, University of Birmingham, United Kingdom. **^10^** Department of Urology, University Hospital Basel, University of Basel, Basel, Switzerland. **^11^** Department of Medicine, Division of Physical Medicine and Rehabilitation, University of British Columbia, Vancouver, British Columbia, Canada. **^12^** GF Strong Rehabilitation Centre, Vancouver Coastal Health, Vancouver, British Columbia, Canada.

**High risk of bias:**

| **Table 1.** A sensitivity analysis comparing studies with a low risk of bias/good quality versus studies with a high risk of bias/poor quality. | | | | | | |
| --- | --- | --- | --- | --- | --- | --- |
|  | **AV̇O_2peak_ (L/min)** | | **RV̇O_2peak_ (mL/kg/min)** | | **PPO (W)** | |
|  | **N [Σ]** | **WMD [95% CIs]** | **N [Σ]** | **WMD [95% CIs]** | **N [Σ]** | **WMD [95% CIs]** |
| **Low risk** | 23 [271] | 0.20 [0.15, 0.25]  ***p* < 0.001**  I^2^ = 45% (***p* = 0.02**) | 30 [351] | 2.7 [2.0, 3.4]  ***p* < 0.001**  I^2^ = 27% (*p* = 0.34) | 16 [208] | 14 [9, 18]  ***p* < 0.001**  I^2^ = 75% (***p* < 0.001**) |
| **High risk** | 21 [174] | 0.23 [0.15, 0.32]  ***p* < 0.001**  I^2^ = 71% (***p* < 0.001**) | 23 [174] | 2.8 [1.8, 3.7]  ***p* < 0.001**  I^2^ = 32% (*p* = 0.17) | 20  [154] | 12 [7, 17]  ***p* < 0.001**  I^2^ = 59% (***p* < 0.001**) |
| **Subgroup differences** | Q = 0.47, df = 1  *p* = 0.49 | | Q = 0.00, df = 1  *p* = 0.96 | | Q = 0.25, df = 1  *p* = 0.62 | |
| Total number of interventions (N) and sum of participants analysed at post-intervention [Σ]. AV̇O_2peak_, absolute peak oxygen consumption; CIs, confidence intervals; PPO, peak power output; RV̇O_2peak_, relative peak oxygen consumption; WMD, weighted mean difference. Thresholds for statistically significant subgroup differences are adjusted for the number of subgroup comparisons (*p*<0.025). Individual subgroup p-values were adjusted for multiple comparisons via the Bonferroni correction method. | | | | | | |

**Table 2.** Studies classified as low or high risk of bias:

| **AV̇O_2peak_ (L/min)** | | **RV̇O_2peak_ (mL/kg/min)** | | **PPO (W)** | |
| --- | --- | --- | --- | --- | --- |
| Low risk | High risk | Low risk | High risk | Low risk | High risk |
| Alrashidi 2021 | Davis 1987 | Akkurt 2017 | Capodaglio 1996 | Akkurt 2017 | de Groot 2003 |
| Berry 2008 | El-Sayed 2005 | Alrashidi 2021 | de Groot 2003 | Alrashidi 2021 | El-Sayed 2005  Froehlich-Grobe 2022 |
| Carty 2012 | Gauthier 2018 | Carty 2012 | El-Sayed 2005  Froehlich-Grobe 2022 | Berry 2008 | Gauthier 2018 |
| Gorgey 2021 | Gurney 1998  Haddad 1997  Hansen 2023 | Cheung 2019 | Gauthier 2018 | Cooney 1986 | Hansen 2023  Hasnan 2013 |
| Goss 1992 | Hasnan 2013 | Cooney 1986 | Gorman 2016  Hansen 2023 | Duran 2001 | Heesterbeek 2005 |
| Hjeltnes 1998 | Jacobs 2009 | DiPiro 2016 | Hasnan 2013 | Hjeltnes 1998 | Hjeltnes 1997 |
| Hoekstra 2013 | Jeon 2010  Koch 1983 | Gant 2018 | Heesterbeek 2005 | Jacobs 2001 | Hooker 1989 |
| Horiuchi 2017 | Mutton 1997 | Gorman 2019 | Hjeltnes 1997 | Krauss 1993 | Jeon 2010  Koch 1983 |
| Jacobs 2001 | Nash 2001 | Hjeltnes 1998 | Hooker 1989 | Nooijen 2015 | Mutton 1997 |
| Kooijmans 2017 | Nightingale 2018 | Hoekstra 2013 | Jeon 2010 | Qiu 2016 | Nash 2001 |
| Krauss 1993 | Pelletier 2015 | Horiuchi 2017 | Kim 2019 | Sutbeyaz 2005 | Nightingale 2018 |
| Lavado 2013 | Piira 2019 | Jacobs 2001 | Nightingale 2018 | Taylor 2014 | Pelletier 2015 |
| Lindberg 2012 | Pollack 1989 | Kim 2014 | Pelletier 2015 | Wilbanks 2016 | Tordi 2001 |
| Midha 1999 | Taylor 1986 | Lindberg 2012 | Rosety-Rodriguez 2014 | Williams 2020 |  |
| Nooijen 2015 | Torhaug 2016 | Lotter 2020 | Taylor 1986 |  |  |
| Ozturk 2021 |  | Midha 1999 | Tordi 2001 |  |  |
| Qiu 2016 |  | Nooijen 2015 | Torhaug 2016 |  |  |
| Wouda 2018 |  | Ozturk 2021 | Yarar-Fisher 2018 |  |  |
|  |  | Qiu 2016 |  |  |  |
|  |  | Sutbeyaz 2005 |  |  |  |
|  |  | Taylor 2014 |  |  |  |
|  |  | Wilbanks 2016 |  |  |  |
|  |  | Williams 2020 |  |  |  |
|  |  | Wouda 2018 |  |  |  |

**Imputed data:**

| **Table 3.** A sensitivity analysis comparing studies with versus without imputed data. | | | | | | |
| --- | --- | --- | --- | --- | --- | --- |
|  | **AV̇O_2peak_ (L/min)** | | **RV̇O_2peak_ (mL/kg/min)** | | **PPO (W)** | |
|  | **N [Σ]** | **WMD [95% CIs]** | **N [Σ]** | **WMD [95% CIs]** | **N [Σ]** | **WMD [95% CIs]** |
| **Not imputed** | 58 [565] | 0.22 [0.16, 0.27]  ***p* < 0.001**  I^2^ = 68% (***p* < 0.001**) | 65 [612] | 2.5 [1.9, 3.1]  ***p* < 0.001**  I^2^ = 49% (***p* < 0.001**) | 53 [525] | 10 [8, 13]  ***p* < 0.001**  I^2^ = 74% (***p* < 0.001**) |
| **Imputed** | 12 [180] | 0.17 [0.14, 0.19]  ***p* < 0.001**  I^2^ = 0% (*p* = 0.70) | 8  [127] | 4.4 [2.4, 6.4]  ***p* < 0.001**  I^2^ = 56% (***p* = 0.03**) | 7  [88] | 13 [6, 19]  ***p* < 0.001**  I^2^ = 76% (***p* < 0.001**) |
| **Extrapolated** | 4  [34] | 0.25 [0.18, 0.31]  ***p* < 0.001**  I^2^ = 14% (*p* = 0.39) | 6  [39] | 3.9 [2.3, 5.4]  ***p* < 0.001**  I^2^ = 51% (*p* = 0.08) | 5  [34] | 18 [11, 25]  ***p* < 0.001**  I^2^ = 17% (*p* = 0.50) |
| **Subgroup differences** | Q = 7.30, df = 2  *p* = 0.03 | | Q = 5.10, df = 2  *p* = 0.08 | | Q = 4.31, df = 2  *p* = 0.12 | |
| Total number of interventions (N) and sum of participants analysed at post-intervention [Σ]. AV̇O_2peak_, absolute peak oxygen consumption; CIs, confidence intervals; PPO, peak power output; RV̇O_2peak_, relative peak oxygen consumption; WMD, weighted mean difference. Thresholds for statistically significant subgroup differences are adjusted for the number of subgroup comparisons (*p*<0.017). Individual subgroup p-values were adjusted for multiple comparisons via the Bonferroni correction method. | | | | | | |

**Table 4.** Studies classified as low or high risk of bias:

| **AV̇O_2peak_ (L/min)** | | **RV̇O_2peak_ (mL/kg/min)** | | **PPO (W)** | |
| --- | --- | --- | --- | --- | --- |
| Not imputed | Imputed/Extrapolated | Not imputed | Imputed/Extrapolated | Not imputed | Imputed/Extrapolated |
| Alrashidi 2021 | Bakkum 2015 | Alrashidi 2021 | Akkurt 2017 | Alrashidi 2021 | Akkurt 2017 |
| Barstow 1996 | **Hasnan 2013** | Beillot 1996 | Capodaglio 1996 | Barstow 1996 | Bakkum 2015 |
| Beillot 1996 | Hooker 1992 | Bombardier 2021 | Carty 2012 | Beillot 1996 | **Cooney 1986** |
| Berry 2008 | Horiuchi 2017 | Brazg 2017 | **Cooney 1986** | Berry 2008 | **de Groot 2003** |
| Bresnahan 2019 | Jeon 2010 | Bresnahan 2019 | **de Groot 2003** | Bombardier 2021 | **Gibbons 2014** |
| Brurok 2011 | Kjaer 2001  **Koch 1983** | Brurok 2011 | **Fukuoka 2006** | Bresnahan 2019 | Hooker 1992  **Koch 1983** |
| Carty 2012 | **Krauss 1993** | Cheung 2019 | **Hjeltnes 1997**  Kesiktas 2021 | Brurok 2011 | Le Foll de Moro 2005 |
| Davis 1987 | Lavado 2013 | DiCarlo 1988 | Nooijen 2015 | Duffell 2019 | Nooijen 2015 |
| DiCarlo 1988 | Le Foll de Moro 2005 | DiPiro 2016 | Schleifer 2022 | Duran 2001 | Tordi 2001 |
| El-Sayed 2005 | Midha 1999 | El-Sayed 2005 | Tordi 2001 | El-Sayed 2005 |  |
| Farkas 2021 | Nooijen 2015 | Farkas 2021  Froehlich-Grobe 2022 | **Wilbanks 2016** | Farkas 2021  Froehlich-Grobe 2022 |  |
| Gass 1980 | Qiu 2016  Schleifer 2022 | Gant 2018 |  | Gauthier 2018 |  |
| Gauthier 2018 |  | Gass 1980 |  | Gibbons 2016 |  |
| Gibbons 2016 |  | Gauthier 2018 |  | Gorgey 2021 |  |
| Gorgey 2021 |  | Gibbons 2016 |  | Grange 2002 |  |
| Goss 1992 |  | Gorman 2016 |  | Han 2016  Hansen 2023 |  |
| Gurney 1998 |  | Gorman 2019 |  | Hasnan 2013 |  |
| Han 2016  Hansen 2023 |  | Graham 2019 |  | Heesterbeek 2005 |  |
| Hjeltnes 1998 |  | Grange 2002 |  | Hjeltnes 1997 |  |
| Hoekstra 2013 |  | Han 2016  Hansen 2023 |  | Hjeltnes 1998 |  |
| Hooker 1995 |  | Hasnan 2013 |  | Hooker 1989 |  |
| Jacobs 2001 |  | Heesterbeek 2005 |  | Hooker 1995 |  |
| Jacobs 2009 |  | Hjeltnes 1998 |  | Jacobs 1997 |  |
| Janssen 2008 |  | Hoekstra 2013 |  | Jacobs 2001 |  |
| Kooijmans 2017 |  | Hooker 1989 |  | Janssen 2008 |  |
| Koontz 2021 |  | Hooker 1995 |  | Jeon 2010 |  |
| Kressler 2013 |  | Horiuchi 2017 |  | Kooijmans 2017 |  |
| Lindberg 2012 |  | Jacobs 1997 |  | Koontz 2021 |  |
| Ma 2019 |  | Jacobs 2001 |  | Krauss 1993 |  |
| Milia 2014 |  | Jeon 2010 |  | Ma 2019 |  |
| Mohr 1997 |  | Kim 2014 |  | Mcleod 2020 |  |
| Mutton 1997 |  | Kim 2015 |  | Milia 2014 |  |
| Nash 2001 |  | Kim 2019 |  | Mutton 1997 |  |
| Nightingale 2018 |  | Koontz 2021 |  | Nash 2001 |  |
| Nooijen 2017 |  | Lindberg 2012 |  | Nightingale 2018 |  |
| Ozturk 2021 |  | Lotter 2020 |  | Nooijen 2017 |  |
| Pelletier 2015 |  | Ma 2019 |  | Pelletier 2015 |  |
| Piira 2019 |  | Midha 1999 |  | Qiu 2016 |  |
| Pollack 1989 |  | Milia 2014 |  | Sutbeyaz 2005 |  |
| Taylor 1986 |  | Nightingale 2018 |  | Taylor 2014 |  |
| Torhaug 2016 |  | Ozturk 2021 |  | van der Scheer 2016  Vestergaard 2022 |  |
| van der Scheer 2016  Vestergaard 2022 |  | Pelletier 2015 |  | Wilbanks 2016 |  |
| Wheeler 2002 |  | Qiu 2016 |  | Williams 2020 |  |
| Wouda 2018 |  | Rosety-Rodriguez 2014 |  |  |  |
|  |  | Sutbeyaz 2005 |  |  |  |
|  |  | Taylor 1986 |  |  |  |
|  |  | Taylor 2014 |  |  |  |
|  |  | Torhaug 2016  Vestergaard 2022 |  |  |  |
|  |  | Williams 2020 |  |  |  |
|  |  | Wouda 2018 |  |  |  |
|  |  | Yarar-Fisher |  |  |  |

Data extrapolated from figures in studies are highlighted in bold.

**Leave-one-out analyses:**

Outliers were identified for the following outcomes:

- AV̇O_2peak_: DiCarlo 1988 and Pollack 1989
- RV̇O_2peak_: DiCarlo 1988
- PPO: Farkas 2021 ACE

Please see the below plots for each CRF outcome with one study omitted at a time and the resulting effect sizes.

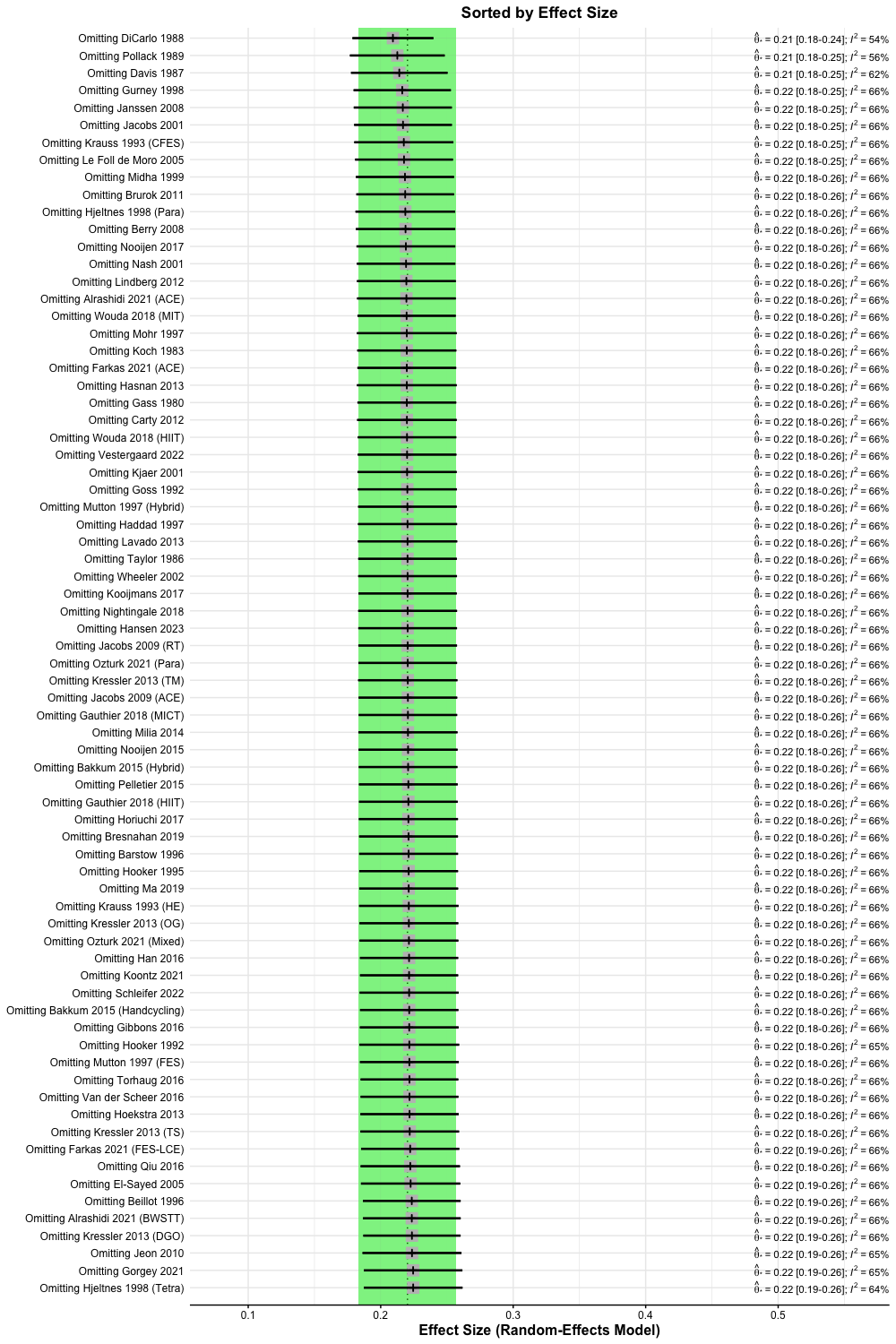

**Figure 1.** Leave-one-out analysis forest plot for interventions reporting change in AV̇O_2peak_, ordered by effect size.

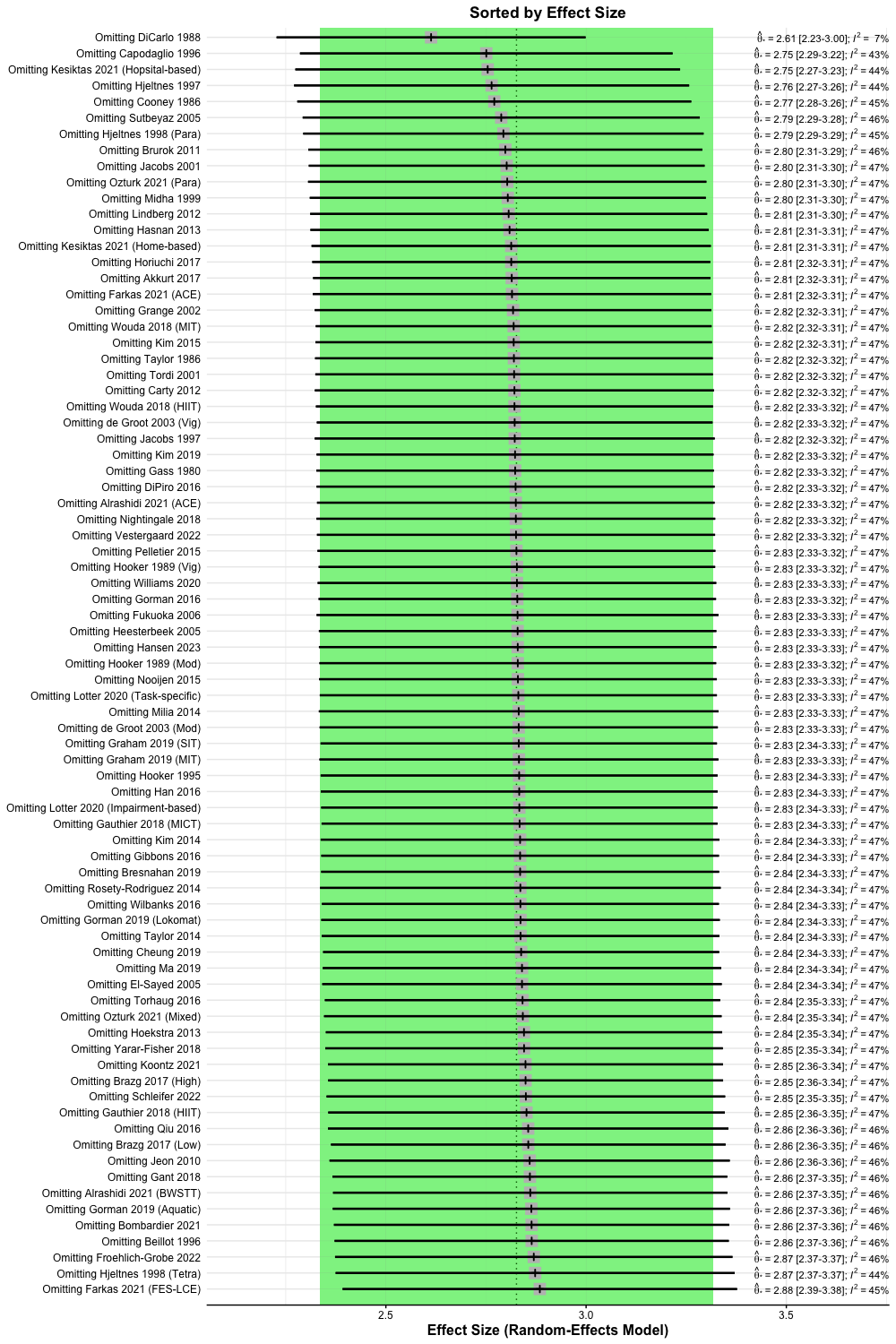

**Figure 2.** Leave-one-out analysis forest plot for interventions reporting change in RV̇O_2peak_, ordered by effect size.

**Figure 3.** Leave-one-out analysis forest plot for interventions reporting change in PPO, ordered by effect size.

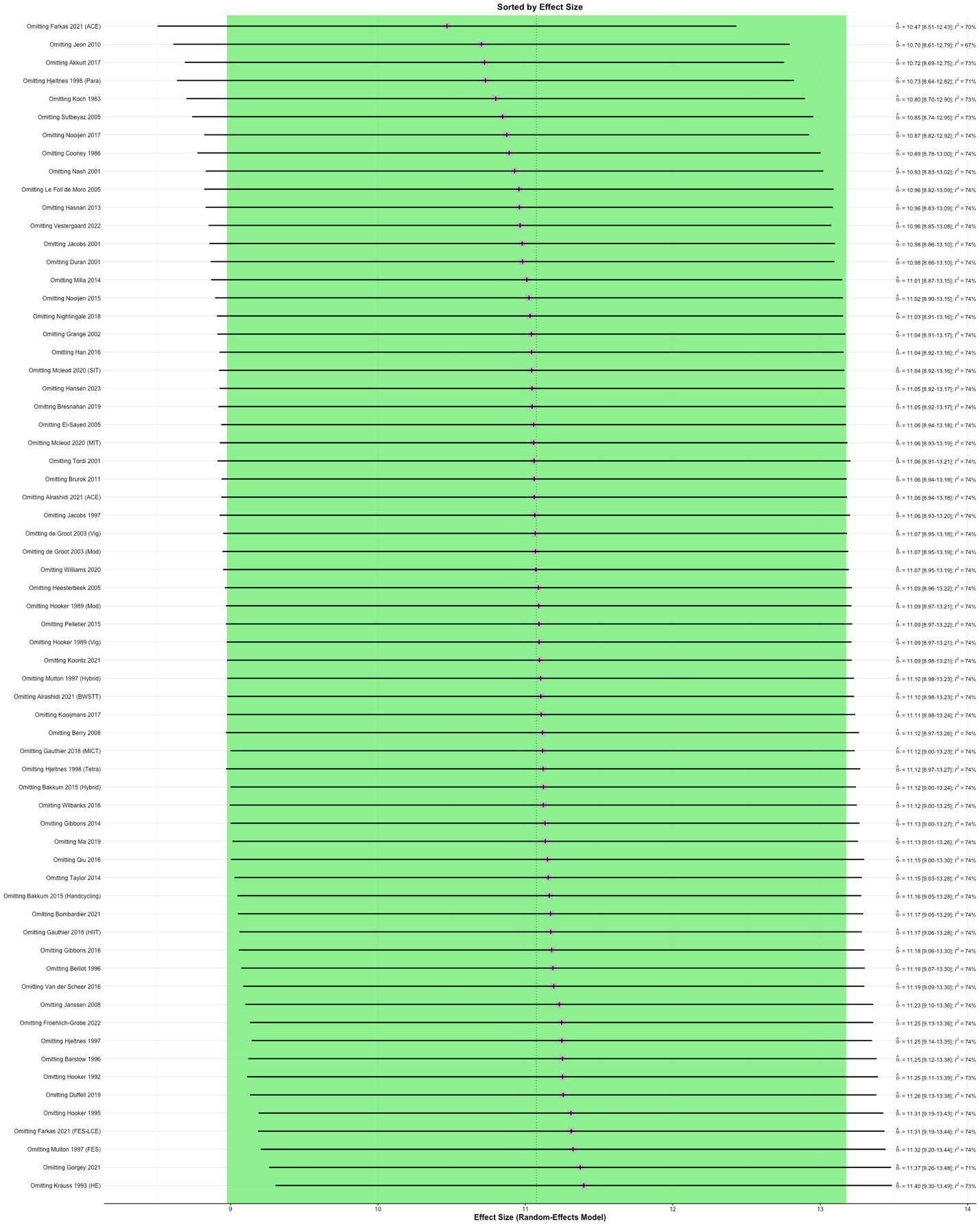
