## Supplementary material for "The effect of exercise on aerobic capacity in individuals with spinal cord injury: A systematic review with meta-analysis and meta-regression": S9 Cardiopulmonary exercise test modalities sub-analysis

**Electronic Supplementary Material 9**

**PLoS Medicine**

Hodgkiss, D.D^1^, Bhangu, G^2,3^, Lunny, C^4^, Jutzeler C.R^5,6^, Chiou S.Y^1,7,8,9^ Walter, M^2,10^, Lucas S.E^1,7^, Krassioukov, A.V.^2,11,12^, Nightingale, T.E.^1,2,9^

**^1^** School of Sport, Exercise and Rehabilitation Sciences, University of Birmingham, UK.

**^2^** International Collaboration on Repair Discoveries (ICORD), University of British Columbia, Vancouver, British Columbia, Canada. **^3^** MD Undergraduate Program, Faculty of Medicine, University of British Columbia, Vancouver, Canada. **^4^** Knowledge Translation Program, Li Ka Shing Knowledge Institute, St. Michael’s Hospital, Toronto, and the University of British Columbia, Vancouver, British Columbia, Canada.

**^5^** Department of Health Sciences and Technology, ETH Zurich, Zurich, Switzerland. **^6^** Schulthess Clinic, Zurich, Switzerland. **^7^** Centre for Human Brain Health, University of Birmingham, United Kingdom. **^8^** MRC Versus Arthritis Centre for Musculoskeletal Ageing Research, University of Birmingham, United Kingdom.

**^9^** Centre for Trauma Science Research, University of Birmingham, United Kingdom. **^10^** Department of Urology, University Hospital Basel, University of Basel, Basel, Switzerland. **^11^** Department of Medicine, Division of Physical Medicine and Rehabilitation, University of British Columbia, Vancouver, British Columbia, Canada. **^12^** GF Strong Rehabilitation Centre, Vancouver Coastal Health, Vancouver, British Columbia, Canada.

***Purpose:*** This supplementary file contains comparisons between exercise interventions that matched the cardiopulmonary exercise test modality to the exercise intervention modality versus those that did not. Forest plots are presented for each cardiorespiratory fitness (CRF) outcome. This was conducted to investigate whether greater effects are observed when using a consistent modality or whether there may be transfer effects between modalities.

***Conclusion:*** Following the adjustment for multiple comparisons, there were no significant differences in AV̇O_2peak_, RV̇O_2peak_ or PPO for studies that matched the modality in the CPET to that in the exercise intervention, versus studies that did not. Despite this, there were trends for significance for AV̇O_2peak_, and RV̇O_2peak_. Future studies may want to consider multiple assessments of CRF using different CPET modalities to test whether improvements are transferrable across exercise modalities. It is currently unclear how much of the improvement in CRF following different exercise interventions is due to central or peripheral training adaptations or merely familiarisation effects or improved movement economy.

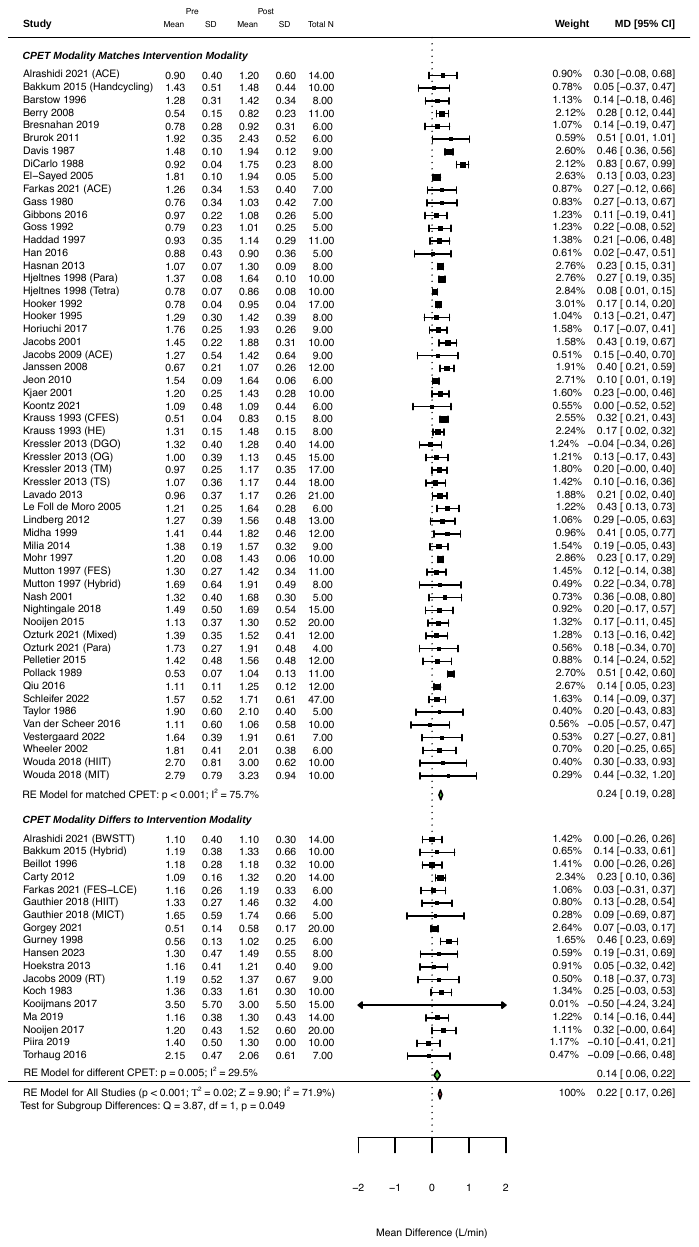

**Figure 1:** Forest plot of absolute peak oxygen consumption with studies grouped into subgroups of 1) studies matching the cardiopulmonary exercise test (CPET) modality with the exercise intervention modality, and 2) studies with different CPET and intervention modalities. Subgroup difference *p*-value was adjusted for multiple comparisons; statistically significant at *p*<0.025.

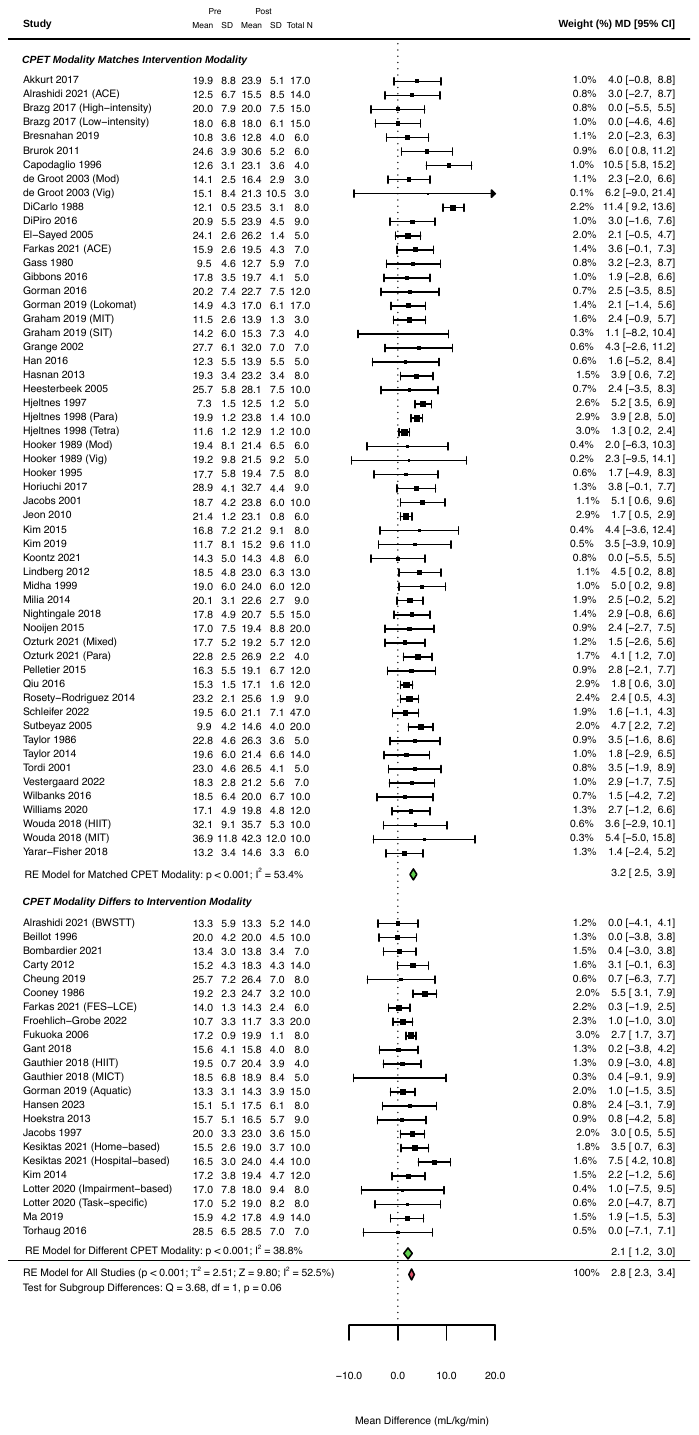

**Figure 2:** Forest plot of relative peak oxygen consumption with studies grouped into subgroups of 1) studies matching the cardiopulmonary exercise test (CPET) modality with the exercise intervention modality, and 2) studies with different CPET and intervention modalities. Subgroup difference *p*-value was adjusted for multiple comparisons; statistically significant at *p*<0.025.

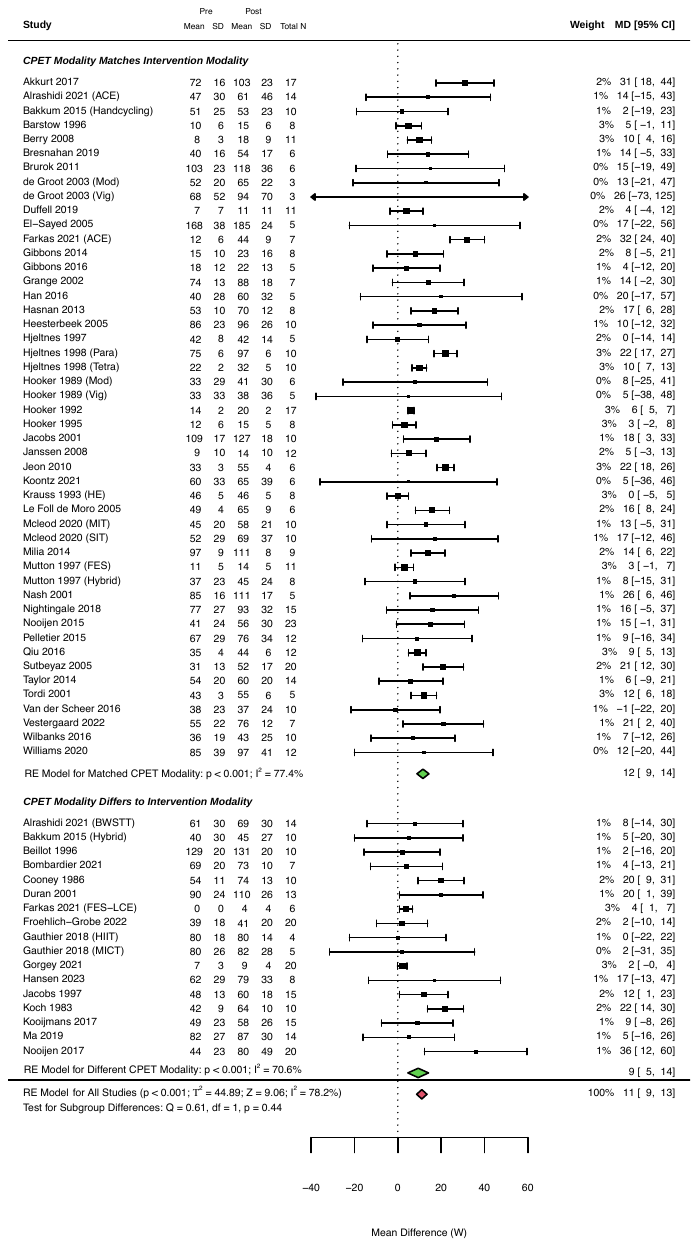

**Figure 3:** Forest plot of peak power output with studies grouped into subgroups of 1) studies matching the cardiopulmonary exercise test (CPET) modality with the exercise intervention modality, and 2) studies with different CPET and intervention modalities. Subgroup difference *p*-value was adjusted for multiple comparisons; statistically significant at *p*<0.025.
