## Supplementary material for "The effect of exercise on aerobic capacity in individuals with spinal cord injury: A systematic review with meta-analysis and meta-regression": S10 Gait training sub-analysis

**Electronic Supplementary Material 10**

**PLoS Medicine**

Hodgkiss, D.D^1^, Bhangu, G^2,3^, Lunny, C^4^, Jutzeler C.R^5,6^, Chiou S.Y^1,7,8,9^ Walter, M^2,10^, Lucas S.E^1,7^, Krassioukov, A.V.^2,11,12^, Nightingale, T.E.^1,2,9^

**^1^** School of Sport, Exercise and Rehabilitation Sciences, University of Birmingham, UK.

**^2^** International Collaboration on Repair Discoveries (ICORD), University of British Columbia, Vancouver, British Columbia, Canada. **^3^** MD Undergraduate Program, Faculty of Medicine, University of British Columbia, Vancouver, Canada. **^4^** Knowledge Translation Program, Li Ka Shing Knowledge Institute, St. Michael’s Hospital, Toronto, and the University of British Columbia, Vancouver, British Columbia, Canada.

**^5^** Department of Health Sciences and Technology, ETH Zurich, Zurich, Switzerland. **^6^** Schulthess Clinic, Zurich, Switzerland. **^7^** Centre for Human Brain Health, University of Birmingham, United Kingdom. **^8^** MRC Versus Arthritis Centre for Musculoskeletal Ageing Research, University of Birmingham, United Kingdom.

**^9^** Centre for Trauma Science Research, University of Birmingham, United Kingdom. **^10^** Department of Urology, University Hospital Basel, University of Basel, Basel, Switzerland. **^11^** Department of Medicine, Division of Physical Medicine and Rehabilitation, University of British Columbia, Vancouver, British Columbia, Canada. **^12^** GF Strong Rehabilitation Centre, Vancouver Coastal Health, Vancouver, British Columbia, Canada.

***Purpose:*** This supplementary file contains a sub-analysis of gait training cardiopulmonary exercise test (CPET) modalities. The purpose was to compare whether there are any transfer effects between a gait training intervention and upper-body exercise (i.e., arm crank ergometry performance in a CPET).

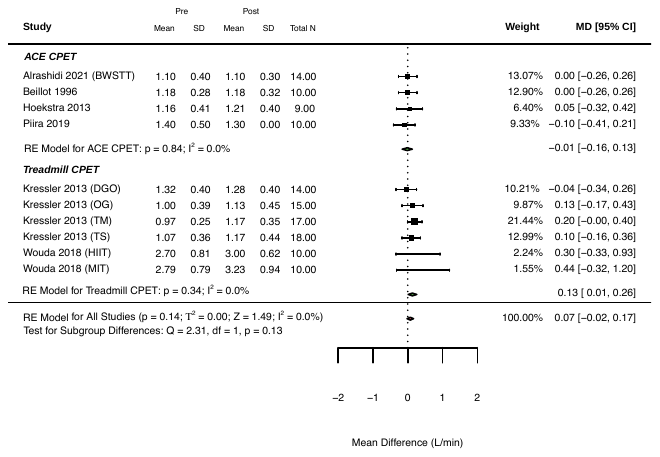
***Conclusion:*** Although there are no significant subgroup differences between arm-crank ergometry and treadmill performance following a gait training exercise intervention, there are larger pooled effect estimates for absolute and relative peak oxygen consumption in interventions using a treadmill CPET.

**Figure 1:** Forest plot of absolute peak oxygen consumption with gait training interventions grouped into subgroups by cardiopulmonary exercise test (CPET) modality. Subgroup difference p-value was adjusted for multiple comparisons; statistically significant at *p*<0.025.

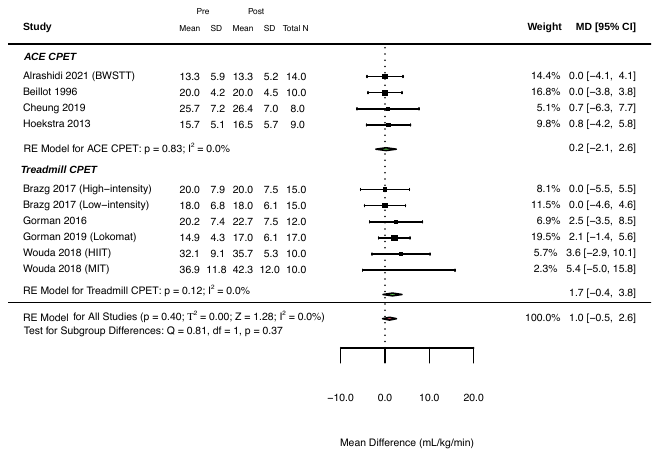

**Figure 2:** Forest plot of relative peak oxygen consumption with gait training interventions grouped into subgroups by cardiopulmonary exercise test (CPET) modality. Subgroup difference p-value was adjusted for multiple comparisons; statistically significant at *p*<0.025.

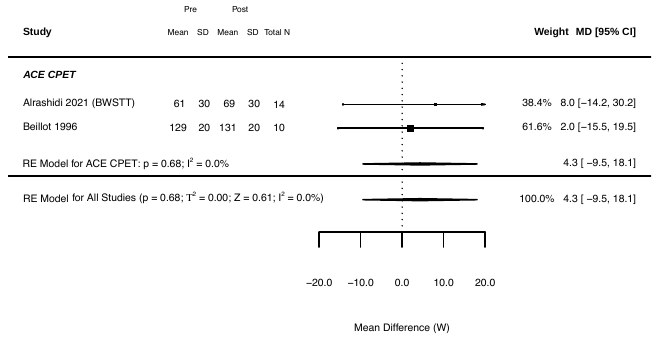

**Figure 3:** Forest plot of peak power output with gait training interventions using arm-crank ergometry (ACE) cardiopulmonary exercise test (CPET). No interventions used treadmill CPET.
