## Supplementary material for "The effect of exercise on aerobic capacity in individuals with spinal cord injury: A systematic review with meta-analysis and meta-regression": S11 Cohort comparisons

**Electronic Supplementary Material 11**

**PLoS Medicine**

Hodgkiss, D.D^1^, Bhangu, G^2,3^, Lunny, C^4^, Jutzeler C.R^5,6^, Chiou S.Y^1,7,8,9^ Walter, M^2,10^, Lucas S.E^1,7^, Krassioukov, A.V.^2,11,12^, Nightingale, T.E.^1,2,9^

**^1^** School of Sport, Exercise and Rehabilitation Sciences, University of Birmingham, UK.

**^2^** International Collaboration on Repair Discoveries (ICORD), University of British Columbia, Vancouver, British Columbia, Canada. **^3^** MD Undergraduate Program, Faculty of Medicine, University of British Columbia, Vancouver, Canada. **^4^** Knowledge Translation Program, Li Ka Shing Knowledge Institute, St. Michael’s Hospital, Toronto, and the University of British Columbia, Vancouver, British Columbia, Canada.

**^5^** Department of Health Sciences and Technology, ETH Zurich, Zurich, Switzerland. **^6^** Schulthess Clinic, Zurich, Switzerland. **^7^** Centre for Human Brain Health, University of Birmingham, United Kingdom. **^8^** MRC Versus Arthritis Centre for Musculoskeletal Ageing Research, University of Birmingham, United Kingdom.

**^9^** Centre for Trauma Science Research, University of Birmingham, United Kingdom. **^10^** Department of Urology, University Hospital Basel, University of Basel, Basel, Switzerland. **^11^** Department of Medicine, Division of Physical Medicine and Rehabilitation, University of British Columbia, Vancouver, British Columbia, Canada. **^12^** GF Strong Rehabilitation Centre, Vancouver Coastal Health, Vancouver, British Columbia, Canada.

***Purpose:*** This supplementary file includes the pooled and individual participant demographics, injury characteristics and cardiorespiratory fitness (CRF) outcome data for cross-sectional studies comparing CRF levels between inactive and active individuals with a spinal cord injury (SCI). The purpose of this secondary meta-analysis was to investigate the potential long-term benefits of sustained, habitual physical activity and exercise on CRF.

***Conclusion:*** Active individuals with SCI exhibit a significantly higher absolute (AV̇O_2peak_) and relative peak oxygen consumption (RV̇O_2peak_), and peak power output (PPO) than inactive individuals with SCI. There was notable heterogeneity in studies assessing RV̇O_2peak,_ which is likely explained by the inclusion of elite athlete vs inactive individuals included in these comparisons. Cumulative participation in physical activity or exercise for a sustained period of time (i.e., individuals performing high levels of physical activity such as elite athletes) likely explains the higher CRF. It should be noted that these cross-sectional cohort comparison studies are prone to confounding and are inherently biased. Nevertheless, these studies provide insight into the impact of prolonged physical activity, which is rarely assessed in prospective longitudinal intervention studies. The longest intervention in our systematic review was 52 weeks, with many being much shorter than this.

| **Table 1:** Overview of participant demographics and injury characteristics from included cross-sectional cohort comparisons studies for each cardiorespiratory fitness outcome. | | |
| --- | --- | --- |
| **Absolute V̇O_2peak_ studies (N = 11)** | | |
|  | **Inactive (Σ participants = 134)** | **Active (Σ participants = 182)** |
| *Participant demographics* | | |
| Age (years) | 33 (27 – 41) | 30 (26 – 42) |
| Sex (M/F) | 118/16 (88%/12%) | 169/13 (93%/7%) |
| *Injury characteristics* | | |
| Neurological level of injury (TETRA/PARA) | 41/85 (31%/63%), Σ^NR^=8 (6%) | 28/137 (15%/75%), Σ^NR^=17 (10%) |
| Severity (complete/incomplete) | 31/1 (23%/1%), Σ^NR^=102 (76%) | 14/8 (8%/2%), Σ^NR^=160 (90%) |
| Time since injury (years) | 9 (5 – 13), Σ^NR^=38 (28%) | 11 (7 – 13), Σ^NR^=83 (46%) |
| *CRF outcome* | | |
| Absolute V̇O_2peak_ (L/min) | 1.19 (0.46 – 1.76) | 1.89 (0.92 – 2.42) |
| **Relative V̇O_2peak_ studies (N = 10)** | | |
|  | **Inactive (Σ participants = 123)** | **Active (Σ participants = 142)** |
| *Participant demographics* | | |
| Age (years) | 32 (27 – 39) | 38 (28 – 46) |
| Sex (M/F) | 107/16 (87%/13%) | 129/13 (83%/17%) |
| *Injury characteristics* | | |
| Neurological level of injury (TETRA/PARA) | 41/74 (33%/60%), Σ^NR^=8 (7%) | 28/97 (20%/68%), Σ^NR^=17 (12%) |
| Severity (complete/incomplete) | 31/1 (25%/1%), Σ^NR^=91 (74%) | 14/8 (10%/6%), Σ^NR^=120 (84%) |
| Time since injury (years) | 9 (5 – 13), Σ^NR^=27 (22%) | 11 (7 – 13), Σ^NR^=43 (14%) |
| *CRF outcome* | | |
| Relative V̇O_2peak_ (mL/kg/min) | 17.2 (5.5 – 25.2) | 28.4 (13.2 – 35.0) |
| **Peak power output studies (N = 6)** | | |
|  | **Inactive (Σ participants = 75)** | **Active (Σ participants = 108)** |
| *Participant demographics* | | |
| Age (years) | 33 (27 – 41) | 38 (25 – 48) |
| Sex (M/F) | 59/16 (79%/21%) | 95/13 (88%/12%) |
| *Injury characteristics* | | |
| Neurological level of injury (TETRA/PARA) | 16/59 (21%/79%) | 9/99 (8%/92%) |
| Severity (complete/incomplete) | 18/1 (24%/1%), Σ^NR^=75 (50%) | 7/4 (6%/4%)*,* Σ^NR^=97 (90%) |
| Time since injury (years) | 11 (8 – 13), Σ^NR^=31 (41%) | 11 (7 – 13), Σ^NR^=69 (64%) |
| *CRF outcome* | | |
| Peak power output (W) | 46 (18 – 63) | 80 (50 – 97) |
| Continuous variables are displayed as weighted means (range: lowest – highest mean values reported from studies). Categorical variables are displayed as n (%). Weighted means were calculated to account for differences in sample size between studies using the following formula: Σn*x̅ /Σn, where Σ = the sum of, n = number of participants in each study and, x̅ = mean CRF outcome of each study. F, females; M, males; NR, not reported; PARA, paraplegia; TETRA, tetraplegia; V̇O_2peak_, peak oxygen consumption; W, watts. | | |

**Table 2:** Summaries of individual cross-sectional cohort comparison studies between inactive and active individuals with spinal cord injury.

| **Author/year/**  **country** | **Category** | **Inactive** | **Active** |
| --- | --- | --- | --- |
| Dallmeijer et al. (1997)  The Netherlands | Population | N = 13 (13M/0F)  Age = 36 ± 9 years  TSI = 5 ± 4 years  Classification = 13 TETRA/0 PARA  Severity = 13 complete/ 0 incomplete | N = 11 (11M/0F)  Age = 29 ± 12 years  TSI = 7 ± 9 years  Classification = 11 TETRA/0 PARA  Severity = 7 complete/ 4 incomplete |
|  | Activity status | Sedentary | Physically active (n=11) with sports participation ranging from 1.5-6.0 hours/week. All subjects participated in quad rugby training (1.5 hours/week), with additional activities including wheeling (n=4), wheelchair dancing (n=1) and table tennis (n=1) |
|  | CRF outcomes | **AV̇O_2peak_ = 0.79 ± 0.22 L/min**  **RV̇O_2peak_ = 9.1 ± 2.5 mL/kg/min**  PPO = NR | **AV̇O_2peak_ = 1.01 ± 0.32 L/min**  **RV̇O_2peak_ = 14.4 ± 4.5 mL/kg/min**  PPO = NR |
|  | CPET modality | Maximal wheelchair ergometry protocol | |
| Davis et al. (1988)  USA | Population | N = 15 (15M/0F)  Age = 27 ± 8 years  TSI = 13 ± 11 years  Classification = 0 TETRA/15 PARA  Severity = NR | N = 15 (15M/0F)  Age = 28 ± 6 years  TSI = 13 ± 11 years  Classification = 0 TETRA/15 PARA  Severity = NR |
|  | Activity status | Sedentary (n=11) or participated in recreational activities, bowling, horse-riding and volleyball less than 3 times per week (n=4) | Seasonal track athletes exercising at least 4 times/week (n=8) or athletes participating in wheelchair basketball, competitive swimming or weightlifting at least 3 times/week (n=3). |
|  | CRF outcomes | **AV̇O_2peak_ = 1.56 ± 0.35 L/min**  RV̇O_2peak_ = 25.2 ± 5.6 mL/kg/min  **PPO = 62 ± 20 W** | **AV̇O_2peak_ = 2.24 ± 0.54 L/min**  RV̇O_2peak_ = 34.2 ± 8.3 mL/kg/min ^a^  **PPO = 97 ± 24 W** |
|  | CPET modality | Continuous multistage arm crank ergometry protocol | |
| Eriksson et al. (1988)  Sweden | Population | N = 22 (22M/0F)*  Age = 31 ± 8 years  TSI = 9 ± 6 years  Classification = 12 TETRA/10 PARA  Severity = NR  **Pooled from quadraplegic and paraplegic untrained groups* | N = 25 (25M/0F)  Age = 31.3 ± 5.0 years  TSI = 12.6 ± 7.9 years  Classification = 8 TETRA/17 PARA  Severity = NR |
|  | Activity status | Untrained individuals with tetraplegia or paraplegia ^b^ | Elite athletes with tetraplegia and paraplegia undertaking regular physical training for 8 ± 3 and 6 ± 7 years, respectively. |
|  | CRF outcomes | AV̇O_2peak_ = 1.21 ± 0.46 L/min  RV̇O_2peak_ = 17.4 ± 6.1 mL/kg/min | AV̇O_2peak_ = 1.82 ± 0.62 L/min  RV̇O_2peak_ = 28.4 ± 9.9 mL/kg/min |
|  | CPET modality | Maximal wheelchair ergometry protocol | |
| Gibbons *et al.* (2016)  England | Population | N = 6 (3M/3F)  Age = 36 ± 9 years  TSI = 12 ± 7 years  Classification = 3 TETRA/3 PARA  Severity = 6 complete/ 0 incomplete | N = 3 (3M/0F)  Age = 42 ± 15 years  TSI = 7 ± 3 years  Classification = 1 TETRA/2 PARA  Severity = 3 complete/ 0 incomplete |
|  | Activity status | Functional Electrical Stimulation-Untrained | Functional Electrical Stimulation-Trained |
|  | CRF outcomes | AV̇O_2peak_ = 0.99 ± 0.38 L/min  RV̇O_2peak_ = 15.3 ± 2.3 mL/kg/min  PPO = 63 ± 45 W | AV̇O_2peak_ = 1.35 ± 0.50 L/min  RV̇O_2peak_ = 18.3 ± 6.5 mL/kg/min  PPO = 77 ± 38 W |
|  | CPET modality | Incremental arm crank ergometry protocol | |
| Hopman et al. (1996)  The Netherlands | Population | N = 13 (10M/3F)  Age = 31 ± 10 years  TSI = 8 ± 5 years  Classification = 13 TETRA/0 PARA  Severity = 12 complete/ 1 incomplete | N = 8 (8M/0F)  Age = 33 ± 13 years  TSI = 8 ± 10 years  Classification = 8 TETRA/0 PARA  Severity = 4 complete/ 4 incomplete |
|  | Activity status | Untrained (n=7), had not participated in any sport for the previous 2 years but performed low volume team sport training (quad rugby, 1/wk, 2 games/month). Sedentary (n = 6), did not participate in any sport ^c^ | Trained (n=8) had participated in sport 2 times/week for the previous 2 years |
|  | CRF outcomes | AV̇O_2peak_ = 0.62 ± 0.22 L/min  RV̇O_2peak_ = 7.8 ± 2.8 mL/kg/min  PPO = 18 ± 14 W | AV̇O_2peak_ = 1.03 ± 0.42 L/min  RV̇O_2peak_ = 14.0 ± 5.7 mL/kg/min  PPO = 50 ± 29 W |
|  | CPET modality | Incremental arm crank ergometry protocol | |
| Huonker et al. (1998)  Germany | Population | N = 20 (20M/0F)  Age = 34 ± 10 years  TSI = NR  Classification = 0 TETRA/20 PARA  Severity = NR | N = 29 (29M/0F)  Age = 32 ± 7 years  TSI = NR  Classification = 0 TETRA/29 PARA  Severity = NR |
|  | Activity status | Sedentary | Paralympic athletes: cross-country sledding (n=12), wheelchair racing (n=10), wheelchair basketball (n=7) |
|  | CRF outcomes | AV̇O_2peak_ = 1.76 ± 0.28 L/min  RV̇O_2peak_ = 23.9 ± 3.8 mL/kg/min  PPO = 50 ± 18 W | AV̇O_2peak_ = 2.42 ± 0.30 L/min  RV̇O_2peak_ = 34.5 ± 4.3 mL/kg/min  PPO = 89 ± 16 W |
|  | CPET modality | Incremental wheelchair ergometry test | |
| Okuma et al. (1989)  Japan | Population | N = 7 (7M/0F)  Age = 31 ± 8 years  TSI = NR  Classification = 0 TETRA/7 PARA  Severity = NR | N = 14 (14M/0F)*  Age = 31 ± 7 years  TSI = NR  Classification = 0 TETRA/10 PARA  Severity = NR  **N=4 non-SCI* |
|  | Activity status | Non-athletic/did not play any sports | Wheelchair marathon competitors |
|  | CRF outcomes | **AV̇O_2peak_ = 1.32 ± 0.26 L/min**  **RV̇O_2peak_ = 22.6 ± 5.0 mL/kg/min**  PPO = NR | **AV̇O_2peak_ = 1.80 ± 0.36 L/min**  **RV̇O_2peak_ = 35.0 ± 4.8 mL/kg/min**  PPO = NR |
|  | CPET modality | Maximal treadmill wheelchair test | |
| Ramkrapes et al. (2021)  Brazil | Population | N = 8 (8M/0F)  Age = 38.50 ± 6.02 years  TSI = 12.25 ± 6.73 years  Classification = 8 TETRA/0 PARA  Severity = NR | N = 13 (13M/0F)  Age = 33.27 ± 5.62 years  TSI = 12.36 ± 2.73 years  Classification = 13 TETRA/0 PARA  Severity = NR |
|  | Activity status | Categorised as insufficiently active (below 14 units) via the Godin-Shephard Leisure-Time Physical Activity Questionnaire. | Categorised as physically active (above 24 units) via the Godin-Shephard Leisure-Time Physical Activity Questionnaire. |
|  | CRF outcomes | AV̇O_2peak_ = 0.46 ± 0.17 L/min  RV̇O_2peak_ = 5.53 ± 2.32 mL/kg/min  PPO = NR | AV̇O_2peak_ = 0.92 ± 0.26 L/min  RV̇O_2peak_ = 13.15 ± 3.73 mL/kg/min  PPO = NR |
|  | CPET modality | Incremental arm crank ergometry protocol | |
| Rawashdeh et al. (2000)  Germany | Population | **INACTIVE**  N = 11 (11M/0F)  Age = 41 ± 14 years  TSI = NR  Classification = 0 TETRA/11 PARA  Severity = NR | **ACTIVE (COMBINED RECREATIONAL AND ENDURANCE ATHLETES)**  N=40 (40M/0F)  Age = 26 ± 9 years  TSI = NR  Classification = 0 TETRA/40 PARA  Severity = NR |
|  | Activity status | **INACTIVE**  Participants were categorised as inactive (0.2 ± 0.4 sport hours per week) | **ACTIVE (COMBINED RECREATIONAL AND ENDURANCE ATHLETES)**  Participants were categorised as recreational athletes by undertaking 3.5 ± 1.5 hours of sport per week or categorised as endurance athletes by undertaking 8.8 ± 4.6 hours of sport per week. |
|  | CRF outcomes | **INACTIVE**  **AV̇O_2peak_ = 1.30 ± 0.40 L/min**  **RV̇O_2peak_ = NR**  **PPO = 38.5 ± 15 W** | **ACTIVE (COMBINED RECREATIONAL AND ENDURANCE ATHLETES)**  **AV̇O_2peak_ = 2.15 ± 0.53 L/min**  **RV̇O_2peak_ = NR**  **PPO = 81 ± 22 W** |
|  | CPET modality | Incremental wheelchair ergometry test | |
| Schmid et al. (1998)  Germany | Population | N = 10 (0M/10F)  Age = 30 ± 13 years  TSI = 12 ± 16 years  Classification = 0 TETRA/10 PARA  Severity = NR | N = 13 (0M/13F)  Age = 28 ± 20 years  TSI = 13 ± 23 years  Classification = 0 TETRA/13 PARA  Severity = NR |
|  | Activity status | Sedentary | German national wheelchair basketball athletes |
|  | CRF outcomes | AV̇O_2peak_ = 1.09 ± 0.62 L/min  **RV̇O_2peak_ = 18.3 ± 10.4 mL/kg/min**  **PPO = 46 (NR)** | AV̇O_2peak_ = 1.90 ± 1.06 L/min ^a^  **RV̇O_2peak_ = 33.7 ± 18.8 mL/kg/min**  **PPO = 60 (NR)** |
|  | CPET modality | Discontinuous graded maximal wheelchair ergometry test | |
| Zwiren et al. (1975)  Israel | Population | N = 9 (9M/0F)  Age = 29 ± 10 years  TSI = 7 (2 – 23) years  Classification = 0 TETRA/9 PARA  Severity = NR | N = 11 (11M/0F)  Age = 28 ± 8 years  TSI = 13 (2 – 27) years  Classification = 0 TETRA/11 PARA  Severity = NR |
|  | Activity status | Sedentary (not undergoing any physical training) | International wheelchair bound athletes regularly engaged in vigorous training |
|  | CRF outcomes | **AV̇O_2peak_ = 1.38 ± 0.32 L/min**  **RV̇O_2peak_ = 19.6 ± 5.5 mL/kg/min**  PPO = NR | **AV̇O_2peak_ = 2.07 ± 0.41 L/min**  **RV̇O_2peak_ = 35.0 ± 7.6 mL/kg/min**  PPO = NR |
|  | CPET modality | Incremental arm crank ergometry test | |

Continuous variables are displayed as mean ± standard deviation. Categorical variables are displayed as n. Cardiorespiratory fitness (CRF) outcomes in bold indicate a significant difference between active and inactive subgroups (*p* < 0.05). AV̇O_2peak_, absolute peak oxygen consumption; F, females; M, males; NR, not reported; PARA, paraplegia; PPO, peak power output; RV̇O_2peak_, relative peak oxygen consumption; TETRA, tetraplegia; TSI, time since injury. ^a^ Calculated using body mass (kg) provided in the paper (therefore no significant differences reported). ^b^ No significant differences reported for CRF outcomes as the inactive cohort was combined from multiple subgroups. ^c^ No significant differences reported for CRF outcomes as untrained and sedentary cohorts were combined to form the inactive cohort.

| **Table 3.** Risk of bias of cross-sectional cohort comparison studies assessed via the NIH Quality Assessment Tool for Observational Cohort and Cross-Sectional Studies (14 items). | | | | | | | | | | | | | | | |
| --- | --- | --- | --- | --- | --- | --- | --- | --- | --- | --- | --- | --- | --- | --- | --- |
| **Author (Year)** | **Q1** | **Q2** | **Q3** | **Q4** | **Q5** | **Q6** | **Q7** | **Q8** | **Q9** | **Q10** | **Q11** | **Q12** | **Q13** | **Q14** | **Overall Quality** |
| Dallmeijer et al. (1997) | **✓** | ❌ | **?** | **✓** | ❌ | ❌ | ❌ | **✓** | **✓** | ❌ | **✓** | **?** | **N/A** | **✓** | **GOOD** |
| Davis et al. (1988) | **✓** | ❌ | **?** | **?** | ❌ | ❌ | ❌ | **✓** | **✓** | ❌ | **✓** | **?** | **N/A** | ❌ | **FAIR** |
| Eriksson et al. (1988) | ❌ | **✓** | **?** | **✓** | ❌ | ❌ | ❌ | **✓** | **✓** | ❌ | **✓** | ❌ | **N/A** | ❌ | **FAIR** |
| Gibbons et al. (2016) | **✓** | **✓** | **?** | **?** | ❌ | ❌ | ❌ | **✓** | **✓** | ❌ | **✓** | **?** | **N/A** | ❌ | **FAIR** |
| Hopman et al. (1996) | **✓** | **✓** | **?** | **?** | ❌ | **✓** | **✓** | **✓** | **✓** | **✓** | **✓** | ❌ | **N/A** | ❌ | **POOR** |
| Huonker et al. (1998) | **✓** | ❌ | **?** | **?** | ❌ | ❌ | ❌ | **✓** | **✓** | ❌ | **✓** | **?** | **N/A** | ❌ | **FAIR** |
| Okuma et al. (1989) | ❌ | **✓** | **?** | **?** | ❌ | ❌ | ❌ | **✓** | **✓** | **✓** | **✓** | **?** | **N/A** | ❌ | **FAIR** |
| Ramkrapes et al. (2021) | **✓** | **✓** | **?** | **?** | **✓** | ❌ | ❌ | **✓** | **✓** | ❌ | **✓** | **?** | **N/A** | **✓** | **GOOD** |
| Rawashdeh et al. (2000) | ❌ | ❌ | **?** | **?** | ❌ | ❌ | ❌ | **✓** | ❌ | ❌ | ❌ | **?** | **N/A** | ❌ | **POOR** |
| Schmid et al. (1998) | **✓** | ❌ | **?** | **?** | ❌ | ❌ | ❌ | **✓** | **✓** | ❌ | **✓** | **?** | **N/A** | ❌ | **FAIR** |
| Zwiren et al. (1975) | **✓** | **✓** | **?** | **?** | ❌ | ❌ | ❌ | **✓** | **✓** | ❌ | **✓** | **?** | **N/A** | ❌ | **FAIR** |
| **Total ✓** | **8** | **6** | **0** | **2** | **1** | **1** | **1** | **11** | **10** | **2** | **10** | **0** | **0** | **2** |  |
| **Total** ❌ | **3** | **5** | **0** | **0** | **10** | **10** | **10** | **0** | **1** | **9** | **1** | **2** | **0** | **9** |  |
| **Total ?** | **0** | **0** | **11** | **9** | **0** | **0** | **0** | **0** | **0** | **0** | **0** | **9** | **0** | **0** |  |
| **✓** = yes, ❌ = no, **?** = cannot determine/not reported | | | | | | | | | | | | | | | |

**Quality Assessment Questions**

1. Was the research question or objective in this paper clearly stated?
2. Was the study population clearly specified and defined?
3. Was the participation rate of eligible persons at least 50%?
4. Were all the subjects selected or recruited from the same or similar populations (including the same time period)? Were inclusion and exclusion criteria for being in the study prespecified and applied uniformly to all participants?
5. Was a sample size justification, power description, or variance and effect estimates provided?
6. For the analyses in this paper, were the exposure(s) of interest measured prior to the outcome(s) being measured?
7. Was the timeframe sufficient so that one could reasonably expect to see an association between exposure and outcome if it existed?
8. For exposures that can vary in amount or level, did the study examine different levels of the exposure as related to the outcome (e.g., categories of exposure, or exposure measured as continuous variable)?
9. Were the exposure measures (independent variables) clearly defined, valid, reliable, and implemented consistently across all study participants?
10. Was the exposure(s) assessed more than once over time?
11. Were the outcome measures (dependent variables) clearly defined, valid, reliable, and implemented consistently across all study participants?
12. Were the outcome assessors blinded to the exposure status of participants?
13. Was loss to follow-up after baseline 20% or less?
14. Were key potential confounding variables measured and adjusted statistically for their impact on the relationship between exposure(s) and outcome(s)?

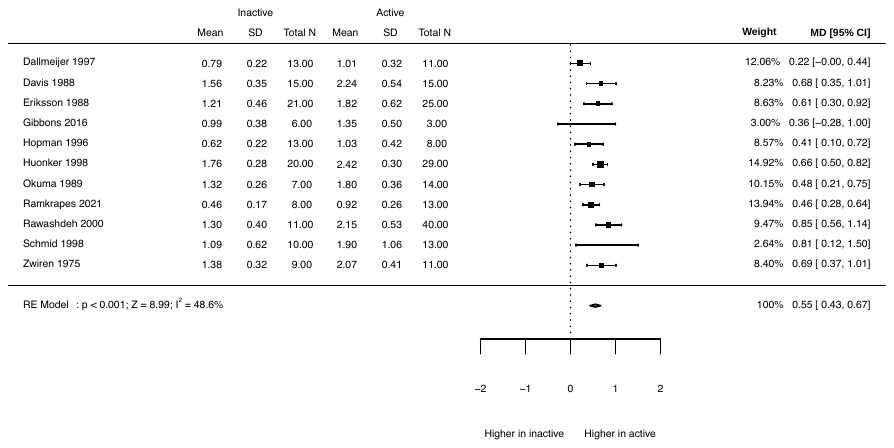

**Figure 1.** Cross-sectional cohort comparison of absolute peak oxygen consumption (L/min) in inactive and active individuals with spinal cord injury.

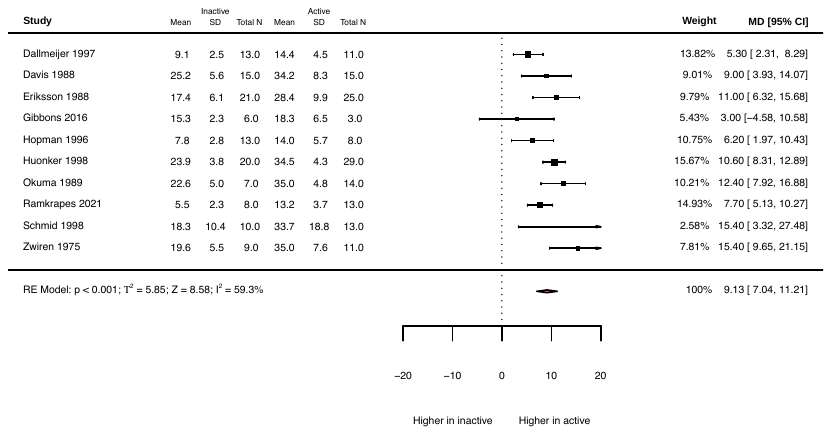

**Figure 2.** Cross-sectional cohort comparison of relative peak oxygen consumption (mL/kg/min) in inactive and active individuals with spinal cord injury.

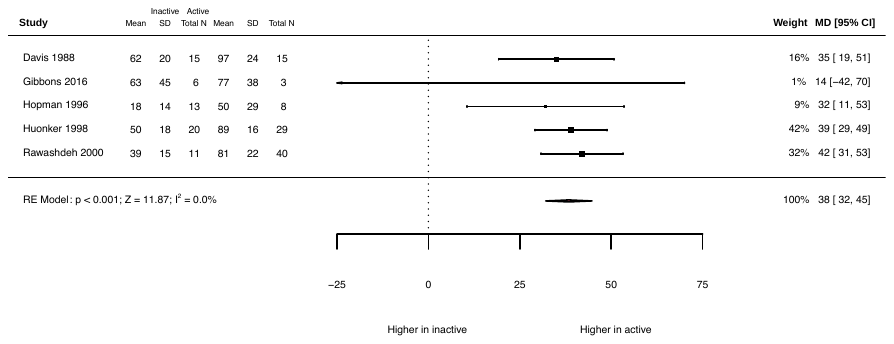

**Figure 3.** Cross-sectional cohort comparison of peak power output (W) in inactive and active individuals with spinal cord injury.

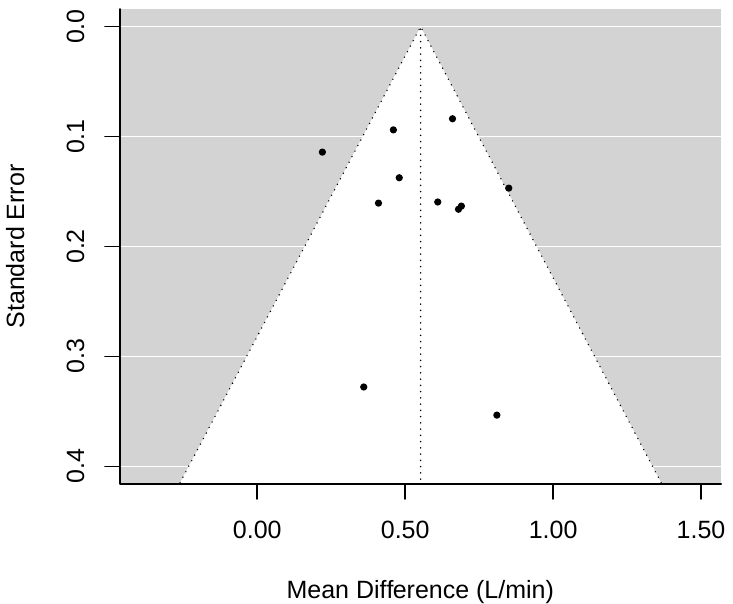

**Figure 4.** Funnel plot of cross-sectional cohort studies comparing absolute peak oxygen consumption between inactive and active individuals. Egger’s test for funnel plot asymmetry was not significant (Z = 0.47, *p* = 0.64).

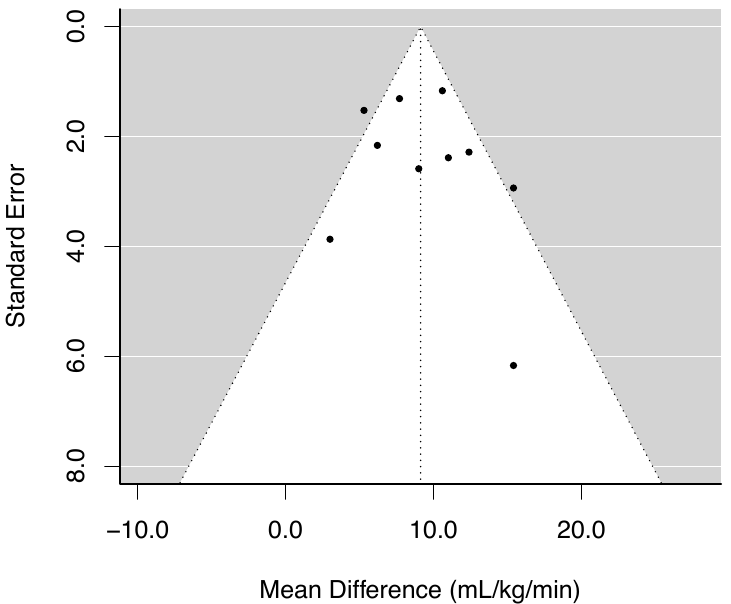

**Figure 5.** Funnel plot of cross-sectional cohort studies comparing relative peak oxygen consumption between inactive and active individuals. Egger’s test for funnel plot asymmetry was not significant (Z = 0.78, *p* = 0.43).

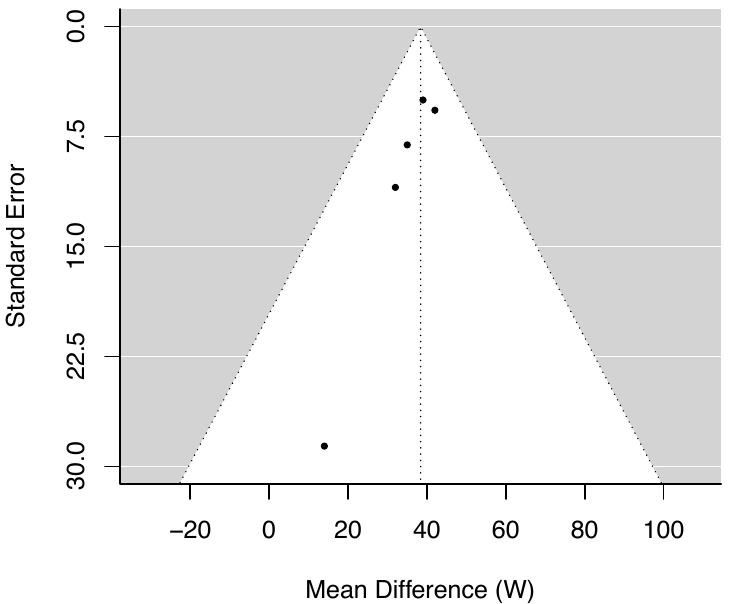

**Figure 6.** Funnel plot of cross-sectional cohort studies comparing peak power output between inactive and active individuals. Egger’s test not performed (<10 studies).

**Sensitivity analysis**

We conducted a sensitivity analysis, whereby we ran separate meta-analyses comparing either inactive individuals vs. active individuals, or inactive individuals vs. elite-level athletes with a spinal cord injury. These were determined based on the descriptions provided in the methods of each study. There was a significantly higher RV̇O_2peak_ [4.7 (2.6, 6.7) mL/kg/min, p<0.001] in ‘active’ compared to inactive individuals (Figure 7), but an even higher RV̇O_2peak_ [11.2 (9.6, 12.9) mL/kg/min, p<0.001] in ‘elite athletes’ compared to inactive (Figure 8).

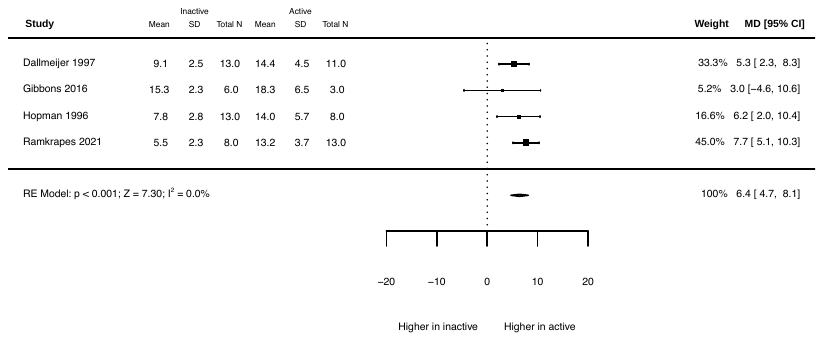

**Figure 7.** A sensitivity analysis of cross-sectional cohort studies comparing relative peak oxygen consumption (mL/kg/min) between inactive and active individuals with a spinal cord injury.

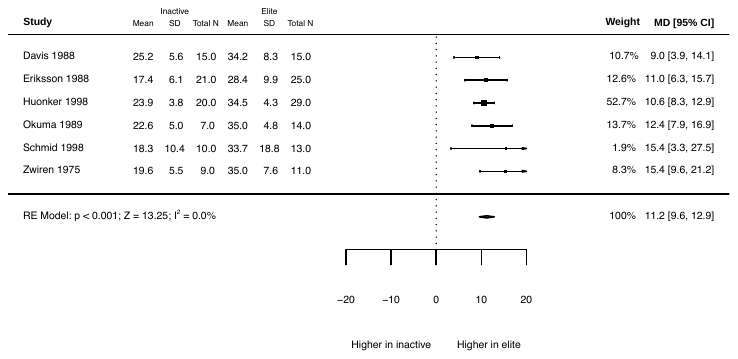

**Figure 8.** A sensitivity analysis of cross-sectional cohort studies comparing relative peak oxygen consumption (mL/kg/min) between inactive individuals and elite-level athletes with spinal cord injury.

**References**

Cross-sectional cohort studies included in the systematic review, sorted alphabetically:

Dallmeijer AJ, Hopman MTE, van der Woude LHV. Lipid, lipoprotein, and apolipoprotein profiles in active and sedentary men with tetraplegia. Arch Phys Med Rehabil 1997;78(11):1173-6.

Davis GM, Shephard RJ. Cardiorespiratory fitness in highly active versus inactive paraplegics. Med Sci Sports Exerc 1988;20(5):463-8.

Eriksson P, Lofstrom L, Ekblom B. Aerobic power during maximal exercise in untrained and well-trained persons with quadriplegia and paraplegia. Scand J Rehab Med 1988;20:141-7.

Gibbons RS, Stock CG, Andrews BJ, Gall A, Shave RE. The effect of FES-rowing training on cardiac structure and function: pilot studies in people with spinal cord injury. Spinal Cord 2016;54(10):822-9.

Hoevenaars D, Holla JFM, Postma K, van der Woude LHV, Janssen TWJ, de Groot S. Associations between meeting exercise guidelines, physical fitness, and health in people with spinal cord injury. Disabil Rehabil 2023;45(6):1030-1037.

Hopman MTE, Dallmeijer, Snoek G, AJ, van der Woude LHV. The effect of training on cardiovascular responses to arm exercise in individuals with tetraplegia. Eur J Appl Physiol Occup Physiol 1996;74:172-9.

Huonker M, Schmid A, Sorichter S, Schmid-Trucksäb A, Mrosek P, Keul J. Cardiovascular differences between sedentary and wheelchair-trained subjects with paraplegia. Med Sci Sports Exerc 1998;30(4):609-13.

Okuma H, Ogata H, Hatada K. Transition of physical fitness in wheelchair marathon competitors over several years. Paraplegia 1989;27(3):237-43.

Ramkrapes APB, Duft RG, Bonfante ILP, Mateus KCS, Trombeta JCS, Rodrigues B, et al. Higher Physical Activity Level Improves Leptin Concentrations in Spinal Cord Injury Subjects. Biomed Res Int. 2021;2021:9415253.

Rawashdeh VM, Heitkamp HC, Jeschke AD. [Dynamic performance of paraplegics as a function of physical training]. MMW Fortschritte der Medizin. 2000;142(16):38-40.

Schmid A, Huonker M, Stober P, Barturen JM, Schmidt-Trucksäss A, Dürr H et al. Physical performance and cardiovascular and metabolic adaptation of elite female wheelchair basketball players in wheelchair ergometry and in competition. Am J Phys Med Rehabil 1998;77(6):527-33.

Zwiren LD, Bar-Or, O. Responses to exercise of paraplegics who differ in conditioning level. Med Sci Sports 1975;7(2):94-8.
