## Supplementary material for "The effect of exercise on aerobic capacity in individuals with spinal cord injury: A systematic review with meta-analysis and meta-regression": S12 Cross-sectional associations

**Electronic Supplementary Material 12**

**PLoS Medicine**

Hodgkiss, D.D^1^, Bhangu, G^2,3^, Lunny, C^4^, Jutzeler C.R^5,6^, Chiou S.Y^1,7,8,9^ Walter, M^2,10^, Lucas S.E^1,7^, Krassioukov, A.V.^2,11,12^, Nightingale, T.E.^1,2,9^

**^1^** School of Sport, Exercise and Rehabilitation Sciences, University of Birmingham, UK.

**^2^** International Collaboration on Repair Discoveries (ICORD), University of British Columbia, Vancouver, British Columbia, Canada. **^3^** MD Undergraduate Program, Faculty of Medicine, University of British Columbia, Vancouver, Canada. **^4^** Knowledge Translation Program, Li Ka Shing Knowledge Institute, St. Michael’s Hospital, Toronto, and the University of British Columbia, Vancouver, British Columbia, Canada.

**^5^** Department of Health Sciences and Technology, ETH Zurich, Zurich, Switzerland. **^6^** Schulthess Clinic, Zurich, Switzerland. **^7^** Centre for Human Brain Health, University of Birmingham, United Kingdom. **^8^** MRC Versus Arthritis Centre for Musculoskeletal Ageing Research, University of Birmingham, United Kingdom.

**^9^** Centre for Trauma Science Research, University of Birmingham, United Kingdom. **^10^** Department of Urology, University Hospital Basel, University of Basel, Basel, Switzerland. **^11^** Department of Medicine, Division of Physical Medicine and Rehabilitation, University of British Columbia, Vancouver, British Columbia, Canada. **^12^** GF Strong Rehabilitation Centre, Vancouver Coastal Health, Vancouver, British Columbia, Canada.

***Purpose:*** This supplementary file includes the pooled and individual participant demographics, injury characteristics and cardiorespiratory fitness (CRF) outcome data for cross-sectional studies investigating the association between habitual physical activity level and CRF. We graphically present individual study association data for each CRF outcome to help capture the importance of the volume and intensity of physical activity in individuals with a spinal cord injury (SCI).

***Conclusion:*** There is only one study using a validated wearable device to monitor habitual physical activity exposure in SCI. This study outlines the CRF benefits of performing moderate-to-vigorous physical activity, as opposed to lower levels of activity. Whilst self-report methods of physical activity are valid, reliable and cost-effective, accurately monitoring physical activity via validated wearable devices should be encouraged.

**Table 1:** Overview of participant demographics and injury characteristics from included cross-sectional studies comparing associations between physical activity dimensions and cardiorespiratory fitness outcomes

| **Absolute V̇O_2peak_ studies (N = 4)** | |
| --- | --- |
| **Σ participants = 422** | |
| *Participant demographics* | |
| Age (years) | 41 (34 – 46) |
| Sex (M/F), n (%) | 341/80 (81%/19%), Σ^NR^=1 (<1%) |
| *Injury characteristics* |  |
| Neurological level of injury (TETRA/PARA),  n (%) | 75/180 (18%/43%), Σ^NR^=167 (40%) |
| Severity (complete/incomplete), n (%) | 193/41 (46%/10%*)*, Σ^NR^=188 (44%) |
| Time since injury (years) | 14 (8 – 19) |
| *Physical activity outcome* | |
| Method (self-reported/wearable device), N | N=3 self-reported, N=1 wearable device |
| *CRF outcome* | |
| Absolute V̇O_2peak_ (L/min) | 1.45 (1.34 – 1.90) |
| **Relative V̇O_2peak_ studies (N = 7)** | |
| **Σ participants = 581** | |
| *Participant demographics* | |
| Age (years) | 41 (34 – 48) |
| Sex (M/F), n (%) | 463/117 (80%/20%), Σ^NR^=1 (<1%) |
| *Injury characteristics* | |
| Neurological level of injury (TETRA/PARA),  n (%) | 140/274 (24%/47%), Σ^NR^=167 (29%) |
| Severity (complete/incomplete), n (%) | 320/73 (55%/13%), Σ^NR^=188 (32%) |
| Time since injury (years) | 15 (8 – 29) |
| *Physical activity outcome* | |
| Method (self-reported/wearable device), N | N=6 self-reported, N=1 wearable device |
| *CRF outcome* | |
| Relative V̇O_2peak_ (mL/kg/min) | 18.2 (13.9 – 24.0) *Σ^NR^=45 (12%) due to males only (Janssen et al., 2002) and exclusion of participants (Latimer et al., 2006) |
| **Peak power output studies (N = 5)** | |
| **Σ participants = 512** | |
| *Participant demographics* | |
| Age (years) | 41 (34 – 46) |
| Sex (M/F), n (%) | 400/111 (78%/22%), Σ^NR^=1 (<1%) |
| *Injury characteristics* | |
| Neurological level of injury (TETRA/PARA),  n (%) | 127/218 (25%/43%), Σ^NR^=167 (33%) |
| Severity (complete/incomplete), n (%) | 273/73 (53%/14%), Σ^NR^=166 (33%) |
| Time since injury (years) | 14 (8 – 19) |
| *Physical activity outcome* | |
| Method (self-reported/wearable device), N | N=3 self-reported |
| *CRF outcome* | |
| Peak power output [W] | 54 (51 – 80)*Σ^NR^=45 (16%) due to males only (Janssen et al., 2002) and exclusion of participants (Latimer et al., 2006) |
| Total number of studies (N) and sum of participants (Σ). Continuous variables (age, TSI and CRF outcomes) are displayed as weighted means (range: lowest – highest mean values reported from studies). Categorical variables are displayed as n (%). Weighted means were calculated to account for differences in sample size between studies using the following formula: Σn*x̅ /Σn, where Σ = the sum of, n = number of participants in each study and, x̅ = mean CRF outcome of each study. NR, not reported. | |

**Table 2:** Summaries of individual cross-sectional comparing association between physical activity and cardiorespiratory fitness outcomes

| **Author/year/ country** | **Population** | **Physical activity outcome description** | **Statistical analysis** | **Findings** |
| --- | --- | --- | --- | --- |
| Hoevenaars et al. (2023)  The Netherlands | **INACTIVE**  N = 42 (26M/16F)  Age = 47.57 ± 10.06 years  TSI = 19.88 ± 10.79 years  Classification = 14 T/27 P  (1 CD)  Severity = 34 comp./ 8 incomp.  *Note: Demographics are reported for individuals with V̇O_2peak_* *data. PPO data was collected in N=38 (Inactive), N=31 (MG guidelines) and N=115 (TW guidelines), of which these demographics are captured in Table 1 above.*  **ACTIVE (MARTIN-GINIS GUIDELINES)**  N = 40 (25M/15F)  Age = 46.29 ± 11.53 years  TSI = 19.11 ± 10.77 years  Classification = 18 T/22 P  Severity = 31 comp./ 9 incomp.  **ACTIVE (TWEEDY GUIDELINES)**  N = 119 (95M/23F)  Age = 44.88 ± 9.24 years  TSI = 17.77 ± 9.85 years  Classification = 43 T/76 P  Severity = 99 comp./ 20 incomp. | **INACTIVE**  Not meeting any exercise guidelines (do not meet the recommended 40 mins/wk).  **ACTIVE (MARTIN-GINIS GUIDELINES)**  Participants exceeding 40 mins/wk of moderate and/or vigorous exercise were categorised as meeting the fitness exercise guidelines of Martin-Ginis et al. (2018). Participants exceeding 90 mins/wk of moderate and/or vigorous exercise were categorised as meeting the cardiometabolic exercise guidelines of Martin-Ginis et al. (2018). These two cohorts were pooled for analysis (MG aerobic guidelines).  **ACTIVE (TWEEDY GUIDELINES)**  Participants whose weekly time spent on moderate exercise exceeded 150 mins or on vigorous exercise exceeded 60 mins, were categorised as meeting the exercise guidelines of Tweedy et al. (2017). A combination of moderate and vigorous exercise was allowed for participants to meet the Tweedy guidelines by multiplying vigorous exercise duration by 2.5 and summing with the total duration of moderate exercise. | Multiple regression analyses were performed to evaluate if meeting SCI-specific aerobic exercise guidelines were associated with fitness and health outcomes. Multiple regressions were performed twice on each fitness and health outcome variable. Once with the MG guidelines group and once with the inactive group as the reference category to allow for multiple comparisons. Outcomes were adjusted for the potential confounders of age, sex, TSI, lesion level, lesion completeness, aetiology of SCI, and educational level. Findings are presented as β (SE) and *p*-value.  One-way ANOVAs were performed with Bonferroni post-hoc pairwise comparisons to identify differences in CRF outcomes between groups. | **INACTIVE**  AV̇O_2peak_ = 1.17 ± 0.45 L/min  RV̇O_2peak_ = 14.1 ± 4.4 mL/kg/min  PPO = 40.8 ± 21.8 W  **ACTIVE (MARTIN-GINIS GUIDELINES)**  AV̇O_2peak_ = 1.20 ± 0.40 L/min  RV̇O_2peak_ = 15.6 ± 5.8 mL/kg/min  PPO = 42.9 ± 19.2 W  **ACTIVE (TWEEDY GUIDELINES)**  AV̇O_2peak_ = 1.44 ± 0.57 L/min  RV̇O_2peak_ = 18.2 ± 7.2 mL/kg/min  PPO = 56.0 ± 26.3 W  *Findings from the multiple regression analyses (with inactive group as a reference)*   - **There were significant differences in AV̇O_2peak_ [0.31 (0.09), *p*=0.001], RV̇O_2peak_ [4.49 (1.20), *p*<0.001] and PPO [18.78 (4.59), *p*<0.001] between the inactive group and the Tweedy guidelines group.** - There were no differences in AV̇O_2peak_ [0.10 (0.11), *p*=0.389], RV̇O_2peak_ [2.46 (1.45), *p*=0.091] or PPO [7.93 (5.77), *p*=0.171] between the inactive group and the Martin-Ginis guidelines group.   *Findings from the multiple regression analyses (with Martin-Ginis guidelines group as a reference)*   - **There were significant differences in AV̇O_2peak_ [0.21 (0.09), *p*=0.022] and PPO [10.85 (4.82), *p*=0.026] between the Martin-Ginis guidelines group and the Tweedy guidelines group.** - There was no difference in RV̇O_2peak_ [2.03 (1.19), *p*=0.089] between the Martin-Ginis guidelines group and the Tweedy guidelines group. |
| Janssen et al. (2002)  The Netherlands | *N =* 166 (146M/20F)*  *Age* = *M*: 34 ± 11 yrs  *F*: 36 ± 12 yrs  *TSI* = *M*: 9 ± 9 yrs  *F*: 6 ± 7 yrs  *Classification* = NR  *Severity* = NR  *AV̇O_2peak_* = 1.52 ± 0.67 L/min  *RV̇O_2peak_* = 21.6 ± 10.7 ml/kg/min  PPO = 58 ± 32  *CPET* = Graded wheelchair exercise test  **precise number of participants for each correlation varies as a result of missing data. Participant demographics are not provided for each subsample.* | *Method* = self-reported but precise measurement tool NR  *PA dimensions* = Hours of sport per week. As this was not recorded for all participants, an activity level was assigned to the participants based on the hours of sport participation (if known) and/or the knowledge they were elite athletes using the following dichotomisation:   1. Sedentary (0 hr/wk) 2. Moderately active (1 – 3 hr/wk) 3. Active (3 – 6 hr/wk), or 4. Very active/athlete (>6 hr/wk)   *Measurement period* = NR | Two-tailed Pearson correlation coefficients were calculated between hours of sport per week and CRF (only men were included in this analysis due to the insufficient number of females).  Two stepwise multiple linear regressions were performed to estimate the most important determinants of CRF (including age, gender, lesion level, TSI, body mass and BMI). Regression 1 (n = 156-162) included activity level as the independent PA exposure variable, whereas regression 2 (n=91 – 92) included hours of sport per week. | - ***AV̇O_2peak_*: Significant (P<0.01) correlation with hours of sport per week (*r*=0.46).** - ***RV̇O_2peak_*: Significant (P<0.01) correlation with hours of sport per week (*r*=0.58).** - ***PPO*: Significant (P<0.05) correlation with hours of sport per week (*r*=0.51).** - *Regression analyses 1*: activity level explained an extra 12% of the variance in AV̇O_2peak_ and PPO, after adjusting for lesion level (most important determinant). Each activity level higher was associated with a 0.2 L/min higher AV̇O_2peak_ and 9W higher PPO. - *Regression analyses 2*: hours of sport per week explained an extra 11% and 12% of the variance in AV̇O_2peak_ and PPO, after accounting for lesion level, which was the most important CRF predictor (independently explaining 46% and 57% of the variance in AV̇O_2peak_ and PPO, respectively). |
| Latimer et al. (2006)  Canada | *N =* 73 (52M/21F)***  *Age* = 39 ± 11 yrs  *TSI* = 11 ± 10 yrs  *Classification* = 37 T/36 P  *Severity* = 41 comp./ 32 incomp.  *AV̇O_2peak_* = NR  *RV̇O_2peak_* = 14.9 ± 5.6 ml/kg/min  *PPO* = 60 ± 33  *CPET* = Incremental exercise protocol on an ACE  **actual sample size for CRF parameters was N = 48 due to exclusion of participants that did not reach V̇O_2peak_ (i.e., RER ≥1.00). Participant demographics are not provided for this subsample.* | *Method* = PARA-SCI  *PA dimensions* = Mild, Moderate, Heavy and Total min/day of LTPA (‘*included all structured and unstructured PA done during respondents free time’*), Lifestyle activity (‘*all other reported physical activities*’) and cumulative activity  *Measurement period* = previous 3 days | One-tailed bivariate correlations were calculated between PARA-SCI scores and CRF.  *Covariate* = Lesion level  Due to the moderating effects of lesion level, CRF outcomes were standardised (converted to a z-score). | - ***RV̇O_2peak_*: Significant correlations with Heavy LTPA (*r*=0.35, P<0.01), as well as Moderate (*r*=0.26, P<0.05) and Heavy (*r*=0.33, P<0.01) cumulative activity.** - ***PPO*: Significant correlations with Moderate (*r*=0.29, P<0.05), Heavy (*r*=0.33, P<0.01) and Total (*r*=0.34, P<0.01) LTPA. Significant correlation (P<0.01) with Heavy cumulative activity (*r*=0.34).** - No significant associations between CRF outcomes and Mild LTPA, Lifestyle activity or cumulative activity. |
| Lannem et al. (2010)  Norway | *N =* 47 (41M/6F) ***  *Age* = 48 ± 8 yrs  *TSI* = 29 ± 5 yrs  *Classification* = 13 T/34 P  *Severity* = 47 comp./ 0 incomp.  *AV̇O_2peak_* = NR  *RV̇O_2peak_* = 17.2 ± 6.5 ml/kg/min ^b^  *PPO* = NR  *CPET* = Incremental exercise protocol on an ACE  **total study sample size was N=116 but VO_2 peak_ data was presented for participants with AIS A-B only* | *Method* = self-reported but precise measurement tool NR  *PA dimensions* = Exercise hours per week. Participants also reported the frequency of exercise in 19 defined activities (NR). Participants were subsequently classified as:   1. Exercisers (> once/wk) 2. Non-exercisers (< once/wk)   *Measurement period* = NR | Spearman’s correlations (rho) were calculated to study the association between exercise hours and CRF. Values were provided by authors for N=47 and subgroups of tetraplegia (N=13) and paraplegia (N=34). | - ***RV̇O_2peak_*: Significant (P<0.001) correlation with exercise hours per week (rho=0.80) within the whole group.** - ***RV̇O_2peak_*: Significant (P<0.001) correlation with exercise hours per week (rho=0.87) for tetraplegia.** - ***RV̇O_2peak_*: Significant (P<0.001) correlation with exercise hours per week (rho=0.77) for paraplegia.** |
| Manns et al. (2005)  USA | *N =* 22 (22M/0F)  *Age* = 39 ± 9 yrs  *TSI* = 17 ± 9 yrs  *Classification* = 0 T/22 P  *Severity* = NR  *AV̇O_2peak_* = 1.90 ± 0.60 L/min  *RV̇O_2peak_* = 24.0 ± 8.5 ml/kg/min  *PPO* = NR  *CPET* = Incremental exercise protocol on an ACE | *Method* = PADS  *PA dimensions* = Total activity score, derived from three subscales: Exercise (8 items), LTPA (7 items) and Household activity (16 items)  *Measurement period* = previous 7 days | Partial correlations were calculated between PA and CRF, with adjustments for covariates.  *Covariates* = Age, TSI and medication use | - *AV̇O_2peak_*: Non-significant (P>0.05) correlation with total activity score (*r*=0.45). - ***RV̇O_2peak_*: Significant (P<0.01) correlation with total activity score (*r*=0.64).** |
| Martin-Ginis et al. (2021)  Canada | *N* = 39 (29M/10F)  *Age* = 42 ± 10 yrs  *TSI* = 13 ± 11 yrs  *Classification* = 15 T/24 P  *Severity* = 39 comp./ 0 incomp.  *AV̇O_2peak_^a^* = 1.10 ± 0.43 L/min  *RV̇O_2peak_* = 13.9 ± 5.5 ml/kg/min  *PPO* = 60 ± 28  *CPET* = Incremental exercise protocol on an electrically braked ACE | *Method* = LTPAQ-SCI  *PA dimensions* = Mild LTPA (*‘very light work’*), Moderate LTPA (*‘some physical effort’*), Heavy LTPA (*‘a lot of physical effort’*) and Total LTPA (all dimensions combined) (all measured as min/wk)  *Measurement period* = previous 7 days | One-tailed Pearson correlation coefficients were calculated between LTPA dimensions and CRF.  *Covariates* = NR  Due to deviation from the normal distribution, a square root transformation was carried out on the LTPAQ-SCI variables prior to analysis. | - ***RV̇O_2peak_*: Significant correlations with Moderate LTPA (*r*=0.28, P<0.05), Heavy LTPA (*r*=0.44, P<0.01) and Total LTPA (*r*=0.33, P<0.05).** - ***PPO*: Significant (P<0.05) correlation only with Heavy LTPA (*r*=0.29).** - No significant associations between CRF and Mild LTPA. |
| Nightingale et al. (2017)  UK | *N* = 33 (27M/6F)  *Age* = 44 ± 9 yrs  *TSI* = 15 ± 10 yrs  *Classification* = 0 T/33 P  *Severity* = 29 comp./ 4 incomp.  *AV̇O_2peak_* = 1.51 ± 0.50 L/min  *RV̇O_2peak_* = 19.8 ± 6.4 ml/kg/min  *PPO* = NR  *CPET* = Incremental exercise protocol on an electrically braked ACE | *Method* = individually calibrated, chest-mounted multi-sensor PA monitor (ActiheartTM, Cambridge Neurotechnology Ltd)  *PA dimensions* = activity energy expenditure (kcal/day), PA level (PAL: total energy expenditure/resting metabolic rate) and based on metabolic equivalents (METs) time spent performing (minutes/day); sedentary activities (< 1.5 METs), light-intensity activity (1.5–2.9 METs) and moderate-to-vigorous intensity physical activity (MVPA; ≥ 3 METs)  *Measurement period* = 7 days | Part correlations were calculated between dimensions of PA and CRF, with adjustments for significant (P < 0.05) covariates, using multiple linear regressions.  *Covariates* = Age and level of spinal cord lesion (both continuous variables), neurological completeness of injury and sex (both categorical variables)  AEE, PAL and MVPA were positively skewed and log-transformed prior to analysis. | - ***AV̇O_2peak_*: Significant (P<0.01) correlations with AEE (*r*=0.43), PAL (*r*=0.39) and MVPA (*r*=0.48).** - ***RV̇O_2peak_*: Significant (P<0.05) correlations with AEE (*r*=0.28), PAL (*r*=0.35), sedentary time (*r*=-0.26) and MVPA (*r*=0.32).** - No significant associations between CRF and light-intensity activity. - When dichotomised into three groups: LOW (<40 mins/wk), MOD (40 – 149 mins/wk) and HIGH >150 mins/wk) there was a significant effect of MVPA volume on both AV̇O2_peak_ and RV̇O_2peak_. *Post Hoc* analyses revealed that V̇O_2peak_ was significantly higher in the HIGH compared to the LOW group (P < 0.003). |

Continuous variables are displayed as mean ± standard deviation. Categorical variables are displayed as n. Cardiorespiratory fitness (CRF) outcomes in bold indicate a significant association (*p*<0.05). ACE, arm crank ergometry; AEE, activity energy expenditure; AV̇O_2peak_, absolute peak oxygen consumption; Comp., motor complete injury; F, females; Incomp., motor incomplete injury; LTPA, leisure time physical activity; LTPAQ-SCI, leisure time physical activity questionnaire for people with spinal cord injury; M, males; METS, metabolic equivalents; MVPA, moderate-to-vigorous physical activity; NR, not reported; P, paraplegia; PA, physical activity; PADS, physical activity and disability survey; PAL, physical activity levels; PARA-SCI, physical activity recall assessment for people with spinal cord injury; PPO, peak power output; RV̇O_2peak_, relative peak oxygen consumption; T, tetraplegia; TSI, time since injury. ^a^ Calculated using body mass (kg) provided in the paper. ^b^ Calculated by combining subgroups RV̇O_2peak_ provided in the paper

| **Table 3.** Risk of bias of cross-sectional association studies assessed via the NIH Quality Assessment Tool for Observational Cohort and Cross-Sectional Studies (14 items). | | | | | | | | | | | | | | | |
| --- | --- | --- | --- | --- | --- | --- | --- | --- | --- | --- | --- | --- | --- | --- | --- |
| **Author (Year)** | **Q1** | **Q2** | **Q3** | **Q4** | **Q5** | **Q6** | **Q7** | **Q8** | **Q9** | **Q10** | **Q11** | **Q12** | **Q13** | **Q14** | **Overall Quality** |
| Hoevenaars et al. (2023) | **✓** | **✓** | **?** | **✓** | ❌ | ❌ | ❌ | **✓** | **✓** | ❌ | **✓** | **?** | **N/A** | **✓** | **GOOD** |
| Janssen et al. (2002) | **✓** | **✓** | **?** | **?** | ❌ | ❌ | ❌ | **✓** | **✓** | **?** | **✓** | ❌ | ❌ | **✓** | **POOR** |
| Lannem et al. (2010) | **✓** | ❌ | **✓** | ❌ | ❌ | ❌ | ❌ | **✓** | ❌ | ❌ | **✓** | **?** | **N/A** | ❌ | **POOR** |
| Latimer et al. (2006) | **✓** | ❌ | **?** | **?** | ❌ | ❌ | ❌ | **✓** | **✓** | ❌ | **✓** | **?** | **N/A** | **✓** | **FAIR** |
| Manns et al. (2005) | **✓** | ❌ | **?** | **?** | ❌ | ❌ | ❌ | ❌ | **✓** | ❌ | **✓** | **?** | **N/A** | **✓** | **FAIR** |
| Martin-Ginis et al. (2021) | **✓** | **✓** | **?** | **?** | ❌ | ❌ | ❌ | **✓** | **✓** | ❌ | **✓** | **?** | **N/A** | **✓** | **FAIR** |
| Nightingale et al. (2017) | **✓** | **✓** | **?** | **?** | **✓** | ❌ | ❌ | **✓** | **✓** | ❌ | **✓** | ❌ | **N/A** | **✓** | **GOOD** |
| **Total ✓** | **7** | **4** | **1** | **0** | **1** | **0** | **0** | **6** | **6** | **0** | **7** | **0** | **0** | **6** |  |
| **Total ❌** | **0** | **3** | **0** | **1** | **6** | **7** | **7** | **1** | **1** | **6** | **0** | **2** | **1** | **1** |  |
| **Total ?** | **0** | **0** | **6** | **5** | **0** | **0** | **0** | **0** | **0** | **1** | **0** | **5** | **0** | **0** |  |
| **✓** = yes, ❌ = no, **?** = cannot determine/not reported | | | | | | | | | | | | | | | |

**Quality Assessment Questions**

1. Was the research question or objective in this paper clearly stated?
2. Was the study population clearly specified and defined?
3. Was the participation rate of eligible persons at least 50%?
4. Were all the subjects selected or recruited from the same or similar populations (including the same time period)? Were inclusion and exclusion criteria for being in the study prespecified and applied uniformly to all participants?
5. Was a sample size justification, power description, or variance and effect estimates provided?
6. For the analyses in this paper, were the exposure(s) of interest measured prior to the outcome(s) being measured?
7. Was the timeframe sufficient so that one could reasonably expect to see an association between exposure and outcome if it existed?
8. For exposures that can vary in amount or level, did the study examine different levels of the exposure as related to the outcome (e.g., categories of exposure, or exposure measured as continuous variable)?
9. Were the exposure measures (independent variables) clearly defined, valid, reliable, and implemented consistently across all study participants?
10. Was the exposure(s) assessed more than once over time?
11. Were the outcome measures (dependent variables) clearly defined, valid, reliable, and implemented consistently across all study participants?
12. Were the outcome assessors blinded to the exposure status of participants?
13. Was loss to follow-up after baseline 20% or less?
14. Were key potential confounding variables measured and adjusted statistically for their impact on the relationship between exposure(s) and outcome(s)?

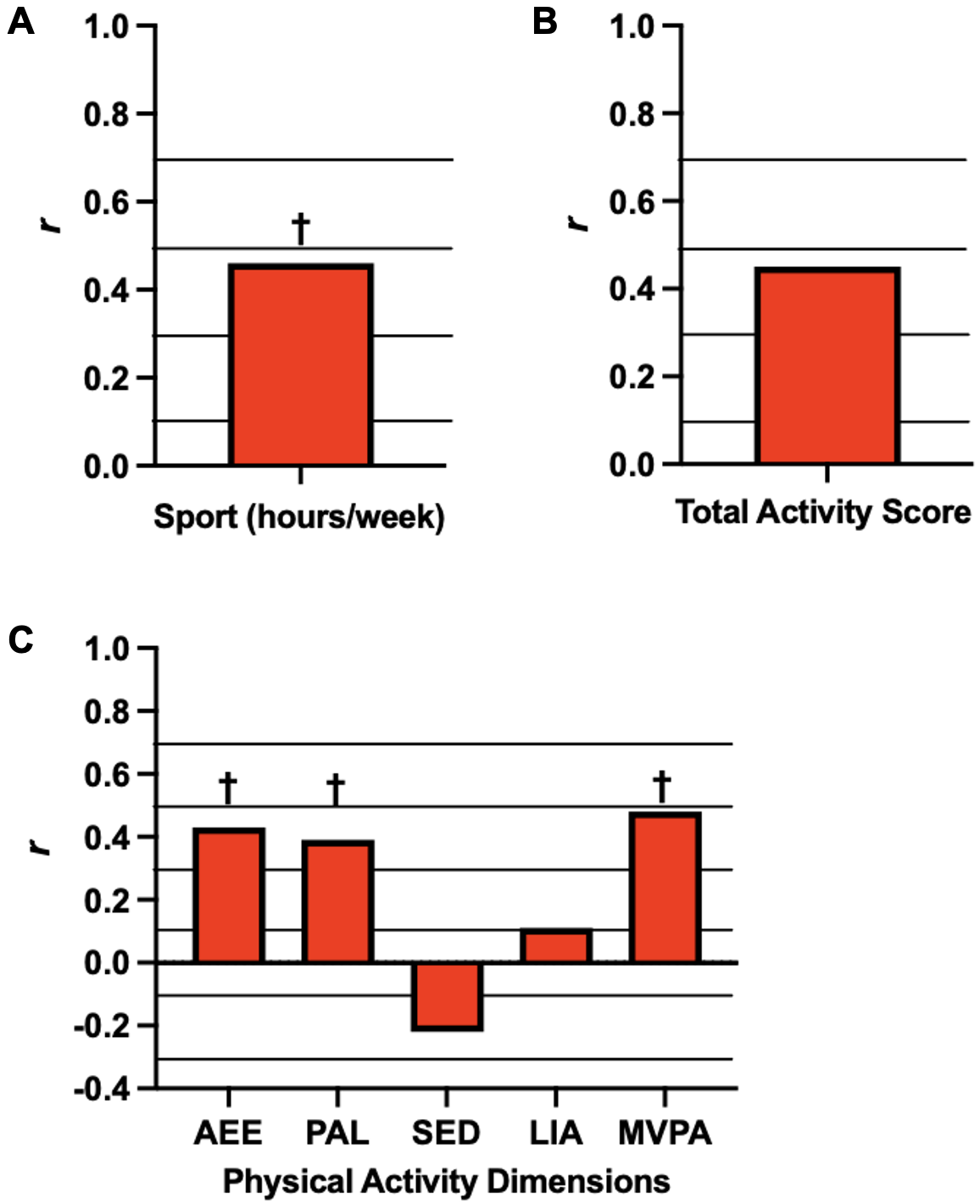

**Figure 1.** Graphical representations of the associations between absolute peak oxygen consumption (AV̇O_2peak_*,* red bars) and habitual physical activity levels. Methods assessing physical activity were self-reported (A and B) and via wearable devices (C). Data has been adapted into figures from A (Janssen *et al*., 2002), B (Manns *et al*., 2005), and C (Nightingale *et al*., 2017). Solid black lines are overlaid to help interpret the magnitude of each correlation: small (*r*>0.1), moderate (*r*>0.3), large (*r*>0.5), and very large (*r*>0.7). Significant associations are denoted by * *p*<0.05, † *p*<0.01. *r* indicates Pearson correlation coefficients. AEE, activity energy expenditure (kcal/day); LIA, light-intensity activity; MVPA, moderate-to-vigorous physical activity; PAL, physical activity levels; SED, sedentary time.

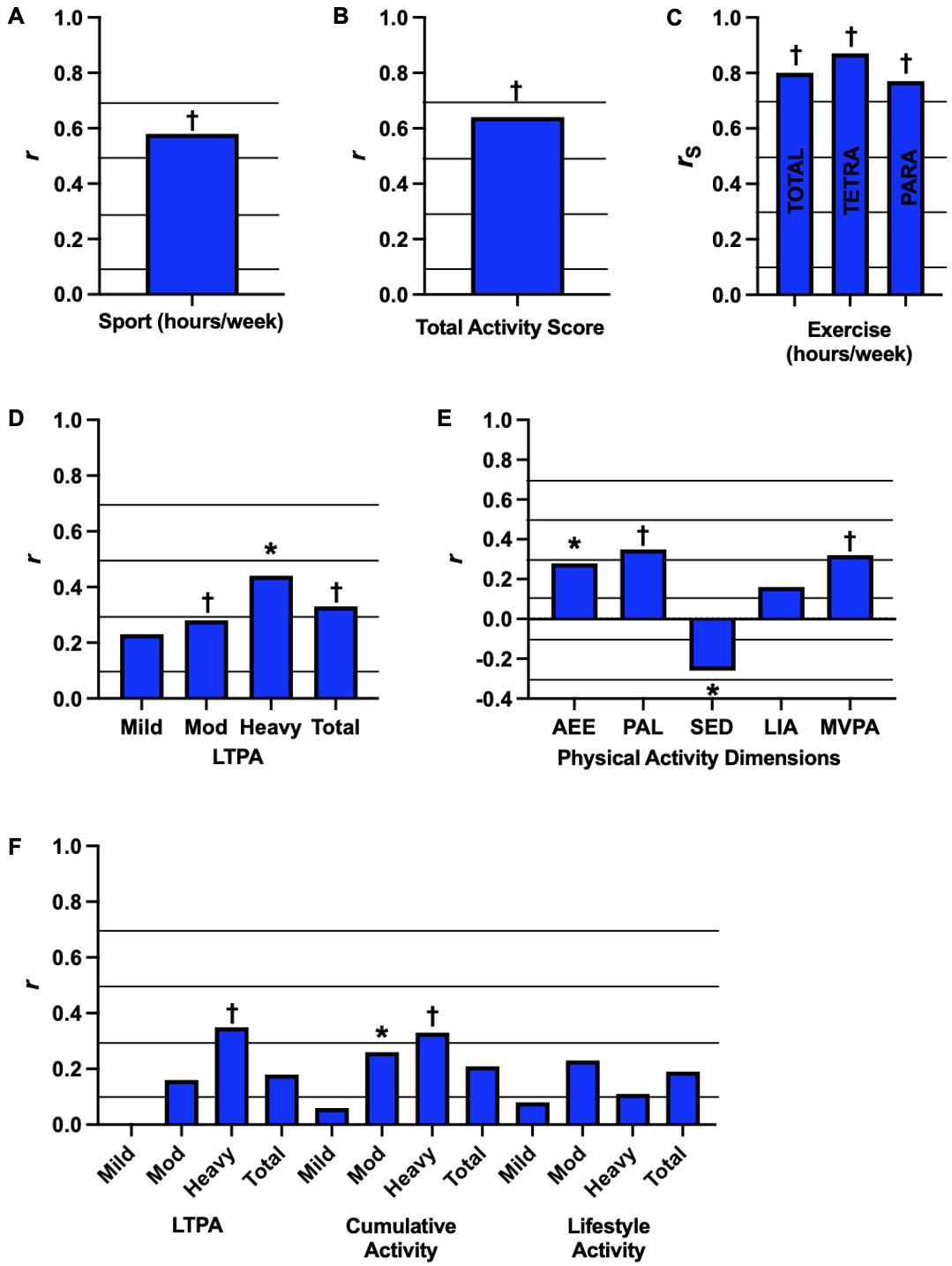
**Figure 2.** Graphical representations of the associations between relative peak oxygen consumption (RV̇O_2peak_, blue bars) and habitual physical activity levels. Methods assessing physical activity were self-reported (A,B,C,D,F) and via wearable devices (E). Data has been adapted into figures from A (Janssen *et al*., 2002), B (Manns *et al*., 2005), C (Lannem *et al*., 2010), D (Martin-Ginis *et al*., 2021), E (Nightingale *et al*., 2017), and F (Latimer *et al*., 2006). Solid black lines are overlaid to help interpret the magnitude of each correlation: small (*r*>0.1), moderate (*r*>0.3), large (*r*>0.5), and very large (*r*>0.7). Significant associations are denoted by * *p*<0.05, † *p*<0.01. *r* and *rho* (*rS*) indicate Pearson and Spearman correlation coefficients, respectively. AEE, activity energy expenditure (kcal/day); LIA, light-intensity activity; LTPA, leisure time physical activity; MVPA, moderate-to-vigorous physical activity; PAL, physical activity levels; SED, sedentary time.

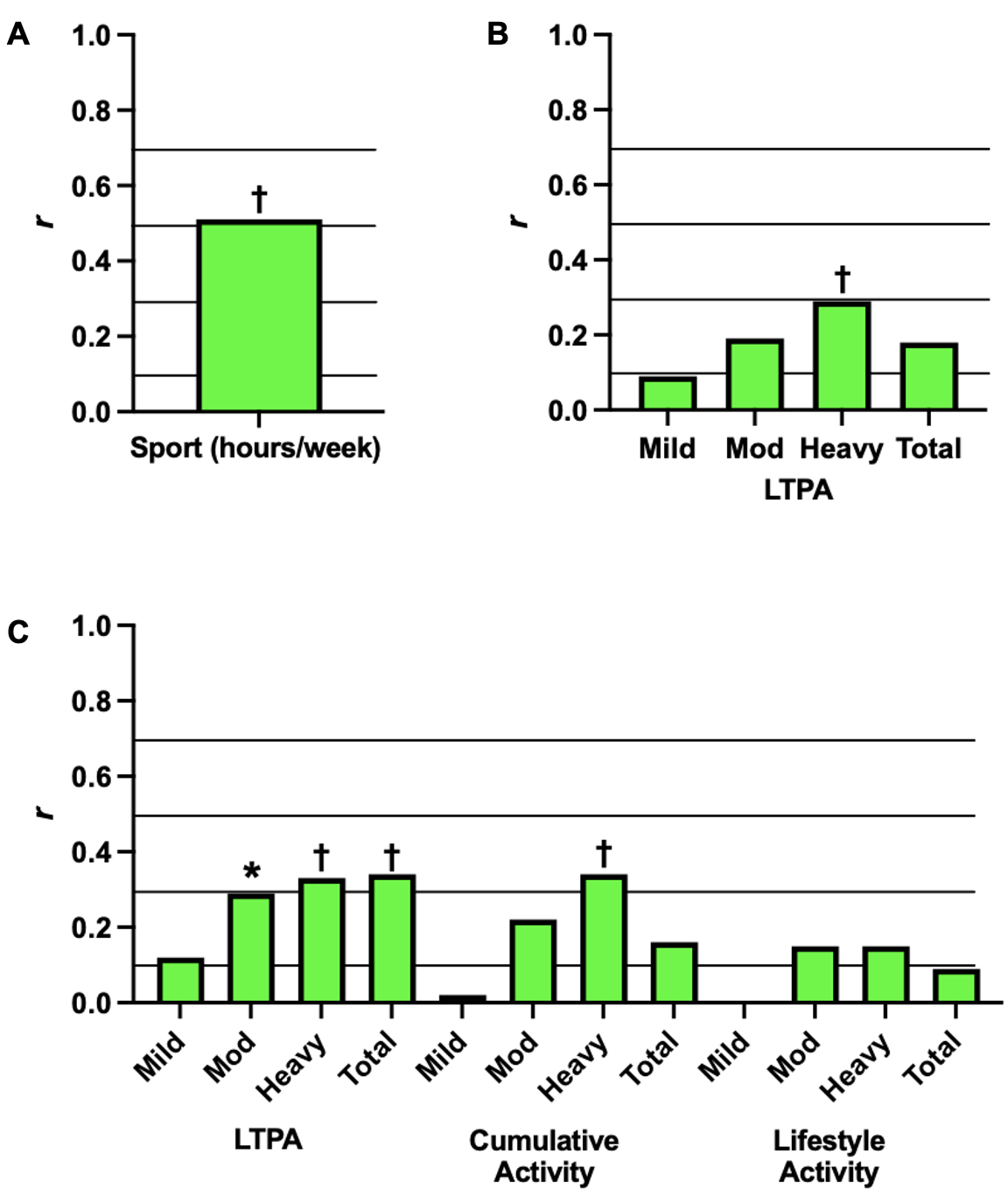

**Figure 3.** Graphical representations of the associations between peak power output (PPO, green bars) and habitual physical activity levels. Methods assessing physical activity were self-reported. Data has been adapted into figures from A (Janssen *et al*., 2002), B (Martin-Ginis *et al*., 2021), and C (Latimer *et al*., 2006). Solid black lines are overlaid to help interpret the magnitude of each correlation: small (*r*>0.1), moderate (*r*>0.3), large (*r*>0.5), and very large (*r*>0.7). Significant associations are denoted by * *p*<0.05, † *p*<0.01. *r* indicates Pearson correlation coefficients. LTPA, leisure time physical activity.

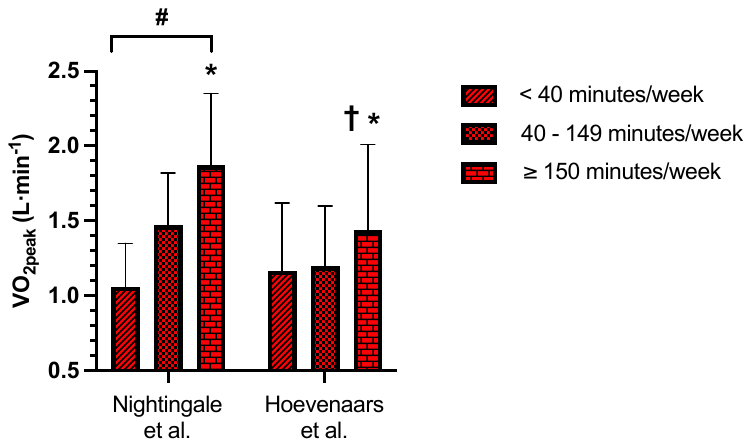

**A**

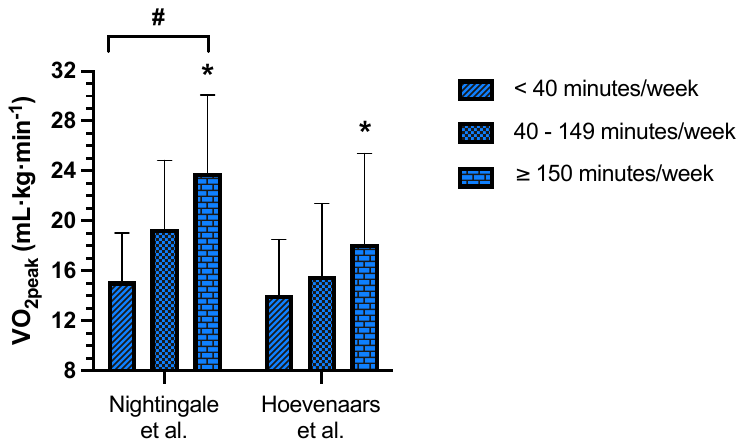

**B**

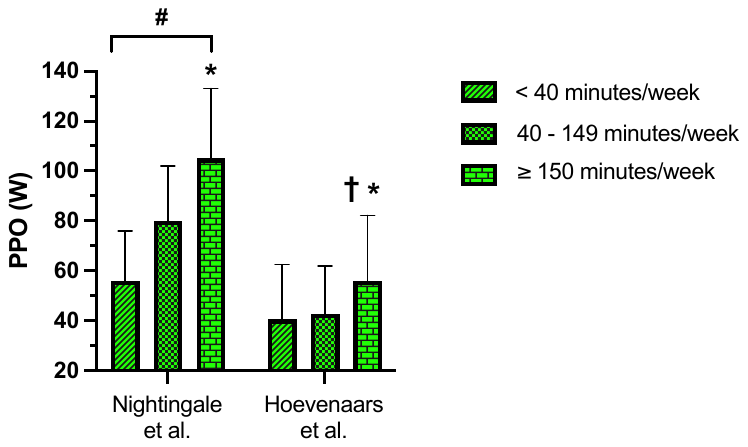

**C**

**Figure 4.** Graphical representations of absolute (A) and relative (B) peak oxygen uptake (VO_2peak_) and peak power output (C; PPO) between individuals with spinal cord injury performing exercise volumes of < 40 minutes/week, 40 – 149 minutes/week and ≥150 minutes/week. Data are presented from Nightingale et al. (2018) and Hoevenaars et al. (2023). ^#^ Significant difference across groups. * Significant difference between < 40 minutes/week and ≥150 minutes/week. ^†^ Significant difference between 40 – 149 minutes/week and ≥150 minutes/week.

**References**

Cross-sectional associations studies included in the systematic review, sorted alphabetically:

Hoevenaars D, Holla JFM, Postma K, van der Woude LHV, Janssen TWJ, de Groot S. Associations between meeting exercise guidelines, physical fitness, and health in people with spinal cord injury. Disabil Rehabil 2023;45(6):1030-1037.

Janssen TWJ, Dallmeijer AJ, Veeger DJHEJ, van der Woude LHV. Normative values and determinants of physical capacity in individuals with spinal cord injury. J Rehabil Res Dev 2002;39(1):29-39.

Lannem AM, Sorensen M, Lidal IB, Hjeltnes N. Perceptions of exercise mastery in persons with complete and incomplete spinal cord injury. Spinal Cord 2010;48:388-92.

Latimer AE, Martin-Ginis KA, Craven CB, Hicks AL. The Physical Activity Recall Assessment for People with Spinal Cord Injury: Validity. Med Sci Sports Exerc 2006;38(2):208-16.

Manns PJ, McCubbin JA, Williams DP. Fitness, Inflammation, and the Metabolic Syndrome in Men With Paraplegia. Arch Phys Med Rehabil 2005;86(6):1176-81.

Martin-Ginis KA, Ubeda-Colomer J, Alrashidi AA, Nightingale TE, Au JS, Currie KD, et al. Construct validation of the leisure time physical activity questionnaire for people with SCI (LTPAQ-SCI). Spinal Cord 2021;59:311-8.

Nightingale TE, Walhin J-P, Thompson D, Bilzon JL. Biomarkers of cardiometabolic health are associated with body composition characteristics but not physical activity in persons with spinal cord injury. J Spinal Cord Med 2017;42(3):328-37.
