## Supplementary material for "The effect of exercise on aerobic capacity in individuals with spinal cord injury: A systematic review with meta-analysis and meta-regression": S13 Observational studies

**Electronic Supplementary Material 13**

**PLoS Medicine**

Hodgkiss, D.D^1^, Bhangu, G^2,3^, Lunny, C^4^, Jutzeler C.R^5,6^, Chiou S.Y^1,7,8,9^ Walter, M^2,10^, Lucas S.E^1,7^, Krassioukov, A.V.^2,11,12^, Nightingale, T.E.^1,2,9^

**^1^** School of Sport, Exercise and Rehabilitation Sciences, University of Birmingham, UK.

**^2^** International Collaboration on Repair Discoveries (ICORD), University of British Columbia, Vancouver, British Columbia, Canada. **^3^** MD Undergraduate Program, Faculty of Medicine, University of British Columbia, Vancouver, Canada. **^4^** Knowledge Translation Program, Li Ka Shing Knowledge Institute, St. Michael’s Hospital, Toronto, and the University of British Columbia, Vancouver, British Columbia, Canada.

**^5^** Department of Health Sciences and Technology, ETH Zurich, Zurich, Switzerland. **^6^** Schulthess Clinic, Zurich, Switzerland. **^7^** Centre for Human Brain Health, University of Birmingham, United Kingdom. **^8^** MRC Versus Arthritis Centre for Musculoskeletal Ageing Research, University of Birmingham, United Kingdom.

**^9^** Centre for Trauma Science Research, University of Birmingham, United Kingdom. **^10^** Department of Urology, University Hospital Basel, University of Basel, Basel, Switzerland. **^11^** Department of Medicine, Division of Physical Medicine and Rehabilitation, University of British Columbia, Vancouver, British Columbia, Canada. **^12^** GF Strong Rehabilitation Centre, Vancouver Coastal Health, Vancouver, British Columbia, Canada.

***Purpose:*** This supplementary file includes the pooled and individual participant demographics, injury characteristics and cardiorespiratory fitness (CRF) outcome data from observational studies assessing changes in CRF. The purpose of this secondary meta-analysis was to investigate whether there are significant improvements in CRF following inpatient rehabilitation [standard of care] and community free-living follow-up.

***Conclusion:*** Absolute and relative peak oxygen consumption are significantly improved following standard of care inpatient rehabilitation but not free-living follow-up. Peak power output (PPO) improved significantly following both standard of care inpatient rehabilitation and free-living follow-up. More research is required to investigate the effectiveness of different exercise interventions above and beyond traditional standard of care inpatient rehabilitation during the sub-acute phase of SCI for improving CRF. Most of the observational data is from studies conducted in the Netherlands, therefore limited information exists on the impact of standard of care rehabilitation from other countries. Nevertheless, even without specific, targeted exercise individuals with SCI improve CRF and PPO after injury through general rehabilitation. Further improvements in PPO are noted after a period of free-living follow-up. Collectively, these data imply a natural improvement in aerobic capacity post-injury. Therefore, to determine effects of exercise training in the sub-acute setting comparisons must be made to existing standard of care rehabilitation in the form of randomised controlled trials.

| **Table 1:** Overview of participant demographics and injury characteristics from included observational studies investigating the impact of rehabilitation or long-term follow-up on cardiorespiratory fitness outcomes. | |
| --- | --- |
| **Absolute V̇O_2peak_ studies (N = 5)** | |
| Σ participants = 343 | |
| *Participant demographics* | |
| Age (years) | 37 (30 – 40) |
| Sex (M/F), n (%) | 274/69 (80%/20%) |
| *Injury characteristics* |  |
| Neurological level of injury (TETRA/PARA), n (%) | 73/168 (21%/49%), Σ^NR^=102 (30%) |
| Severity (complete/incomplete), n (%) | 160/81 (46.5%/23.5%), Σ^NR^=102 (30%) |
| Time since injury: acute (days) and chronic (years) | Acute: 100 (73 – 331)  Chronic: 15 *N=1 |
| *CRF outcome* | |
| Absolute V̇O_2peak_ (L/min) | 0.99 (0.63 – 1.67) |
| **Relative V̇O_2peak_ studies (N = 4)** | |
| Σ participants = 170 | |
| *Participant demographics* | |
| Age (years) | 32 (25 – 40) |
| Sex (M/F), n (%) | 147/23 (86%/14%)) |
| *Injury characteristics* | |
| Neurological level of injury (TETRA/PARA), n (%) | 28/40 (16.5%/23.5%), Σ^NR^=102 (60%) |
| Severity (complete/incomplete), n (%) | 49/19 (29%/11%), Σ^NR^=102 (60%) |
| Time since injury: acute (days) and chronic (years) | Acute: 119 (77 – 331)  Chronic: 15 ^a^ |
| *CRF outcome* | |
| Relative V̇O_2peak_ (mL/kg/min) | 12.9 (8.9 – 21.6) |
| **Peak power output studies (N = 5)** | |
| Σ participants = 343 | |
| *Participant demographics* | |
| Age (years) | 37 (30 – 40) |
| Sex (M/F), n (%) | 274/69 (80%/20%) |
| *Injury characteristics* | |
| Neurological level of injury (TETRA/PARA), n (%) | 73/168 (21%/49%), Σ^NR^=102 (30%) |
| Severity (complete/incomplete), n (%) | 160/81 (46.5%/23.5%), Σ^NR^=102 (30%) |
| Time since injury: acute (days) and chronic (years) | Acute: 100 (73 – 331)  Chronic: 15 *N=1 |
| *CRF outcome* | |
| Peak power output (W) | 37 (31 – 65) |
| Continuous variables are displayed as weighted means (range: lowest – highest mean values reported from studies). Categorical variables are displayed as n (%).Weighted means were calculated to account for differences in sample size between studies using the following formula: Σn*x̅ /Σn, where Σ = the sum of, n = number of participants in each study and, x̅ = mean CRF outcome of each study. a Data taken from Janssen et al. (2002); no TSI reported in Szymczak et al. (2022). | |

**Table 2:** Summaries of individual observational studies.

| **ACUTE SCI** | | | |
| --- | --- | --- | --- |
| **Author/year/ country** | **Population** | **Rehabilitation/Exercise** | **Cardiorespiratory fitness outcomes** |
| Dallmeijer et al. (1999)  The Netherlands | *N =* 20 (16M/4F)  *Age* = 40 ± 15 yrs  *TSI* = 331 ± 142 days  *Classification* = 9 T/11 P  *Severity* = 15 comp./ 5 incomp.  *CPET* = Wheelchair ergometer exercise test | *Phase of rehabilitation =* Observation after discharge from rehabilitation programme  *Length of follow-up =* 1.2 ± 0.3 years  *Rehabilitation information =* Mean sport activity was 2.5 ± 2.0 hours/week, with N=8 reporting no sport activity. | *AV̇O_2peak_* *(N=18)   - Discharge: 1.10 ± 0.48 L/min - 1-year post discharge: 1.14 ± 0.59 L/min   *RV̇O_2peak_* *(N=18)   - Discharge: 14.8 ± 6.7 mL/kg/min - 1-year post discharge: 14.3 ± 7.8 mL/kg/min   *PPO* *(N=19)   - **Discharge: 39 ± 27 W** - **1-year post discharge: 44 ± 31 W** |
| Haisma et al. (2008)  The Netherlands | *N =* 176 (133M/43F)  *Age* = 40 ± 14 yrs  *TSI* = 88 ± 61 days  *Classification* = 55 T/121 P  *Severity* = 117 comp./ 59 incomp.  *CPET* = Graded maximal treadmill exercise test | *Phase of rehabilitation =* Inpatient rehabilitation  *Length of follow-up =*  Pre: 88 ± 61 days  Discharge: 290 ± 140 days  *Rehabilitation information =* NR | *AV̇O_2peak_*   - Baseline: 1.03 ± 0.36 L/min - 3-months: 1.15 ± 0.42) L/min *(N=124) - Discharge: 1.22 ± 0.44 L/min *(N=160) - 1-year post discharge: 1.32 ± 0.51 L/min *(N=133)   *RV̇O_2peak_*   - NR   *PPO*   - Baseline: 31 ± 18 W - 3-months: 37 ± 21 W *(N=124) - Discharge: 41 ± 23 W *(N=160) - 1 year-post discharge: 48 ± 25 W *(N=133) |
| Leving et al. (2021)  The Netherlands | *N =* 8 (5M/3F)  *Age* = 40 ± 17 yrs  *TSI* = 0.2 ± 0.05 yrs  *Classification* = 1 T/7 P  *Severity* = 5 comp./ 3 incomp.  *CPET* = Graded wheelchair treadmill exercise test | *Phase of rehabilitation =* Active inpatient rehabilitation  *Length of follow-up =* 5weeks (N=2 assessed 1-2 weeks following discharge)  *Rehabilitation information =* NR | *AV̇O_2peak_* *(N=5) ^b^   - Baseline: 1.20 (0.71) L/min - Discharge: 1.20 (1.05) L/min   *RV̇O_2peak_*   - NR   *PPO* *(N=6) ^b^   - **Baseline: 40 (51) W** - **Discharge: 48 (56) W**   **Note: Unable to report in this secondary meta-analysis due to data presented as median (IQR).* |
| Stewart et al. (2000)  USA | *N =* 102 (83M/19F)  *Age* = 30 ± 10 yrs  *TSI* = 77 ± 108 days  *Classification* = NR  *Severity* = 91% motor complete (AIS A), 9% sensory incomplete (AIS B,C,D)  *CPET* = Wheelchair ergometer and arm crank ergometer exercise tests | *Phase of rehabilitation =* Inpatient rehabilitation  *Length of follow-up =* 96 days for individuals with tetraplegia and 76 days for other individuals  *Rehabilitation information =* Participants were randomly assigned to wheelchair ergometry or arm crank ergometry tests at admission and were tested on the same ergometer at discharge. No N is provided for each test. | *AV̇O_2peak_*   - **Baseline: 0.63 ± 0.28 L/min** - **Discharge: 0.78 ± 0.32 L/min**   *RV̇O_2peak_*   - **Baseline: 8.9 ± 3.9 mL/kg/min** - **Discharge: 11.0 ± 4.4 mL/kg/min**   *PPO*   - **Baseline: 36 ± 17 W** - **Discharge: 44 ± 20 W** |
| **CHRONIC SCI** | | | |
| Janssen et al. (1996)  The Netherlands | *N =* 37 (37M/0F)  *Age* = 21 – 41 yrs  *TSI* = NR  *Classification* = 11 T/0 P  *Severity* = 11 comp./ 0 incomp.  *CPET* = Graded wheelchair exercise test | *Phase of rehabilitation =* Long-term observation  *Length of follow-up =* 35 ± 2 months  *Rehabilitation information =* NR | *AV̇O_2peak_*   - Baseline: 1.67 ± 0.47 L/min - Follow-up: 1.75 ± 0.55 L/min   *RV̇O_2peak_*   - Baseline: 21.6 ± 7.4 mL/kg/min - Follow-up: 21.7 ± 7.9 mL/kg/min   *PPO*   - **Baseline: 65 ± 26 W** - **Follow-up: 72 ± 27 W** |
| Szymczak et al. (2022)  Poland | *N =* 11 (11M/0F)  *Age* = 37 ± 12 yrs  *TSI* = 15 ± 9 yrs  *Classification* = 8 T/29 P  *Severity* = 23 comp./ 14 incomp.  *CPET* = Graded arm-crank ergometry exercise test | *Phase of rehabilitation =* Observation of changes in CRF of national-level wheelchair rugby players following a training programme in the lead-in to an international tournament.  *Length of follow-up =* 30 weeks  *Programme information =* Training was split into microcycles to build aerobic capacity, strength and speed endurance, and speed and tactics in the lead-in to the tournament. Players performed a range of activities (e.g., wheelchair push, gym/fitness, wheelchair rugby half court play etc.), at a range of exercise intensities (50 – 90%max), session frequencies (1 – 3 times per week) and durations (15 – 150 mins). | *AV̇O_2peak_*   - NR   *RV̇O_2peak_*   - Baseline: 16.8 ± 3.5 mL/kg/min - Follow-up: 15.9 ± 3.5 mL/kg/min   *PPO*   - NR |

Continuous variables are displayed as mean ± standard deviation. Categorical variables are displayed as n. Cardiorespiratory fitness (CRF) outcomes in bold indicate a significant difference between time points (*p* < 0.05). AV̇O_2peak_, absolute peak oxygen consumption; CD, cannot determine; Comp., motor complete injury; CPET, cardiopulmonary exercise test; F, females; Incomp., motor incomplete injury; M, males; NR, not reported; P, paraplegia; PPO, peak power output; RV̇O_2peak_, relative peak oxygen consumption; T, tetraplegia; TSI, time since injury. ^a^ Calculated using body mass (kg) provided in the paper (therefore no significant differences reported). ^b^ Presented as median (range).

| **Table 3.** Risk of bias of observational studies assessed via the NIH Quality Assessment Tool for Observational Cohort and Cross-Sectional Studies (14 items). | | | | | | | | | | | | | | | |
| --- | --- | --- | --- | --- | --- | --- | --- | --- | --- | --- | --- | --- | --- | --- | --- |
| **Author (Year)** | **Q1** | **Q2** | **Q3** | **Q4** | **Q5** | **Q6** | **Q7** | **Q8** | **Q9** | **Q10** | **Q11** | **Q12** | **Q13** | **Q14** | **Overall Quality** |
| Dallmeijer et al. (1999) | **✓** | **✓** | **?** | ❌ | **✓** | **✓** | **✓** | ❌ | ❌ | ❌ | **✓** | **?** | ❌ | ❌ | **POOR** |
| Haisma et al. (2008) | **✓** | **✓** | **?** | **✓** | ❌ | **✓** | **✓** | ❌ | **✓** | ❌ | **✓** | ❌ | ❌ | **✓** | **GOOD** |
| Janssen et al. (1996) | **✓** | **✓** | **?** | **✓** | ❌ | **✓** | **✓** | **✓** | **✓** | ❌ | **✓** | **?** | **✓** | **✓** | **GOOD** |
| Leving et al. (2019) | **✓** | **✓** | **?** | **✓** | ❌ | **✓** | **✓** | **?** | **?** | **✓** | **✓** | ❌ | **?** | ❌ | **POOR** |
| Stewart et al. (2000) | **✓** | **✓** | **?** | **✓** | **✓** | **✓** | ❌ | **✓** | **✓** | ❌ | **✓** | **?** | **✓** | **✓** | **GOOD** |
| Szymczak et al (2022) | **✓** | **✓** | **?** | **✓** | ❌ | **✓** | **✓** | ❌ | **✓** | ❌ | ❌ | ❌ | **✓** | ❌ | **FAIR** |
| **Total ✓** | **6** | **6** | **0** | **5** | **2** | **6** | **5** | **2** | **4** | **1** | **5** | **0** | **3** | **3** |  |
| **Total ❌** | **0** | **0** | **0** | **1** | **4** | **0** | **1** | **3** | **1** | **5** | **1** | **3** | **2** | **3** |  |
| **Total ?** | **0** | **0** | **5** | **0** | **0** | **0** | **0** | **1** | **1** | **0** | **0** | **3** | **1** | **0** |  |
| **✓** = yes, ❌ = no, **?** = cannot determine/not reported | | | | | | | | | | | | | | | |

**Quality Assessment Questions**

1. Was the research question or objective in this paper clearly stated?
2. Was the study population clearly specified and defined?
3. Was the participation rate of eligible persons at least 50%?
4. Were all the subjects selected or recruited from the same or similar populations (including the same time period)? Were inclusion and exclusion criteria for being in the study prespecified and applied uniformly to all participants?
5. Was a sample size justification, power description, or variance and effect estimates provided?
6. For the analyses in this paper, were the exposure(s) of interest measured prior to the outcome(s) being measured?
7. Was the timeframe sufficient so that one could reasonably expect to see an association between exposure and outcome if it existed?
8. For exposures that can vary in amount or level, did the study examine different levels of the exposure as related to the outcome (e.g., categories of exposure, or exposure measured as continuous variable)?
9. Were the exposure measures (independent variables) clearly defined, valid, reliable, and implemented consistently across all study participants?
10. Was the exposure(s) assessed more than once over time?
11. Were the outcome measures (dependent variables) clearly defined, valid, reliable, and implemented consistently across all study participants?
12. Were the outcome assessors blinded to the exposure status of participants?
13. Was loss to follow-up after baseline 20% or less?
14. Were key potential confounding variables measured and adjusted statistically for their impact on the relationship between exposure(s) and outcome(s)?

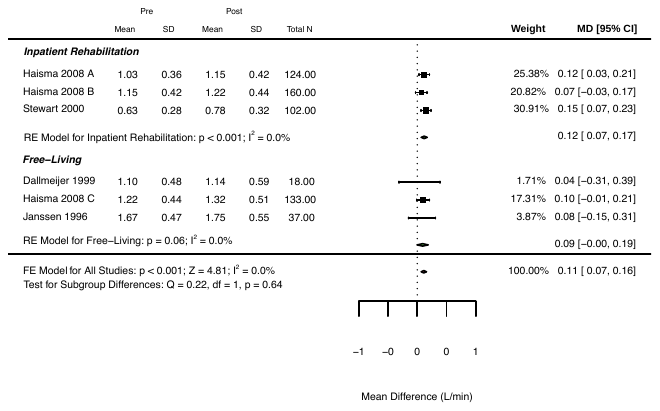

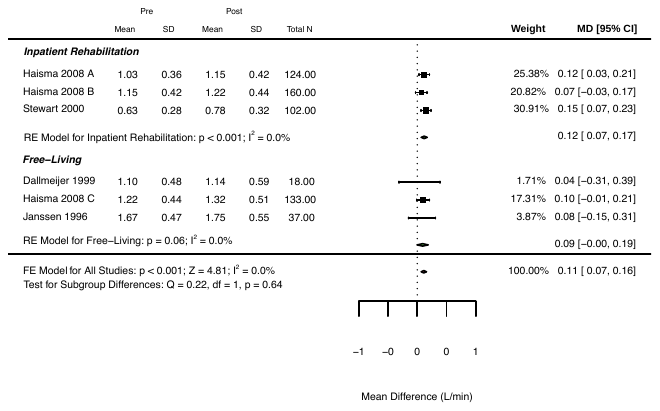

**Figure 1.** Changes in absolute peak oxygen consumption (L/min) following inpatient rehabilitation or a community free-living follow-up period. ^A^ Baseline to 3-months, ^B^ 3-months to discharge, ^C^ Discharge to follow-up. RE, random effects.

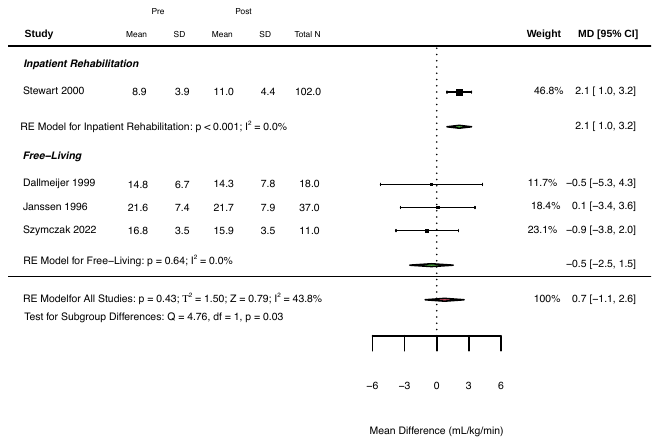

**Figure 2.** Changes in relative peak oxygen consumption (mL/kg/min) following inpatient rehabilitation or a community free-living follow-up period. RE, random effects.

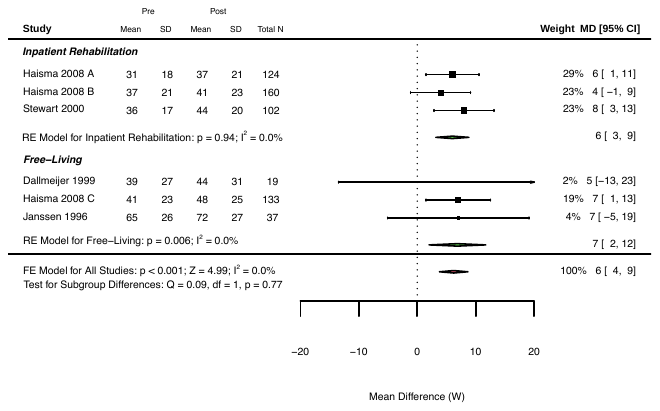

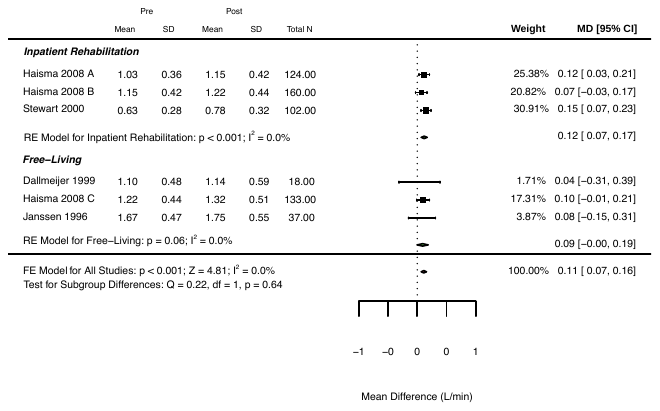

**Figure 3.** Changes in peak power output following inpatient rehabilitation or a community free-living follow-up period. RE, random effects.

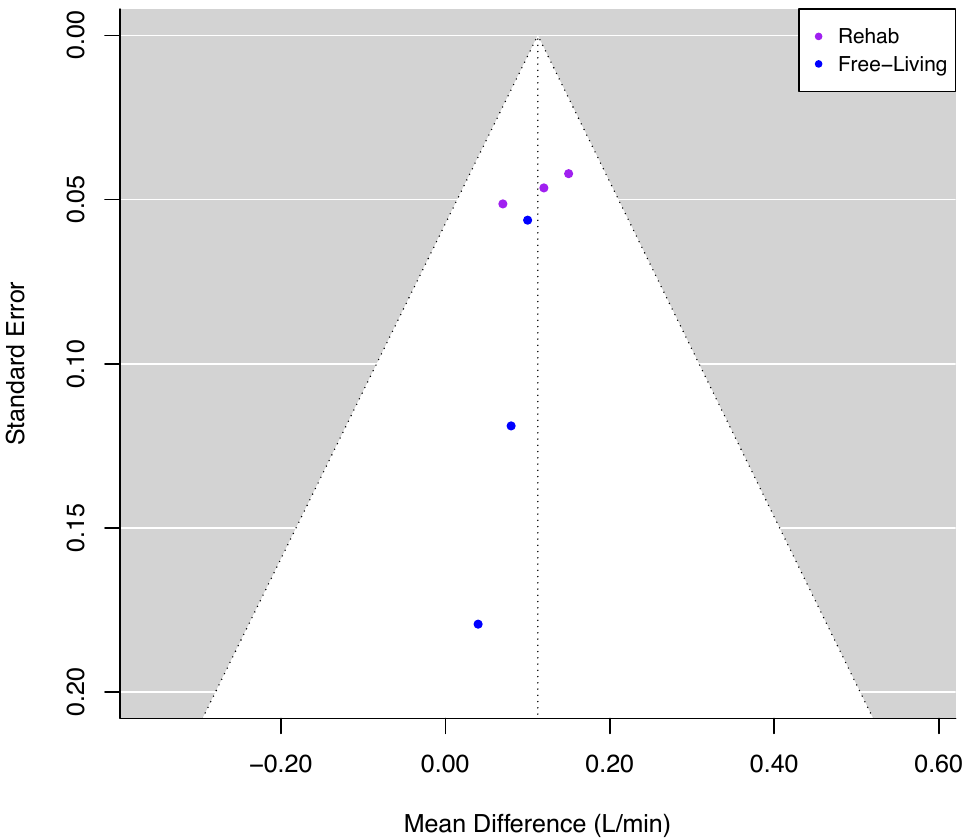

**Figure 4:** Funnel plot of absolute peak oxygen consumption with studies sub-grouped based on type of observational period [i.e., inpatient rehabilitation (rehab) or community free-living follow-up (free-living)]. Egger’s test not performed (<10 studies).

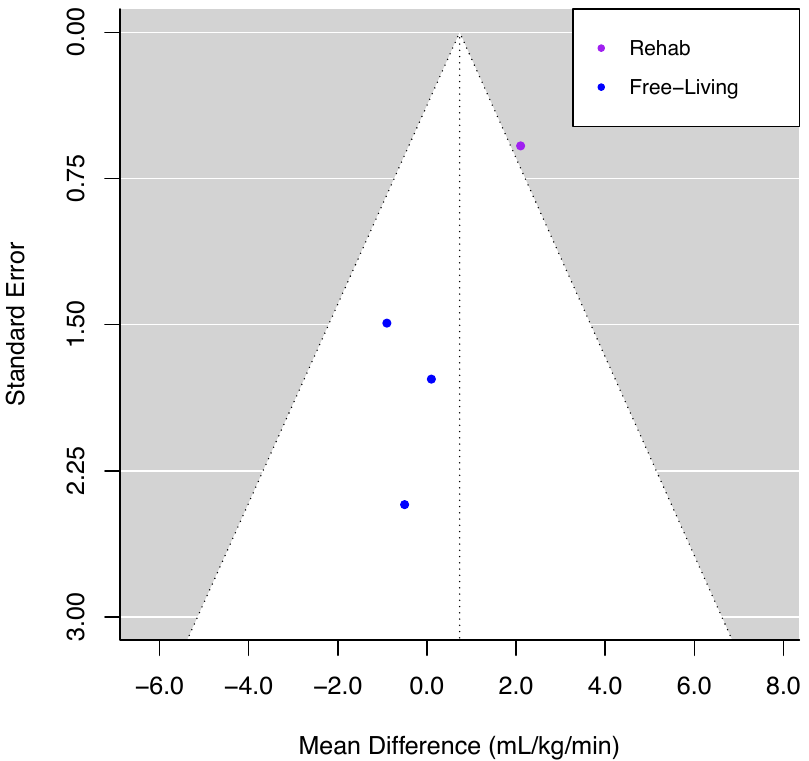

**Figure 5:** Funnel plot of relative peak oxygen consumption with studies sub-grouped based on type of observational period [i.e., inpatient rehabilitation (rehab) or community free-living follow-up (free-living)]. Egger’s test not performed (<10 studies).

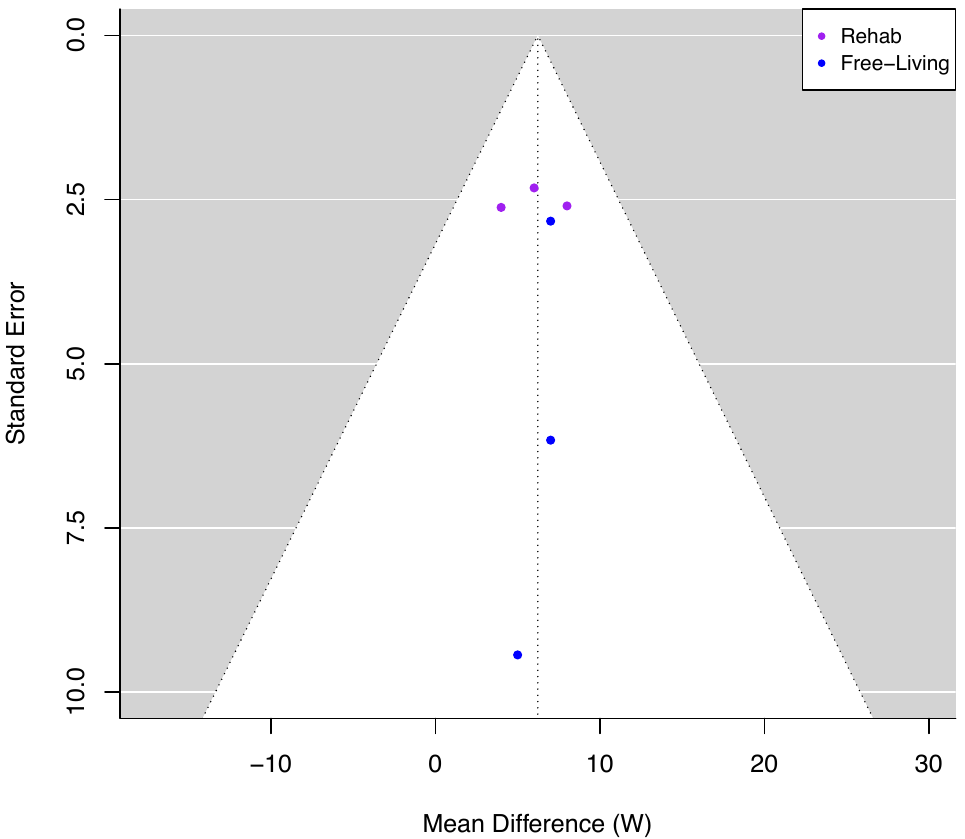

**Figure 6:** Funnel plot of peak power output with studies sub-grouped based on type of observational period [i.e., inpatient rehabilitation (rehab) or community free-living follow-up (free-living)]. Egger’s test not performed (<10 studies).

**References**

Observational studies included in the systematic review, sorted alphabetically:

Dallmeijer AJ vdWL, Hollander PA, Angenot EL. Physical performance in persons with spinal cord injuries after discharge from rehabilitation. Med Sci Sports Exerc 1999;31(8):1111-7.

Haisma JA, Post MW, van der Woude LH, Stam HJ, Bergen MP, Sluis TA, et al. Functional independence and health-related functional status following spinal cord injury: a prospective study of the association with physical capacity. J Rehabil Med 2008;40(10):812-8.

Janssen TW, van Oers CA, Rozendaal EP, Willemsen EM, Hollander AP, van der Woude LH. Changes in physical strain and physical capacity in men with spinal cord injuries. Med Sci Sports Exerc 1996;28(5):551-9.

Leving MT, de Groot S, Woldring FAB, Tepper M, Vegter RJK, van der Woude LHV. Motor learning outcomes of handrim wheelchair propulsion during active spinal cord injury rehabilitation in comparison with experienced wheelchair users. Disabil Rehabil 2021;43(10):1429-42.

Stewart MW, Melton-Rogers SL, Morrison S, Figoni SF. The measurement properties of fitness measures and health status for persons with spinal cord injuries. Arch Phys Med Rehabil 2000;81(4):394-400.
