## Supplementary material for "The effect of exercise on aerobic capacity in individuals with spinal cord injury: A systematic review with meta-analysis and meta-regression": S14 RCT exercise intensity comparisons

**Electronic Supplementary Material 14**

**PLoS Medicine**

Hodgkiss, D.D^1^, Bhangu, G^2,3^, Lunny, C^4^, Jutzeler C.R^5,6^, Chiou S.Y^1,7,8,9^ Walter, M^2,10^, Lucas S.E^1,7^, Krassioukov, A.V.^2,11,12^, Nightingale, T.E.^1,2,9^

**^1^** School of Sport, Exercise and Rehabilitation Sciences, University of Birmingham, UK.

**^2^** International Collaboration on Repair Discoveries (ICORD), University of British Columbia, Vancouver, British Columbia, Canada. **^3^** MD Undergraduate Program, Faculty of Medicine, University of British Columbia, Vancouver, Canada. **^4^** Knowledge Translation Program, Li Ka Shing Knowledge Institute, St. Michael’s Hospital, Toronto, and the University of British Columbia, Vancouver, British Columbia, Canada.

**^5^** Department of Health Sciences and Technology, ETH Zurich, Zurich, Switzerland. **^6^** Schulthess Clinic, Zurich, Switzerland. **^7^** Centre for Human Brain Health, University of Birmingham, United Kingdom. **^8^** MRC Versus Arthritis Centre for Musculoskeletal Ageing Research, University of Birmingham, United Kingdom.

**^9^** Centre for Trauma Science Research, University of Birmingham, United Kingdom. **^10^** Department of Urology, University Hospital Basel, University of Basel, Basel, Switzerland. **^11^** Department of Medicine, Division of Physical Medicine and Rehabilitation, University of British Columbia, Vancouver, British Columbia, Canada. **^12^** GF Strong Rehabilitation Centre, Vancouver Coastal Health, Vancouver, British Columbia, Canada.

***Purpose:*** This supplementary file includes the pooled and individual participant demographics, injury characteristics and exercise intervention parameters of randomised-controlled trials assessing the change in cardiorespiratory fitness following an exercise intervention of either low or moderate vs. vigorous or supramaximal exercise intensity. The purpose of this secondary meta-analysis was to investigate whether there are any additional CRF benefits when performing vigorous or supramaximal intensity exercise.

***Conclusion:*** Changes in CRF were not significantly different between low or moderate and vigorous or supramaximal intensity exercise interventions. This suggests that a similar magnitude of change in CRF following a low or moderate intensity exercise intervention *may possibly* be achieved with vigorous or supramaximal intensity exercise. At present there are too few studies in individuals with SCI comparing either time-matched or energy-matched vigorous-intensity exercise interventions to provide meaningful conclusions regarding whether more vigorous-intensity exercise can elicit superior improvements or achieve the same benefits in a more time efficient manner, respectively.

**Table 1:** Characteristics of randomised controlled trials (low or moderate vs. vigorous or supramaximal intensity interventions) reporting on each cardiorespiratory fitness outcome.

|  | Cardiorespiratory fitness outcome  Total number of interventions [sum of participants]  Mean (range) | | | | | |
| --- | --- | --- | --- | --- | --- | --- |
|  | AV̇O_2peak_ (L/min) | | RV̇O_2peak_ (mL/kg/min) | | PPO (W) | |
|  | LOW or MOD | VIG or SUPRA | LOW or MOD | VIG or SUPRA | LOW or MOD | VIG or SUPRA |
|  | 2 [15] | 2 [14] | 6 [42] | 6 [41] | 4 [24] | 4 [22] |
| *Participant demographics* | | | | | | |
| Age (years) | 37 (34 – 43) | 45 (34 – 50) | 43 (31 – 52) | 45 (30 – 50) | 42 (31 – 52) | 40 (30 – 47) |
| Baseline CRF | 2.41 (1.65 – 2.79) | 2.31 (1.33 – 2.70) | 22.0 (11.5 – 36.9) | 21.9 (14.2 – 32.1) | 50 (33 – 80) | 55 (33 – 80) |
| *Sex* | | | | | | |
| Male | 1 [5] | - | 1 [5] | 1 [3] | 1 [5] | 2 [13] |
| Female | - | - | - | - | - | - |
| Mixed (% F) | 1 [10] (20%) | 2 [14] (21%) | 4 [34] (32%) | 4 [34] (26%) | 3 [19] (53%) | 2 [9] (33%) |
| Not reported/cannot determine | - | - | 1 [3] | 1 [4] | - | - |
| *Injury characteristics* | | | | | | |
| *Time since injury (years)* | 4 (0 – 12) | 2 (0 – 6) | 6 (0 – 13) | 5 (0 – 10) | 6 (0 – 13) | 4 (0 – 10) |
| Acute (<1-year) | 1 [10] | 1 [10] | 2 [13] | 2 [13] | 2 [13] | 2 [13] |
| Chronic (>1-year) | 1 [5] | 1 [4] | 3 [23] | 4 [28] | 1 [5] | 2 [9] |
| Mixed (% acute) | - | - | 1 [6] (17%) | - | 1 [6] (17%) | - |
| Not reported/cannot determine | - | - | - | - | - | - |
| *Neurological level of injury (TETRA/PARA)* | | | | | | |
| TETRA | - | - | - | - | - | - |
| PARA | - | - | 1 [3] | - | 1 [3] | - |
| Mixed (% PARA) | 2 [15] (67%) | 2 [14] (43%) | 5 [39] (56%) | 6 [41] (44%) | 3 [21] (67%) | 4 [22] (64%) |
| Not reported/cannot determine | - | - | - | - | - | - |
| *Severity* | | | | | | |
| Motor-complete | - | - | - | 1 [4] | - | - |
| Motor-incomplete | 1 [10] | 1 [10] | 3 [28] | 2 [25] | 1 [3] | - |
| Mixed (% incomp.) | 1 [5] (20%) | 1 [4] (25%) | 2 [8] (25%) | 2 [7] (29%) | 2 [15] (67%) | 3 [17] (65%) |
| Not reported/cannot determine | - | - | 1 [6] | 1 [5] | 1 [6] | 1 [5] |
| *Exercise intervention parameters* | | | | | | |
| *Type* | | | | | | |
| Upper-body aerobic exercise | 1 [5] | 1 [4] | 4 [17] | 4 [16] | 4 [24] | 4 [22] |
| Upper-body resistance training/circuits | - | - | - | - | - | - |
| Functional electrical stimulation | - | - | - | - | - | - |
| Gait/locomotor training | 1 [10] | 1 [10] | 2 [25] | 2 [25] | - | - |
| Mixed/multimodal | - | - | - | - | - | - |
| Behaviour change | - | - | - | - | - | - |
| *Relative intensity* | | | | | | |
| Light | - | - | 1 [15] |  | - | - |
| Moderate | 2 [15] | - | 5 [27] |  | 4 [24] | - |
| Moderate-to-vigorous | - | - | - |  | - | - |
| Vigorous | - | 2 [14] | - | 5 [37] | - | 3 [12] |
| Supramaximal | - | - | - | 1 [4] | - | 1 [10] |
| Mixed/cannot determine | - | - | - | - | - | - |
| *Relative intensity prescription* | | | | | | |
| VO_2_ (%peak, %reserve) | - | - | 1 [3] | - | - | - |
| Heart rate (%HRR, %HR_peak_, _-%HR_max_) | 1 [10] | 1 [10] | 4 [34] | 4 [33] | 2 [9] | 2 [8] |
| RPE | 1 [5] | 1 [4] | 1 [5] | 1 [4] | 1 [5] | 1 [4] |
| Workload (%PPO, %MTP,  -_%1RM) | - | - | - | - | - | - |
| Mixed/cannot determine | - | - | - | 1 [4] | 1 [10] | 1 [10] |
| *Session duration (min)* | 40 (30 – 45) | 26 (25 – 30) | 45 (20 – 60) | 40 (20 – 60) | 27 (20 – 60) | 20 (5 – 60) |
| *Frequency (sessions/week)* | 3 (N/A) | 2 (2 – 3) | 3 (3 – 4) | 3 (2 – 4) | 3 (N/A) | 3 (N/A) |
| < 3 | - | 1 [10] | - | 2 [14] | - | - |
| ≥ 3 and < 5 | 2 [15] | 1 [4] | 6 [42] | 4 [27] | 4 [24] | 4 [22] |
| ≥ 5 | - | - | - | - | - | - |
| Not reported | - | - | - | - | - | - |
| *Volume (min/week)* | 120 (90 – 135) | 61 (50 – 90) | 156 (60 – 240) | 133 (40 – 240) | 81 (60 – 180) | 61 (15 – 180) |
| SCI-specific exercise guidelines [fitness (40 – 89 min/wk)] | - | 1 [10]  50 (N/A) | 1 [6]  60 (N/A) | 3 [19]  51 (40 – 60) | 2 [16]  60 (N/A) | 2 [15]  30 (15 – 60) |
| SCI-specific exercise guidelines [cardiometabolic (90 – 149 min/wk)] | 2 [15]  120 (90 – 135) | 1 [4]  90 (N/A) | 3 [18]  115 (90 – 135) | 1 [4]  90 (N/A) | 1 [5]  90 (N/A) | 1 [4]  90 (N/A) |
| Achieving general population exercise guidelines (≥150 min/wk) | - | - | 2 [18]  230 (180 – 240) | 2 [18]  230 (180 – 240) | 1 [3]  180 (N/A) | 1 [3]  180 (N/A) |
| Cannot classify | - | - | - | - | - | - |
| *Length (weeks)* | 10 (6 – 12) | 10 (6 – 12) | 8 (6 – 12) | 8 (6 – 12) | 6 (5 – 8) | 6 (5 – 8) |
| ≤ 6 weeks | 1 [5] | 1 [4] | 3 [23] | 3 [23] | 2 [15] | 2 [14] |
| > 6 and ≤ 12 weeks | 1 [10] | 1 [10] | 3 [19] | 3 [18] | 2 [9] | 2 [8] |
| > 12 weeks | - | - | - | - | - | - |
| *Adverse events reported* | | | | | | |
| Bone, joint or muscular pain | - | 1 [1] | 1 [1] | 1 [1] | - | 1 [1] |
| Autonomic or cardiovascular function | - | - | - | - | - | 1 [1] |
| Skin irritation or pressure sores | - | - | - | - | - | - |
| Other ^a^ | - | - | - | - | - | - |
| Total number of studies (N) and participants, (Σ) along with descriptive characteristics for the primary meta-analysis included in this systematic review that describes Δ in CRF outcomes in response to prospective, well-characterised exercise interventions lasting >2 weeks (e.g., combining exercise intervention-arms from RCTs and pre-post studies). Continuous variables are displayed as weighted means (range: lowest – highest mean values reported from studies). Categorical variables are displayed as n (%). Weighted means were calculated to account for differences in sample size between studies using the following formula: Σn*x̅ /Σn, where Σ = the sum of, n = number of participants in each study and, x̅ = mean CRF outcome of each study. F, females; HR _max_, maximal heart rate; HR _peak_, peak heart rate; HRR, heart rate reserve; 1RM, one repetition maximum; M, males; MTP, maximal tolerated power; NR, not reported; PARA, paraplegia; PPO, peak power output; TETRA, tetraplegia; V̇O_2 peak_, peak oxygen consumption; W, watts. ^a^ Other adverse events included: anxiety, nausea, dizziness and issues with testing equipment. | | | | | | |

| **Table 2:** Summaries of individual randomised controlled trials (low or moderate intensity vs. vigorous or supramaximal intensity). | | | | |
| --- | --- | --- | --- | --- |
| **Author/Year/**  **Country** | **Group** | **Population** | **Training Details** | **Cardiorespiratory Fitness Outcomes** |
| Brazg et al. (2017)  USA | Low or Mod | *N =* 15 (11 M/ 4 F)    *Age =* 49 ± 8.1 years  *TSI =* 7.7 ± 7.9 years (0 acute/ 15 chronic)  *Classification =* 10 T/ 5 P  *Severity =* 0 comp./ 15 incomp.  *CPET =* Modified graded peak treadmill test  *CPET same modality of intervention? =* Yes | *Type of Exercise =* Gait (locomotor) training  *Relative Intensity =* 50-60% HR_max_ (RPE 11-13)  *Session Duration (min) =* 60  *Frequency (times/week) =* 4  *Intervention Length (weeks) =* 6  *Adverse Events =* 1 individual terminated participation due to an increase in his back pain | *AV̇O_2peak_ (L/min)*   - *Pre:* NR - *Post:* NR   *RV̇O_2peak_ (mL/kg/min)*   - *Pre:* 18 ± 6.8 - *Post:* 18 ± 6.1   *PPO (W)*   - *Pre:* NR - *Post:* NR |
|  | Vig or Supra | *See above: cross-over of participants following a 4-week washout period.* | *Type of Exercise =* Gait (locomotor) training  *Relative Intensity =* 70-85% HR_max_ (RPE 15-17)  *Session Duration (min) =* 60  *Frequency (times/week) =* 4  *Intervention Length (weeks) =* 6  *Adverse Events =* None | *AV̇O_2peak_ (L/min)*   - *Pre:* NR - *Post:* NR   *RV̇O_2peak_ (mL/kg/min)*   - *Pre:* 20 ± 7.9 - *Post:* 20 ± 7.5   *PPO (W)*   - *Pre:* NR - *Post:* NR |
| de Groot et al. (2003)  Norway | Low or Mod | *N =* 3 (1 M/ 2 F)    *Age =* 52 ± 2 years  *TSI =* 0.32 ± 0.26 years (3 acute/ 0 chronic)  *Classification =* 0 T/ 3 P  *Severity =* 0 comp./ 3 incomp.  *CPET =* Graded ACE  *CPET same modality of intervention? =* Yes | *Type of Exercise =* ACE interspersed with boxing, push-ups and ball throwing  *Relative Intensity =* 40-55% HRR  *Session Duration (min) =* 60  *Frequency (times/week) =* 3  *Intervention Length (weeks) =* 8  *Adverse Events =* NR | *AV̇O_2peak_ (L/min)*   - *Pre:* NR - *Post:* NR   *RV̇O_2peak_ (mL/kg/min)*   - *Pre:* 14.1 ± 2.5 - *Post:* 16.4 ± 2.9   *PPO (W)*   - *Pre:* 52 ± 20 - *Post:* 65 ± 22 |
|  | Vig or Supra | *N =* 3 (3 M/ 0F)    *Age =* 39 ± 2 years  *TSI =* 0.26 ± 0.20 years (3 acute/ 0 chronic)  *Classification =* 1 T/ 2 P  *Severity =* 2 comp./ 1 incomp.  *CPET =* Graded ACE  *CPET same modality of intervention? =* Yes | *Type of Exercise =* ACE interspersed with boxing, push-ups and ball throwing  *Relative Intensity =* 70-80% HRR  *Session Duration (min) =* 60  *Frequency (times/week) =* 3  *Intervention Length (weeks) =* 8  *Adverse Events =* NR | *AV̇O_2peak_ (L/min)*   - *Pre:* NR - *Post:* NR   *RV̇O_2peak_ (mL/kg/min)*   - *Pre:* 15.1 ± 8.4 - *Post:* 21.3 ± 10.5   *PPO (W)*   - *Pre:* 68 ± 52 - *Post:* 94 ± 70 |
| Gauthier et al. (2018)  Canada | Low or Mod | *N =* 5 (5 M/ 0 F)    *Age =* 43.2 ± 18.5 years  *TSI =* 11.5 ± 10.3 years (0 acute/ 5 chronic)  *Classification =* 1 T/ 4 P  *Severity =* 4 comp./ 1 incomp.  *CPET =* Progressive ACE  *CPET same modality of intervention? =* No | *Type of Exercise =* Wheelchair propulsion  *Relative Intensity =* 4-5 RPE (CR10 scale)  *Session Duration (min) =* 30  *Frequency (times/week) =* 3  *Intervention Length (weeks) =* 6  *Adverse Events =* NR | *AV̇O_2peak_ (L/min)*   - *Pre:* 1.65 ± 0.59 - *Post:* 1.74 ± 0.66   *RV̇O_2peak_ (mL/kg/min)*   - *Pre:* 18.5 ± 6.8 - *Post:* 18.9 ± 8.4   *PPO (W)*   - *Pre:* 80 ± 26 - *Post:* 82 ± 28 |
|  | Vig or Supra | *N =* 4 (3 M/ 1 F)    *Age =* 33.9 ± 3 years  *TSI =* 6 ± 3.6 years (0 acute/ 4 chronic)  *Classification =* 1 T/ 3 P  *Severity =* 3 comp./ 1 incomp.  *CPET =* Progressive ACE  *CPET same modality of intervention? =* No | *Type of Exercise =* Wheelchair propulsion  *Relative Intensity =* High intensity 6-8 RPE, low intensity 1-2 RPE (CR10 scale)  *Session Duration (min) =* 30 (30s high intensity, 60s low intensity bouts)  *Frequency (times/week) =* 3  *Intervention Length (weeks) =* 6  *Adverse Events =* One dropout due to development of significant shoulder pain | *AV̇O_2peak_ (L/min)*   - *Pre:* 1.33 ± 0.27 - *Post:* 1.46 ± 0.32   *RV̇O_2peak_ (mL/kg/min)*   - *Pre:* 19.5 ± 0.70 - *Post:* 20.4 ± 3.9   *PPO (W)*   - *Pre:* 80 ± 18 - *Post:* 80 ± 14 |
| Graham et al. (2019)  USA | Low or Mod | *N =* 3 (sex NR)    *Age =* 51.3 ± 1.2 years  *TSI =* NR (0 acute/ 3 chronic)  *Classification =* 1 T/ 2 P  *Severity =* 2 comp./ 1 incomp.  *CPET =* ACE  *CPET same modality of intervention? =* Yes | *Type of Exercise =* ACE  *Relative Intensity =* 55% V̇O_2peak_  *Session Duration (min) =* 30  *Frequency (times/week) =* 3  *Intervention Length (weeks) =* 6  *Adverse Events =* NR | *AV̇O_2peak_ (L/min)*   - *Pre:* NR - *Post:* NR   *RV̇O_2peak_ (mL/kg/min)*   - *Pre:* 11.5 ± 2.6 - *Post:* 13.9 ± 1.3   *PPO (W)*   - *Pre:* NR - *Post:* NR |
|  | Vig or Supra | *N =* 4 (sex NR)    *Age =* 49.4 ± 13 years  *TSI =* NR (0 acute/ 4 chronic)  *Classification =* 2 T/ 2 P  *Severity =* 4 comp./ 0 incomp.  *CPET =* ACE  *CPET same modality of intervention? =* Yes | *Type of Exercise =* ACE  *Relative Intensity =* 25% HRR and 50% PPO  *Session Duration (min) =* 20 (4-min at 25% HRR, 30-s at 50% PPO; 2-min recovery to finish at 25% HRR)  *Frequency (times/week) =* 2  *Intervention Length (weeks) =* 6  *Adverse Events =* NR | *AV̇O_2peak_ (L/min)*   - *Pre:* NR - *Post:* NR   *RV̇O_2peak_ (mL/kg/min)*   - *Pre:* 14.2 ± 6.0 - *Post:* 15.3 ± 7.3   *PPO (W)*   - *Pre:* NR - *Post:* NR |
| Hooker and Wells (1989)  USA | Low or Mod | *N =* 6 (3 M/ 3 F)    *Age =* 31.3 ± 4.2 years  *TSI =* 12.88 ± 6.8 years (1 acute/ 5 chronic)  *Classification =* 1 T/ 5 P  *Severity =* NR  *CPET =* Incremental discontinuous wheelchair ergometer exercise  *CPET same modality of intervention? =* Yes | *Type of Exercise =* Wheelchair ergometry  *Relative Intensity =* 50-60% HRR  *Session Duration (min) =* 20  *Frequency (times/week) =* 3  *Intervention Length (weeks) =* 8  *Adverse Events =* NR | *AV̇O_2peak_ (L/min)*   - *Pre:* NR - *Post:* NR   *RV̇O_2peak_ (mL/kg/min)*   - *Pre:* 19.4 ± 8.1 - *Post:* 21.4 ± 6.5   *PPO (W)*   - *Pre:* 33 ± 29 - *Post:* 41 ± 30 |
|  | Vig or Supra | *N =* 5 (3 M/ 2 F)    *Age =* 30.4 ± 5 years  *TSI =* 10.2 ± 7.9 years (0 acute/ 5 chronic)  *Classification =* 2 T/ 3 P  *Severity =* NR  *CPET =* Incremental discontinuous wheelchair ergometer exercise  *CPET same modality of intervention? =* Yes | *Type of Exercise =* Wheelchair ergometry  *Relative Intensity =* 70-80% HRR  *Session Duration (min) =* 20  *Frequency (times/week) =* 3  *Intervention Length (weeks) =* 8  *Adverse Events =* NR | *AV̇O_2peak_ (L/min)*   - *Pre:* NR - *Post:* NR   *RV̇O_2peak_ (mL/kg/min)*   - *Pre:* 19.2 ± 9.8 - *Post:* 21.5 ± 9.2   *PPO (W)*   - *Pre:* 33 ± 33 - *Post:* 38 ± 36 |
| Mcleod et al. (2020)  Canada | Low or Mod | *N =* 10 (5 M/ 5 F)    *Age =* 45 ± 17 years  *TSI =* 0.15 ± 0.12 years (10 acute/ 0 chronic)  *Classification =* 5 T/ 5 P  *Severity =* 1 comp./ 9 incomp.  *CPET =* ACE  *CPET same modality of intervention? =* Yes | *Type of Exercise =* ACE  *Relative Intensity =* 12 RPE (45% PPO)  *Session Duration (min) =* 20  *Frequency (times/week) =* 3  *Intervention Length (weeks) =* 5  *Adverse Events =* NR | *AV̇O_2peak_ (L/min)*   - *Pre:* NR - *Post:* NR   *RV̇O_2peak_ (mL/kg/min)*   - *Pre:* NR - *Post:* NR   *PPO (W)*   - *Pre:* 45 ± 20 - *Post:* 58 ± 21 |
|  | Vig or Supra | *N =* 10 (10 M/ 0 F)    *Age =* 47 ± 15 years  *TSI =* 0.20 ± 0.19 years (10 acute/ 0 chronic)  *Classification =* 4 T/ 6 P  *Severity =* 1 comp./ 9 incomp.  *CPET =* ACE  *CPET same modality of intervention? =* Yes | *Type of Exercise =* ACE  *Relative Intensity =* ‘All out efforts’ and 10% PPO active recovery  *Session Duration (min) =* 5 (3 x 20-s supramaximal bouts interspersed by 120-s active recovery)  *Frequency (times/week) =* 3  *Intervention Length (weeks) =* 5  *Adverse Events =* 1 individual experienced post-exercise hypotension | *AV̇O_2peak_ (L/min)*   - *Pre:* NR - *Post:* NR   *RV̇O_2peak_ (mL/kg/min)*   - *Pre:* NR - *Post:* NR   *PPO (W)*   - *Pre:* 52 ± 29 - *Post:* 69 ± 37 |
| Wouda et al. (2018)  Norway | Low or Mod | *N =* 10 (8 M/ 2 F)    *Age =* 34 ± 15 years  *TSI =* 0.18 ± 0.09 years (10 acute/ 0 chronic)  *Classification =* 4 T/ 6 P  *Severity =* 0 comp./ 10 incomp.  *CPET =* Maximal graded treadmill test  *CPET same modality of intervention? =* Yes | *Type of Exercise =* Gait training (walking or running)  *Relative Intensity =* 70% HR_peak_  *Session Duration (min) =* 45  *Frequency (times/week) =* 3  *Intervention Length (weeks) =* 12  *Adverse Events =* NR | *AV̇O_2peak_ (L/min)*   - *Pre:* 2.79 ± 0.79 - *Post:* 3.23 ± 0.94   *RV̇O_2peak_ (mL/kg/min)*   - *Pre:* 36.9 ± 11.8 - *Post:* 42.3 ± 12.0   *PPO (W)*   - *Pre:* NR - *Post:* NR |
|  | Vig or Supra | *N =* 10 (8 M/ 2 F)    *Age =* 50 ± 15 years  *TSI =* 0.19 ± 0.08 years (10 acute/ 0 chronic)  *Classification =* 7 T/ 3 P  *Severity =* 0 comp./ 10 incomp.  *CPET =* Maximal graded treadmill test  *CPET same modality of intervention? =* Yes | *Type of Exercise =* Gait training (walking or running)  *Relative Intensity =* 85-95% HR_peak_ and 70% HR_peak_  *Session Duration (min) =* 25 (4 x 4-min intervals at 85-95% HR_peak_ interspersed with 3 x 3-min recovery at 70% HR_peak_)  *Frequency (times/week) =* 2  *Intervention Length (weeks) =* 12  *Adverse Events =* NR | *AV̇O_2peak_ (L/min)*   - *Pre:* 2.70 ± 0.81 - *Post:* 3.00 ± 0.62   *RV̇O_2peak_ (mL/kg/min)*   - *Pre:* 32.1 ± 9.1 - *Post:* 35.7 ± 5.3   *PPO (W)*   - *Pre:* NR - *Post:* NR |
| Data is presented as mean ± standard deviation, unless otherwise stated. Significant changes in CRF outcomes from pre- to post-intervention are highlighted in bold (p<0.05). ACE, arm crank ergometry; AV̇O_2peak_, absolute peak oxygen consumption; CPET, cardiopulmonary exercise test; F, females; HR_max_, maximum heart rate; HR_peak_, peak heart rate; HRR, heart rate reserve; M, male; Mod, moderate-intensity; NR, not reported; P, paraplegia; PPO, peak power output; RCT, randomised-controlled trial; RPE, rating of perceived exertion; RV̇O_2peak_, relative peak oxygen consumption; Supra, supramaximal-intensity; T, tetraplegia; TSI, time since injury; Vig, vigorous-intensity; V̇O_2peak_, peak oxygen consumption; W, watts. | | | | |

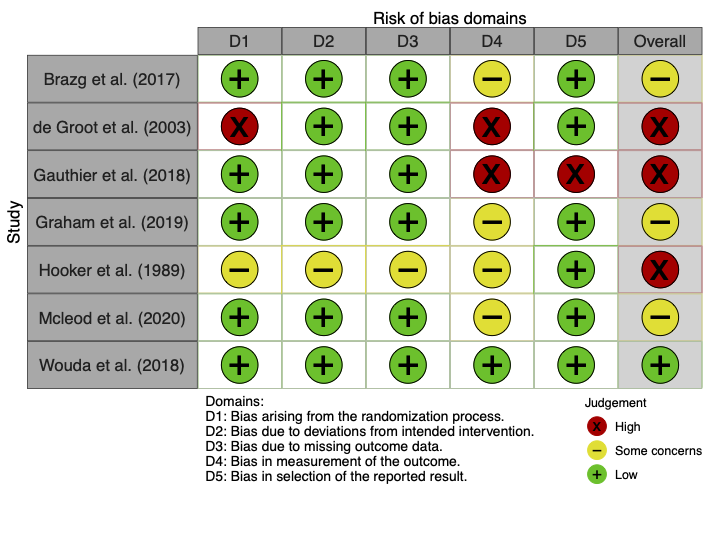

**Figure 1:** Traffic light plot for risk of bias in the intensity comparison RCTs, assessed via the Cochrane RoB 2 tool.

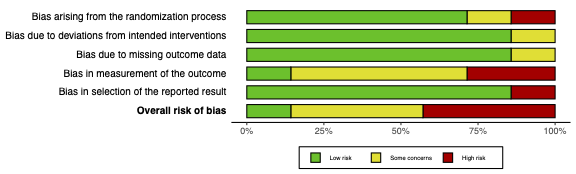

**Figure 2:** Summary plot for risk of bias in the intensity comparison RCTs, assessed via the Cochrane RoB 2 tool.

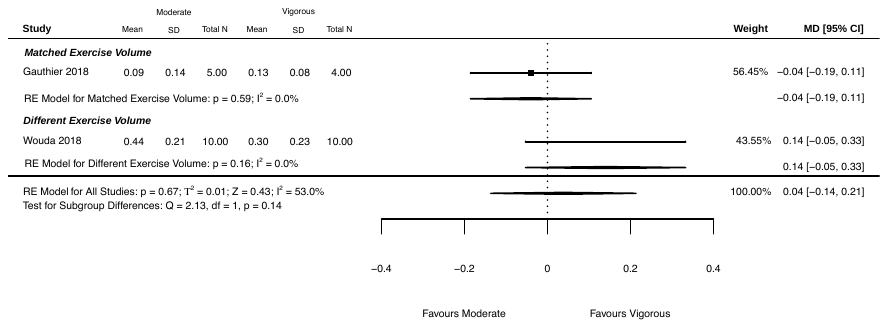

Low or Mod

Vig or Supra

Favours Low or Mod

Favours Vig or Supra

**Figure 3:** Changes in absolute peak oxygen consumption following low or moderate and vigorous or supramaximal intensity exercise intervention comparisons in randomised-controlled trial studies. Interventions are grouped into subgroups depending on whether intensity groups performed the same weekly exercise volume. MD, mean difference; Mod, moderate-intensity; N, number of participants; RE, random effects; SD, standard deviation; Supra, supramaximal-intensity; Vig, vigorous-intensity.

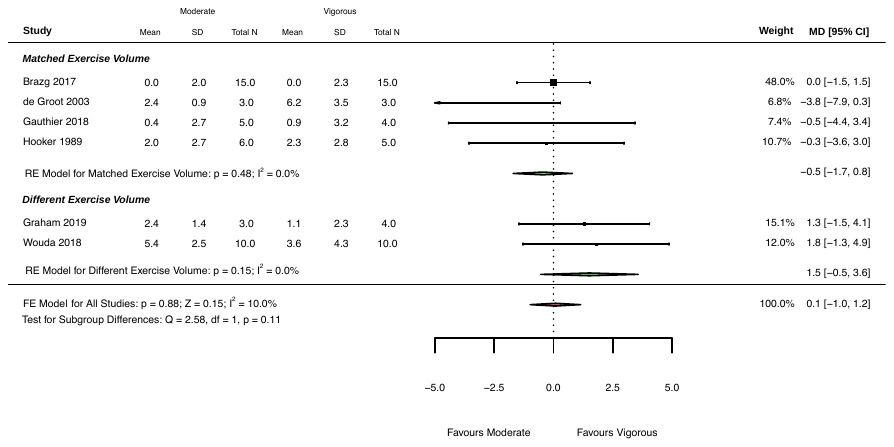

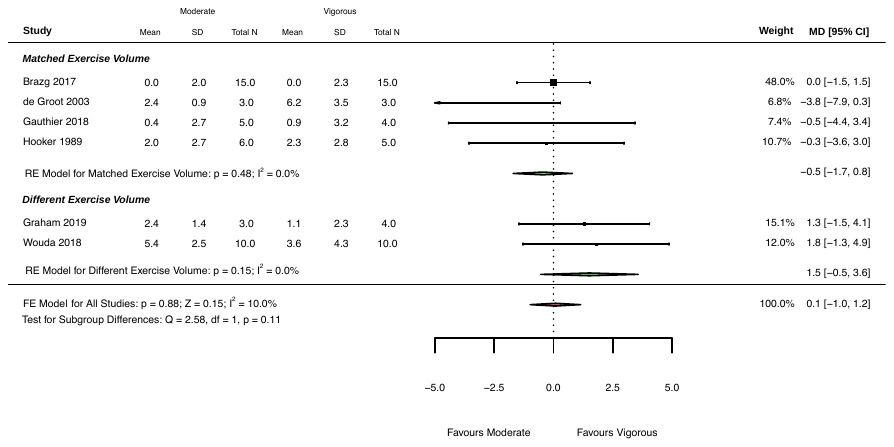

Low or Mod

Vig or Supra

Favours Low or Mod

Favours Vig or Supra

**Figure 4:** Changes in relative peak oxygen consumption following low or moderate and vigorous or supramaximal intensity exercise intervention comparisons in randomised-controlled trial studies. Interventions are grouped into subgroups depending on whether intensity groups performed the same weekly exercise volume. MD, mean difference; Mod, moderate-intensity; N, number of participants; RE, random effects; SD, standard deviation; Supra, supramaximal-intensity; Vig, vigorous-intensity.

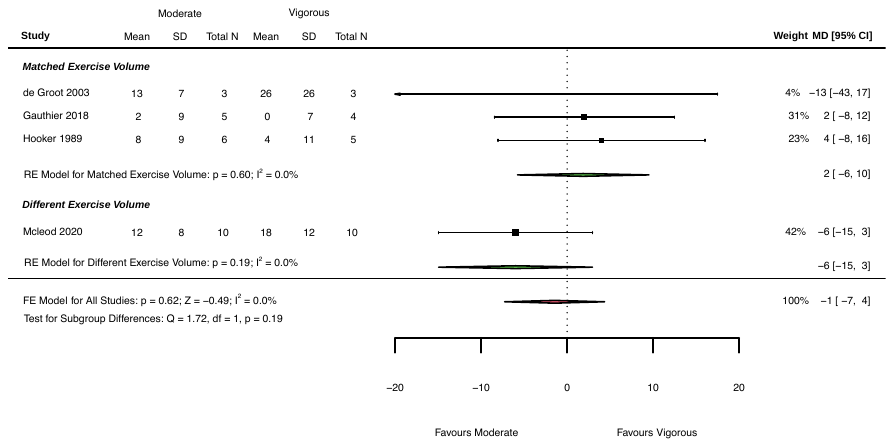

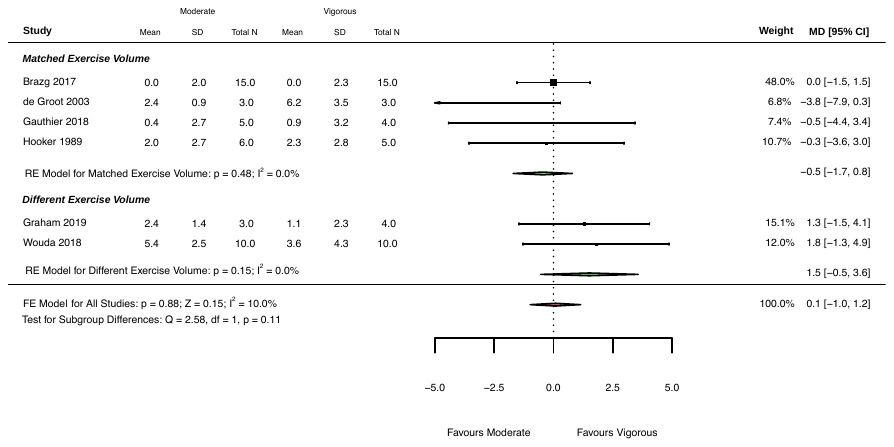

Low or Mod

Vig or Supra

Favours Low or Mod

Favours Vig or Supra

**Figure 5:** Changes in peak power output following low or moderate and vigorous or supramaximal intensity exercise intervention comparisons in randomised-controlled trial studies. Interventions are grouped into subgroups depending on whether intensity groups performed the same weekly exercise volume. MD, mean difference; Mod, moderate-intensity; N, number of participants; RE, random effects; SD, standard deviation; Supra, supramaximal-intensity; Vig, vigorous-intensity.

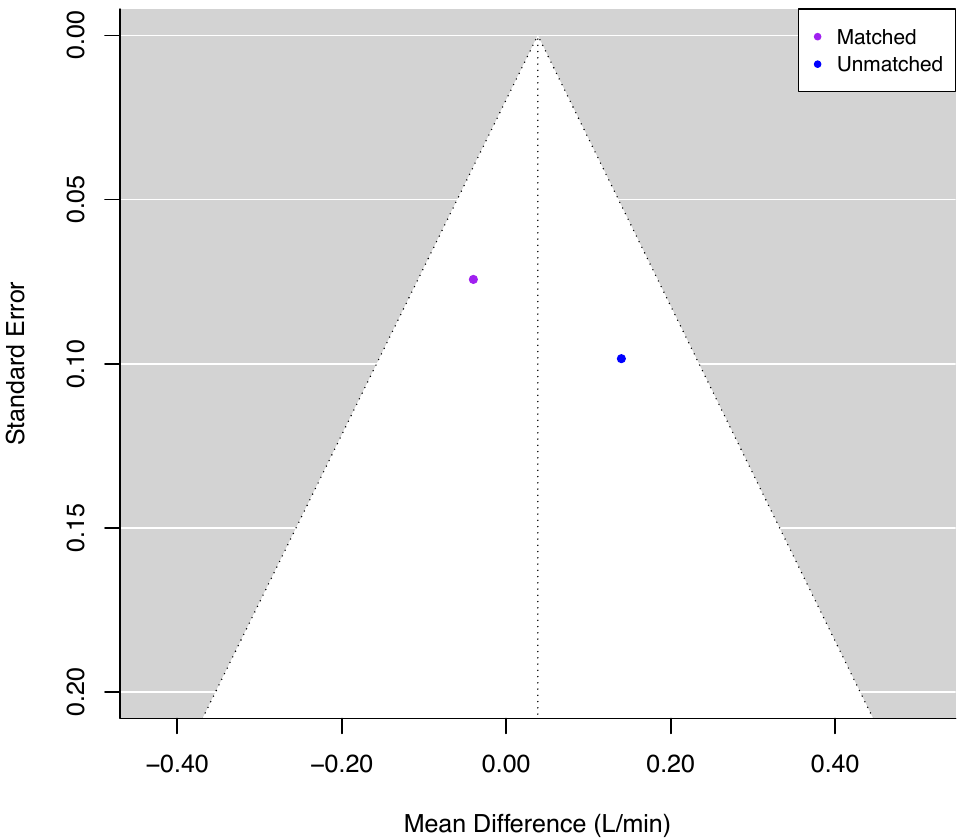

**Figure 6:** Funnel plot of absolute peak oxygen consumption with studies sub-grouped based on whether interventions matched exercise volume between intensity groups (matched) or did not (unmatched). Egger’s test not performed (<10 studies).

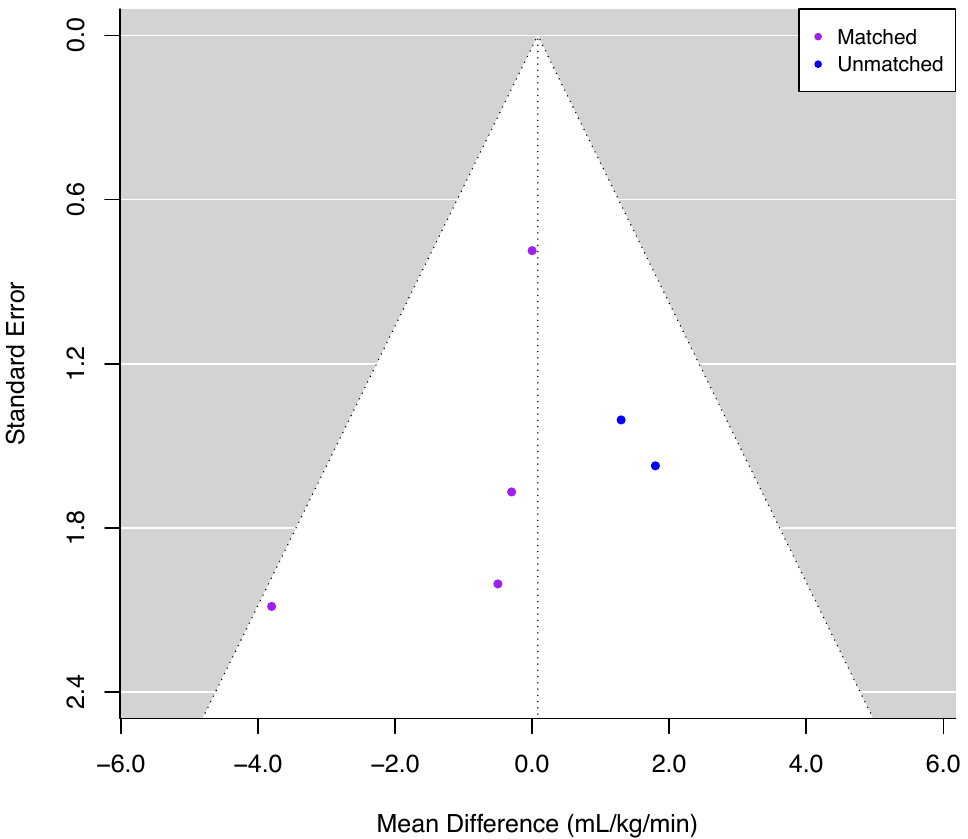

**Figure 7:** Funnel plot of relative peak oxygen consumption with studies sub-grouped based on whether interventions matched exercise volume between intensity groups (matched) or did not (unmatched). Egger’s test not performed (<10 studies).

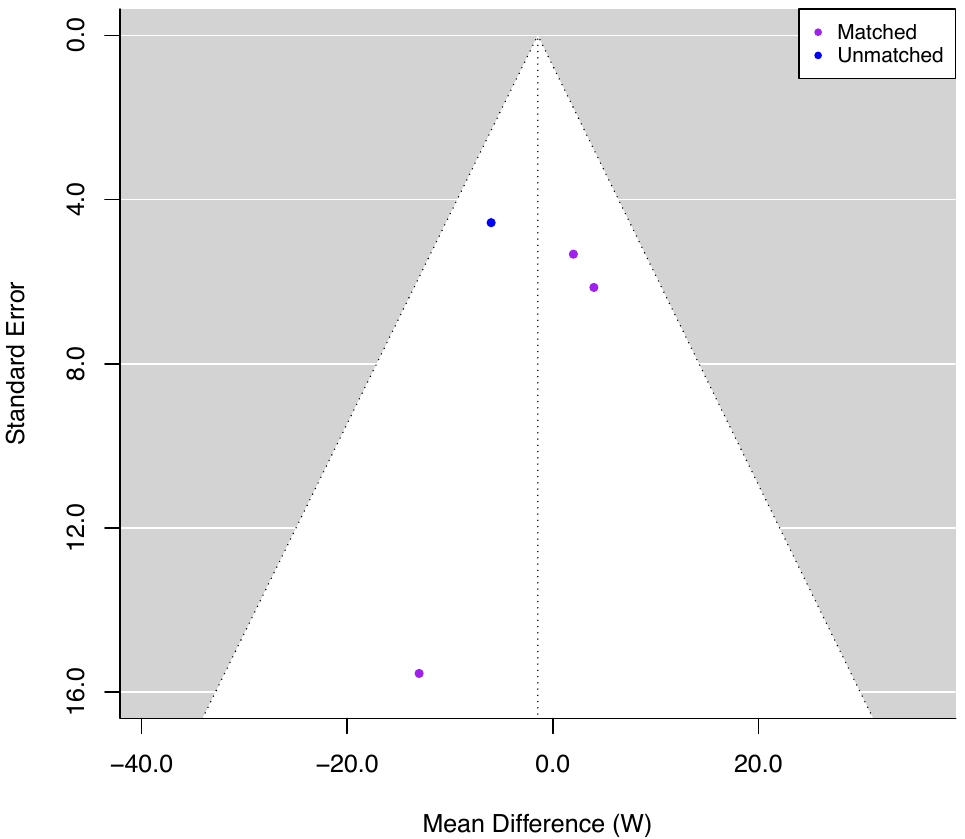

**Figure 8:** Funnel plot of peak power output with studies sub-grouped based on whether interventions matched exercise volume between intensity groups (matched) or did not (unmatched). Egger’s test not performed (<10 studies).

**References**

RCT intensity comparison (low or moderate-intensity vs vigorous or supramaximal-intensity) studies included in the systematic review, sorted alphabetically:

Brazg G, Fahey M, Holleran CL, Connolly M, Woodward J, Hennessy PW, et al. Effects of Training Intensity on Locomotor Performance in Individuals With Chronic Spinal Cord Injury: A Randomized Crossover Study. Neurorehabil Neural Repair. 2017;31: 944–954.

de Groot PCE, Hjeltnes N, Heijboer AC, Stal W, Birkeland K. Effect of training intensity on physical capacity, lipid profile and insulin sensitivity in early rehabilitation of spinal cord injured individuals. Spinal Cord. 2003;41: 673–679.

Gauthier C, Brosseau R, Hicks AL, Gagnon DH. Feasibility, Safety, and Preliminary Effectiveness of a Home-Based Self-Managed High-Intensity Interval Training Program Offered to Long-Term Manual Wheelchair Users. Rehabil Res Pract. 2018;2018: 8209360.

Graham K, Yarar-Fisher C, Li J, McCully KM, Rimmer JH, Powell D, et al. Effects of High-Intensity Interval Training Versus Moderate-Intensity Training on Cardiometabolic Health Markers in Individuals With Spinal Cord Injury: A Pilot Study. Top Spinal Cord Inj Rehabil. 2019;25: 248–259.

Hooker SP, Wells CL. Effects of low- and moderate-intensity training in spinal cord-injured persons. Med Sci Sports Exerc. 1989;21: 18–22.

Mcleod JC, Diana H, Hicks AL. Sprint interval training versus moderate-intensity continuous training during inpatient rehabilitation after spinal cord injury: a randomized trial. Spinal Cord. 2020;58: 106–115.

Wouda MF, Lundgaard E, Becker F, Strøm V. Effects of moderate- and high-intensity aerobic training program in ambulatory subjects with incomplete spinal cord injury-a randomized controlled trial. Spinal Cord. 2018;56: 955–963.
